# Effectiveness of Tocilizumab, Sarilumab, and Anakinra for critically ill patients with COVID-19 The REMAP-CAP COVID-19 Immune Modulation Therapy Domain Randomized Clinical Trial

**DOI:** 10.1101/2021.06.18.21259133

**Authors:** The REMAP-CAP Investigators, Lennie P.G. Derde

## Abstract

**BACKGROUND:** The interleukin-6 receptor antagonist tocilizumab improves outcomes in critically ill patients with coronavirus disease 2019 (COVID-19). However, the effectiveness of other immune modulating agents is unclear.

**METHODS:** We evaluated four immunomodulatory agents in an ongoing international, multifactorial, adaptive platform trial. Adult participants with COVID-19 were randomized to receive tocilizumab, sarilumab, anakinra, or standard care (control). In addition, a small group (n=21) of participants were randomized to interferon-β1a. The primary outcome was an ordinal scale combining in-hospital mortality (assigned -1) and days free of organ support to day 21. The trial used a Bayesian statistical model with pre-defined triggers for superiority, equivalence or futility.

**RESULTS:** Statistical triggers for equivalence between tocilizumab and sarilumab; and for inferiority of anakinra to the other active interventions were met at a planned adaptive analysis. Of the 2274 critically ill participants enrolled, 972 were assigned to tocilizumab, 485 to sarilumab, 378 to anakinra and 418 to control. Median organ support-free days were 7 (interquartile range [IQR] –1, 16), 9 (IQR –1, 17), 0 (IQR –1, 15) and 0 (IQR –1, 15) for tocilizumab, sarilumab, anakinra and control, respectively. Median adjusted odds ratios were 1.46 (95%CrI 1.13, 1.87), 1.50 (95%CrI 1.13, 2.00), and 0.99 (95%CrI 0.74, 1.35) for tocilizumab, sarilumab and anakinra, yielding 99.8%, 99.8% and 46.6% posterior probabilities of superiority, respectively, compared to control. Median adjusted odds ratios for hospital survival were 1.42 (95%CrI 1.05,1.93), 1.51 (95%CrI 1.06, 2.20) and 0.97 (95%CrI 0.66, 1.40) for tocilizumab, sarilumab and anakinra respectively, compared to control, yielding 98.8%, 98.8% and 43.6% posterior probabilities of superiority, respectively, compared to control. All treatments appeared safe.

**CONCLUSIONS:** In patients with severe COVID-19 receiving organ support, tocilizumab and sarilumab are similarly effective at improving survival and reducing duration of organ support. Anakinra is not effective in this population. (ClinicalTrials.gov number: NCT02735707)

## Background

Both corticosteroids and tocilizumab, an interleukin-6 receptor antagonist (IL-6ra), reduce the need for organ support and increase survival for hospitalized patients with COVID-19.^1–4^ Sarilumab, another IL-6ra, also improved outcomes.^4^ However, sarilumab was evaluated in a small number of patients, and the comparative effectiveness of these two agents, and the effectiveness of other immunomodulators, is not known.

The demonstrated efficacy of modulating the host inflammatory response in patients with severe COVID-19 suggests that alternative approaches may also be effective. Interleukin-1 (IL-1) mediates a range of cellular responses involved in acute inflammation,^5^ and is therefore a potential therapeutic target in COVID-19.^6,7^ A recombinant form of the endogenous IL-1 receptor antagonist, anakinra, is widely used to treat autoinflammatory diseases. Coronaviruses such as SARS-CoV-2 also dampen the host interferon response to infection.^8–12^ Moreover, interferon-β may attenuate lung injury by enhancing endothelial barrier function.^13–15^

We reported initial findings from this platform when the statistical trigger for superiority of tocilizumab (and later sarilumab) compared with control was met. We now report additional conclusions from the Immune Modulation Therapy domain of the Randomized, Embedded, Multifactorial Adaptive Platform Trial for Community-Acquired Pneumonia (REMAP-CAP) following statistical triggers for equivalence between tocilizumab and sarilumab; and for inferiority of anakinra to the other active interventions.

## Methods

### Trial Design and Oversight

REMAP-CAP is an international, adaptive platform trial evaluating treatment strategies for participants with severe pneumonia including COVID-19.^16^ Eligible participants are randomized to multiple interventions across multiple domains. A ‘domain’ covers a common therapeutic area and contains two or more interventions. The REMAP-CAP trial is guided by a master (‘core’) protocol with individual appendices for each domain, regional governance, and adaptations during a declared pandemic. The platform initially included only participants admitted to an intensive care unit (ICU) and receiving respiratory or cardiovascular organ support (referred to as the Severe State, or critically ill patients). We subsequently added a Moderate State, enrolling hospitalized participants not receiving respiratory or cardiovascular organ support, referred to as non-critically ill patients. The trial is managed by a blinded International Trial Steering Committee (ITSC) and an unblinded independent Data and Safety Monitoring Board. It is approved by relevant regional ethics committees and conducted in accordance with Good Clinical Practice guidelines and the principles of the Declaration of Helsinki. Written or verbal informed consent, in accordance with regional legislation, is obtained from all participants or their surrogates.

The trial has multiple funders and multiple regional sponsors and drug supply is supported by several pharmaceutical companies (see **Funding**). The funders, sponsors, and pharmaceutical companies had no role in designing the trial, analyzing data, writing the manuscript, or making the decision to submit for publication. All authors vouch for the data and analyses, as well as for the fidelity of this report to the trial protocol and statistical analysis plan.

### Participants

Initially, participants aged ≥ 18 years, within 24 hours of receiving respiratory or cardiovascular organ support in an ICU with suspected or microbiologically confirmed COVID-19 could be enrolled. We refer to these participants here as critically ill. Respiratory organ support was defined as invasive or non-invasive mechanical ventilation including via high flow nasal cannula if the flow rate was >30 L/min and the fraction of inspired oxygen was >0.4. Cardiovascular organ support was defined as the intravenous infusion of any vasopressor or inotrope.

Exclusion criteria included presumption that death was imminent with lack of commitment to full support, and prior participation in REMAP-CAP within 90 days. Additional exclusion criteria, specific for the Immune Modulation Therapy domain, are listed in the Supplementary appendix. Later, the domain opened to enrollment for anakinra, interferon-β1a and control for non-critically ill patients, defined as hospitalized adult patients not receiving respiratory or cardiovascular organ support in ICU, with suspected or microbiologically confirmed COVID-19 (**Figure S1, Supplementary appendix**).

### Randomization

The Immune Modulation Therapy domain initially included five interventions: two IL-6 receptor antagonists (tocilizumab and sarilumab), the IL-1 receptor antagonist anakinra, interferon-β1a, and control (no immune modulation). Investigators at each site selected *a priori* at least two of the available interventions to which participants would be randomized; initially one intervention had to be control. Participants were randomized via a centralized computer program starting with balanced assignment for each intervention, including control, with proportions at each site dependent on the number of interventions available at each site. An *a priori* negative interaction between interferon-β1a and corticosteroid interventions was prespecified, because of a previously reported potential hazard of the combination treatment.^17^ Clinical use of corticosteroids excluded randomization to interferon-β1a, resulting in low rate of recruitment when corticosteroids became standard of care. The control arm was closed on November 19, 2020 when the statistical trigger for superiority of tocilizumab (and later sarilumab) compared with control was met.^4^ We describe the adaptions to this domain during the trial in **Figure S1** (**Supplementary appendix**).

### Procedures

Tocilizumab, at a dose of 8mg/kg of actual body weight (up to a maximum of 800mg), was administered as an intravenous infusion over one hour; this dose could be repeated 12-24 hours later at the discretion of the treating clinician if clinical improvement was judged insufficient. Sarilumab, 400mg, was administered as a single intravenous infusion. Anakinra was administered intravenously as 300mg loading dose, followed by 100mg every 6 hours for 14 days or until either free from invasive mechanical ventilation for more than 24 hours, or discharge from ICU. In participants who had a creatinine clearance <30ml/min or were receiving renal replacement therapy, the dosing interval was increased to 12 hours. All investigational drugs were dispensed by local pharmacies and were open label.

Other aspects of patient management were provided according to each site’s standard of care. Participants could also be randomized to other interventions within other domains, based on domains active at the site, participant eligibility, and consent. Although clinical staff were aware of individual participant intervention assignment, neither they nor the ITSC were provided any information about randomization ratios or aggregate patient outcomes.

### Outcome Measures

The primary outcome was an ordinal scale that is a composite of in-hospital mortality and duration of respiratory and cardiovascular organ support, censored at 21 days, where all deaths within hospital and up to day 90 were assigned the worst outcome (–1). Among survivors, respiratory and cardiovascular organ support-free days were calculated up to day 21, such that a higher number represents faster recovery. Secondary outcomes were all prespecified and are listed in the **Supplementary appendix**. Adherence to allocated intervention was defined as receipt of at least one dose of the allocated drug for the active interventions, and receiving no immune modulator for the control arm.

### Statistical Analysis

The Statistical Analysis Plan for the Immune Modulation Therapy domain was written blinded to all data other than the statistical triggers from the prior adaptive analyses and posted online (www.remapcap.org) before data lock and final analyses (**Supplementary appendix, Appendix 1**).

REMAP-CAP uses a Bayesian design,^16^ the primary model adjusted for location (site, nested within country), age (categorized into six groups), sex, and time-period (two-week calendar epochs) to account for rapid changes in clinical care and outcomes over time during the pandemic. The temporal adjustment models the change in organ support-free days over time, and allows comparison of non-concurrently randomized interventions across time periods.^18,19^ The model contained treatment effects for each intervention compared with control within each domain and pre-specified treatment-by-treatment interactions across domains. The treatment effects for tocilizumab and sarilumab were nested in the primary model.^4^ Prior distributions for individual treatment effects presented here were neutral. Sensitivity analyses were performed with time and site factors removed and for the per protocol population.

The primary analysis (**Supplementary appendix, Appendix 2**) was conducted by the Statistical Analysis Committee, and included all participants with suspected or proven COVID-19 randomized to any domain up to April 10, 2021, (the date the statistical triggers described above were met), who had completed at least 21 days follow up, for whom an outcome was known. The inclusion of additional data from participants enrolled outside the Immune Modulation Therapy domain provided robust estimation of the coefficients of other covariates, as per the principle of the REMAP-CAP design.^16^ The model included covariate terms reflecting each patient’s domain eligibility, such that the estimate of an intervention’s effectiveness, relative to any other intervention within that domain, was generated from those patients that were eligible to be randomized to those interventions within the domain.

The odds ratio for the primary outcome was modeled such that a parameter >1 reflected an increase in the cumulative odds for the organ support-free days outcome, implying benefit. Missing outcomes were not imputed. The model was fit using a Markov Chain Monte Carlo algorithm that drew iteratively (20,000 draws) from the joint posterior distribution. This allows calculation of posterior odds ratios with their 95% credible intervals (CrI) and the probability that each intervention (including control) was optimal in the domain, that an intervention was superior compared with control (efficacy), that two non-control interventions were equivalent, or an intervention was futile compared with control. The pre-defined statistical triggers for trial conclusions and disclosure of results are described in the Statistical Appendix to the protocol (www.remapcap.org).

Analysis of the primary outcome was repeated in a second model using only data from Severe State participants and domains that had stopped and were unblinded at the time of analysis with no adjustment for assignment in other ongoing domains (**Supplementary appendix, Appendix 3**). The secondary outcomes were also analyzed in this second population. Further details of all analyses are provided and pre-specified analyses are listed in the **Supplementary appendix**. Included are analyses of interactions with the Therapeutic Anticoagulation domain and Immunoglobulin domain, because they have also been unblinded.^20,21^ We evaluated participants co-randomized in the Immunoglobulin domain or Therapeutic Anticoagulation domain, for whom data on major thrombotic events (MTE) were available, by treatment allocated in the Immune Modulation Therapy domain and bleeding events for those co-randomized to both the Therapeutic Anticoagulation and Immune Modulation Therapy domain. Prespecified subgroup analyses are reported for participants with or without invasive mechanical ventilation and for CRP terciles. Secondary analyses are also reported for tocilizumab and sarilumab independently and without nesting as described in the Statistical Analysis Plan (**Supplementary appendix, Appendix 1**).

## Results

Statistical triggers for equivalence between tocilizumab and sarilumab; and for inferiority of anakinra to the other active interventions in critically ill participants were met at a planned adaptive analysis communicated to the ITSC on April 9, 2021. The ITSC closed all arms of the domain on April 10, 2021.

At that time, 6023 participants had been randomized in at least one domain in REMAP-CAP and 2274 critically ill participants had been randomized in the Immune Modulation Therapy domain (972 tocilizumab, 485 sarilumab, 378 anakinra, 21 interferon-β1a and 418 control) in 133 sites across 9 countries (**Figure 1**). 39 of these participants subsequently withdrew consent, and 19 had missing primary outcomes. Only five non-critically ill participants were randomized, two to anakinra and three to control. Baseline characteristics and a description of the non-critically ill participants (n=5) and crude results for interferon-β1a (n=19) are given in the **Supplementary appendix**.

**Figure 1.**
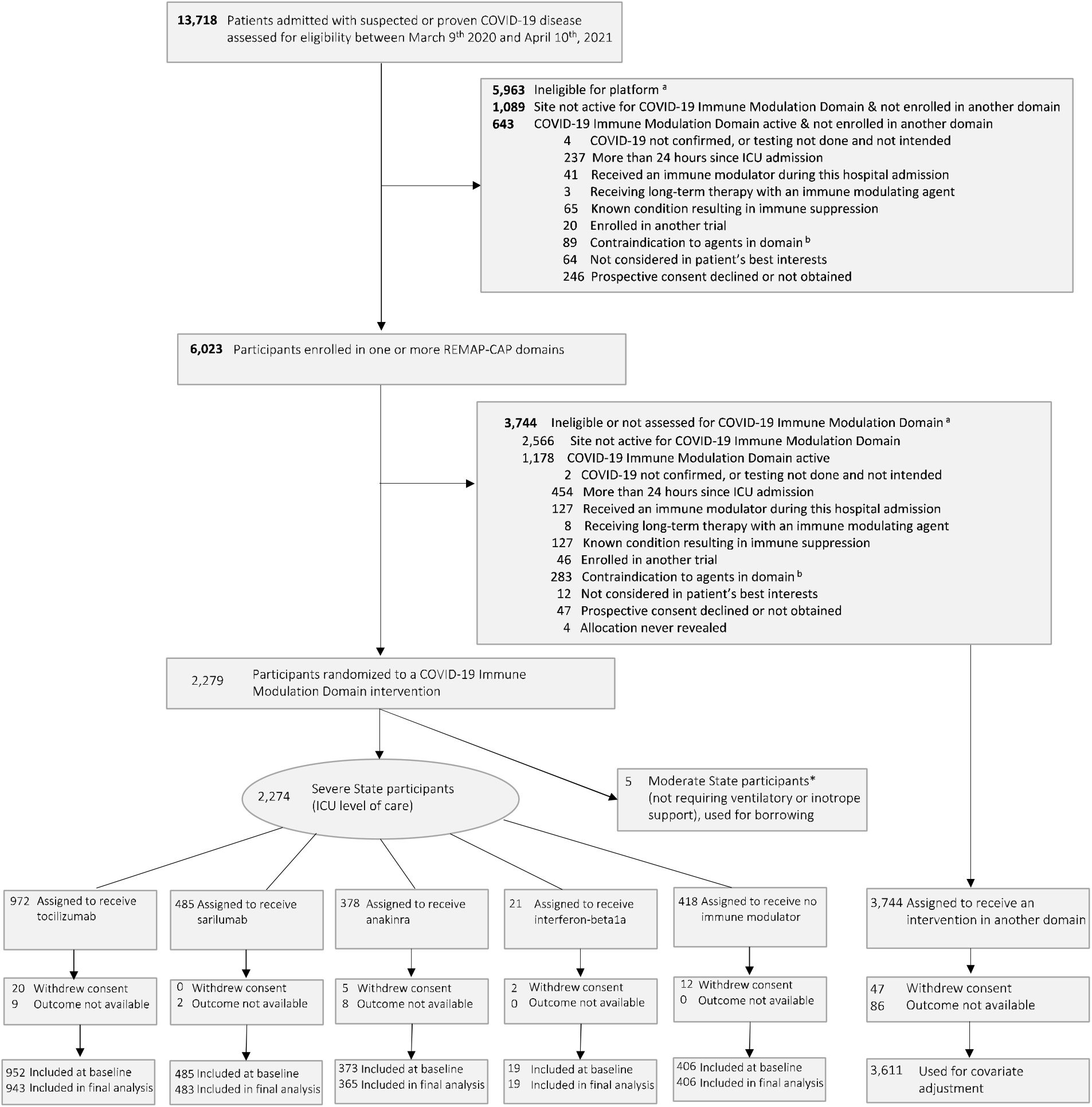
Screening, enrollment, randomization and inclusion in analysis. a = Patients could meet more than one ineligibility criterion. Full details are provided in the supplement b = Contraindications include known hypersensitivity to an agent specified as an intervention; known or suspected pregnancy; known hypersensitivity to proteins produced by E. coli (exclusion from receiving anakinra); baseline alanine aminotransferase or an aspartate aminotransferase that is more than five times the upper limit of normal (exclusion from receiving tocilizumab or sarilumab); baseline platelet count < 50 × 10^9^ / L (exclusion from receiving tocilizumab or sarilumab) *Commencement of organ support in ICU was used instead of date and time of ICU admission for patients who had already received an allocation in the Moderate State

### Patients

Baseline characteristics for critically ill participants were balanced across intervention groups (**Tables 1 and S1)**. All but four participants were receiving respiratory support at the time of randomization, including high flow nasal oxygen (536/2235, 24%), non-invasive (958/2235; 42.9%) and invasive (735/2235; 32.9%) mechanical ventilation. Adherence was 96.7% for tocilizumab, 96.0% for sarilumab and 94.6% for anakinra. Of 906 patients receiving tocilizumab, 295 received more than one dose (32.6%). Of participants allocated to the control group, 7/402 (1.7%) received one or more of the drugs available in this domain. Patients could be co-enrolled in other domains (**Table S2**).

**Table 1.** Participant Characteristics at Baseline (Severe State)*

|  | **Tocilizumab**<br>**(n = 952)** | **Sarilumab**<br>**(n = 485)** | **Anakinra**<br>**(n = 373)** | **Control**<br>**(n = 406)** |
| --- | --- | --- | --- | --- |
| Age - mean (SD), years | 60.8 (12.2) | 59.0 (13.2) | 59.8 (11.9) | 61.1 (12.9) |
| Male sex - n (%) | 656 (68.9) | 326 (67.2) | 269 (72.1) | 285 (70.2) |
| Race / Ethnicity <sup>a</sup> - n / N (%) |  |  |  |  |
| White | 515 / 724 (71.1) | 368 / 453<br>(81.2) | 184 / 254<br>(72.4) | 234 / 317<br>(73.8) |
| Asian | 123 / 724 (17.0) | 53 / 453 (11.7) | 39 / 254 (15.4) | 53 / 317 (16.7) |
| Black | 38 / 724 (5.2) | 9 / 453 (2.0) | 9 / 254 (3.5) | 10 / 317 (3.2) |
| Mixed | 13 / 724 (1.8) | 1 / 453 (0.2) | 6 / 254 (2.4) | 6 / 317 (1.9) |
| Other | 35 / 724 (4.8) | 22 / 453 (4.9) | 16 / 254 (6.3) | 14 / 317 (4.4) |
| Body-mass index <sup>b</sup> - median (IQR),<br>kg/m<sup>2</sup> | 30.4 (26.6 -<br>34.9)<br>(n = 862) | 31.2 (27.7 -<br>36.3)<br>(n = 419) | 29.7 (26.3 -<br>35.3)<br>(n = 332) | 30.9 (27.1 -<br>34.9)<br>(n = 385) |
| APACHE II score <sup>c</sup> - median (IQR) | 13.0 (8.0 - 19.0)<br>(n = 934) | 12.0 (7.0 -<br>20.0)<br>(n = 475) | 13.0 (8.0 -<br>19.0)<br>(n = 365) | 12.0 (8.0 -<br>18.0)<br>(n = 394) |
| Confirmed SARS-CoV-2 infection <sup>d</sup> - n /<br>N (%) | 802 / 942 (85.1) | 429 / 484<br>(88.6) | 319 / 369<br>(86.4) | 348 / 406<br>(85.7) |
| Preexisting condition - n / N (%) |  |  |  |  |
| Diabetes | 281 / 949 (29.6) | 108 / 484<br>(22.3) | 125 / 370<br>(33.8) | 152 / 406<br>(37.4) |
| Respiratory disease | 218 / 949 (23.0) | 117 / 484<br>(24.2) | 81 / 370 (21.9) | 100 / 406<br>(24.6) |
| Asthma/COPD | 183 / 949 (19.3) | 99 / 484 (20.5) | 67 / 370 (18.1) | 89 / 406 (21.9) |
| Other | 40 / 949 (4.2) | 26 / 484 (5.4) | 17 / 370 (4.6) | 17 / 406 (4.2) |
| Kidney disease | 66 / 866 (3.2) | 30 / 446 (1.5) | 22 / 340 (1.1) | 43 / 377 (2.1) |
| Severe cardiovascular disease | 86 / 930 (9.2) | 33 / 474 (7.0) | 41 / 367 (11.2) | 47 / 401 (11.7) |
| Any immunosuppressive condition | 25 / 948 (2.6) | 11 / 484 (2.3) | 6 / 370 (1.6) | 18 / 406 (4.4) |
| Cancer | 7 / 948 (0.7) | 3 / 484 (0.6) | 3 / 370 (0.8) | 10 / 406 (2.5) |
| Chronic immunosuppressive<br>therapy | 10 / 949 (1.1) | 8 / 484 (1.7) | 4 / 370 (1.1) | 7 / 406 (1.7) |
| Other | 13 / 948 (1.4) | 2 / 484 (0.4) | 3 / 370 (0.8) | 5 / 406 (1.2) |
| Liver cirrhosis / failure | 3 / 930 (0.3) | 0 / 474 (0.0) | 1 / 370 (0.3) | 2 / 401 (0.5) |
| Time to enrollment - median (IQR) |  |  |  |  |
| From hospital admission - days | 1.4 (0.9 - 3.3) | 1.6 (0.9 - 3.5) | 1.6 (0.9 - 3.8) | 1.2 (0.8 - 2.8) |
| From ICU admission - hours | 13.4 (6.9 - 19.1) | 15.1 (7.8 -<br>19.9) | 13.6 (7.3 -<br>19.7) | 14.0 (6.8 -<br>19.5) |
| Acute respiratory support - n (%) |  |  |  |  |
| None / supplemental oxygen only | 1 (0.1) | 0 (0.0) | 1 (0.3) | 2 (0.5) |
| High-flow nasal cannula | 226 (23.7) | 96 (19.8) | 101 (27.1) | 110 (27.1) |
| Noninvasive ventilation only | 404 (42.4) | 241 (49.7) | 133 (35.7) | 171 (42.1) |
| Invasive mechanical ventilation | 320 (33.6) | 148 (30.5) | 138 (37.0) | 122 (30.0) |

**Table 1. Participant Characteristics at Baseline (Severe State)\***
|  | **Tocilizumab**<br>**(n = 952)** | **Sarilumab**<br>**(n = 485)** | **Anakinra**<br>**(n = 373)** | **Control**<br>**(n = 406)** |
| --- | --- | --- | --- | --- |
| ECMO | 1 (0.1) | 0 (0.0) | 0 (0.0) | 1 (0.2) |
| Vasopressor support - n (%) | 179 (18.8) | 77 (15.9) | 81 (21.7) | 79 (19.5) |
| PaO<sub>2</sub> / FiO<sub>2</sub> - median (IQR) | 110 (86 - 148)<br>(n = 872) | 116 (89 - 152)<br>(n = 430) | 106 (84 - 148)<br>(n = 330) | 118 (89 - 169.5)<br>(n = 359) |
| Median laboratory values (IQR) <sup>f</sup> |  |  |  |  |
| C-reactive protein, µg/mL | 132 (69 - 201)<br>(n = 783) | 120 (70 - 199)<br>(n = 419) | 112 (70 - 189)<br>(n = 324) | 129 (71 - 208)<br>(n = 255) |
| D-dimer, µg/L | 946 (483 - 2475)<br>(n = 564) | 947 (420 - 2216)<br>(n = 304) | 1006 (460 - 2363)<br>(n = 256) | 1010 (500 - 2115)<br>(n = 175) |
| Received therapies at randomization -<br>n / N (%) |  |  |  |  |
| Steroids | 770 / 938 (82.1) | 422 / 472<br>(89.4) | 317 / 369<br>(85.9) | 269 / 402<br>(66.9) |
| Remdesivir | 272 / 938 (29.0) | 140 / 472<br>(29.7) | 109 / 369<br>(29.5) | 105 / 402<br>(26.1) |

\* Percentages may not sum to 100 because of rounding. SD denotes standard deviation; APACHE, Acute Physiology and Chronic Health Evaluation; SARS-CoV-2, Severe Acute Respiratory Syndrome Coronavirus; ICU, intensive care unit; COPD, chronic obstructive pulmonary disease; IQR, interquartile range; ECMO, extracorporeal membrane oxygenation.
a Data collection not approved in Canada and continental Europe. "Other" includes "declined" and "multiple".
b Body-mass index is the weight in kilograms divided by the square of the height in meters.
c Range 0 to 71, with higher scores indicating greater severity of illness.
d SARS-CoV2 infection was confirmed by respiratory tract polymerase chain reaction test.
e Range 3 to 15, with higher scores indicating greater consciousness, using values closest to randomization but prior to the use of sedative agents.
f Values were from the sample collected closest to randomization, up to 8 hours prior to randomization. If no samples were collected up to 8 hours prior to time of randomization, the sample collected closest to the time of randomization up to 2 hours after randomization was used (other than PaO<sub>2</sub> / FiO<sub>2</sub> which was a pre-randomization value only)

### Primary Outcome

Median organ support-free days were 7 (interquartile range [IQR] –1, 16), 9 (IQR –1, 17), 0 (IQR –1, 15) for tocilizumab, sarilumab and anakinra compared to 0 (IQR –1, 15) for the control group (**Table 2** and **Figure 2**). Compared with control, median adjusted odds ratios for organ support-free days (primary model) were 1.46 (95%CrI 1.13, 1.87) for tocilizumab, 1.50 (95%CrI 1.13, 2.00) for sarilumab and 0.99 (95%CrI 0.74, 1.35) for anakinra, yielding 99.8%, 99.8% and 46.6% posterior probabilities of superiority to control respectively. The posterior probability of equivalence between tocilizumab and sarilumab was 92.1% at the adaptive analysis, triggering the platform conclusion. Once all participants had completed follow-up, the probability of equivalence dropped to 84.9%, though not because sarilumab appeared less favorable. The probability that each intervention was optimal (the best in the domain) was 28% for tocilizumab and 46.5% for sarilumab, and the probability that sarilumab was non-inferior to tocilizumab was 98.9% (**Supplementary appendix Figure S2 and S3**). The probability that anakinra was optimal was 0.02%, triggering the inferiority threshold of <0.33%.

**Table 2.** Primary and Secondary Outcomes.

| Outcome/Analysis | Tocilizumab<br>(N=943) | Sarilumab<br>(N=483) | Anakinra<br>(N=365) | Control<br>(N=406) |
| --- | --- | --- | --- | --- |
| **Primary outcome** |  |  |  |  |
| Median (IQR) | 7 (–1 to 16) | 9 (–1 to 17) | 0 (–1 to 15) | 0 (–1 to 15) |
| Adjusted OR - mean (SD) | 1.47 (0.19) | 1.52 (0.22) | 1.00 (0.16) | 1 |
| - median (95% CrI) | 1.46 (1.13 to 1.87) | 1.50 (1.13 to 2.00) | 0.99 (0.74 to 1.35) | 1 |
| Probability of superiority to control, % | 99.8 | 99.8 | 46.6 | - |
| Probability of equivalence<br>(tocilizumab/sarilumab), % | 84.9 |  |  | - |
| Probability of being optimal in this domain, % * | 28.0 | 46.5 | 0.02 |  |
| **Subcomponents of primary outcome** |  |  |  |  |
| In-hospital deaths, n (%) | 317/943 (33.6) | 158/483 (32.7) | 145/365 (39.7) | 150/406 (36.9) |
| Days free from organ support in survivors,<br>median (IQR) | 15 (7.25 to 18) | 15 (9 to 18) | 14 (3.75 to 18) | 13 (4 to 17) |
| **Primary hospital survival** |  |  |  |  |
| Adjusted OR - mean (SD) | 1.44 (0.23) | 1.54 (0.29) | 0.99 (0.19) | 1 |
| - median (95% CrI) | 1.42 (1.05 to 1.93) | 1.51 (1.06 to 2.20) | 0.97 (0.66 to 1.40) | 1 |
| Probability of superiority to control, % | 98.8 | 98.8 | 43.6 | - |
| **Secondary analysis of primary outcome** |  |  |  |  |
| Adjusted OR - mean (SD) | 1.50 (0.19) | 1.59 (0.22) | 1.07 (0.17) | 1 |
| - median (95% CrI) | 1.49 (1.16 to 1.91) | 1.57 (1.20 to 2.06) | 1.06 (0.78 to 1.44) | 1 |
| Probability of superiority to control, % | 99.9 | 99.9 | 64.6 | - |
| **Secondary analysis of hospital survival** |  |  |  |  |
| Adjusted OR - mean (SD) | 1.45 (0.23) | 1.59 (0.29) | 1.03 (0.20) | 1 |
| - median (95% CrI) | 1.43 (1.05 to 1.95) | 1.57 (1.10 to 2.22) | 1.01 (0.70 to 1.49) | 1 |
| Probability of superiority to control, % | 99.0 | 99.3 | 52.9 | - |

The primary analysis of organ support-free days (OSFD) and in-hospital mortality used data from all participants (Moderate and Severe State) enrolled in the trial who met COVID-19 criteria and were randomized within at least one domain, for whom the outcome was known (n=5852), adjusting for age, sex, time period, site, region, domain and intervention eligibility and intervention assignment.
Other analyses were restricted to n=3848 participants enrolled in the Immune Modulation domain and any domains that have ceased recruitment (Corticosteroid; COVID-19 Antiviral, Anticoagulation and Immunoglobulin domains), adjusting for age, sex, time period, site, region, domain and intervention eligibility and intervention assignment. Definitions of outcomes are provided in Methods and the study protocol. All models are structured such that a higher OR or HR is favorable.
IQR – Interquartile range; OR - odds ratio; SD - standard deviation; CrI - credible interval
\* The posterior probability that pooled IL-6ra (calculated from the secondary model) is optimal in the domain was 68.2%

**Figure 2.**
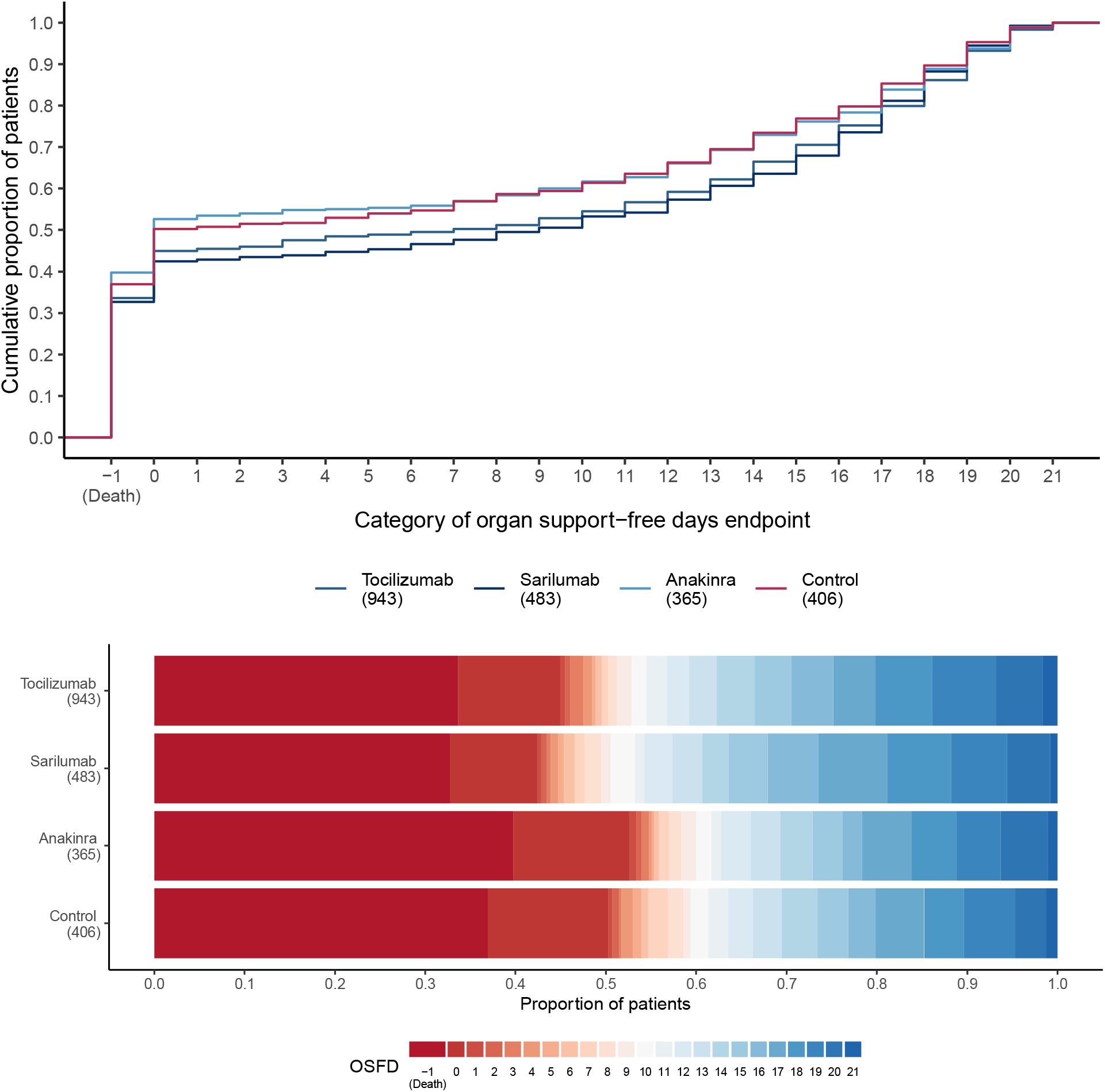
Distributions of organ support–free days. Panel A) shows the cumulative proportion (y-axis) for each intervention group by day (x-axis), with death listed first. Curves that rise more slowly indicate a more favorable distribution in the number of days alive and free of organ support. The height of each curve at “–1” indicates the in-hospital mortality rate for each intervention. The height of each curve at any point, for example, at day = 10, indicates the proportion of patients with organ support-free days (OSFD) of 10 or lower (i.e. 10 or worse). The difference in height of the two curves at any point represents the difference in the percentile in the distribution of OSFDs associated with that number of days alive and free of organ support. Panel B) shows organ support–free days as horizontally stacked proportions by intervention group. Red represents worse outcomes and blue represents better outcomes. The median adjusted odds ratios from the primary analysis, using a Bayesian cumulative logistic model, were 1.46 (95%CrI 1.13, 1.87) for tocilizumab, 1.50 (95%CrI 1.13, 2.00) for sarilumab, and 0.99 (95%CrI 0.74, 1.35) for anakinra, yielding 99.8%, 99.8% and 46.5% posterior probabilities of superiority, respectively, compared to control.

### Secondary Outcomes and Subgroup Analyses

Hospital survival rates were 66.4% for tocilizumab, 67.3% for sarilumab, 60.3% for anakinra, and 63.1%, for control. Compared with control, median adjusted odds ratios for hospital survival were 1.42 (95%CrI 1.05, 1.93) for tocilizumab, 1.51 (95%CrI 1.06, 2.20) for sarilumab and 0.97 (95%CrI 0.66, 1.40) for anakinra, yielding 98.8%, 98.8% and 43.6% respective posterior probabilities of the interventions being superior to control. Tocilizumab and sarilumab were both effective across all secondary outcomes, including 90-day survival, and both led to more rapid ICU and hospital discharge. (**Figure 3 and Supplementary appendix Table S4)**. The odds ratios for organ support-free days for tocilizumab and sarilumab were consistent across models with nested, independent, and pooled treatment effects (**Supplementary appendix Figure S4**). There was no evidence of any effect of anakinra in any of the secondary outcome analyses. The rates of serious adverse events were similar between all interventions (**Supplementary appendix Table S5**).

**Figure 3.**
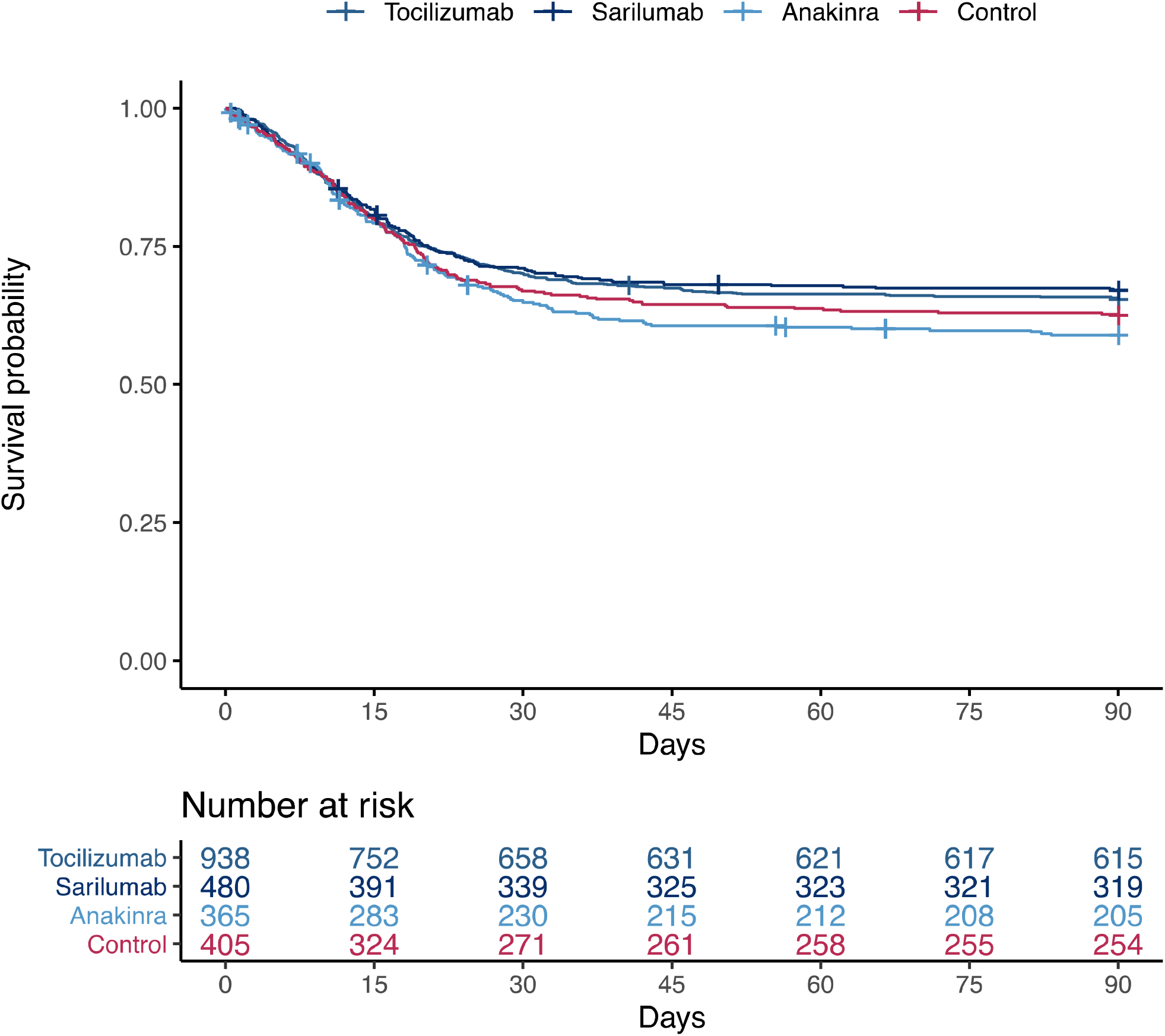
Time to event analysis. This plot is restricted to participants in the Severe State randomized to the Immune Modulation Therapy domain or another unblinded domain. Kaplan–Meier curve for survival up to 90 days according to individual interventions is shown. There were 323, 161, 160 and 151 deaths in the tocilizumab, sarilumab, anakinra and control groups respectively. This resulted in a hazard ratio of 1.39 (95%CrI 1.11 to 1.74) for tocilizumab, 1.44 (95%CrI 1.11 to 1.89) for sarilumab and 1.13 (95%CrI 0.87 to 1.49) for anakinra, yielding 99.9%, 99,6% and 82.3% respective posterior probabilities of superiority to control. These “survival hazard ratios” are defined as the reciprocal of the mortality hazard ratio to be consistent with the convention that odds ratios and hazard ratios >1 imply benefit.

The effects of both tocilizumab and sarilumab were similar for participants who were and were not invasively mechanically ventilated, and across CRP terciles. Subgroup results for anakinra showed no beneficial effect (**Supplementary appendix Figure S5 and S6**). The sensitivity analyses on the primary outcome were consistent with the primary analysis (**Supplementary appendix, Table S3; Appendix 2 and 3**).

## Discussion

In critically ill patients with COVID-19, tocilizumab and sarilumab are both effective and likely equivalent in improving survival and reducing duration of organ support. Benefits of IL-6ra are consistent across primary and secondary outcomes, and across subgroups and secondary analyses. Anakinra is inferior to IL-6ra and no more effective than control.

REMAP-CAP has previously reported the efficacy of tocilizumab and sarilumab compared to standard of care in critically ill patients.^4^ The RECOVERY trial similarly showed the effectiveness of tocilizumab in a broader group of hospitalized patients.^22^ There has been less evidence about the efficacy of sarilumab. We report that the two agents are equally effective when compared directly. The comparison met the trial criteria for equivalence with a posterior probability of >0.90 at the time of a planned adaptive analysis; it fell below that threshold as full data became available only because of a minimal increase in the likelihood that sarilumab was superior to tocilizumab. In the face of an urgent global need and potential limitations of drug supply, the two agents can be considered equally effective, and this can help ensure as many patients as possible receive effective treatments.

We did not observe a beneficial effect of anakinra in critically ill patients with COVID-19. Previous studies and a recent meta-analysis provided a rationale for the use of anakinra in COVID-19. ^23^ Anakinra treatment using soluble urokinase plasminogen activator receptor (sUPAR) to identify an at-risk group of patients for early targeting of IL-1 was effective in moderate COVID-19 in a recent study.^24^ There are several possible explanations for our findings. Firstly, we could have chosen the wrong dose. However, our choice of administration regimen was informed by pharmacometric modelling data, and it is unlikely that predicted drug concentrations varied significantly. Secondly, blocking IL-1 (and IL-1 mediated IL-6 release) may benefit non-critically ill, but not critically ill patients. Such potential differential effects could not be assessed in this study as a result of the low recruitment in non-critically ill patients. Finally, we did not use a strategy of early targeting of the IL-1 pathway in selected patients as used in the SAVE-MORE trial,^24^ because sUPAR is not commonly available.

REMAP-CAP’s pragmatic, international design means that our results are likely generalizable to the wider critically ill patient population with COVID-19. Importantly, even once we demonstrated individual benefits of tocilizumab and sarilumab, the adaptive design allowed continued randomization to evaluate the comparative effectiveness of these two interventions, as well as to evaluate other immune modulation therapies. This allowed maximal learning about treatment effects while improving standard care, having removed the less effective control group from further assignment.^25^

The trial has limitations. It uses an open-label design, although awareness of intervention assignment is unlikely to affect the mortality component of the primary outcome. The effect of interferon-β1a could not be assessed with only 21 patients randomized to this intervention, as the use of corticosteroids excluded patients from this treatment. Interferon-β1a did not improve clinical outcomes in the SOLIDARITY Trial.^26^

In conclusion, in adult patients with COVID-19 receiving organ support in intensive care, the IL-6 receptor antagonists, tocilizumab and sarilumab, are similarly effective at improving survival and reducing duration of organ support. Anakinra is not effective in this population.

## Supporting information

Appendix 1 Statistical Analysis Plan

Appendix 2 Primary analysis

Appendix 3 Secondary analysis

## Data Availability

Data will be available to researchers on request subject to sponsor restrictions, please contact

http://www.remapcap.org

## Data Availability

Data will be available to researchers on request subject to sponsor restrictions, please contact

http://www.remapcap.org

## Acknowledgements

We are grateful to the NIHR Clinical Research Network (UK), UPMC Health System Health Services Division (US), and the Direction de la Recherche Clinique et de l’Innovation de l’AP-HP (France) for their support of participant recruitment. We are grateful for the supply of study drugs in the United Kingdom and Europe from Roche Products Ltd, Sanofi (Aventis Pharma Ltd), Swedish Orphan Biovitrum AB (Sobi™) and Faron Pharmaceuticals

