## Supplementary Appendix for "Effectiveness of Tocilizumab, Sarilumab, and Anakinra for critically ill patients with COVID-19 The REMAP-CAP COVID-19 Immune Modulation Therapy Domain Randomized Clinical Trial"

### 1. REMAP-CAP investigators

1.1. Immune Modulation Therapy domain Writing Committee

Lennie P.G. Derde, MD, PhD ^1^; Anthony C. Gordon, MBBS, MD ^2,3^; Paul R. Mouncey, MSc ^4^; Farah Al-

Beidh, PhD ^2^; Kathryn M. Rowan, PhD ^4^; Alistair D. Nichol, MD, PhD ^5,6,7^; Yaseen M. Arabi, MD ^8^; Djillali Annane, MD, PhD ^9,10,11^; Abi Beane, PhD ^12^; Richard Beasley ^13^; Zahra Bhimani, MPH ^14^; Marc J.M. Bonten, MD, PhD ^1^; Charlotte A. Bradbury, MBChB, PhD ^15^; Frank M. Brunkhorst, MD, PhD ^16^; Adrian Buzgau, MSc ^5^; Meredith Buxton ^17^; Allen C. Cheng, MD, PhD ^5,18^; Nicola Cooper ^2,3^; Matt Cove ^19^; Olaf L. Cremer, MD, PhD ^1^; Michelle A. Detry, PhD ^20^; Eamon J. Duffy, BPharm ^21^; Lise J. Estcourt, MBBCh, PhD ^22^; Mark Fitzgerald, PhD ^20^; James Galea ^23^; Herman Goossens, PhD ^24^; Rashan Haniffa, PhD ^25,26,27^; Thomas E. Hills, PhD ^13,28^; Nao Ichihara ^29^; Andrew King ^23^; Francois Lamontagne, MD ^30^; Patrick R. Lawler, MD, MPH ^31,32^; Helen L. Leavis, MD, PhD ^1;^ Roger J. Lewis, MD, PhD ^20,33^; Kelsey M. Linstrum, MS ^34^; Edward Litton, MD, PhD ^35,36^; John C. Marshall, MD ^14^; Florian B. Mayr, MD, MPH ^34^; Danny McAuley, MD ^37,38^; Anna McGlothlin, PhD ^20^; Shay P McGuinness, MD ^5,21,28^; Bryan J. McVerry, MD ^34^; Stephanie K. Montgomery, MSc ^34^; Susan C. Morpeth, MD, PhD ^28,39^; Srinivas Murthy, MD ^40^; Mihai G. Netea ^41^; Kayode Ogungbenro ^23^; Katrina Orr, BPharm ^42^; Rachael L. Parke, PhD ^13,43^; Jane C. Parker, BN ^5^; Asad E. Patanwala, PharmD, MPH ^44,45^; Ville Pettila, MD ^46^; Luis Felipe Reyes ^47,48^; Ashish Sanil ^20^; Hiroki Saito ^49^; Marlene S. Santos, MD, MSHS ^14^; Christina T. Saunders, PhD ^20^; Manoj Saxena ^50^; Christopher W. Seymour, MD, MSc ^34^; Manu Shankar-Hari, MD, PhD ^51,52^; Wendy I. Sligl, MD, MSc ^53^; Alexis F. Turgeon, MD, MSc ^54^; Anne M. Turner, MPH ^13^; Steven Tong ^55^; Suvi Vaara ^46^; Taryn Youngstein ^2,3^; Ryan Zarychanski, MD, MSc ^56^; Cameron Green, MSc ^5^; Alisa M. Higgins, PhD ^5^; Colin J. McArthur, MD ^28^; Lindsay R. Berry, PhD ^20^; Elizabeth Lorenzi, PhD ^20^; Scott Berry, PhD ^20^; Steve A. Webb, MD, PhD ^5,57^; Derek C. Angus, MD, MPH ^34^; Frank L. van de Veerdonk, MD, PhD ^41^.

1. University Medical Center Utrecht, Utrecht, The Netherlands
2. Imperial College London, London, United Kingdom
3. Imperial College Healthcare NHS Trust, St. Mary's Hospital, London, United Kingdom
4. Intensive Care National Audit & Research Centre (ICNARC), London, United Kingdom
5. Monash University, Melbourne, Australia
6. University College Dublin, Dublin, Ireland
7. Alfred Health, Melbourne, Australia
8. King Saud bin Abdulaziz University for Health Sciences and King Abdullah International Medical Research Center, Riyadh, Kingdom of Saudi Arabia
9. Hospital Raymond Poincaré (Assistance Publique Hópitaux de Paris), Garches, France
10. Université Versailles SQY - Université Paris Saclay, Montigny-le-Bretonneux, France
11. Université Paris Saclay - UVSQ – INSERM, Garches, France
12. University of Oxford, Oxford, United Kingdom
13. Medical Research Institute of New Zealand (MRINZ), Wellington, New Zealand
14. St. Michael's Hospital Unity Health, Toronto, Canada
15. University of Bristol, Bristol, United Kingdom
16. Jena University Hospital, Jena, Germany
17. Global Coalition for Adaptive Research, San Francisco, California
18. Alfred Health, Melbourne, Australia
19. National University of Singapore
20. Berry Consultants, Austin, United States
21. Auckland District Health Board, Auckland, New Zealand
22. NHS Blood and Transplant, Oxford, United Kingdom
23. University of Manchester, Manchester, United Kingdom
24. University of Antwerp, Wilrijk, Belgium
25. University of Oxford, Bangkok, Thailand
26. University College London Hospital, London, United Kingdom
27. National Intensive Care Surveillance (NICST), Colombo, Sri Lanka
28. Auckland City Hospital, Auckland, New Zealand
29. Graduate School of Medicine, The University of Tokyo, Tokyo, Japan
30. Université de Sherbrooke, Sherbrooke, Quebec, Canada
31. University Health Network, Toronto, Canada
32. University of Toronto, Toronto, Canada
33. Harbor-UCLA Medical Center, Torrance, CA, United States
34. University of Pittsburgh, Pittsburgh, United States
35. Fiona Stanley Hospital, Perth, Australia
36. University of Western Australia, Perth, Australia
37. Queen’s University Belfast, Belfast, Northern Ireland
38. Royal Victoria Hospital, Belfast, Northern Ireland
39. Middlemore Hospital, Auckland, New Zealand
40. University of British Columbia, Vancouver, Canada
41. Radboudumc, Nijmegen, The Netherlands
42. Fiona Stanley Hospital, Perth, Australia
43. University of Auckland, Auckland, New Zealand
44. University of Sydney, Sydney, Australia
45. Royal Prince Alfred Hospital, Sydney, Australia
46. University of Helsinki and Helsinki University Hospital, Helsinki, Finland
47. Universidad de La Sabana, Chia, Colombia
48. Clínica Universidad de La Sabana, Chia, Colombia
49. Yokohama City Seibu Hospital, Yokohama
50. The George Institute for Global Health, Sydney, Australia
51. King's College London, London, United Kingdom
52. Guy's and St Thomas' NHS Foundation Trust, London, United Kingdom
53. University of Alberta, Edmonton, Canada
54. Université Laval, Québec City, Canada
55. Peter Doherty Institute for Infection and Immunity, Melbourne, Australia
56. University of Manitoba, Winnipeg, Canada
57. St John of God Hospital, Subiaco, Australia
    1.2. REMAP-CAP Trial Investigators and Collaborators

**International Trial Steering Committee:**

Farah Al-Beidh, Derek Angus, Djillali Annane, Yaseen Arabi, Abi Beane, Wilma van Bentum-Puijk, Scott Berry, Zahra Bhimani, Marc Bonten, Charlotte Bradbury, Frank Brunkhorst, Meredith Buxton, Allen Cheng, Lennie Derde, Lise Estcourt, Herman Goossens, Anthony Gordon, Cameron Green, Rashan Haniffa, Francois Lamontagne, Patrick Lawler, Kelsey Linstrum, Edward Litton, John Marshall, Colin McArthur, Daniel McAuley, Shay McGuinness, Bryan McVerry, Stephanie Montgomery, Paul Mouncey, Srinivas Murthy, Alistair Nichol, Rachael Parke, Jane Parker, Kathryn Rowan, Marlene Santos, Christopher Seymour, Manu Shankar-Hari, Alexis Turgeon, Anne Turner, Frank van de Veerdonk, Steve Webb (Chair), Ryan Zarychanski

**Regional Management Committees**

***Australia and New Zealand Regional Management Committee***

Yaseen Arabi, Lewis Campbell, Allen Cheng, Lennie Derde, Andrew Forbes, David Gattas, Cameron Green, Stephane Heritier, Peter Kruger, Edward Litton, Colin McArthur (Deputy Executive Director), Shay McGuinness (Chair), Alistair Nichol, Rachael Parke, Jane Parker, Sandra Peake, Jeffrey Presneill, Ian Seppelt, Tony Trapani, Anne Turner, Steve Webb (Executive Director), Paul Young

***Canadian Regional Management Committee***

Zahra Bhimani, Brian Cuthbertson, Rob Fowler, Francois Lamontagne, John Marshall (Executive Director), Venika Manoharan, Srinivas Murthy (Deputy Executive Director), Marlene Santos, Alexis Turgeon, Ryan Zarychanski

***Critical Care Asia Regional Management Committee***

Diptesh Aryal, Abi Beane (Chair), Arjen M Dondrop, Cameron Green, Rashan Haniffa (Executive Director), Madiha Hashmi, Deva Jayakumar, John Marshall, Colin McArthur, Srinivas Murthy, Timo Tolppa, Vanessa Singh, Steve Webb

***European Regional Management Committee***

Farah Al-Beidh, Derek Angus, Djillali Annane, Wilma van Bentum-Puijk, Scott Berry, Marc Bonten (Executive Director), Nicole Brillinger, Frank Brunkhorst, Maurizio Cecconi, Lennie Derde (Chair), Stephan Ermann, Bruno Francois, Herman Goossens, Anthony Gordon, Cameron Green, Sebastiaan Hullegie, Rene Markgraff, Colin McArthur, Paul Mouncey, Alistair Nichol, Mathias Pletz, Pedro Povoa, Gernot Rohde, Kathryn Rowan, Lorraine Parker, Irma Scheepstra-Beukers, Steve Webb

***United States Regional Management Committee***

Brian Alexander, Derek Angus (Executive Director), Kim Basile, Meredith Buxton (Chair), Timothy Girard, Christopher Horvat, David Huang, Kelsey Linstrum, Florian Mayr, Bryan McVerry, Stephanie Montgomery, Christopher Seymour

**Regional Coordinating Centers**

***Australia, CCA region, and Saudi Arabia:*** The Australia and New Zealand Intensive Care Research Centre (ANZIC-RC), Monash University

***Canada:*** St. Michael’s Hospital, Unity Health Toronto

***Europe:*** University Medical Center Utrecht (UMCU)

***New Zealand:*** The Medical Research Institute of New Zealand (MRINZ)

***United States:*** Global Coalition for Adaptive Research (GCAR), and University of Pittsburgh Medical Center

***Critical Care Asia:*** NICS MORU.

**Domain-Specific Working Groups**

***Antibiotic and Macrolide Duration Domain-Specific Working Group***

Richard Beasley, Marc Bonten, Allen Cheng (Chair), Nick Daneman, Lennie Derde, Robert Fowler, David Gattas, Anthony Gordon, Cameron Green, Peter Kruger, Colin McArthur, Steve McGloughlin, Susan Morpeth, Srinivas Murthy, Alistair Nichol, Mathias Pletz, David Paterson, Gernot Rohde, Steve Webb

***Corticosteroid Domain-Specific Working Group***

Derek Angus (Chair), Wilma van Bentum-Puijk, Lennie Derde, Anthony Gordon, Sebastiaan Hullegie, Peter Kruger, Edward Litton, John Marshall, Colin McArthur, Srinivas Murthy, Alistair Nichol, Bala Venkatesh, Steve Webb

***Influenza Antiviral Domain-Specific Working Group***

Derek Angus, Scott Berry, Marc Bonten, Allen Cheng, Lennie Derde, Herman Goossens, Sebastiaan Hullegie, Menno de Jong, John Marshall, Colin McArthur, Srinivas Murthy (Chair), Tim Uyeki, Steve Webb

***COVID-19 Antiviral Domain-Specific Working Group***

Derek Angus, Yaseen Arabi (Chair), Kenneth Baillie, Richard Beasley, Scott Berry, Marc Bonten, Allen Cheng, Menno de Jong, Lennie Derde, Eamon Duffy, Rob Fowler, Herman Goossens, Anthony Gordon, Cameron Green, Thomas Hills, Colin McArthur, Susan Morpeth, Srinivas Murthy, Alistair Nichol, Katrina Orr, Rachael Parke, Jane Parker, Asad Patanwala, Kathryn Rowan, Steven Tong, Tim Uyeki, Frank van de Veerdonk, Steve Webb

***COVID-19 Immune Modulation Domain-Specific Working Group***

Derek Angus, Yaseen Arabi, Kenneth Baillie, Richard Beasley, Scott Berry, Marc Bonten, Frank Brunkhorst, Allen Cheng, Nichola Cooper, Olaf Cremer, Menno de Jong, Lennie Derde (Chair), Eamon Duffy, James Galea, Herman Goossens, Anthony Gordon, Cameron Green, Thomas Hills, Andrew King, Helen Leavis, John Marshall, Florian Mayr, Colin McArthur, Bryan McVerry, Susan Morpeth, Srinivas Murthy, Mihai Netea, Alistair Nichol, Kayode Ogungbenro, Katrina Orr, Jane Parker, Asad Patawala, Ville Pettilä (Deputy Chair), Kathryn Rowan, Manoj Saxena, Christopher Seymour, Wendy Sligl, Steven Tong, Tim Uyeki, Suvi Vaara, Frank van de Veerdonk, Steve Webb, Taryn Youngstein

***COVID-19 Immune Modulation-2 Domain-Specific Working Group***

Derek Angus, Scott Berry, Lennie Derde, Cameron Green, David Huang, Florian Mayr, Bryan McVerry, Stephanie Montgomery, Christopher W. Seymour (Chair), Steve Webb

***Therapeutic Anticoagulation Domain-Specific Working Group***

Derek Angus, Diptesh Aryal, Scott Berry, Shailesh Bihari, Charlotte Bradbury, Marc Carrier, Dean Fergusson, Robert Fowler, Ewan Goligher (Deputy Chair), Anthony Gordon, Christopher Horvat, David Huang, Beverley Hunt, Devachandran Jayakumar, Anand Kumar, Mike Laffan, Patrick Lawler, Sylvain Lother, Colin McArthur, Bryan McVerry, John Marshall, Saskia Middeldorp, Zoe McQuilten, Matthew Neal, Alistair Nichol, Christopher Seymour, Roger Schutgens, Simon Stanworth, Alexis Turgeon, Steve Webb, Ryan Zarychanski (Chair)

***Vitamin C Domain-Specific Working Group***

Neill Adhikari (Chair), Derek Angus, Djillali Annane, Matthew Anstey, Yaseen Arabi, Scott Berry, Emily Brant, Angelique de Man, Lennie Derde, Anthony Gordon, Cameron Green, David Huang, Francois Lamonagne (Chair), Edward Litton, John Marshall, Marie-Helene Masse, Colin McArthur, Shay McGuinness, Paul Mouncey, Srinivas Murthy, Rachael Parke, Alistair Nichol, Tony Trapani, Andrew Udy, Steve Webb

***COVID-19 Immunoglobulin Domain-Specific Working Group***

Derek Angus, Donald Arnold, Phillipe Begin, Scott Berry, Richard Charlewood, Michael Chasse, Mark Coyne, Jamie Cooper, James Daly, Lise Estcourt (Chair, UK lead), Dean Fergusson, Anthony Gordon, Iain Gosbell, Heli Harvala-Simmonds, Tom Hills (New Zealand lead), Christopher Horvat, David Huang, Sheila MacLennan, John Marshall, Colin McArthur (New Zealand lead), Bryan McVerry (USA lead), David Menon, Susan Morpeth, Paul Mouncey, Srinivas Murthy, John McDyer, Zoe McQuilten (Australia lead), Alistair Nichol (Ireland lead), Nicole Pridee, David Roberts, Kathryn Rowan, Christopher Seymour, Manu Shankar-Hari (UK lead), Helen Thomas, Alan Tinmouth, Darrell Triulzi, Alexis Turgeon (Canada lead), Tim Walsh, Steve Webb, Erica Wood, Ryan Zarychanski (Canada lead)

***Simvastatin Domain-Specific Working Group***

Derek Angus, Yaseen Arabi, Abi Beane, Carolyn Calfee, Anthony Gordon, Cameron Green, Rashan Haniffa, Deva Jayakumar, Peter Kruger, Patrick Lawler, Edward Litton, Colin McArthur, Daniel McAuley (Chair), Bryan McVerry, Matthew Neal, Alistair Nichol, Cecilia O’Kane, Murali Shyamsundar, Pratik Sinha, Taylor Thompson, Steve Webb, Ian Young

***Antiplatelet Domain-Specific Working Group***

Derek Angus, Scott Berry, Shailesh Bihari, Charlotte Bradbury (Chair), Marc Carrier, Timothy Girard, Ewan Goligher, Anthony Gordon, Ghady Haidar, Christopher Horvat, David Huang, Beverley Hunt, Anand Kumar, Patrick Lawler, Patrick Lawless, Colin McArthur, Bryan McVerry, John Marshall, Zoe McQuilten, Matthew Neal, Alistair Nichol, Christopher Seymour, Simon Stanworth, Steve Webb, Alexandra Weissman, Ryan Zarychanski

***Mechanical Ventilation Domain***

Derek Angus, Wilma van Bentum-Puijk, Lewis Campbell, Lennie Derde, Niall Ferguson, Timothy Girard, Ewan Goligher, Anthony Gordon, Cameron Green, Carol Hodgson, Peter Kruger, John Laffey, Edward Litton, John Marshall, Colin McArthur, Daniel McAuley, Shay McGuinness, Alistair Nichol (Chair) Neil Orford, Kathryn Rowan, Ary Neto, Steve Webb

***ACE-2 RAS Domain***

Rebecca Baron, Lennie Derde, Slava Epelman, Claudia Frankfurter, David Gattas, Frank Gommans, Anthony Gordon, Rashan Haniffa, David Huang, Edy Kim, Francois Lamontagne, Patrick Lawler (Chair), David Leaf, John Marshall, Colin McArthur, Bryan McVerry, Daniel McAuley, Muthiah Vaduganathan, Roland van Kimmenade, Frank van de Veerdonk, Steve Webb

**Statistical Analysis Committee**

Michelle Detry, Mark Fitzgerald, Roger Lewis (Chair), Anna McGlothlin, Ashish Sanil, Christina Saunders

**Statistical Design Team**

Lindsay Berry, Scott Berry, Elizabeth Lorenzi

**Data Coordinating Team**

Adrian Buzgau, Cameron Green, Alisa Higgins

**Project Management**

***Australia and Saudi Arabia:*** Jane Parker, Vanessa Singh, Claire Zammit

***Canada:*** Zahra Bhimani, Marlene Santos

***CCA:*** Abi Beane, Rashan Haniffa, Timo Tolppa

***Europe:*** Wilma van Bentum-Puijk, Lorraine Parker, Irma Scheepstra-Beukers, Erika Groeneveld, Svenja Peters, Clementina Okundaye, Denise van Hout, Albertine Smit, Linda Rikkert, Sara Bari, Kik Raymakers, Marion Kwakkenbos-Craanen, Sophie Post, Gerwin Schreuder.

***Germany:*** Nicole Brillinger, Rene Markgraf

***Global:*** Cameron Green

***Ireland:*** Kate Ainscough, Kathy Brickell, Peter Doran, Patrick Murray

***New Zealand:*** Anne Turner

***United Kingdom:*** Farah Al-Beidh, Aisha Anjum, Janis-Best Lane, Elizabeth Fagbodun, Lorna Miller, Paul Mouncey, Karen Parry-Billings, Sam Peters, Alvin Richards-Belle, Michelle Saull, Stefan Sprinckmoller, Daisy Wiley

***United States of America:*** Kim Basile, Meredith Buxton, Kelsey Linstrum, Stephanie Montgomery, Renee Wunderley

**Database Providers**

***Research Online:*** Marloes van Beurden, Evelien Effelaar, Joost Schotsman,

***Spinnaker Software:*** Craig Boyd, Cain Harland, Audrey Shearer, Jess Wren

***University of Pittsburgh Medical Center:*** Giles Clermont, William Garrard, Christopher Horvat, Kyle Kalchthaler, Andrew King, Daniel Ricketts, Salim Malakoutis, Oscar Marroquin, Edvin Music, Kevin Quinn

***NICS MORU:*** Udara Attanayaka, Abi Beane, Sri Darshana, Rashan Haniffa, Pramodya Ishani, Issrah Jawad, Upulee Pabasara, Timo Tolppa, Ishara Udayanga.

**Clinical Trials Groups**

The REMAP-CAP platform is supported by the Australian and New Zealand Intensive Care Society Clinical Trials Group, the Canadian Critical Care Clinical Trials Group, the UK Critical Care Research Group and the International Forum of Acute Care Trialists.

REMAP-CAP was supported in the UK by the NIHR Clinical Research Network and we acknowledge the contribution of Kate Gilmour, Karen Pearson, Chris Siewerski, Sally-Anne Hurford, Emma Marsh, Debbie Campbell, Penny Williams, Kim Shirley, Meg Logan, Jane Hanson, Anne Oliver, Mihaela Sutu, Sheenagh Murphy, Latha Aravindan, Joanne Collins, Holly Monaghan, Adam Unsworth, Seonaid Beddows, Laura Ann Dawson, Sarah Dyas, Adeeba Asghar, Kate Donaldson, Tabitha Skinner, Nhlanhla Mguni, Natasha Muzengi, Ji Luo, Joanna O’Reilly, Chris Levett, Alison Potter, David Porter, Teresa Lockett, Jazz Bartholomew, Clare Rook, Hannah Williams, Alistair S Hall, Hilary Campbell, Holly Speight, Sandra Halden, Susan Harrison, Mobeena Naz, Charles Rounds, Kaatje Lomme, Johnathan Sheffield, William Van’t Hoff, James D Williamson, Catherine Birch, Morwenna Brend, Emma Chambers, Sarah Crawshaw, Chelsea Drake, Heather Harper, Stephen Lock, Eleanor OKell, Amber Hayes, Susan Walker, Jayne Goodwin, Helen Hodgson, Yvette Ellis, Dawn Williamson, Madeleine Bayne, Shane Jackson, Rahim Byrne, Sonia McKenna, Alison Clinton, NIHR Urgent Public Health Group: https://www.nihr.ac.uk/documents/urgent-public-health-group-members/24638#Members

**Other supporting networks**

REMAP-CAP was supported in France by the CRICS-TRIGGERSEP network

REMAP-CAP was supported in Ireland by the Irish Critical Care Clinical Trials Group and we acknowledge the contribution of Kate Ainscough, Kathy Brickell and Peter Doran.

REMAP-CAP was supported in the Netherlands by the Research Collaboration Critical Care the Netherlands (RCC-Net).

REMAP-CAP was supported in Canada but the Canadian Institutes of Health Research and St. Michael’s Unity Health

**Site Investigators and Research Coordinators**

***Australia:***

*The Alfred Hospital*: Andrew Udy, Phoebe McCracken, Meredith Young, Jasmin Board, Emma Martin;

*Ballarat Health Services*: Khaled El-Khawas, Angus Richardson, Dianne Hill, Robert J Commons, Hussam Abdelkharim;

*Bendigo Hospital*: Cameron Knott, Julie Smith, Catherine Boschert;

*Caboolture Hospital*: Julia Affleck, Yogesh Apte, Umesh Subbanna, Roland Bartholdy, Thuy Frakking;

*Campbelltown Hospital*: Karuna Keat, Deepak Bhonagiri, Ritesh Sanghavi, Jodie Nema, Megan Ford;

*Canberra Hospital*: Harshel G. Parikh, Bronwyn Avard, Mary Nourse;

*Concord Repatriation General Hospital*: Winston Cheung, Mark Kol, Helen Wong, Asim Shah, Atul Wagh;

*Eastern Health (Box Hill, Maroondah & Angliss Hospitals)*: Joanna Simpson, Graeme Duke, Peter Chan, Brittney Carter, Stephanie Hunter;

*Flinders Medical Centre*: Shailesh Bihari, Russell D Laver, Tapaswi Shrestha, Xia Jin;

*Fiona Stanley Hospital*: Edward Litton, Adrian Regli, Susan Pellicano, Annamaria Palermo, Ege Eroglu;

*Footscray Hospital*: Craig French, Samantha Bates, Miriam Towns, Yang Yang, Forbes McGain;

*Gold Coast University Hospital*: James McCullough, Mandy Tallott;

*John Hunter Hospital*: Nikhil Kumar, Rakshit Panwar, Gail Brinkerhoff, Cassandra Koppen, Federica Cazzola;

*Launceston General Hospital*: Matthew Brain; Sarah Mineall;

*Lyell McEwin Hospital*: Roy Fischer, Vishwanath Biradar, Natalie Soar;

*Logan Hospital*: Hayden White, Kristen Estensen, Lynette Morrison, Joanne Sutton, Melanie Cooper;

*Monash Health (Monash Medical Centre, Dandenong Hospital & Casey Hospital):* Yahya Shehabi, Wisam Al-Bassam, Amanda Hulley; Umesh Kadam, Kushaharan Sathianathan;

*Nepean Hospital*: Ian Seppelt, Christina Whitehead, Julie Lowrey, Rebecca Gresham, Kristy Masters;

*Princess Alexandra Hospital*: Peter Kruger, James Walsham, Mr Jason Meyer, Meg Harward, Ellen Venz;

*The Prince Charles Hospital*: Kara Brady, Cassandra Vale, Kiran Shekar, Jayshree Lavana, Dinesh Parmar;

*The Queen Elizabeth Hospital*: Sandra Peake, Patricia Williams, Catherine Kurenda;

*Rockhampton Hospital*: Helen Miles, Antony Attokaran;

*Royal Adelaide Hospital*: Samuel Gluck, Stephanie O’Connor, Marianne Chapman, Kathleen Glasby;

*Royal Darwin Hospital*: Lewis Campbell, Kirsty Smyth, Margaret Phillips;

*Royal Melbourne Hospital*: Jeffrey Presneill, Deborah Barge, Kathleen Byrne, Alana Driscoll, Louise Fortune;

*Royal North Shore Hospital*: Pierre Janin, Elizabeth Yarad, Frances Bass, Naomi Hammond, Anne O’Connor;

*Royal Perth Hospital*: Sharon Waterson, Steve Webb, Robert McNamara;

*Royal Prince Alfred Hospital*: David Gattas, Heidi Buhr, Jennifer Coles;

*Sir Charles Gardiner Hospital*: Sacha Schweikert, Bradley Wibrow, Matthew Anstey, Rashmi Rauniyar;

*St George Hospital*: Kush Deshpande, Pam Konecny, Jennene Miller, Adeline Kintono, Raymond Tung

*St. John of God Midland Public and Private Hospitals*: Ed Fysh, Ashlish Dawda, Bhaumik Mevavala;

*St. John of God Hospital, Murdoch*: Annamaria Palermo, Adrian Regli, Bart De Keulenaer;

*St. John of God Hospital, Subiaco*: Ed Litton, Janet Ferrier;

*St. Vincent’s Hospital (NSW)*: Priya Nair, Hergen Buscher, Claire Reynolds, Sally Newman;

*St. Vincent’s Hospital (VIC)*: John Santamaria, Leanne Barbazza, Jennifer Homes, Roger Smith;

*Sunshine Coast University Hospital*: Peter Garrett, Lauren Murray, Jane Brailsford, Loretta Forbes, Teena Maguire;

*Sunshine Hospital*: Craig French, Gerard Fennessy, John Mulder, Rebecca Morgan, Rebecca McEldrew;

*The Sutherland Hospital*: Anas Naeem, Laura Fagan, Emily Ryan;

*Toowoomba Hospital*: Vasanth Mariappa, Judith Smith;

*University Hospital Geelong*: Scott Simpson, Matthew Maiden, Allison Bone, Michelle Horton, Tania Salerno;

*Wollongong Hospital*: Martin Sterba, Wenli Geng;

***Belgium:***

*Ghent University Hospital*: Pieter Depuydt, Jan De Waele, Liesbet De Bus, Jan Fierens, Stephanie Bracke, Joris Vermassen, Daisy Vermeiren;

***Canada*:**

*Brantford General Hospital:* Brenda Reeve, William Dechert;

Institut universitaire de cardiologie et de pneumologie de Québec: Francois Lellouche, Patricia Lizotte

*Centre Hospitalier de l’Universite de Montreal:* Michaël Chassé, François Martin Carrier, Dounia Boumahni, Fatna Benettaib, Ali Ghamraoui;

*CHU de Québec – Université Laval*: Alexis Turgeon, David Bellemare, Marie-Claude Boulanger, Ève Cloutier, Olivier Costerousse, Rana Daher, François Lauzier, Charles Francoeur;

*Centre Hospitalier Universitaire de Sherbrooke:* François Lamontagne, Frédérick D’Aragon, Elaine Carbonneau, Julie Leblond;

Grace Hospital: Gloria Vazquez-Grande, Nicole Marten

Grand River Hospital (Kitchener): Theresa Liu, Atif Siddiqui

*Health Sciences Centre, Winnipeg:* Ryan Zarychanski, Gloria Vazquez-Grande, Nicole Marten, RN, Maggie Wilson;

*Hôpital du Sacré Coeur de Montréal:* Martin Albert, Karim Serri, Alexandros Cavayas, Mathilde Duplaix, Virginie Williams;

*Juravinski Hospital:* Bram Rochwerg, Tim Karachi, Simon Oczkowski, John Centofanti, Tina Millen

McGill University Health Centre: Josie Campisi, Kosar Khwaja,

*Niagara Health (St. Catherine’s Hospital):* Erick Duan, Jennifer Tsang, Lisa Patterson;

*Regina General Hospital*: Eric Sy, Chiraag Gupta, Sandy Kassir, Jonathan Mailman, Stephen Lee;

*Royal Alexandra Hospital*: Demetrios Kutsogiannis, Patricia Thompson

*Sunnybrook Health Sciences Centre*: Rob Fowler, Neill Adhikari, Maneesha Kamra, Nicole Marinoff

*St. Boniface General Hospital*: Ryan Zarychanski, Nicole Marten

*St. Joseph’s Healthcare Hamilton*: Deborah Cook, Frances Clarke

*St. Mary's General Hospital (Kitchener)*: Rebecca Kruisselbrink, Atif Siddiqui

*St. Michael’s Hospital*: John Marshall, Laurent Brochard, MD, Karen Burns, MD, Gyan Sandhu, Imrana Khalid;

*The Ottawa Hospital:* Shane English, Irene Watpool, Rebecca Porteous, Sydney Miezitis, Lauralyn McIntyre;

*University Health Network:* Elizabeth Wilcox, Lorenzo del Sorbo, Hesham Abdelhady, Tina Romagnuolo

*University of Alberta:* Wendy Sligl, Nadia Baig, Oleksa Rewa, Sean Bagshaw;

*William Osler Health System:* Alexandra Binnie, Elizabeth Powell, Alexandra McMillan, Tracy Luk, Noah Aref;

***Croatia:***

*General Hospital Pozega*: Zdravko Andric, Sabina Cviljevic, Renata Đimoti, Marija Zapalac, Gordan *Mirković;*

*University Hospital of Infectious Diseases “Dr Fran Milhajevid”*: Vladimir Krajinovic,

Marko Kutleša, Viktor Kotarski;

*University Hospital of Zagreb*: Ana Vujaklija Brajković, Jakša Babel, Helena Sever, Lidija Dragija, Ira Kušan;

***Finland:***

*Helsinki University Hospital*: Suvi Vaara, Tero Varpula, Tuomas Oksanen, Leena Pettilä, Jonna Heinonen, Ville Pettilä;

*Tampere University Hospital:* Anne Kuitunen, Sari Karlsson, Annukka Vahtera, Heikki Kiiski, Sanna Ristimäki*;*

***France:***

*Ambroise Pare Hospital*: Amine Azaiz, Cyril Charron, Mathieu Godement, Guillaume Geri, Antoine Vieillard-Baron;

*Centre Hospitalier de Melun*: Franck Pourcine, Mehran Monchi;

*Centre Hospitalier Simone Veil, Beauvais*: David Luis, Romain Mercier, Anne Sagnier, Nathalie Verrier, Cecile Caplin, Jack Richecoeu, Daniele Combaux;

*Centre Hospitalier Sud Essonne*: Shidasp Siami, Christelle Aparicio, Sarah Vautier, Asma Jeblaoui, Delphine Lemaire-Brunel;

*Centre Hospitalier Tenon*: Muriel Fartoukh, Laura Courtin, Vincent Labbe, Guillaume Voiriot, Sara Nesrine Salhi;

*Centre Hospitalier Victor Dupouy*: Gaetan Plantefeve, Cécile Leparco, Damien Contou;

*CH de Mont de Marsan* : Arnaud Sement, Alexandre Gachet, Alexis Hanisch, Abdelmagid Haffiane, Anne-Hélène Boivin, Amelie Barreau, Elodie Guerineau, Séverine Poupblanc;

*CH des pays de Morlaix* : Pierre Yves Egreteau, Montaine Lefevre, Simon Bocher, Guillaume Le Loup, Lenaïg Le Guen, Vanessa Carn, Melanie Bertel;

CHR d'Orleans: Grégoire Muller, Mai-Anh Nay, Toufik Kamel, Dalila Benzekri, MD, Sophie Jacquier, Isabelle Runge, Armelle Mathonnet, François Barbier, MD, Anne Bretagnol;

*CHRU Tours Hopital Bretonneau*: Emmanuelle Mercier, Delphine Chartier, Charlotte Salmon, Pierre-François Dequin, Denis Garot;

*CHU Dupuytren, Limoges;*

*Hôpital Civil, Hôpitaux Universitaires de Strasbourg;*

*Hôpital de Hautepierre, Hôpitaux Universitaires de Strasbourg*: Francis Schneider, Vincent Castelain, Guillaume Morel, Sylvie L’Hotellier;

*Hôpital Simon Veil*, *Eaubonne*: Evelina Ochin, Christian Vanjak, Patrick Rouge, Lynda Bendjemar;

*Hospital Nord Franche-Comté*: Julio Badie, Fernando Daniel Berdaguer, Sylvain Malfroy, Chaouki Mezher, Charlotte Bourgoin, Guy Moneger, Elodie Bouvier;

*Lariboisière Hospital*: Bruno Megarbane, Sebastian Voicu, Nicolas Deye, Isabelle Malissin, Laetitia Sutterlin, Aymen Mrad, Adrien Pépin Lehalleur, Giulia Naim, Philippe Nguyen, Jean-Michel Ekhérian, Yvonnick Boué, Georgios Sidéris, Dominique Vodovar, Emmanuelle Guérin, Caroline Grant;

*Raymond Poincaré Hospital*: Djillali Annane, Pierre Moine, Nicholas Heming, Virginie Maxime, Isabelle Bossard, Tiphaine Barbarin Nicholier, Bernard Clair, David Orlikowski, Rania Bounab, Lilia Abdeladim;

*Vendee Hospital*: Gwenhael Colin, Vanessa Zinzoni, Natacha Maquigneau, Matthieu Henri-Lagarrigue, Caroline Pouplet;

***Germany:***

*Carl-Thiem-Klinikum Cottbus gGmbH*: Jens Soukup, Richard Wetzold, Madlen Löbel, Dr. Ing, Lisa Starke, Patrick Grimm;

*Charité - Universitätsmedizin Berlin*: André Finn, Gabriele Kreß, Uwe Hoff, Carl Friedrich Hinrichs, Jens Nee;

*Elisabeth Krankenhaus Essen*: Ingo Voigt, Robert Schueler, Elisabeth Blank, Vanessa Hüning, Melanie Steffen, Patricia Goralski;*Jena University Hospital*: Mathias W. Pletz, Stefan Hagel, Juliane Ankert, Steffi Kolanos, Frank Bloos;

*Klinikum Dortmund gGmbH*: Daniela Nickoleit-Bitzenberger, Bernhard Schaaf, Werner Meermeier, Katharina Prebeg, Harun Said Azzaui, Martin Hower, Klaus-Gerd Brieger, Corinna Elender, Timo Sabelhaus, Ansgar Riepe, Ceren Akamp, Julius Kremling, Daniela Klein, Elke Landsiedel-Mechenbier;

*University Hospital of Leipzig*: Sirak Petros, Kevin Kunz, Bianka Schütze;

*Universitätsklinikum Hamburg-Eppendorf*: Stefan Kluge, Axel Nierhaus, Dominik Jarczak, Kevin Roedl;

*Universitätsklinikum Köln*: Matthias Kochanek, Giusi Rueß-Paterno, Josette Mc-Kenzie, Dennis Eichenauer, Alexander Shimabukuro-Vornhagen;

*University Hospital of Frankfurt*: Gernot Gerhard Ulrich Rohde, Achim Grünewaldt, Jörg Bojunga;

*University Hospital of Würzburg*: Dirk Weismann, Anna Frey; Maria Drayss, M.E. Goebeler, Thomas Flor, Gertrud Fragner, Nadine Wahl, Juliane Totzke, Cyrus Sayehli;

*Vivantes Klinikum Neukölln*: Lorenz Reill, Michael Distler, Astrid Maselli;

***Hungary:***

*Almási Balogh Pál Hospital, Ózd*: János Bélteczki, István Magyar, Ágnes Fazekas, Sándor Kovács, Viktória Szőke;

*Jósa András County Hospital, Nyíregyháza*: Gábor Szigligeti, János Leszkoven;

***India:***

India Apollo Speciality Hospital - OMR, Chennai: Devachandran Jayakumar, Suresh Babu;

Apollo Main Hospital, Chennai: C Vignesh, Augustian James

Apollo Speciality Vanagaram, Vanagaram, Chennai: R Ebenezer, S Krishnamurthy, Lakshmi Ranganathan, Manisha;

***Ireland:***

*Beacon Hospital Dublin*: Daniel Collins, Kathy Brickell, Liadain Reid, Michelle Smyth, Patrick Breen, Sandra Spain;

*Beaumont Hospital*: Gerard Curley, Natalie McEvoy, Pierce Geoghegan, Jennifer Clarke;

*Galway University Hospitals*: John Laffey, Bairbre McNicholas, Michael Scully, Siobhan Casey, Maeve Kernan, Aoife Brennan, Ritika Rangan, Riona Tully, Sarah Corbett, Aine McCarthy, Oscar Duffy, David Burke;

*St Vincent’s University Hospital, Dublin*: Alistair Nichol, Kathy Brickell, Michelle Smyth, Leanne MC Hayes, Liadain Reid, Lorna Murphy, Andy Neill, Bryan Reidy, Michael O’Dwyer, Donal Ryan, Kate Ainscough;

*Cork University Hospital*: Prof Joe Eustace  PI, Dr Patrick Seigne , Ann-Marie O’Callaghan & Fionnuala O’Brien

*Waterford University Hospital*: Dr Sheeba Hakak,Dr Ray Kelly,Dr Craig Joyce,Dr Kirsten Joyce,Dr Wahid  Altaf, Leo Walsh,  Aisling Murphy

***Italy:***

*Humanitas Research Hospital:* Romina Aceto, Alessio Aghemo, Salvatore Badalamenti, Enrico Brunetta, Maurizio Cecconi, Michele Ciccarelli, Elena Constantini, Massimiliano Greco, Marco Folci, Carlo Selmi, Antonio Voza;

***Nepal:***

Nepal Grande International Hospital : Sushil Khanal, Sameena Amatya;

HAMS Hospital: Hem Raj Paneru, Sabin Koirala, Pratibha Paudel;

Nepal Mediciti Hospital: Diptesh Aryal, Kanchan Koirala, Namrata Rai, Subekshya Luitel;

Tribhuvan University Teaching Hospital : Hem Raj Paneru, Binita Bhattarai;

***Netherlands:***

*Canisius Wilhelmina Ziekenhuis*: Oscar Hoiting, Marco Peters, Els Rengers, Mirjam Evers, Anton Prinssen;

*Deventer Hospital*: Huub L.A. van den Oever, Arriette Kruisdijk-Gerritsen;

*Jeroen Bosch Ziekenhuis*: Koen Simons, Tamara van Zuylen, Angela Bouman;

*Meander Medisch Centrum*: Laura van Gulik;

*Radboud University Medical Center Nijmegen*: Mihai Netea, Jeroen Schouten, Peter Pickkers, Noortje Roovers, Margreet Klop-Riehl, Hetty van der Eng, Frank van de Veerdonk, Sonja Sloots-Cuppen, Lieke Preijers, Nienke van Oosten;

*UMC Leiden*: Evert de Jonge, Jeanette Wigbers, Michael del Prado;

*UMC Utrecht*: Marc Bonten, Olaf Cremer, Lennie Derde, Emma Rademaker, Jelle Haitsma Mulier, Birgit Romberg; Helen Leavis, Roger Schutgens, Marjolein van Opdorp, Henny Ophorst-den Besten, Karen Brakké

*Ziekenhuis Gelderse Vallei Ede*: Sjoerd van Bree, Marianne Bouw-Ruiter, Barbara Festen, Fiona van Gelder, Mark van Iperen, Margreet Osinga, Roel Schellaars, Dave Tjan, Ruben van der Wekken, Max Melchers, Arthur van Zanten;

*OLVG Amsterdam*: Nicole Juffermans, Matty Koopmans, Romein Dujardin, Bashar Alderink

*Hagaziekenhuis Den Haag*: Kees van Nieuwkoop, Thomas Ottens, Yorik Visser, Lettie van den Berg, Annemarie van der Kraan-Donker

*Bernhoven ziekenhuis Uden*: Kitty Slieker, Esther Ewalds, Arnate Sanders, Wendy Wittenberg, Heidi Peters Geurts;

***New Zealand*:**

*Auckland City Hospital, Cardiothoracic and Vascular ICU:* Shay McGuinness, Rachael Parke, Eileen Guilder, Magdalena Butler, Keri-Anne Cowdrey, Melissa Woollett;

*Auckland City Hospital, DCCM:* Colin McArthur, Thomas Hills, Lynette Newby, Yan Chen, Catherine Simmonds, Rachael McConnochie, Caroline O’Connor;

*Christchurch Hospital:* Jay Ritzema Carter, Seton Henderson, Kymbalee Van Der Heyden, Jan Mehrtens, Anna Morris, Stacey Morgan;

*Middlemore Hospital:* Tony Williams, Alex Kazemi, Susan Morpeth, Rima Song, Vivian Lai, Dinuraj Girijadevi;

*North Shore Hospital:* Robert Everitt, Robert Russell, Danielle Hacking;

*Rotorua Hospital:* Ulrike Buehner, Erin Williams;

*Tauranga Hospital:* Troy Browne, Kate Grimwade, Jennifer Goodson, Owen Keet, Owen Callender;

*Waikato Hospital:* Robert Martynoga, Kara Trask, Amelia Butler, PGCert,

*Wellington Hospital:* Paul Young, PhD, Chelsea Young, PGDip, Eden Lesona, Shaanti Olatunji, Leanlove Navarra, Raulle Sol Cruz

*Whangarei Hospital:* Katherine Perry, Ralph Fuchs, Bridget Lambert;

*Taranaki Base Hospital:* Jonathan Albrett, Carolyn Jackson, Simon Kirkham;

***Pakistan:***

Ziauddin Hospital Clifton Campus: Madiha Hashmi; Ashok Panjwani; Zulfiqar Ali Umrani, Shoaib Siddiq; Mohiuddin Shaikh; National Institute of Cardiovascular Diseases Pakistan: Nawal Salahuddin, Sobia Masood;

***Portugal*:**

*Hospital de Abrantes:* Nuno José Teodoro Amaro dos Santos Catorze, Tiago Nuno Alfaro Lima Pereira, Ricardo Manuel Castro Ferreira, Joana Margarida Pereira Sousa Bastos, Teresa Margarida Oliveira Batista;

***Romania*:**

*"Dr. Victor Babes" Clinical Hospital of Infectious and Tropical Diseases Bucharest:* Simin Aysel Florescu, Delia Stanciu, Mihaela Florentina Zaharia, Alma Gabriela Kosa, Daniel Codreanu;

***Saudi Arabia*:**

*King Abdulaziz Medical City- Riyadh:* Yaseen M Arabi,  Eman Al Qasim, Lolowa Alswaidan, Mohamed M Hegazy, Hatim Arishi, Ali Al Amri, Samah Y AlQahtani, Brintha Naidu, Haytham Tlayjeh, Sajid Hussain, Farhan [Al Enezi](https://cams.ksau-hs.edu.sa/index.php/en/faculty-staff/directory/dean-s-office/academic-and-student-affairs-male/274-dr-farhan-al-enezi), Sheryl Ann Abdukahil;

***Spain*:**

*Hospital del Mar:* Rosana Muñoz-Bermúdez, Judith Marin-Corral, Anna Salazar Degracia, Francisco Parrilla Gómez, Maria Isabel Mateo López;

*Reina Sofia University Hospital:* Rafael León López, Jorge Rodríguez-Gómez, Sheila Cárcel, Rosario Carmona, Carmen de la Fuente, Marina Rodriguez;

***United Kingdom*:**

*Aberdeen Royal Infirmary:* Callum Kaye, Amanda Coutts, Lynn MacKay;

*Addenbrooke's Hospital:* Charlotte Summers, Petra Polgarova, Neda Farahi, Eleonore Fox;

*Alder Hey Children’s NHS Foundation Trust:* Stephen J McWilliam, Daniel B Hawcutt, Laura Rad, Laura

O’Malley, Jennifer Whitbread, Dawn Jones, Rachael Dore, Paula Saunderson;

*Alexandra Hospital Redditch:* Olivia Kelsall, Nicholas Cowley, Laura Wild, Jessica Thrush, Hannah

Wood, Karen Austin;

*Altnagelvin Hospital:* Adrian Donnelly, Martin Kelly, Naoise Smyth, Sinéad O’Kane, Declan McClintock,

Majella Warnock, Ryan Campbell, Edmund McCallion;

*Antrim Area Hospital:* Paul Johnson, Shirley McKenna, Joanne Hanley, Andrew Currie, Barbara Allen,

Clare McGoldrick, Moyra McMaster;

*Barnet Hospital:* Rajeev Jha, Michael Kalogirou, Christine Ellis, Vinodh Krishnamurthy, Aibhilin

O’Connor, Saranya Thurairatnam;

*Basildon Universty Hospital:* Dipak Mukherjee, Agilan Kaliappan, Mark Vertue, Anne Nicholson, Joanne

Riches, Gracie Maloney, Lauren Kittridge, Amanda Solesbury, Angelo Ramos;

*Belfast Health and Social Care Trust (Belfast City Hospital, Mater Infirmorium, Royal Victoria Hospital):*

Jon Silversides, Peter McGuigan, Kathryn Ward, Aisling O’Neill, Stephanie Finn, Chris Wright, Jackie

Green, Érin Collins;

*Brighton and Sussex University Hospitals Trust:* Barbara Phillips, Laura Oritz-Ruiz de Gordoa;

*Calderdale and Huddersfield Foundation Trust:* Jez Pinnell, Matt Robinson, Lisa Gledhill, Tracy Wood;

*Cardiff and Vale University Health Board:* Matt Morgan, Jade Cole, Helen Hill, Michelle Davies,

Angharad Williams, Emma Thomas, Rhys Davies, Matt Wise;

*Charing Cross Hospital:* David Antcliffe, Maie Templeton, Roceld Rojo, Phoebe Coghlan, Joanna Smee,

Gareth Barker;

*Chesterfield Royal Hospital:* Euan Mackay, Jon Cort, Amanda Whileman, Thomas Spencer, Nick Spittle,

Sarah Beavis, Anand Padmakumar, Katie Dale, Joanne Hawes, Emma Moakes, Rachel Gascoyne, Kelly

Pritchard, Lesley Stevenson, Justin Cooke, Karolina Nemeth-Roszpopa;

*The Christie NHS Foundation Trust:* Vidya Kasipandian, Amit Patel, Suzanne Allibone, Roman Mary

Genetu;

*Colchester Hospital:* Mohamed Ramali, Ooi HC, Alison Ghosh, Rawlings Osagie, Malka Jayasinghe

Arachchige, Melissa Hartley;

*Countess of Chester Hospital:* Peter Bamford, Andrew Reid, Kathryn Cawley, Maria Faulkner, Charlotte

Pickering;

*Croydon University Hospital:* Ashok Sundar Raj, Georgios Tsinaslanidis, Reena Nair Khade, Gloria

Nwajei Agha, Rose Nalumansi Sekiwala;

*Cumberland Infirmary:* Tim Smith, Chris Brewer, Jane Gregory;

*Darlington Memorial Hospital:* James Limb, Amanda Cowton, Julie O’Brien, Kelly Postlethwaite;

*Derriford Hospital:* Nikitas Nikitas, Colin Wells, Liana Lankester, Helen McMillan;

*Dorset County Hospital:* Mark Pulletz, Patricia Williams, Jenny Birch, Sophie Wiseman, Sarah Horton;

*East Kent Hospitals (Queen Elizabeth the Queen Mother Hospital):* Ana Alegria, Salah Turki, Tarek

Elsefi, Nikki Crisp, Louise Allen;

*East Lancashire Hospitals NHS Trust (Royal Blackburn Hospital):* Nicholas Truman, Matthew Smith, Sri

Chukkambotla, Wendy Goddard, Stephen Duberley, Meherunnisa Khan, Aayesha Kazi;

*Freeman Hospital and Royal Victoria Infirmary, Newcastle upon Tyne*: Iain J McCullagh, Tom Cairns,

Helen Hanson, Bijal Patel, Ian Clement;

*Frimley Health NHS Foundation Trust:* Omar Touma, Susan Holland, Christopher Hodge, Holly Taylor,

Meera Alderman, Nicky Barnes, Joana Da Rocha, Catherine Smith, Nicole Brooks, Thanuja

Weerasinghe, Julie-Ann Sinclair, Yousuf Abusamra, Ronan Doherty, Joanna Cudlipp, Rajeev Singh, Haili

Yu, Admad Daebis, Christopher NG, Sara Kendrick, Anita Saran, Ahmed Makky, Danni Greener, Louise

Rowe-Leete, Alexandra Edwards, Yvonne Bland, Rozzie Dolman, Tracy Foster;

*Gateshead Health NHS Trust:* Vanessa Linnett, Amanda Sanderson, Jenny Ritzema, Helen Wild,

Rachael Lucas, Yvonne Marriott;

*George Eliot Hospital:* Divya Khare, Meredith Pinder, Amitha Gopinath, Thogulava Kannan, Steven

Dean, Piyush Vanmali;

*Glan Clwyd Hospital:* Richard Pugh, Richard Lean, Xinyi Qiu, Jeremy Scanlan, Andrew Evans, Gwyneth

Davies, Joanne Lewis;

*Glangwili General Hospital:* Yvonna Plesnikova, Ahmed Ben Khoud, Samantha Coetzee;

*Glasgow Royal Infirmary:* Kathryn Puxty, Susanne Cathcart, Dominic Rimmer, Catherine Bagot,

Kathryn Scott, Laila Martin;

*Glenfield Hospital Leicester:* Hakeem Yusuff, Graziella Isgro, Chris Brightling, Michelle Bourne, Michelle

Craner, Rebecca Boyles;

*Grange University Hospital:* Tamas Szakmany, Shiney Cherian, Gemma Williams, Christie James, Abby

Waters;

*Great Western Hospitals NHS Foundation Trust:* Rachel Prout, Roger Stedman, Louisa Davies,

Suzannah Pegler, Lynsey Kyeremeh, Louise Moorhouse;

*Guy’s & St Thomas’ NHS Foundation Trust:* Manu Shankar-Hari, Gill Arbane, Marina Marotti, Aneta

Bociek, Sara Campos;

*Hammersmith Hospital:* Stephen Brett, Sonia Sousa Arias, Rebecca Elin Hall;

*Homerton University Hospital NHS Foundation Trust:* Susan Jain, Abhinav Gupta, Catherine Holbrook,

Pierre Antoine;

*James Cook University Hospital:* Jeremy Henning, Stephen Bonner, Keith Hugill, Emanuel Cirstea, Dean

Wilkinson, Jessica Jones, Mohammed Nagy Tawfik Altomy;

*James Paget University Hospitals:* Michal Karlikowski, Helen Sutherland, Elva Wilhelmsen, Jane

Woods, Julie North;

*Kettering General Hospital:* Dhinesh Sundaran, Laszlo Hollos, Anna Williams, Margaret Turns, Joanne

Walsh;

*King’s College Hospital (Denmark Hill site):* Phil Hopkins, John Smith, Harriet Noble, Kevin O’Reilly,

Reena Mehta, Onyee Wong, Esther Makanju, Deepak Rao, Nyma Sikondari, Sian Saha, Ele Corcoran,

Evita Pappa, Maeve Cockrell, Clare Donegan, Morteza Balaie;

*Lancashire Teaching Hospitals NHS Foundation Trust:* Shondipon Laha, Mark Verlander, Alexandra

Williams, Avinash Kumar Jha;

*Leeds Teaching Hospitals Trust:* Elankumaran Paramasivam, Elizabeth Wilby, Bethan Ogg, Clare

Howcroft, Angelique Aspinwall, Sam Charlton, Richard Gould, Deena Mistry, Sidra Awan, Caroline

Bedford, Joanne Carr-Wilkinson;

*Leicester General Hospital:* Andrew Hall, Jill Cooke, Caroline Gardiner-Hill, Carolyn Maloney, Nigel

Brunskill, Olivia Watchorn, Chloe Hardy;

*Leicester Royal Infirmary:* Hafiz R Qureshi, Neil Flint, Sarah Nicholson, Sara Southin, Andrew Nicholson,

Amardeep Ghattaoraya;

*Lewisham and Greenwich NHS Trust:* Dr Daniel Harding, Sinead O’Halloran, Amy Collins, Emma Smith,

Estefania Trues;

*Liverpool Foundation Trust Aintree:* Barbara Borgatta, Ian Turner-Bone, Amie Reddy, Laura Wilding;

*Liverpool Heart and Chest Hospital:* Craig Wilson, Zuhra Surti;

*Luton and Dunstable University Hospital:* Loku Chamara Warnapura, Ronan Agno, Prasannakumari

Sathianathan, Deborah Shaw, Nazia Ijaz, Adam Spong, Suganya Sabaretnam, Dean Burns, Eva Lang,

Margaret Louise Tate;

*Maidstone and Tunbridge Wells NHS Trust:* David Golden, Miriam Davey, Rebecca Seaman, Alexander

Osborne;

*Manchester Royal Infirmary:* Jonathan Bannard-Smith, Richard Clark, Kathrine Birchall, Joanne Henry,

Fiona Pomeroy, Rachael Quayle, Katharine Wylie, Anila Sukuraman, Maya John, Sindhu Sibin;

*Medway Maritime Hospital:* Arystarch Makowski, Beata Misztal, Syeda Haider, Angela Liao, Rebecca

Squires;

*Milton Keynes University Hospital:* Richard Stewart, Esther Mwaura, Louise Mew, Lynn Wren, Felicity

Willams, Sara-Beth Sutherland, Rashmi Rebello;

*Mid & South Essex NHS Foundation Trust:* Aneta Oborska, Abdul Kayani, Selver Kalchko-Veyssal,

Rajalakshmi Orath Prabakaran, Bernard Hadebe, Selver KalchkoVeyssal;

*Musgrove Park Hospital:* Richard Innes, Patricia Doble, Libby Graham, Charmaine Shovelton, Tessa

Dean;

*Nevill Hall Hospital:* Vincent Hamlyn, Nancy Hawkins, Anna Roynon-Reed, Sean Cutler, Sarah Lewis;

*Newham University Hospital:* Juan Martin Lazaro, Tabitha Newman;

*Ninewells Hospital:* Pauline Austin, Susan Chapman, Louise Cabrelli;

*Norfolk and Norwich University Hospital:* Simon Fletcher, Jurgens Nortje, Deirdre Fottrell-Gould,

Georgina Randell, Katie Stammers, Gail Healey, Marta Goncalves Pinto;

*Northampton General Hospital:* Mohsin Zaman, Einas Elmahi, Andrea Jones, Kathryn Hall;

*Northern General Hospital, Sheffield:* Gary H Mills, Ajay Raithatha, Kris Bauchmuller, Kim Ryalls, Kate

Harrington, Helen Bowler, Jas Sall, Richard Bourne;

*Northwick Park Hospital NHS Trust:* Jamie Gross, Natalie Massey, Olumide Adebambo, Matilda Long,

Kiran Tony;

*North Manchester General Hospital:* Zoe Borrill, Tracy Duncan, Andrew Ustianowski, Alison Uriel, Ayaa

Eltayeb, Jordan Alfonso, Samuel Hey, Joanne Shaw, Claire Fox, Gabriella Lindergard, Bethan Charles,

Bethany Blackledge, Karen Connolly, Jade Harris;

*North Middlesex University Hospital:* Jeronimo Moreno Cuesta, Kugan Xavier, Dharam Purohit, Munzir

Elhassan, Anne Haldeos, Rachel Vincent, Marwa Abdelrazik, Samuel Jenkins, Arunkumar Ganesan,

Rohit Kumar, David Carter, Dhanalakshmi Bakthavatsalam, Alasdair Frater, Malik Tahir Saleem;

*Oxford University Hospitals:* Matthew Rowland, Paula Hutton, Archana Bashyal, Neil Davidson, Clare

Hird;

*Pilgrim Hospital Boston:* Manish Chhablani, Gunjan Phalod, Amy Kirkby, Simon Archer, Kimberley

Netherton;

*Princess Royal Hospital:* Denise Skinner, Jane Gaylard, Julie Newman;

*Princess of Wales Hospital:* Sonia Sathe, Lisa Roche, Ellie Davies;

*Poole Hospital:* Henrik Reschreiter, Julie Camsooksai, Sarah Patch, Sarah Jenkins, Charlotte Humphrey;

*Queen Alexandra Hospital Portsmouth:* David Pogson, Steve Rose, Zoe Daly, Lutece Brimfield, Angie

Nown;

*Queen Elizabeth Hospital, Birmingham:* Dhruv Parekh, Colin Bergin, Michelle Bates, Christopher

McGhee, Daniella Lynch, Khushpreet Bhandal, Kyriaki Tsakiridou, Amy Bamford, Lauren Cooper, Tony

Whitehouse, Tonny Veenith, Elliot Forster, Martin O'Connell;

*Queen Elizabeth University Hospital, Glasgow:* Malcolm A.B. Sim, Sophie Kennedy Hay, Steven

Henderson, Maria Nygren, Eliza Valentine;

*Queen’s Hospital, Burton:* Amro Katary, Gillian Bell, Louise Wilcox, Michail Mataliotakis, Paul Smith,

Murtaza Asif Ali, Agah Isguzar;

*Queen’s Hospital, Romford:* Mandeep-Kaur Phull, Abbas Zaidi, Tatiana Pogreban, Lace Paulyn

Rosaroso;

*Queens Medical Centre and Nottingham City Hospital:* Daniel Harvey, Benjamin Lowe, Megan

Meredith, Lucy Ryan, DREEAM Research Team;

*The Rotherham NHS Foundation Trust:* Anil Hormis, Rachel Walker, Dawn Collier, Sarah Kimpton,

Susan Oakley;

*Royal Alexandra Hospital:* Kevin Rooney, Natalie Rodden, Nicola Thomson, Deborah McGlynn, Lynn

Abel, Lisa Gemmell, Radha Sundaram, James Hornsby;

*Royal Berkshire Hospital:* Andrew Walden, Liza Keating, Matthew Frise, Sabi Gurung Rai, Shauna

Bartley;

*Royal Bournemouth and Christchurch Hospitals:* Martin Schuster-Bruce, Sally Pitts, Rebecca Miln,

Laura Purandare, Luke Vamplew;

*Royal Brompton Hospital:* Brijesh Patel, Debra Dempster, Mahitha Gummadi, Natalie Dormand, Shu

Fang Wang;

*Royal Cornwall NHS Trust:* Michael Spivey, Sarah Bean, Karen Burt, Lorraine Moore, Fiona Hammonds,

Carol Richards;

*Royal Devon and Exeter NHS Foundation Trust:* Christopher Day, Letizia Zitter, Sarah Benyon;

*Royal Glamorgan Hospital:* Jayaprakash Singh, Ceri Lynch, Lisa Roche, Justyna Mikusek, Bethan

Deacon, Keri Turner;

*Royal Gwent Hospital*: Evelyn Baker, John Hickey, Shreekant Champanerkar, Lindianne Aitken,

Lorraine LewisProsser, Christie James;

*Royal Hallamshire Hospital, Sheffield:* Gary H Mills, Norfaizan Ahmad, Matt Wiles, Jayne Willson;

*Royal Hampshire Hospitals:* Irina Grecu, Jane Martin, Caroline Wrey Brown, Ana-Marie Arias, Emily

Bevan, Samantha Westlake;

*Royal Infirmary of Edinburgh:* Thomas H Craven, David Hope, Jo Singleton, Sarah Clark, Corrienne

McCulloch, Simon Biddie;

*Royal Liverpool University Hospital:* Ingeborg D Welters, David Oliver Hamilton, Karen Williams,

Victoria Waugh, David Shaw, Suleman Mulla, Alicia Waite, Jaime Fernandez Roman, Maria Lopez

Martinez, Brian Johnston;

*Royal London Hospital:* Zudin Puthucheary, Timothy Martin, Filipa Santos, Ruzena Uddin, Maria

Fernandez, Fatima Seidu, Alastair Somerville, Mari-Liis Pakats, Salma Begum, Tasnin Shahid;

*The Royal Free Hospital:* Sanjay Bhagani, Mark De Neef, Sara Mingo Garcia, Amitaa Maharajh, Aarti

Nandani, Jade Dobson, Gloria Fernando, Christine Eastgate, Keith Gomez, Zakee Abdi;

*The Royal Marsden NHS Foundation Trust:* Kate Colette Tatham, Shaman Jhanji, Ethel Black, Arnold

Dela Rosa, Ryan Howle, Ravishankar Rao Baikady;

*The Royal Oldham Hospital:* Redmond P Tully, Andrew Drummond, Joy Dearden, Jennifer E Philbin,

Sheila Munt;

*The Royal Wolverhampton NHS Trust:* Shameer Gopal, Jagtar- Singh Pooni, Saibal Ganguly, Andrew

Smallwood, Stella Metherell;

*Royal Papworth Hospital:* Alain Vuylsteke, Charles Chan, Saji Victor, COVID Research Team, Papworth

Hospital;

*Royal Stoke Hospital:* Ramprasad Matsa, Minerva Gellamucho, Michelle Davies;

*Royal Surrey County Hospital:* Ben Creagh-Brown, Cheryl Marriot, Armorel Salberg, Louisa Zouita,

Sarah Stone, Natalia Michalak, Sinead Donlon, Shelia Mtuwa, Irving Mayangao, Jerik Verula, Dorota

Burda, Celia Harris, Emily Jones, Paul Bradley, Esther Tarr, Lesley Harden, Charlie Piercy;

*Royal United Hospital Bath:* Jerry Nolan, Ian Kerslake, Tim Cook, Tom Simpson, James Dalton, Carrie

Demetriou, Sarah Mitchard, Lidia Ramos, Katie White, Toby Johnson, William Headdon, Stephen

Spencer, Alison White, Lucy Howie;

*Russells Hall Hospital:* Michael Reay, Steve Jenkins, Angela Watts, Eleanor Traverse, Stacey Jennings,

Vikram Anumakonda, Caroline Tuckwell, Karen Pearson, Kath Harrow, Julie Matthews, Karen

McGarry, Vanessa Moore, Lucie Smith, Anna Summerfield;

*Salisbury NHS Foundation Trust:* Phil Donnison, Ruth Casey, Ben Irving, Wadzanai Matimba-Mupaya,

Catherine Reed, Alpha Anthony, Fiona Trim, Lenka Cambalova, Debra Robertson, Anna Wilson;

*Salford Royal NHS Foundation Trust:* Paul Dark, Alice Harvey, Reece Doonan, Liam McMorrow, Karen

Knowles, Jessica Pendlebury, Stephanie Lee, Jane Perez, Bethan Charles, Tracy Marsden, Melanie Taylor, Angiy Michael, Matthew Collis, Andrew Claxton, Wadih Habeichi, Dan Horner, Melanie Slaughter, Vicky Thomas, Nicola Proudfoot, Claire Keatley;

*Sandwell and West Birmingham NHS Trust:* Jonathan Hulme, Santhana Kannan, Fiona Kinney, Ho Jan

Senya, Anne Hayes;

*Sherwood Forest Hospitals NHS Foundation Trust:* Valli Ratnam, Mandy Gill, Jill Kirk, Sarah Shelton;

*South Tyneside District Hospital:* Christian Frey, Riccardo Scano, Madeleine McKee, Peter Murphy;

*Southmead Hospital:* Matt Thomas, Ruth Worner, Beverley Faulkner, Emma Gendall, Kati Hayes,

Hayley Blakemore, Borislava Borislavova;

*St. Bartholomew’s Hospital:* Colin Hamilton-Davies, Carmen Chan, Celina Mfuko, Hakam Abbass,

Vineela Mandadapu;

*St. George’s Hospital:* Susannah Leaver, Kamal Patel, Sarah Farnell-Ward, Romina Pepermans Saluzzio,

Sam Rawlins, Christine Sicat;

*St. Mary’s Hospital:* Anthony Gordon; Dorota Banach, Ziortza Fernández de Pinedo Artaraz, Leilani

Cabreros, Victoria Latham;

*St. Peter’s Hospital, Chertsey:* Ian White, Maria Croft, Nicky Holland, Rita Pereira;

*Stepping Hill Hospital, Stockport:* Ahmed Zaki, David Johnson, Hywel Garrard, Vera Juhaz, Louise

Brown, Abigail Pemberton;

*Sunderland Royal Hospital:* Alistair Roy, Anthony Rostron, Lindsey Woods, Sarah Cornell;

*Swansea Bay University Health Board:* Suresh Pillai, Rachel Harford, Helen Ivatt, Debra Evans, Suzanne

Richards, Eilir Roberts, James Bowen, James Ainsworth;

*Torbay and South Devon NHS Foundation Trust:* Angela Foulds, Adam Revill;

*United Lincolnshire NHS Trust:* Russell Barber, Anette Hilldrith, Gunjan Phalod;

*University Hospitals Bristol & Weston NHS Foundation Trust:* Jeremy Bewley, Katie Sweet, Lisa

Grimmer, Rebekah Johnson, Rachel Wyatt, Karen Morgan, Siby Varghese, Charlotte Bradbury, Joanna

Willis, Emma Stratton, Laura Kyle, Daniel Putensen, Kay Drury, Agnieszka Skorko;

*University Hospitals Coventry & Warwickshire NHS Trust:* Pamela Bremmer, Geraldine Ward,

Christopher Bassford;

*University Hospital of North Tees:* Farooq Brohi, Vijay Jagannathan, Michele Clark, Sarah Purvis, Bill

Wetherill;

*University Hospital Southampton NHS Foundation Trust:* Ahilanandan Dushianthan, Rebecca Cusack,

Kim de Courcy-Golder, Karen Salmon, Rachel Burnish, Simon Smith, Susan Jackson, Winningtom Ruiz,

Zoe Duke, Magaret Johns, Michelle Male, Kirsty Gladas, Satwinder Virdee, Jacqueline Swabe, Helen

Tomlinson;

*Warwick Hospital:* Ben Attwood, Penny Parsons, Bridget Campbell, Alex Smith;

*Watford General Hospital:* Valerie J Page, Xiao Bei Zhao, Deepali Oza, Gail Abrahamson, Ben Sheath,

Chiara Ellis;

*Western General Hospital, Edinburgh:* Jonathan Rhodes, Thomas Anderson, Sheila Morris;

*Whipps Cross Hospital:* Charlotte Xia Le Tai, Amy Thomas, Alexandra Keen, Carey Tierney, Nimca

Omer, Gina Bacon;

*Whiston Hospital*: Ascanio Tridente, Karen Shuker, Jeanette Anders, Sandra Greer, Paula Scott, Amy

Millington, Philip Buchanan, Jodie Kirk

*Wirral University Teaching Hospital NHSFT:* Craig Denmade, Girendra Sadera, Reni Jacob, Cathy Jones,

Debbie Hughes;

*Worcester Royal Hospital:* Stephen Digby, Nicholas Cowley, Laura Wild, Jessica Thrush, Hannah Wood,

Karen Austin;

*Wrexham Maelor Betsi Cadwaladr University Hospital*: David Southern, Harsha Reddy, Sarah Hulse,

Andrew Campbell, Mark Garton, Claire Watkins, Sara Smuts;

*Wrightington, Wigan and Leigh Teaching Hospitals NHS Foundation Trust:* Alison Quinn, Benjamin

Simpson, Catherine McMillan, Cheryl Finch, Claire Hill, Josh Cooper;

*Wye Valley NHS Trust:* Joanna Budd, Charlotte Small, Ryan O’Leary, Janine Birch, Emma Collins,

Andrew Holland;

*Wythenshawe Hospital*: Peter D G Alexander, Tim Felton, Susan Ferguson, Katharine Sellers, Luke

Ward;

*York Teaching Hospital*: David Yates, Isobel Birkinshaw, Kay Kell, Zoe Scott, Harriet Pearson;

***United States of America:***

*University of Pittsburgh Research Staff:* CRISMA Center—Kelsey Linstrum, Stephanie Montgomery, Kim Basile, Dara Stavor, Dylan Burbee, Amanda McNamara, Renee Wunderley, Nicole Bensen, Aaron Richardson; MACRO Center—Peter Adams, Tina Vita, Megan Buhay, Denise Scholl, Matthew Gilliam, James Winters, Kaleigh Doherty, Emily Berryman

*UPMC Hospital Champions:* UPMC Altoona—Mehrdad Ghaffari, UPMC East—Meghan Fitzpatrick; UPMC Jameson and Horizon —Kavitha Bagavathy; UPMC Mercy—Mahwish Hussain, Chenell Donadee; UPMC Williamsport—Emily Brant; UPMC McKeesport—Kayla Bryan-Morris, John Arnold and Bob Reynolds; UPMC Hamot—Gregory Beard; UPMC Presbyterian—Bryan McVerry, David Huang, Ghady Haidar, Alexandra Weissman, Florian Mayr; UPMC Passavant – Matthew Gingo;

*UPMC COVID Therapeutics Committee:* Erin McCreary, Elise Martin, Ryan Bariola, Alex Viehman, Jessica Daley, Alyssa Lopus, Mark Schmidhofer, UPMC Directors of Pharmacy

*UPMC ICU Service Center:* Rachel Sackrowitz, Chenell Donadee, Aimee Skrtich

*UPMC Wolff Center:* Tami Minnier, Mary Kay Wisniewski, Katelyn Mayak

*UPMC eRecord Team:* Richard Ambrosino, Sherbrina Keen, Sue Della Toffalo, Martha Stambaugh, Ken Trimmer, Reno Perri, Sherry Casali, Rebecca Medva, Brent Massar, Ashley Beyerl, Jason Burkey, Sheryl Keeler, Maryalyce Lowery, Lynne Oncea, Jason Daugherty, Chanthou Sevilla, Amy Woelke, Julie Dice, Lisa Weber, Jason Roth, Cindy Ferringer, Deborah Beer, Jessica Fesz, Lillian Carpio

*Data Collection/Curation Team:* Salim Malakouti (Computer Science, University of Pittsburgh), Edvin Music and Dan Ricketts (CRISMA Center), Andrew King (Biomedical Informatics, University of Pittsburgh), Gilles Clermont (Critical Care Medicine), Robert Bart (UPMC Health Services Division*)*

*UPMC Clinical Analytics:* Oscar Marroquin, Kevin Quinn, William Garrard, Kyle Kalchthaler

*UPMC Office of Healthcare Innovation:* Derek Angus

*Department of Emergency Medicine:* Alexandra Weissman, Donald Yealy, David Barton, Nadine Talia

*Department of Critical Care Medicine:* David Huang, Florian Mayr, Andrew Schoenling, Mark Andreae, Varun Shetty, Emily Brant, Brian Malley, Chenell Donadee, Derek Angus, Christopher Horvat, Christopher Seymour, Timothy Girard, Gilles Clermont, Rachel Sackrowitz, Robert Bart

*Division of Infectious Diseases:* Ghady Haidar

*Division of Pulmonary, Allergy, and Critical Care Medicine:* William Bain, Ian Barbash, Mark Brown, Antu Das, Meghan Fitzpatrick, Christopher Franz, Stefanie Hannan, Georgios Kitsios, Ritchie Koshy, Sophia Lieber, Emily Lyons, John McDyer, Bryan McVerry, Kaveh Moghbeli, Iulia Popescu, Brian Rosborough, Faraaz Shah, Tomeka Suber

*Global Coalition for Adaptive Research (GCAR):* Meredith Buxton, Brian Alexander, Tracey Roberts

*UPMC Laboratory Services*: Alan H. Wells

*Berry Consultants:* Roger Lewis, Michelle Detry, Anna McGlothlin, Christina Saunders, Mark Fitzgerald, Ashish Sanil, Scott Berry

**Clinical Trials Groups**

The REMAP-CAP platform is supported by the Australian and New Zealand Intensive Care Society Clinical Trials Group, the Canadian Critical Care Trials Group, the Irish Critical Care Clinical Trials Network, the UK Critical Care Research Group, and the International Forum of Acute Care Trialists.

REMAP-CAP was supported in the UK by the NIHR Clinical Research Network and we acknowledge the contribution of Kate Gilmour, Karen Pearson, Chris Siewerski, Sally-Anne Hurford, Emma Marsh, Debbie Campbell, Penny Williams, Kim Shirley, Meg Logan, Jane Hanson, Anne Oliver, Mihaela Sutu, Sheenagh Murphy, Latha Aravindan, Joanne Collins, Holly Monaghan, Adam Unsworth, Seonaid Beddows, Laura Ann Dawson, Sarah Dyas, Adeeba Asghar, Kate Donaldson, Tabitha Skinner, Nhlanhla Mguni, Natasha Muzengi, Ji Luo, Joanna O’Reilly, Chris Levett, Alison Potter, David Porter, Teresa Lockett, Jazz Bartholomew, Clare Rook, Hannah Williams, Alistair S Hall, Hilary Campbell, Holly Speight, Sandra Halden, Susan Harrison, Mobeena Naz, Charles Rounds, Kaatje Lomme, Johnathan Sheffield, William Van’t Hoff, James D Williamson, Catherine Birch, Morwenna Brend, Emma Chambers, , Sarah Crawshaw, Chelsea Drake, Heather Harper, Stephen Lock, Eleanor OKell, Amber Hayes, Susan Walker, Jayne Goodwin, Helen Hodgson, Yvette Ellis, Dawn Williamson, Madeleine Bayne, Shane Jackson, Rahim Byrne, Sonia McKenna, Alison Clinton, NIHR Urgent Public Health Group: <https://www.nihr.ac.uk/documents/urgent-public-health-group-members/24638#Members>

REMAP-CAP was supported in Ireland by the Irish Critical Care Clinical Trials Network and we acknowledge the contribution of Kate Ainscough, Kathy Brickell and Peter Doran.

REMAP-CAP was supported in France by the CRICS-TRIGGERSEP network.

REMAP-CAP was supported in the Netherlands by the Research Collaboration Critical Care the Netherlands (RCC-Net).

REMAP-CAP was supported in the United States by the Translational Breast Cancer Research Consortium, the UPMC Learning While Doing Program, and the Global Coalition for Adaptive Research

2. Additional methods
   2.1. REMAP-CAP eligibility criteria for suspected or proven COVID-19

###### Platform inclusion criteria:

During a pandemic, the platform inclusion criteria as listed in the Core Protocol (www.remapcap.org) are modified as described in the Pandemic Appendix to the Core Protocol ([www.remapcap.org](http://www.remapcap.org)). As such, the COVID-19 platform level inclusion criteria are:

In order to be eligible to participate in the pandemic aspects of REMAP-CAP, a patient must meet the following criteria:

- Adult patient admitted to hospital with acute illness due to suspected or proven pandemic infection
- Either Severe State or Moderate State patient. Patients were categorized into one of two states, which were mutually exclusive:
  - Severe State, defined by receiving respiratory or cardiovascular organ failure support in an intensive care unit (ICU).
    - Respiratory organ support is defined as invasive or non-invasive mechanical ventilation including via high flow nasal cannula if flow rate >30 L/min and FiO_2_ >0.4. If non-invasive ventilation would normally be provided but is being withheld, due to infection control concerns associated with aerosol generating procedures, then the patient still meets the severe disease state criteria.
    - Cardiovascular organ support was defined as the intravenous infusion of any vasopressor or inotrope.
    - Pandemic surge capacity means that provision of advanced organ support may need to occur in locations that do not usually provide ICU-level care. Therefore, an ICU is defined as an area within the hospital that is repurposed so as to be able to deliver one or more of the qualifying organ failure supports (non-invasive ventilation, invasive ventilation, and vasopressor therapy)
  - Moderate State, defined by not being admitted to an ICU, or admitted to an ICU but not receiving organ support (organ support defined as for the Severe State)

Patients were assigned to either Moderate State or Severe State at the time of assessment of edibility. Further details are provided in the Pandemic Appendix to the Core Protocol ([www.remapcap.org](http://www.remapcap.org)).

###### Platform exclusion criteria:

- Death is deemed to be imminent and inevitable during the next 24 hours AND one or more of the participant, substitute decision maker or attending physician are not committed to full active treatment
- Patient is expected to be discharged from hospital today or tomorrow
- More than 14 days have elapsed while admitted to hospital with symptoms of an acute illness due to suspected or proven pandemic infection
- Previous participation in this REMAP within the last 90 days

###### Immune Modulation Therapy domain specific inclusion criteria:

- Adult patient admitted to hospital with acute illness due to suspected or proven pandemic (COVID-19) infection
- Microbiological testing for SARS-CoV-2 of upper or lower respiratory tract secretions or both has occurred or is intended to occur

###### Immune Modulation Therapy domain specific exclusion criteria:

- More than 24 hours has elapsed since ICU admission
- Patient has already received any dose of one or more of any form of interferon, anakinra, tocilizumab, or sarilumab during this hospitalization or is on long-term therapy with any of these agents prior to this hospital admission
- Known condition or treatment resulting in ongoing immune suppression including neutropenia prior to this hospitalization
- Patient has been randomized in a trial evaluating an immune modulation agent for proven or suspected COVID-19 infection, where the protocol of that trial requires ongoing administration of study drug
- The treating clinician believes that participation in the domain would not be in the best interests of the patient

###### Intervention specific exclusion criteria:

- Known hypersensitivity to an agent specified as an intervention in this domain will exclude a patient from receiving that agent
- Intention to prescribe systemic corticosteroids for any reason, other than participation in the Corticosteroid domain of this platform, will result in exclusion from receiving IFN-β1a
- Known hypersensitivity to proteins produced by E. coli will result in exclusion from receiving anakinra
- Known or suspected pregnancy will result in exclusion from the anakinra, IFN-β1a, tocilizumab, and sarilumab interventions. It is normal clinical practice that women admitted who are in an age group in which pregnancy is possible will have a pregnancy test conducted. The results of such tests will be used to determine interpretation of this exclusion criteria.
- A baseline alanine aminotransferase or an aspartate aminotransferase that is more than five times the upper limit of normal will result in exclusion from receiving tocilizumab or sarilumab
- A baseline platelet count < 50 x 10^9^ / L will result in exclusion from receiving tocilizumab or sarilumab
2.2. Additional statistical methods

###### Secondary outcomes:

- Hospital mortality (dichotomous)
- 90-day survival (time to event)
- Respiratory support-free days
- Cardiovascular support-free days
- Time to ICU discharge (censored at day 90)
- Time to hospital discharge (censored at day 90)
- WHO ordinal scale (range: 0-8, where 0 = no illness, 1-7 = increasing level of care, and 8 = death) assessed at day 14.^1^ In this analysis categories 0,1,2 have been condensed into one category for all patients discharge from hospital.
- Progression to invasive mechanical ventilation, extracorporeal membrane oxygenation (ECMO), or death among those not ventilated at baseline

###### Exploratory outcomes:

In exploratory analyses, major thrombotic events (MTE) and death were analyzed as a composite outcome defined as myocardial infarction, pulmonary embolism, ischemic stroke, systemic arterial embolism, and in-hospital death up to day 90.

###### Analysis populations:

The SAP for the immune modulation domain analysis defines five analysis populations.

1. The primary analysis population includes the REMAP-CAP COVID-19 severe and moderate intent-to-treat (ITT) population
2. The Unblinded ITT population is defined as all severe patients randomized in one or more unblinded domains within the pandemic stratum without a prior randomization in the moderate state. The unblinded ITT population consists of 3848 patients randomized in the Immune Modulation Therapy, Corticosteroid, Antiviral, Anticoagulation, or Immunoglobulin domains. There are 31 patients within this population that are missing values of OSFD and in-hospital mortality.
3. The Unblinded non-negative COVID-19 population is defined as all patients in the Unblinded ITT population after removing those with >1 negative test for COVID-19 and no positive tests. The unblinded ITT population restricted to non-negative COVID-19 consists of 3613 patients. There are 29 patients within this population that are missing values of OSFD and in-hospital mortality.
4. The Immune Modulation ITT population consists of patients in the Unblinded ITT population that were randomized to the Immune Modulation domain within the pandemic stratum. The Immune Modulation ITT population consists of 2226 patients. There are 19 patients within this population that are missing values of OSFD and in-hospital mortality.
5. The Immune Modulation Per Protocol (PP) population consists of the patients in the Immune Modulation ITT population who have been treated as per protocol. The Immune Modulation per protocol population consists of 2107 patients. There are 13 patients within this population that are missing values of OSFD and in-hospital mortality.

20 patients had randomizations in both Moderate and Severe state. All of them had known OSFD outcomes for both states. These patients thus each contribute 2 outcomes to the model.

###### Data management and summaries:

Data management and summaries were created with the use of R software, version 3.6.0; the primary analysis was computed with R software, version 4.0.0, with the use of the rstan package, version 2.21.1. Additional data management and analyses were performed with SQL Server 2016; SPSS software, version 26; and Stata software, version 14.2.

3. Additional results and analyses
   3.1. Site participation and exclusions

*Site Participation in the Immune Modulation Domain:*

During the study period, April 19 2020 to April 10 2021, 127 sites were open for enrolment in the Immune Modulation Therapy domain. There were 4 sites open for recruitment in the Moderate state (3 UK, 1 Australia) and 123 sites for recruitment in the Severe State (110 UK, 11 Netherlands, 3 Ireland, 2 Australia, 2 New-Zealand, 1 Canada, 1 Finland, 1 Italy, 1 Saudi-Arabia).

###### Platform exclusions:

From the patient flow (Figure 1 main manuscript):

**7695 participants were excluded** from the REMAP-CAP platform:

- **5963** participants were ineligible for the platform
- Of **1089** participants, the site was not active for COVID-19 Immune Modulation Therapy domain and they were not enrolled in another domain
- Of **643** participants, the site was active for the COVID-19 Immune Modulation Therapy domain and they were not enrolled in another domain. They were excluded from the Immune Modulation Therapy domain for the following reasons (participants could be excluded for more than one reason):
  - 4 COVID-19 not confirmed, or testing not done and not intended
  - 237 More than 24 hours since ICU admission
  - 41 Received an immune modulator during this hospital admission
  - 3 Receiving long-term therapy with an immune modulating agent
  - 65 Known condition resulting in immune suppression
  - 20 Enrolled in another trial
  - 89 Contraindication to agents in domain ^
  - 64 Not considered in patient’s best interests
  - 246 Prospective consent declined or not obtained
  3.2. Additional analyses
       3.2.1. Statistical analysis plan

The full additional analyses and data specified in the statistical analysis plan are provided in the “REMAP-CAP Unblinded Analysis Report” at the end of this supplement (Appendix 1).

3.2.2. Summary of results of interferon-β1a

It is important to note that because any clinical use of corticosteroids was an exclusion criterion for interferon-ß1a assignment, recruitment was extremely low after corticosteroids became standard of care (June 17, 2020). The full data on the interferon-ß1a treatment arm is provided in the “ITSC Secondary Analysis Report” at the end of this supplement.

A total of 21 patients were randomized to interferon-ß1a treatment. After withdrawal of consent for two patients, results are available for 19 patients. Median organ support-free days was 0 (IQR -1,13.50) for the interferon-ß1a treatment arm and 0 (IQR -1, 15) for the control group. Compared to control, the median adjusted odds ratio (primary model) for interferon-ß1a was 1.67 (95%CrI 0.49, 5.73) yielding a 79.4% posterior probability of superiority. The hospital mortality was 36.8% for interferon-ß1a and 36.9% for control. Compared with control, median adjusted odds ratio for hospital survival was 1.72 (95%CrI 0.48, 6.58) for interferon-ß1a yielding a 77.4% posterior probability of being superior to control.

3.2.3. Moderate State participants

Only five participants were randomized to an intervention in the Immune Modulation Therapy domain (three to control and two to anakinra). For Moderate State participants progressing to Severe State, and who had already received an allocation in the Moderate State, commencement of organ support in ICU was used instead of date and time of ICU admission to indicate the transition in State. Baseline characteristics of these patients are shown in Table S1b. These patients were included in the primary model, but not in any secondary model. The results are not presented separately.

#### Table S1: Baseline Characteristics by disease state

| **Table S1a. Participant Characteristics at Baseline (Severe State)*** | | | | | | |
| --- | --- | --- | --- | --- | --- | --- |
|  | **Tocilizumab**  **(*n* = 952)** | **Sarilumab**  **(*n* = 485)** | **Anakinra**  **(*n* = 373)** | **Interferon-β-1a**  **(*n* = 19)** | **Control**  **(*n* = 406)** | **All Patients**  **(*n* = 2235)** |
| Age - mean (SD), years | 60.8 (12.2) | 59.0 (13.2) | 59.8 (11.9) | 64.1 (8.7) | 61.1 (12.9) | 60.3 (12.5) |
| Male sex - n (%) | 656 (68.9) | 326 (67.2) | 269 (72.1) | 14 (73.7) | 285 (70.2) | 1550 (69.4) |
| Race / Ethnicity ^a^ - n / N (%) |  |  |  |  |  |  |
| White | 515 / 724 (71.1) | 368 / 453 (81.2) | 184 / 254 (72.4) | 13 / 18 (72.2) | 234 / 317 (73.8) | 1314 / 1766 (74.4) |
| Asian | 123 / 724 (17.0) | 53 / 453 (11.7) | 39 / 254 (15.4) | 3 / 18 (16.7) | 53 / 317 (16.7) | 271 / 1766 (15.3) |
| Black | 38 / 724 (5.2) | 9 / 453 (2.0) | 9 / 254 (3.5) | 1 / 18 (5.6) | 10 / 317 (3.2) | 67 / 1766 (3.8) |
| Mixed | 13 / 724 (1.8) | 1 / 453 (0.2) | 6 / 254 (2.4) | 0 / 18 (0.0) | 6 / 317 (1.9) | 26 / 1766 (1.5) |
| Other | 35 / 724 (4.8) | 22 / 453 (4.9) | 16 / 254 (6.3) | 1 / 18 (5.6) | 14 / 317 (4.4) | 88 / 1766 (5.0) |
| Body-mass index ^b^ - median (IQR), kg/m^2^ | 30.4 (26.6 - 34.9)  (*n* = 862) | 31.2 (27.7 - 36.3)  (*n* = 419) | 29.7 (26.3 - 35.3)  (*n* = 332) | 30.1 (26.9 - 35.1)  (*n* = 19) | 30.9 (27.1 - 34.9)  (*n* = 385) | 30.5 (26.8 - 35.3)  (*n* = 2017) |
| APACHE II score ^c^ - median (IQR) | 13.0 (8.0 - 19.0)  (*n* = 934) | 12.0 (7.0 - 20.0)  (*n* = 475) | 13.0 (8.0 - 19.0)  (*n* = 365) | 13.0 (7.0 - 21.5)  (*n* = 19) | 12.0 (8.0 - 18.0)  (*n* = 394) | 13.0 (8.0 - 19.0)  (*n* = 2187) |
| Confirmed SARS-CoV-2 infection ^d^ - n / N (%) | 802 / 942 (85.1) | 429 / 484 (88.6) | 319 / 369 (86.4) | 15 / 19 (78.9) | 348 / 406 (85.7) | 1913 / 2220 (86.2) |
| Preexisting condition - n / N (%) |  |  |  |  |  |  |
| Diabetes | 281 / 949 (29.6) | 108 / 484 (22.3) | 125 / 370 (33.8) | 7 / 19 (36.8) | 152 / 406 (37.4) | 673 / 2228 (30.2) |
| Respiratory disease | 218 / 949 (23.0) | 117 / 484 (24.2) | 81 / 370 (21.9) | 3 / 19 (15.8) | 100 / 406 (24.6) | 519 / 2228 (23.3) |
| Asthma/COPD | 183 / 949 (19.3) | 99 / 484 (20.5) | 67 / 370 (18.1) | 2 / 19 (10.5) | 89 / 406 (21.9) | 440 / 2228 (19.7) |
| Other | 40 / 949 (4.2) | 26 / 484 (5.4) | 17 / 370 (4.6) | 2 / 19 (10.5) | 17 / 406 (4.2) | 102 / 2228 (4.6) |
| Kidney disease | 66 / 866 (3.2) | 30 / 446 (1.5) | 22 / 340 (1.1) | 1 / 17 (0.0) | 43 / 377 (2.1) | 162 / 2046 (7.9) |
| Severe cardiovascular disease | 86 / 930 (9.2) | 33 / 474 (7.0) | 41 / 367 (11.2) | 1 / 19 (5.3) | 47 / 401 (11.7) | 208 / 2191 (9.5) |
| Any immunosuppressive condition | 25 / 948 (2.6) | 11 / 484 (2.3) | 6 / 370 (1.6) | 0 / 19 (0.0) | 18 / 406 (4.4) | 60 / 2227 (2.7) |
| Cancer | 7 / 948 (0.7) | 3 / 484 (0.6) | 3 / 370 (0.8) | 0 / 19 (0.0) | 10 / 406 (2.5) | 23 / 2227 (1.0) |
| Chronic immunosuppressive therapy | 10 / 949 (1.1) | 8 / 484 (1.7) | 4 / 370 (1.1) | 0 / 19 (0.0) | 7 / 406 (1.7) | 29 / 2228 (1.3) |
| Other | 13 / 948 (1.4) | 2 / 484 (0.4) | 3 / 370 (0.8) | 0 / 19 (0.0) | 5 / 406 (1.2) | 23 / 2227 (1.0) |
| Liver cirrhosis / failure | 3 / 930 (0.3) | 0 / 474 (0.0) | 1 / 370 (0.3) | 1 / 19 (5.3) | 2 / 401 (0.5) | 7 / 2191 (0.3) |
| Time to enrollment - median (IQR) |  |  |  |  |  |  |
| From hospital admission - days | 1.4 (0.9 - 3.3) | 1.6 (0.9 - 3.5) | 1.6 (0.9 - 3.8) | 1.7 (0.9 - 3.6) | 1.2 (0.8 - 2.8) | 1.4 (0.9 - 3.2) |
| From ICU admission - hours | 13.4 (6.9 - 19.1) | 15.1 (7.8 - 19.9) | 13.6 (7.3 - 19.7) | 7.7 (5.3 - 17.3) | 14.0 (6.8 - 19.5) | 14.0 (7.0 - 19.5) |
| Acute respiratory support - n (%) |  |  |  |  |  |  |
| None / supplemental oxygen only | 1 (0.1) | 0 (0.0) | 1 (0.3) | 0 (0.0) | 2 (0.5) | 4 (0.2) |
| High-flow nasal cannula | 226 (23.7) | 96 (19.8) | 101 (27.1) | 3 (15.8) | 110 (27.1) | 536 (24.0) |
| Noninvasive ventilation only | 404 (42.4) | 241 (49.7) | 133 (35.7) | 9 (47.4) | 171 (42.1) | 958 (42.9) |
| Invasive mechanical ventilation | 320 (33.6) | 148 (30.5) | 138 (37.0) | 7 (36.8) | 122 (30.0) | 735 (32.9) |
| ECMO | 1 (0.1) | 0 (0.0) | 0 (0.0) | 0 (0.0) | 1 (0.2) | 2 (0.1) |
| Vasopressor support - n (%) | 179 (18.8) | 77 (15.9) | 81 (21.7) | 5 (26.3) | 79 (19.5) | 421 (18.8) |
| Acute kidney replacement therapy - n / N (%) | 3 / 948 (0.3) | 2 / 483 (0.4) | 1 / 370 (0.3) | 0 / 19 (0.0) | 2 / 406 (0.5) | 8 / 2226 (0.4) |
| PaO_2_ / FiO_2_ - median (IQR) | 110 (86 - 148)  (*n* = 872) | 116 (89 - 152)  (*n* = 430) | 106 (84 - 148)  (*n* = 330) | 110 (101 - 127)  (*n* = 19) | 118 (89 - 169.5)  (*n* = 359) | 113 (87 - 153)  (*n* = 2010) |
| Glasgow coma scale score ^e^ - median (IQR) | 15 (15 - 15)  (*n* = 857) | 15 (15 - 15)  (*n* = 423) | 15 (15 - 15)  (*n* = 332) | 15 (14.2 - 15)  (*n* = 18) | 15 (15 - 15)  (*n* = 385) | 15 (15 - 15)  (*n* = 2015) |
| Median laboratory values (IQR) ^f^ |  |  |  |  |  |  |
| C-reactive protein, µg/mL | 132 (69 - 201)  (*n* = 783) | 120 (70 - 199)  (*n* = 419) | 112 (70 - 189)  (*n* = 324) | 111 (63 - 151)  (*n* = 14) | 129 (71 - 208)  (*n* = 255) | 124 (70 - 198)  (*n* = 1795) |
| D-dimer, µg/L | 946 (483 - 2475)  (*n* = 564) | 947 (420 - 2216)  (*n* = 304) | 1006 (460 - 2363)  (*n* = 256) | 927 (554 - 3049)  (*n* = 10) | 1010 (500 - 2115)  (*n* = 175) | 980 (472 - 2321)  (*n* = 1309) |
| Ferritin, ng/mL | 1111 (618 - 1793)  (*n* = 557) | 990 (584 - 1607)  (*n* = 267) | 918 (550 - 1703)  (*n* = 205) | 1474 (474 - 1982)  (*n* = 11) | 887 (414 - 1603)  (*n* = 204) | 1006 (548 - 1705)  (*n* = 1244) |
| Neutrophils, x10^9^/L | 8.0 (5.6 - 10.9)  (*n* = 814) | 7.7 (5.4 - 10.7)  (*n* = 469) | 7.8 (5.7 - 10.7)  (*n* = 344) | 10.7 (6.8 - 12.9)  (*n* = 14) | 7.9 (5.3 - 11.0)  (*n* = 288) | 7.9 (5.5 - 10.9)  (*n* = 1929) |
| Lymphocytes, x10^9^/L | 0.7 (0.5 - 1.0)  (*n* = 816) | 0.7 (0.5 - 1.0)  (*n* = 466) | 0.7 (0.5 - 1.0)  (*n* = 345) | 0.6 (0.6 - 1.1)  (*n* = 14) | 0.7 (0.5 - 1.0)  (*n* = 288) | 0.7 (0.5 - 1.0)  (*n* = 1929) |
| Platelet count, x10^9^/L | 241 (180 - 311)  (*n* = 939) | 238 (185 - 308)  (*n* = 481) | 252 (195 - 331)  (*n* = 370) | 240 (173 - 306)  (*n* = 19) | 235 (177 - 296)  (*n* = 404) | 241 (183 - 310)  (*n* = 2213) |
| Lactate, mmol/L | 1.3 (1.0 - 1.7)  (*n* = 850) | 1.3 (1.0 - 1.7)  (*n* = 418) | 1.3 (1.0 - 1.7)  (*n* = 334) | 1.2 (1.0 - 1.6)  (*n* = 18) | 1.4 (1.0 - 1.9)  (*n* = 359) | 1.3 (1.0 - 1.7)  (*n* = 1979) |
| Creatinine, mg/dL | 0.9 (0.7 - 1.1)  (*n* = 942) | 0.8 (0.7 - 1.1)  (*n* = 479) | 0.8 (0.7 - 1.1)  (*n* = 370) | 0.8 (0.7 - 1.2)  (*n* = 19) | 0.9 (0.7 - 1.2)  (*n* = 404) | 0.8 (0.7 - 1.1)  (*n* = 2214) |
| Bilirubin, mg/dL | 0.5 (0.4 - 0.8)  (*n* = 910) | 0.6 (0.4 - 0.8)  (*n* = 463) | 0.6 (0.4 - 0.8)  (*n* = 364) | 0.8 (0.6 - 1.1)  (*n* = 19) | 0.5 (0.4 - 0.8)  (*n* = 389) | 0.6 (0.4 - 0.8)  (*n* = 2145) |
| Received therapies at randomization - n / N (%) |  |  |  |  |  |  |
| Steroids | 770 / 938 (82.1) | 422 / 472 (89.4) | 317 / 369 (85.9) | 11 / 19 (57.9) | 269 / 402 (66.9) | 1789 / 2200 (81.3) |
| Remdesivir | 272 / 938 (29.0) | 140 / 472 (29.7) | 109 / 369 (29.5) | 3 / 19 (15.8) | 105 / 402 (26.1) | 629 / 2200 (28.6) |

* Percentages may not sum to 100 because of rounding. SD denotes standard deviation; APACHE, Acute Physiology and Chronic Health Evaluation; SARS-CoV-2, Severe Acute Respiratory Syndrome Coronavirus; ICU, intensive care unit; COPD, chronic obstructive pulmonary disease; IQR, interquartile range; ECMO, extracorporeal membrane oxygenation.

^a^ Data collection not approved in Canada and continental Europe. “Other” includes “declined” and “multiple”.

^b^ Body-mass index is the weight in kilograms divided by the square of the height in meters.

^c^ Range 0 to 71, with higher scores indicating greater severity of illness.

^d^ SARS-CoV2 infection was confirmed by respiratory tract polymerase chain reaction test.

^e^ Range 3 to 15, with higher scores indicating greater consciousness, using values closest to randomization but prior to the use of sedative agents.

^f^ Values were from the sample collected closest to randomization, up to 8 hours prior to randomization. If no samples were collected up to 8 hours prior to time of randomization, the sample collected closest to the time of randomization up to 2 hours after randomization was used (other than PaO_2_ / FiO_2_ which was a pre-randomization value only)

| **Table S1b. Participant Characteristics at Baseline (Moderate State)*** | | | |
| --- | --- | --- | --- |
|  | **Control**  **(*n* = 3)** | **Anakinra**  **(*n* = 2)** | **All Patients**  **(*n* = 5)** |
| Age - mean (SD), years | 67.0 (13.7) | 36.0 (17.0) | 54.6 (21.3) |
| Male sex - n (%) | 1 (33.3) | 1 (50.0) | 2 (40.0) |
| Race / Ethnicity ^a^ - n / N (%) |  |  |  |
| White | 3 / 3 (100.0) | 2 / 2 (100.0) | 5 / 5 (100.0) |
| Asian | 0 / 3 (0.0) | 0 / 2 (0.0) | 0 / 5 (0.0) |
| Black | 0 / 3 (0.0) | 0 / 2 (0.0) | 0 / 5 (0.0) |
| Mixed | 0 / 3 (0.0) | 0 / 2 (0.0) | 0 / 5 (0.0) |
| Other | 0 / 3 (0.0) | 0 / 2 (0.0) | 0 / 5 (0.0) |
| Body-mass index ^b^ - median (IQR), kg/m^2^ | 32.0 (31.9 - 36.1)  (*n* = 3) | 30.4 (28.0 - 32.7)  (*n* = 2) | 32.0 (31.8 - 35.1)  (*n* = 5) |
| APACHE II score ^c^ - median (IQR) | 13.0 (13.0 - 18.0)  (*n* = 3) | 6.5 (5.8 - 7.2)  (*n* = 2) | 13.0 (8.0 - 13.0)  (*n* = 5) |
| Confirmed SARS-CoV-2 infection ^d^ - n / N (%) | 3 / 3 (100.0) | 2 / 2 (100.0) | 5 / 5 (100.0) |
| Preexisting condition - n / N (%) |  |  |  |
| Diabetes | 0 / 3 (0.0) | 0 / 2 (0.0) | 0 / 5 (0.0) |
| Respiratory disease | 1 / 3 (33.3) | 0 / 2 (0.0) | 1 / 5 (20.0) |
| Asthma/COPD | 1 / 3 (33.3) | 0 / 2 (0.0) | 1 / 5 (20.0) |
| Other | 0 / 3 (0.0) | 0 / 2 (0.0) | 0 / 5 (0.0) |
| Kidney disease | 0 / 3 (0.0) | 0 / 2 (0.0) | 0 / 5 (0.0) |
| Severe cardiovascular disease | 0 / 3 (0.0) | 0 / 2 (0.0) | 0 / 5 (0.0) |
| Any immunosuppressive condition | 0 / 3 (0.0) | 0 / 2 (0.0) | 0 / 5 (0.0) |
| Cancer | 0 / 3 (0.0) | 0 / 2 (0.0) | 0 / 5 (0.0) |
| Chronic immunosuppressive therapy | 0 / 3 (0.0) | 0 / 2 (0.0) | 0 / 5 (0.0) |
| Other | 0 / 3 (0.0) | 0 / 2 (0.0) | 0 / 5 (0.0) |
| Liver cirrhosis / failure | 0 / 3 (0.0) | 0 / 2 (0.0) | 0 / 5 (0.0) |
| Time to enrollment - median (IQR) |  |  |  |
| From hospital admission - days | 0.9 (0.8 - 5.4) | 2.7 (2.4 - 3.1) | 2.0 (0.9 - 3.4) |
| From ICU admission - hours | 14.3 (13.7 - 17.4) | 24.5 (23.5 - 25.5) | 20.4 (14.3 - 22.5) |
| Acute respiratory support - n (%) |  |  |  |
| None / supplemental oxygen only | 3 (100.0) | 2 (100.0) | 5 (100.0) |
| High-flow nasal cannula | 0 (0.0) | 0 (0.0) | 0 (0.0) |
| Noninvasive ventilation only | 0 (0.0) | 0 (0.0) | 0 (0.0) |
| Invasive mechanical ventilation | 0 (0.0) | 0 (0.0) | 0 (0.0) |
| ECMO | 0 (0.0) | 0 (0.0) | 0 (0.0) |
| Vasopressor support - n (%) | 0 (0.0) | 0 (0.0) | 0 (0.0) |
| Acute kidney replacement therapy - n / N (%) | 0 / 3 (0.0) | 0 / 2 (0.0) | 0 / 5 (0.0) |
| PaO_2_ / FiO_2_ - median (IQR) | 131 (114.5 - 263.5)  (*n* = 3) | 106 (106 - 106)  (*n* = 1) | 118 (104 - 197)  (*n* = 4) |
| Glasgow coma scale score ^e^ - median (IQR) | 15 (12 - 15)  (*n* = 3) | 15 (15 - 15)  (*n* = 2) | 15 (15 - 15)  (*n* = 5) |
| Median laboratory values (IQR) ^f^ |  |  |  |
| C-reactive protein, µg/mL | 148 (148 - 148)  (*n* = 1) | 69 (69 - 69)  (*n* = 1) | 109 (89 - 128)  (*n* = 2) |
| D-dimer, µg/L | ---  (*n* = 0) | 130 (130 - 130)  (*n* = 1) | 130 (130 - 130)  (*n* = 1) |
| Ferritin, ng/mL | ---  (*n* = 0) | 784 (784 - 784)  (*n* = 1) | 784 (784 - 784)  (*n* = 1) |
| Neutrophils, x10^9^/L | 12.5 (10.3 - 12.7)  (*n* = 3) | 4.8 (4.7 - 5.0)  (*n* = 2) | 8.1 (5.1 - 12.5)  (*n* = 5) |
| Lymphocytes, x10^9^/L | 1.7 (1.4 - 3.5)  (*n* = 3) | 1.8 (1.6 - 2.0)  (*n* = 2) | 1.7 (1.3 - 2.3)  (*n* = 5) |
| Platelet count, x10^9^/L | 262 (246 - 272)  (*n* = 3) | 301 (280 - 321)  (*n* = 2) | 262 (260 - 282)  (*n* = 5) |
| Lactate, mmol/L | 1.0 (0.9 - 1.0)  (*n* = 3) | 0.9 (0.9 - 0.9)  (*n* = 1) | 0.9 (0.9 - 1.0)  (*n* = 4) |
| Creatinine, mg/dL | 0.6 (0.6 - 0.8)  (*n* = 3) | 0.7 (0.6 - 0.7)  (*n* = 2) | 0.6 (0.6 - 0.7)  (*n* = 5) |
| Bilirubin, mg/dL | 1.1 (0.8 - 1.4)  (*n* = 3) | 0.3 (0.3 - 0.3)  (*n* = 1) | 0.8 (0.6 - 1.2)  (*n* = 4) |
| Received therapies at randomization - n / N (%) |  |  |  |
| Steroids | 1 / 3 (33.3) | 2 / 2 (100.0) | 3 / 5 (60.0) |
| Remdesivir | 0 / 3 (0.0) | 1 / 2 (50.0) | 1 / 5 (20.0) |

* Percentages may not sum to 100 because of rounding. SD denotes standard deviation; APACHE, Acute Physiology and Chronic Health Evaluation; SARS-CoV-2, Severe Acute Respiratory Syndrome Coronavirus; ICU, intensive care unit; COPD, chronic obstructive pulmonary disease; IQR, interquartile range; ECMO, extracorporeal membrane oxygenation.

^a^ Data collection not approved in Canada and continental Europe. “Other” includes “declined” and “multiple”.

^b^ Body-mass index is the weight in kilograms divided by the square of the height in meters.

^c^ Range 0 to 71, with higher scores indicating greater severity of illness.

^d^ SARS-CoV2 infection was confirmed by respiratory tract polymerase chain reaction test.

^e^ Range 3 to 15, with higher scores indicating greater consciousness, using values closest to randomization but prior to the use of sedative agents.

^f^ Values were from the sample collected closest to randomization, up to 8 hours prior to randomization. If no samples were collected up to 8 hours prior to time of randomization, the sample collected closest to the time of randomization up to 2 hours after randomization was used (other than PaO_2_ / FiO_2_ which was a pre-randomization value only)

#### Table S2. Co-enrolment into other REMAP-CAP domains for Severe State participants enrolled in Immune Modulation Therapy domain

| **Domain** | **Tocilizumab (N=947)** | **Sarilumab (N=482)** | **Anakinra (N=373)** | **Interferon-β1a**  **(N=19)** | **Control (N=405)** |
| --- | --- | --- | --- | --- | --- |
| *Corticosteroid domain* | | | | | |
| No hydrocortisone, N | 16 | 0 | 0 | 1 | 9 |
| Fixed dose hydrocortisone, N | 891 | 482 | 372 | 15 | 350 |
| Hydrocortisone for shock, N | 19 | 0 | 1 | 1 | 18 |
| *Covid-19 Antiviral domain* | | | | | |
| No antiviral, N | 91 | 16 | 6 | 2 | 115 |
| Lopinavir/ritonavir, N | 63 | 10 | 11 | 5 | 86 |
| Hydroxychloroquine, N | 10 | 0 | 1 | 0 | 17 |
| Lopinavir/ritonavir plus hydroxychloroquine, N | 5 | 0 | 0 | 0 | 3 |
| *Covid-19 Immunoglobulin, N* | | | | | |
| No convalescent plasma, N | 208 | 119 | 39 | 7 | 95 |
| Convalescent plasma, N | 252 | 143 | 52 | 4 | 108 |
| *Therapeutic Anticoagulation, N* | | | | | |
| Thromboprophylaxis, N | 80 | 54 | 18 | 3 | 71 |
| Therapeutic anticoagulation, N | 90 | 40 | 22 | 7 | 77 |

#### Table S3. Sensitivity analyses of the primary outcomes

| **Outcome/Analysis** | **Tocilizumab (N=947)** | **Sarilumab (N=482)** | **Anakinra (N=373)** | **Interferon-β1a**  **(N=19)** | **Control (N=405)** |
| --- | --- | --- | --- | --- | --- |
| **Secondary Analysis of Primary Outcome**, for Unblinded ITT population with nesting between IL-6ra intervention effects | | | | | |
| Adjusted OR - mean (SD) | 1.52 (0.19) | 1.57 (0.20) | 1.07 (0.17) | 1.37 (0.56) | 1 |
| - median (95% CrI) | 1.51 (1.18 to 1.93) | 1.56 (1.19 to 2.02) | 1.06 (0.77 to 1.44) | 1.27 (0.58 to 2.75) | 1 |
| Probability of superiority to control, % | >99.9 | 99.9 | 64.7 | 72.8 | - |
| **Secondary Analysis of Hospital Survival**, for Unblinded ITT population with nesting between IL-6ra intervention effects | | | | | |
| Adjusted OR - mean (SD) | 1.47 (0.23) | 1.55 (0.27) | 1.03 (0.20) | 1.42 (0.72) | 1 |
| - median (95% CrI) | 1.45 (1.07 to 1.97) | 1.53 (1.11 to 2.15) | 1.01 (0.70 to 1.49) | 1.26 (0.52 to 3.22) | 1 |
| Probability of superiority to control, % | 99.3 | 99.6 | 52.4 | 68.9 | - |
| **Secondary Analysis of Primary Outcome**, for Unblinded ITT population with independent IL-6ra intervention effects | | | | | |
| Adjusted OR - mean (SD) | 1.50 (0.19) | 1.59 (0.22) | 1.07 (0.17) | 1.36 (0.55) | 1 |
| - median (95% CrI) | 1.49 (1.16 to 1.91) | 1.57 (1.20 to 2.06) | 1.06 (0.78 to 1.44) | 1.26 (0.58 to 2.65) | 1 |
| Probability of superiority to control, % | 99.9 | 99.9 | 64.6 | 71.9 | - |
| **Secondary Analysis of Hospital Survival**, for Unblinded ITT population with independent IL-6ra intervention effects | | | | | |
| Adjusted OR - mean (SD) | 1.45 (0.23) | 1.59 (0.29) | 1.03 (0.20) | 1.42 (0.72) | 1 |
| - median (95% CrI) | 1.43 (1.05 to 1.95) | 1.57 (1.10 to 2.22) | 1.01 (0.70 to 1.49) | 1.26 (0.51 to 3.27) | 1 |
| Probability of superiority to control, % | 99.0 | 99.3 | 52.9 | 69.1 | - |
| **Secondary Analysis of Primary Outcome**, for Unblinded ITT population with pooled IL-6ra intervention effects* | | | | | |
| Adjusted OR - mean (SD) | 1.53 (0.19) | | 1.07 (0.17) | 1.36 (0.56) | 1 |
| - median (95% CrI) | 1.52 (1.19 to 1.94) | | 1.05 (0.78 to 1.45) | 1.26 (0.58 to 2.69) | 1 |
| Probability of superiority to control, % | >99.9 | | 63.9 | 72.4 | - |
| **Secondary Analysis of Hospital Survival**, for Unblinded ITT population with pooled IL-6ra intervention effects* | | | | | |
| Adjusted OR - mean (SD) | 1.49 (0.23) | | 1.03 (0.20) | 1.42 (0.72) | 1 |
| - median (95% CrI) | 1.48 (1.09 to 2.00) | | 1.01 (0.69 to 1.47) | 1.27 (0.51 to 3.25) | 1 |
| Probability of superiority to control, % | 99.4 | | 51.6 | 69.2 | - |
| **Secondary Analysis of Primary Outcome**, for Unblinded ITT population with additional unblinded interaction effects* | | | | | |
| Adjusted OR - mean (SD) | 1.45 (0.40) | | 1.08 (0.19) | 1.35 (0.55) | 1 |
| - median (95% CrI) | 1.39 (0.82 to 2.38) | | 1.07 (0.76 to 1.49) | 1.25 (0.56 to 2.68) | 1 |
| Probability of superiority to control, % | 88.9 | | 64.9 | 71.5 | - |
| **Secondary Analysis of Hospital Survival**, for Unblinded ITT population with additional unblinded interaction effects* | | | | | |
| Adjusted OR - mean (SD) | 2.11 (0.90) | | 1.05 (0.23) | 1.41 (0.72) | 1 |
| - median (95% CrI) | 1.93 (0.90 to 4.35) | | 1.02 (0.68 to 1.56) | 1.26 (0.50 to 3.26) | 1 |
| Probability of superiority to control, % | 95.4 | | 53.9 | 68.6 | - |
| **Secondary Analysis of Primary Outcome**, for Unblinded ITT population restricted to non-negative COVID-19 | | | | | |
| Adjusted OR - mean (SD) | 1.55 (0.20) | 1.69 (0.25) | 1.11 (0.18) | 1.08 (0.50) | 1 |
| - median (95% CrI) | 1.53 (1.18 to 1.99) | 1.68 (1.26 to 2.23) | 1.10 (0.80 to 1.51) | 0.99 (0.41 to 2.31) | 1 |
| Probability of superiority to control, % | 99.9 | >99.9 | 72.1 | 49.3 | - |
| **Secondary Analysis of Hospital Survival**, for Unblinded ITT population restricted to non-negative COVID-19 | | | | | |
| Adjusted OR - mean (SD) | 1.48 (0.24) | 1.68 (0.31) | 1.06 (0.21) | 1.04 (0.58) | 1 |
| - median (95% CrI) | 1.46 (1.06 to 2.00) | 1.65 (1.15 to 2.37) | 1.04 (0.70 to 1.52) | 0.90 (0.33 to 2.51) | 1 |
| Probability of superiority to control, % | 99.0 | 99.8 | 57.6 | 42.1 | - |
| **Secondary Analysis of Primary Outcome**, for Immune Modulation Therapy ITT population | | | | | |
| Adjusted OR - mean (SD) | 1.55 (0.19) | 1.67 (0.25) | 1.16 (0.19) | 1.29 (0.53) | 1 |
| - median (95% CrI) | 1.54 (1.22 to 1.95) | 1.65 (1.23 to 2.22) | 1.14 (0.83 to 1.57) | 1.20 (0.55 to 2.59) | 1 |
| Probability of superiority to control, % | >99.9 | >99.9 | 79.6 | 68.3 | - |
| **Secondary Analysis of Hospital Survival**, for Immune Modulation Therapy ITT population | | | | | |
| Adjusted OR - mean (SD) | 1.55 (0.25) | 1.78 (0.23) | 1.14 (0.23) | 1.42 (0.71) | 1 |
| - median (95% CrI) | 1.53 (1.13 to 2.09) | 1.74 (1.21 to 2.54) | 1.12 (0.76 to 1.66) | 1.27 (0.51 to 3.19) | 1 |
| Probability of superiority to control, % | 99.8 | 99.9 | 70.9 | 69.4 | - |
| **Secondary Analysis of Primary Outcome**, for Unblinded ITT population with site and time factors removed | | | | | |
| Adjusted OR - mean (SD) | 1.42 (0.17) | 1.39 (0.18) | 0.99 (0.14) | 1.12 (0.46) | 1 |
| - median (95% CrI) | 1.41 (1.12 to 1.79) | 1.37 (1.07 to 1.77) | 0.98 (0.74 to 1.30) | 1.03 (0.48 to 2.23) | 1 |
| Probability of superiority to control, % | 99.8 | 99.2 | 43.6 | 53.6 | - |
| **Secondary Analysis of Hospital Survival**, for Unblinded ITT population with site and time factors removed | | | | | |
| Adjusted OR - mean (SD) | 1.37 (0.20) | 1.37 (0.22) | 0.97 (0.17) | 1.26 (0.62) | 1 |
| - median (95% CrI) | 1.35 (1.02 to 1.80) | 1.35 (1.00 to 1.88) | 0.95 (0.68 to 1.33) | 1.13 (0.47 to 2.87) | 1 |
| Probability of superiority to control, % | 98.1 | 97.5 | 38.7 | 60.3 | - |

* results are provided for the pooled IL-6ra

#### Table S4. Secondary outcomes

| **Outcome/Analysis** | **Tocilizumab (N=947)** | **Sarilumab (N=482)** | | **Anakinra (N=373)** | **Interferon-β1a (N=19)** | **Control (N=405)** |
| --- | --- | --- | --- | --- | --- | --- |
| **90-day Survival (time to event)** |  | | | | | |
| Adjusted HR - mean (SD) | 1.40 (0.16) | 1.46 (0.20) | | 1.15 (0.16) | 1.26 (0.51) | 1 |
| - median (95% CrI) | 1.39 (1.11 to 1.74) | 1.44 (1.11 to 1.89) | | 1.13 (0.87 to 1.49) | 1.16 (0.60 to 2.52) | 1 |
| Probability of superiority to control, % | 99.9 | 99.6 | | 82.3 | 65.8 | - |
| **Freedom from progression to intubation, ECMO, or death** |  |  | |  |  |  |
| Adjusted OR - mean (SD) | 1.70 (0.29) | 1.46 (0.29) | | 1.19 (0.26) | 1.33 (0.79) | 1 |
| - median (95% CrI) | 1.67 (1.20 to 2.36) | 1.43 (0.98 to 2.10) | | 1.16 (0.76 to 1.78) | 1.15 (0.39 to 3.35) | 1 |
| Probability of superiority to control, % | 99.9 | 96.9 | | 75.0 | 59.6 | - |
| **Respiratory support-free days** |  |  | |  |  |  |
| Median (IQR) | 6 (-1,16) | 8 (-1,16.5) | | 0 (-1,15) | 0 (-1,13) | 0 (-1,14) |
| Adjusted OR - mean (SD) | 1.52 (0.19) | 1.61 (0.23) | | 1.07 (0.17) | 1.36 (0.55) | 1 |
| - median (95% CrI) | 1.51 (1.18 to 1.92) | 1.60 (1.21 to 2.09) | | 1.06 (0.78 to 1.43) | 1.27 (0.58 to 2.72) | 1 |
| Probability of superiority to control, % |  |  | |  |  | - |
| Days free from respiratory support in survivors, median (IQR) | 14 (6,18) | 15 (8,18) | | 13.5 (3.75,18) | 9.5 (0,17.25) | 13 (3.5,17) |
| **Cardiovascular support-free days** |  | | | | | |
| Median (IQR) | 10 (-1,21) | 18 (-1,21) | | 10 (-1,21) | 7 (-1,19) | 15 (-1,21) |
| Adjusted OR - mean (SD) | 1.46 (0.19) | 1.60 (0.24) | | 1.09 (0.18) | 1.08 (0.47) | 1 |
| - median (95% CrI) | 1.44 (1.12 to 1.87) | 1.58 (1.19 to 2.12) | | 1.07 (0.78 to 1.47) | 0.99 (0.44 to 2.20) | 1 |
| Probability of superiority to control, % | 99.8 | 99.9 | | 66.6 | 49.3 | - |
| Days free from cardiovascular support in survivors, median (IQR) | 21 (16,21) | 21 (18,21) | | 21 (15,21) | 16.5 (7.75,21) | 21 (17,21) |
| **Time to ICU discharge** | | |  | | | |
| Adjusted HR - mean (SD) | 1.30 (0.11) | 1.42 (0.14) | | 1.10 (0.12) | 1.21 (0.33) | 1 |
| - median (95% CrI) | 1.29 (1.10 to 1.52) | 1.41 (1.16 to 1.72) | | 1.10 (0.89 to 1.36) | 1.17 (0.65 to 1.95) | 1 |
| Probability of superiority to control, % | 99.9 | >99.9 | | 80.2 | 71 | - |
| **Time to hospital discharge** | | |  | | | |
| Adjusted HR - mean (SD) | 1.31 (0.11) | 1.31 (0.14) | | 1.05 (0.12) | 1.18 (0.34 | 1 |
| - median (95% CrI) | 1.31 (1.11 to 1.55) | 1.30 (1.06 to 1.59) | | 1.04 (0.84 to 1.30) | 1.15 (0.62 to 1.94) | 1 |
| Probability of superiority to control, % | > 99.9 | 99.4 | | 64.7 | 68.6 | - |
| **WHO scale at day 14** | | |  | | | |
| Adjusted OR - mean (SD) | 1.54 (0.19) | 1.67 (0.24) | | 1.19 (0.18) | 1.22 (0.50) | 1 |
| - median (95% CrI) | 1.53 (1.20 to 1.95) | 1.65 (1.25 to 2.18) | | 1.18 (0.87 to 1.60) | 1.13 (0.52 to 2.46) | 1 |
| Probability of superiority to control, % | >99.9 | >99.9 | | 85.6 | 62 | - |
| **Progression to invasive mechanical ventilation, ECMO or death, restricted to those not intubated at baseline** | | |  | | | |
| Free of invasive mechanical ventilation at baseline, n | 613 | 331 | | 228 | 12 | 276 |
| Progression to intubation, ECMO or death, n (%) | 266/613 (43.4) | 149/331 (45) | | 122/228 (53.5) | 6/12 (50) | 147/276 (53.3) |
| Adjusted OR - mean (SD) | 1.70 (0.29) | 1.46 (0.29) | | 1.19 (0.26) | 1.33 (0.79) | 1 |
| - median (95% CrI) | 1.67 (1.20 to 2.36) | 1.43 (0.98 to 2.10) | | 1.16 (0.76 to 1.78) | 1.15 (0.39 to 3.35) | 1 |
| Probability of superiority to control, % | 99.9 | 96.9 | | 75.0 | 59.6 | - |
| Individual components of progression * |  | | | | | |
| Intubation, n (%) | 214 (34.9) | 110 (33.2) | | 103 (45.2) | 6 (50) | 121 (43.8) |
| ECMO, n (%) | 6 (1) | 3 (0.9) | | 2 (0.9) | 0 (0) | 3 (1.1) |
| Death, n (%) | 173 (28.2) | 102 (30.8) | | 75 (32.9) | 3 (25) | 88 (31.9) |
| **Secondary analysis of major thrombotic events or death*** |  | | | | | |
| Adjusted OR - mean (SD) | 1.78 (0.34) | | | 1.01 (0.29) | 1.63 (1.00) | 1 |
| - median (95% CrI) | 1.75 (1.22 to 2.55) | | | 0.97 (0.57 to 1.69) | 1.38 (0.51 to 4.17) | 1 |
| Probability of superiority to control, % | 99.9 | | | 46.0 | 73.0 | - |
| **Secondary analysis of major thrombotic events*** |  | | | | | |
| Adjusted OR - mean (SD) | 1.24 (0.37) | | | 1.79 (0.91) | 3.07 (3.07) | 1 |
| - median (95% CrI) | 1.19 (0.68 to 2.09) | | | 1.58 (0.66 to 4.10) | 2.16 (0.52 to 11.15) | 1 |
| Probability of superiority to control, % | 72.9 | | | 84.4 | 84.3 | - |
| **Secondary analysis of major bleeding events*** |  | | | | | |
| Adjusted OR - mean (SD) | 1.46 (0.93) | | | 1.31 (1.26) | 0.88 (0.96) | 1 |
| - median (95% CrI) | 1.24 (0.39 to 3.89) | | | 0.96 (0.23 to 4.44) | 0.59 (0.12 to 3.47) | 1 |
| Probability of superiority to control, % | 63.7 | | | 47.4 | 27.0 | - |

* results are provided for the pooled IL-6ra

#### Table S5. Primary safety analysis of serious adverse events

|  | **Tocilizumab (N=947)** | **Sarilumab (N=482)** | | **Anakinra (N=373)** | **Interferon-β1a (N=19)** | **Control (N=405)** |
| --- | --- | --- | --- | --- | --- | --- |
| **For the Immune Modulation ITT Population** | | | | | | |
| Adjusted OR - mean (SD) | 1.15 (0.41) | | 1.68 (0.79) | 1.32 (0.60) | 1.26 (1.16) | 1 |
| - median (95% CrI) | 1.08 (0.53 to 2.15) | | 1.52 (0.65 to 3.66) | 1.20 (0.51 to 2.77) | 0.92 (0.22 to 4.33) | 1 |
| Probability of superiority to control, % | 59.2 | | 84.0 | 66.9 | 45.6 | - |

ORs above 1 indicate fewer SAEs, and ORs below 1 indicate more SAEs.

#### Table S6. Predefined subgroup analysis of the primary outcomes by CRP terciles

| **Outcome/Analysis** | **Tocilizumab (N=947)** | **Sarilumab (N=482)** | **Anakinra (N=373)** | **Control (N=405)** |
| --- | --- | --- | --- | --- |
| **Secondary Analysis of Primary Outcome**, model restricted to Immune Modulation Therapy Domain participants, according to **CRP tercile subgroups** | | | | |
| **CRP lowest tercile**  Adjusted OR - mean (SD) | 1.30 (0.23) | 1.42 (0.30) | 1.04 (0.25) | 1 |
| - median (95% CrI) | 1.28 (0.91 to 1.82) | 1.39 (0.93 to 2.09) | 1.01 (0.64 to 1.60) | 1 |
| Probability of superiority to control, % | 92.4 | 94.4 | 52.5 | - |
| **CRP middle tercile**  Adjusted OR - mean (SD) | 1.33 (0.23) | 1.37 (0.28) | 0.92 (0.20) | 1 |
| - median (95% CrI) | 1.31 (0.95 to 1.83) | 1.34 (0.91 to 2.01) | 0.90 (0.58 to 1.38) | 1 |
| Probability of superiority to control, % | 94.8 | 93.3 | 31 | - |
| **CRP highest tercile**  Adjusted OR - mean (SD) | 1.90 (0.32) | 1.88 (0.38) | 1.34 (0.32) | 1 |
| - median (95% CrI) | 1.87 (1.35 to 2.59) | 1.85 (1.24 to 2.69) | 0.83 to 2.05) | 1 |
| Probability of superiority to control, % | >99.9 | 99.8 | 87.1 | - |
| **Secondary Analysis of Hospital Survival,** model restricted to Immune Modulation Therapy Domain participants, according to CRP tercile subgroups | | | | |
| **CRP lowest tercile**  Adjusted OR - mean (SD) | 1.09 (0.24) | 1.33 (0.35) | 0.85 (0.24) | 1 |
| - median (95% CrI) | 1.07 (0.70 to 1.63) | 1.29 (0.79 to 2.15) | 0.82 (0.48 to 1.41) | 1 |
| Probability of superiority to control, % | 62.4 | 84 | 23.6 | - |
| **CRP middle tercile**  Adjusted OR - mean (SD) | 1.32 (0.29) | 1.38 (0.36) | 0.99 (0.27) | 1 |
| - median (95% CrI) | 1.29 (0.85 to 1.95) | 1.34 (0.81 to 2.20) | 0.95 (0.57 to 1.62) | 1 |
| Probability of superiority to control, % | 87.6 | 87.1 | 43.3 | - |
| **CRP highest tercile**  Adjusted OR - mean (SD) | 1.80 (0.38) | 1.86 (0.47) | 1.23 (0.35) | 1 |
| - median (95% CrI) | 1.76 (1.18 to 1.95) | 1.80 (1.11 to 2.95) | 1.18 (0.68 to 2.06) | 1 |
| Probability of superiority to control, % | 99.7 | 99.1 | 72.8 | - |

subgroup analyses are performed by baseline C-reactive protein (CRP) subgroup. The CRP subgroups are defined based on terciles of CRP. Patients with CRP lower than 85 𝜇g/ml are included in the lowest CRP subgroup. Patients with CRP greater/equal to 85 and less than 169 𝜇g/ml are included in the middle CRP subgroup. Patients with CRP greater or equal to 169 𝜇g/ml are included in the highest CRP

subgroup. Patients with unknown CRP are included in a fourth “unknown” subgroup.

#### Table S7. Predefined subgroup analysis of the Primary Outcomes by baseline mechanical ventilation status

| **Outcome/Analysis** | **Tocilizumab (N=947)** | **Sarilumab (N=482)** | **Anakinra (N=373)** | **Control (N=405)** |
| --- | --- | --- | --- | --- |
| **Secondary Analysis of Primary Outcome**, model restricted to Immune Modulation Therapy Domain participants, according to **baseline mechanical ventilation status** | | | | |
| **No invasive mechanical ventilation at baseline**  Adjusted OR - mean (SD) | 1.62 (0.20) | 1.56 (0.23) | 1.11 (0.19) | 1 |
| - median (95% CrI) | 1.61 (1.26 to 2.07) | 1.54 (1.16 to 2.08) | 1.09 (0.78 to 1.53) | 1 |
| Probability of superiority to control, % | >99.9 | 99.9 | 69.5 | - |
| **Invasive mechanical ventilation at baseline**  Adjusted OR - mean (SD) | 1.22 (0.19) | 1.40 (0.28) | 0.89 (0.19) | 1 |
| - median (95% CrI) | 1.20 (0.81 to 1.64) | 1.38 (0.94 to 2.02) | 0.87 (0.57 to 1.31) | 1 |
| Probability of superiority to control, % | 87.6 | 94.8 | 25.4 | - |
| **Secondary Analysis of Hospital Survival,** model restricted to Immune Modulation Therapy Domain participants, according to **baseline mechanical ventilation status** | | | | |
| **No invasive mechanical ventilation at baseline**  Adjusted OR - mean (SD) | 1.56 (0.25) | 1.51 (0.29) | 1.13 (0.25) | 1 |
| - median (95% CrI) | 1.54 (1.12 to 2.11) | 1.48 (1.02 to 2.16) | 1.10 (0.72 to 1.70) | 1 |
| Probability of superiority to control, % | 99.6 | 98.0 | 67.8 | - |
| **Invasive mechanical ventilation at baseline**  Adjusted OR - mean (SD) | 1.18 (0.23) | 1.57 (0.39) | 0.84 (0.22) | 1 |
| - median (95% CrI) | 1.16 (0.79 to 1.70) | 1.53 (0.95 to 2.45) | 0.81 (0.50 to 1.34) | 1 |
| Probability of superiority to control, % | 76.8 | 95.9 | 20.8 | - |

#### Figure S1. Alterations to the Immune Modulation Therapy domain of REMAP-CAP

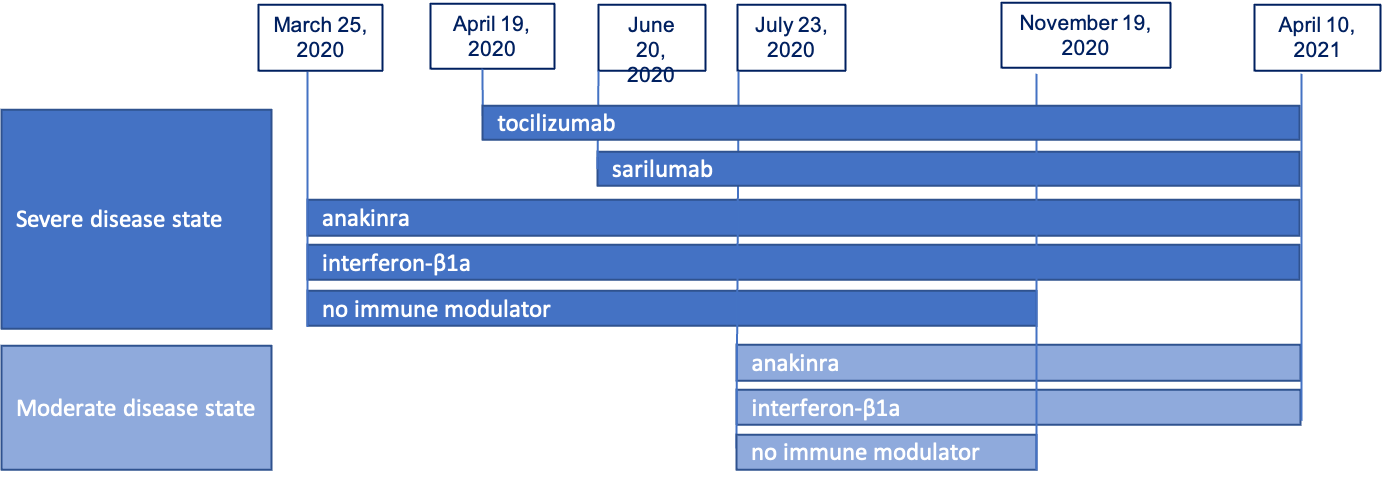

#### Figure S2. Forest Plot of Organ Support Free days for sarilumab compared to tocilizumab

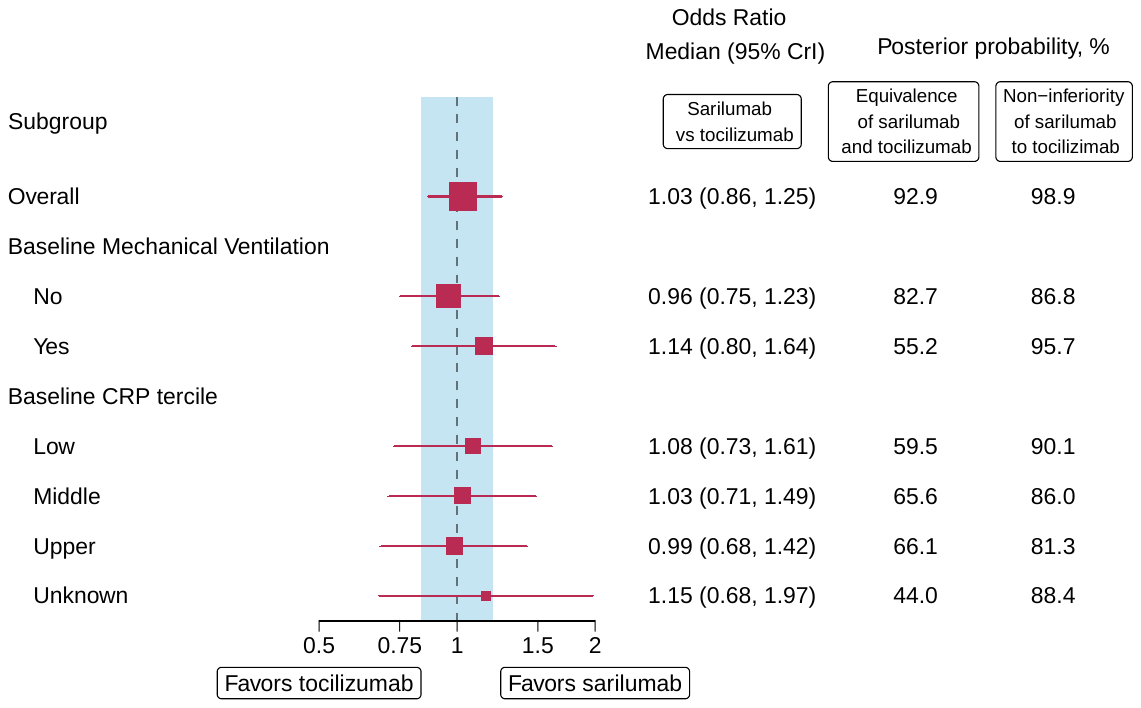

#### Figure S3. Forest Plot of Hospital Survival for sarilumab compared to tocilizumab

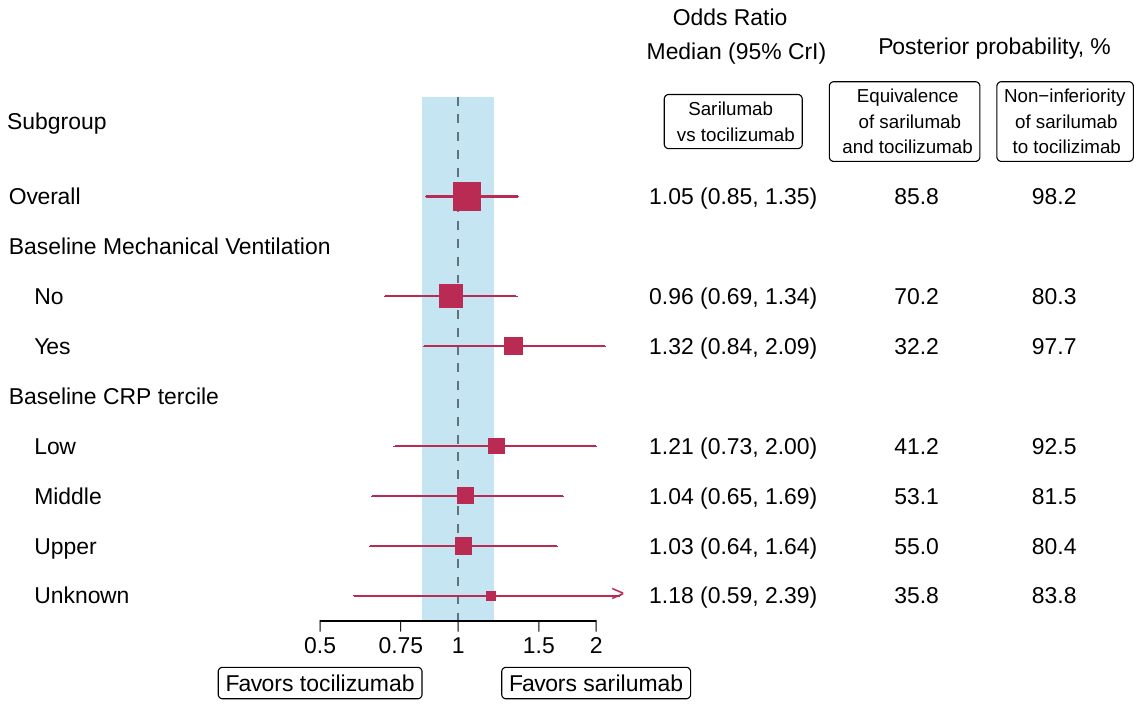

#### Figure S4. Organ Support Free Days - odds ratios and 95% CrI for tocilizumab, sarilumab, and pooled IL-6ra in models with nested, independent, and pooled treatment effects

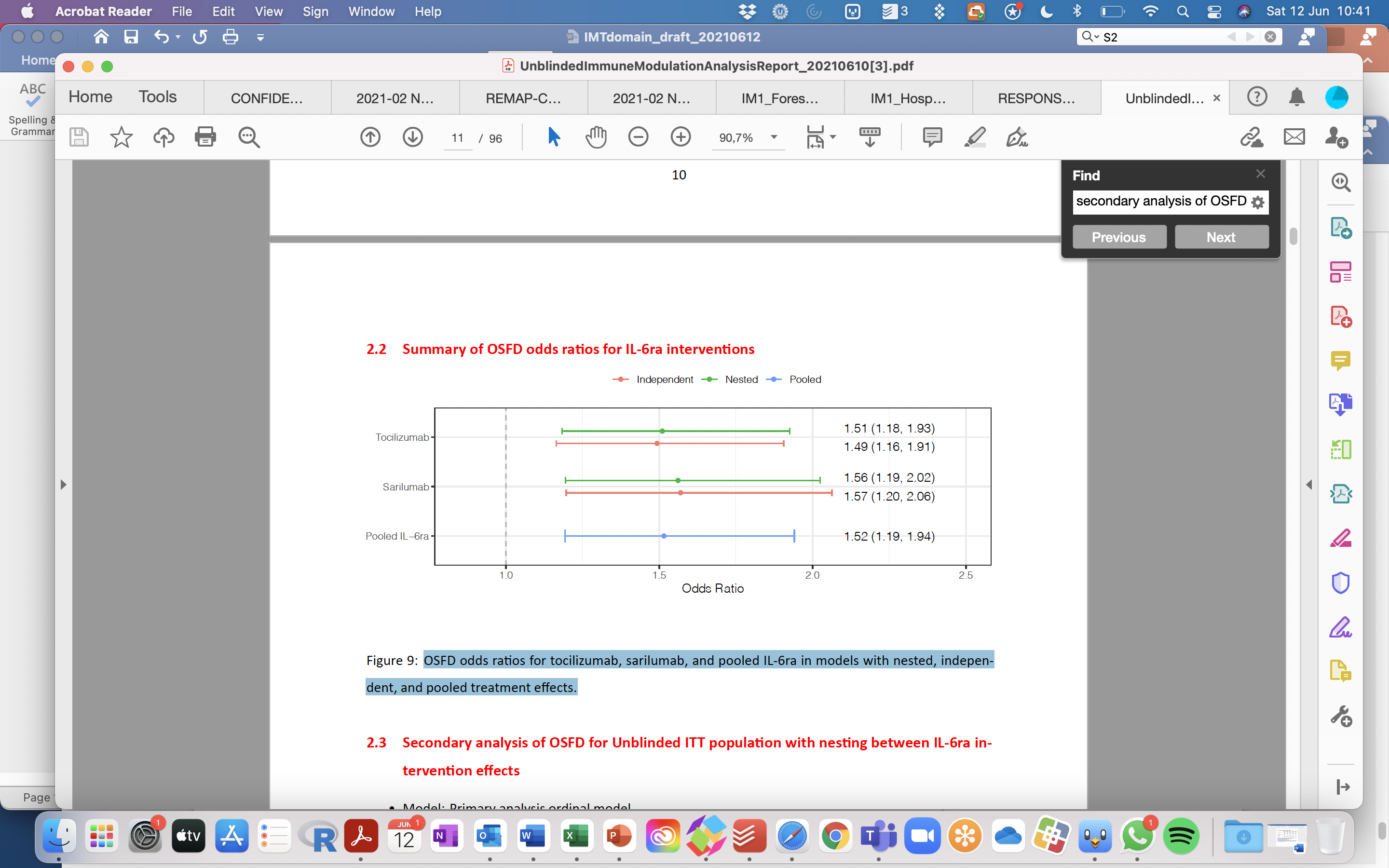

#### Figure S5. Forest Plot of Organ Support Free Days by subgroup

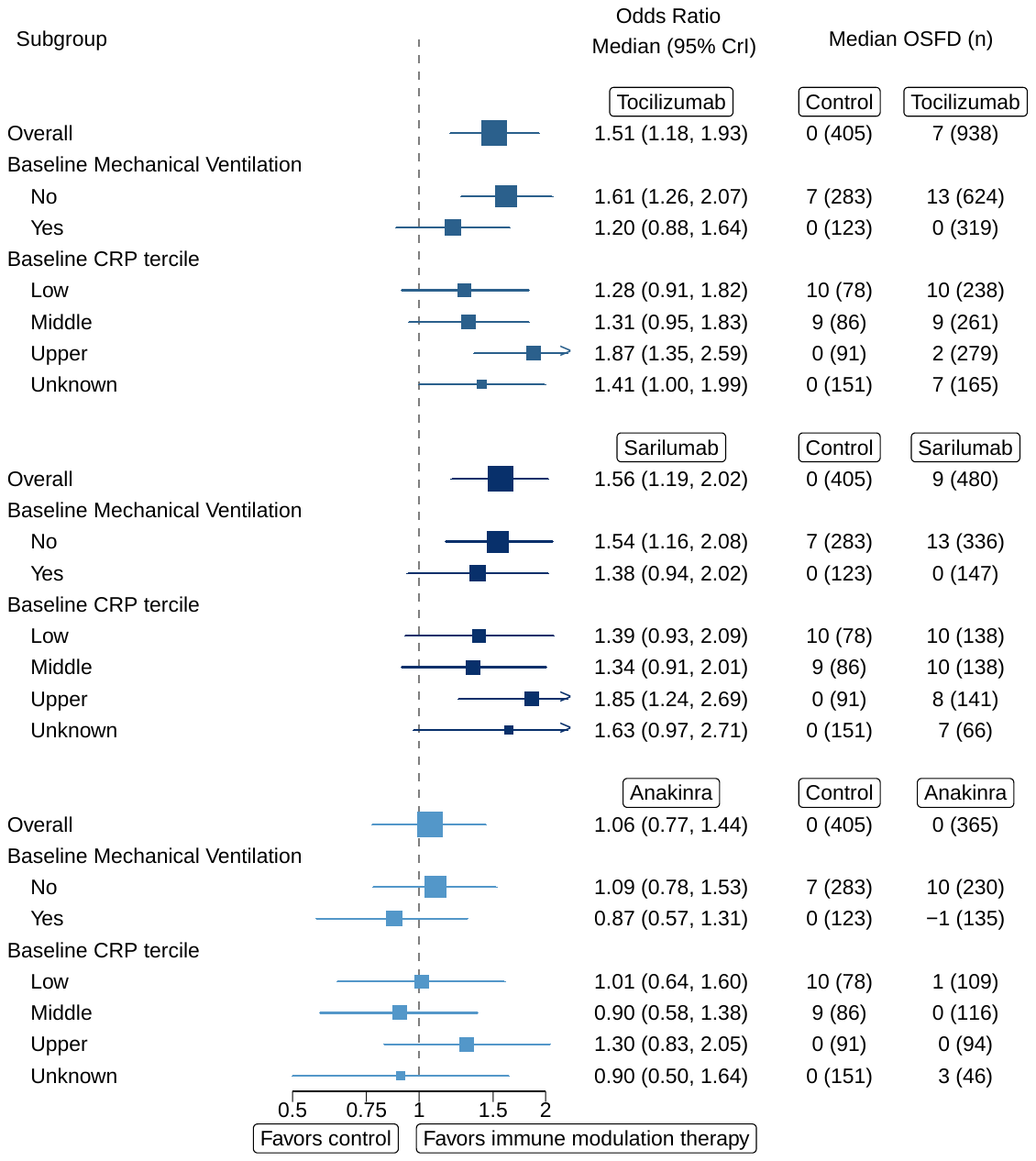

#### Figure S6. Forest Plot of Hospital Survival by subgroup

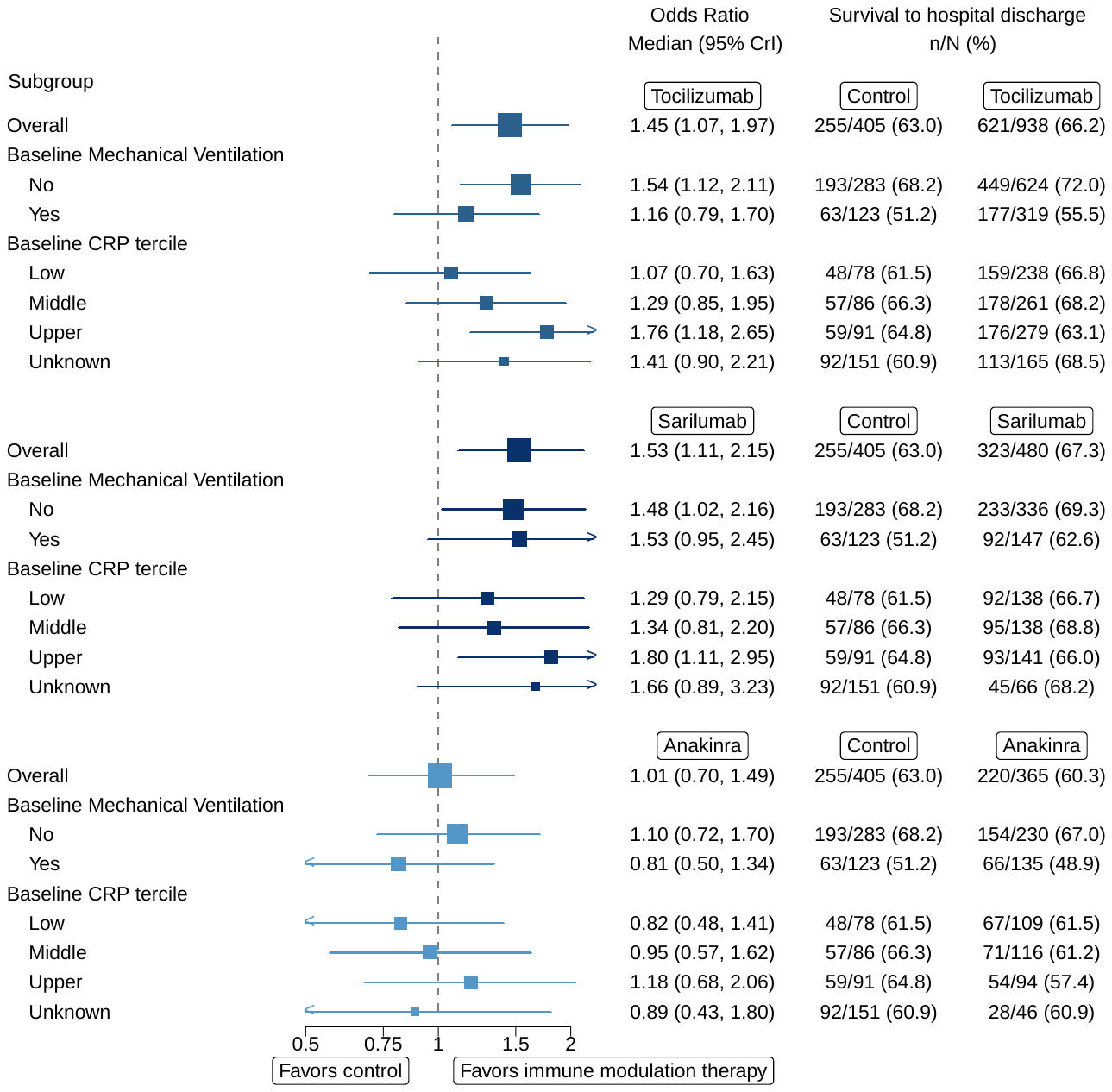

#### Appendix 1: Statistical Analysis Plan

#### Appendix 2: Statistical Analysis Committee Primary Analysis Report for the Immune Modulation Therapy Domain

#### Appendix 3: ITSC Secondary Analysis Report for the Immune Modulation Therapy Domain
