## Appendix 1 Statistical Analysis Plan for "Effectiveness of Tocilizumab, Sarilumab, and Anakinra for critically ill patients with COVID-19 The REMAP-CAP COVID-19 Immune Modulation Therapy Domain Randomized Clinical Trial"

REMAP-CAP
Randomized, Embedded, Multifactorial Adaptive Platform trial for Community-Acquired Pneumonia

### Statistical Analysis Plan for the Immune Modulation Therapy Domain for Patients with COVID-19 Pandemic Infection Suspected Or Proven (PISOP)

COVID-19 Immune Modulation Therapy Domain SAP Version 1.1 dated 15 April 2021

### 1. COVID-19 IMMUNE MODULATION THERAPY DOMAIN SAP VERSION

The version is in this document's header and on the cover page.

#### 1.1. Version history

Version 1: Finalized on November 25, 2020.

Version 1.1: Finalised on April 15, 2021

### 2. SAP AUTHORS

Scott Berry, Berry Consultants, Austin, TX, USA

Lindsay Berry, Berry Consultants, Austin, TX, USA

Elizabeth Lorenzi, Berry Consultants, Austin, TX, USA

Steven Webb, Monash University, Melbourne, Victoria, Australia

Derek C. Angus, University of Pittsburgh and UPMC Health System, PA, USA

Lennie Derde, UMC Utrecht, Netherlands

Paul Mouncey, ICNARC, London UK

Anthony Gordon, Imperial College, London, UK

### 3. INTRODUCTION

This statistical plan for the analysis of the Immune Modulation Therapy Domain in the pandemic stratum of the REMAP-CAP trial is an appendix to the Pandemic Appendix to Core (PAtC) Statistical Analysis Plan (SAP). This plan details the statistical analyses in the original REMAP-CAP core SAP and the pandemic stratum SAP applied to the analysis of the tocilizumab interventions in the Immune Modulation Therapy Domain. This plan is prespecified for the imminent unblinding of the data for the tocilizumab and control interventions in the Immune Modulation Therapy Domain within the pandemic infection suspected or proven (PISOP) (COVID-19) stratum.

The standard of care (control) intervention in the Immune Modulation Therapy Domain was halted in the PISOP stratum following the disclosure from the DSMB that the tocilizumab intervention, an IL-6 receptor antagonist, had met the pre-specified threshold for efficacy.

The REMAP-CAP ITSC publicly disclosed the efficacy of tocilizumab on November 19, 2020 (Greenwich Mean Time (GMT)) coinciding with the closure of the control intervention in the Immune Modulation Therapy Domain within the PISOP stratum of REMAP-CAP. REMAP-CAP explores multiple treatment domains by randomizing patients within multiple domains simultaneously. The adaptive platform trial was designed to produce modular results for individual interventions or full domains upon reaching platform conclusions. The predefined statistical trigger for efficacy for tocilizumab was met, and hence the results for the tocilizumab and immune modulation control interventions will be unblinded and made public. This document prespecifies the analysis plan for this unblinding.

The authors of this document are blinded to all individual data other than publicly disclosed results and that the statistical trigger for efficacy has been reached for tocilizumab. Given the recent increased enrollment to the Immune Modulation Therapy Domain, the primary analysis for this SAP will be conducted when the patient last randomized, before closing of the immune modulation control arm, reaches 21 days of follow-up (completion of the primary end-point).

When the full analysis model is run, once all patients have completed the primary follow-up period, if the other IL-6 receptor antagonist (sarilumab) has met a pre-specified threshold for superiority, efficacy, equivalence or futility, then it will be added to this analysis and reporting. In that case, both tocilizumab and sarilumab will be included in the analyses described below for tocilizumab. In addition, the analyses below will be run with the tocilizumab and sarilumab arms pooled and also the tocilizumab and sarilumab arms will be compared for equivalence.

#### 3.1 Additions for version 1.1

On April 8 2021, the DSMB informed the ITSC that tocilizumab and sarilumab had met the predefined triggers for equivalence, and that anakinra had met the predefined trigger for inferiority in the severe state. The decision to close the Interferon-beta-1a intervention for operational futility had also been approved. Therefore, further randomization to the domain was stopped on April 10 and unblinding and analysis of all existing patients and interventions is planned. At the time up updating this SAP, the authors are blinded to all individual data other than publicly disclosed statistical triggers that have been met.

This analysis will follow the previous version 1 of this SAP with the following additions / changes.

1. The primary analysis model now includes both moderate and severe state patients in the PISOP stratum. As the overwhelming majority of patients in this domain have been recruited in the severe state the main focus of reporting will be the severe state only. However, summaries/results for the moderate state may be reported where appropriate.
2. The unblinded ITT population will now include patients from the therapeutic anti-coagulation and immunoglobulin domains. Therefore, these additional interactions will be reported. As we know tocilizumab and sarilumab have met the equivalence trigger we will primarily report them as a pooled IL-6 RA group in these interaction analyses.
3. The previous tocilizumab / IL-6 RA specific populations will now be the Immune Modulation specific population and include all patients and all interventions in this domain within the severe state
4. As tocilizumab and sarilumab have met the trigger for equivalence it will be important to understand how similar the treatment effects of the two interventions are, and if there are any differences between them. The primary model run by the SAC includes these interventions as a nested analysis. For the secondary analyses, the models will be run without borrowing between the interventions to fully understand and explore similarities and differences.
5. We will also report the pooled IL-6 RA group result (including the posterior probability that the pooled group is optimal [domain superiority]), as well as the individual intervention effects (as above). We will also conduct the previous post-hoc subgroup analyses of mechanical ventilation vs no mechanical ventilation at baseline.
6. Analysis of anakinra will include comparisons to the pooled IL-6 RA group and also control. As described in the pandemic appendix to core, if an active intervention has a greater than

90% probability of being inferior to the control, then it will be deemed to be harmful in this population.

7. As the results of patients co-randomized in the therapeutic anti-coagulation and immunoglobulin domain have been unblinded then additional outcomes of bleeding and thrombotic events will be available. In those patients with these results, we will analyze major bleeding and major thrombotic events by immune modulation treatment, using a pooled IL-6RA group.
8. As previously decided, future interaction analyses with the corticosteroid domain will only include fixed-dose steroids as the active treatment group (shock-based steroids will be excluded).

### 4. DESIGN CONSIDERATIONS

REMAP-CAP is designed with a Bayesian analysis as the primary analysis method for the trial. There is one overarching Bayesian model, prespecified in the SAP, driving all adaptations, statistical triggers, and result summaries. That primary statistical analysis model will be used to report the results for the tocilizumab intervention in the Immune Modulation Therapy Domain within the severe state of the PISOP stratum.

The decision to use a Bayesian analysis was driven in part by the uncertainty of the extent of the pandemic. The sample size could be small or large, and there may be unexpected external events, such as other trial results, that alter the design of REMAP-CAP. Given the expected evolution of the design and uncertain sample size, the Bayesian approach is more appropriate.

REMAP-CAP defines several statistical triggers within the trial that, at any analysis of the trial, would result in public disclosure and a declaration of a platform conclusion.

The following internal statistical triggers were defined for the Immune Modulation Therapy Domain:

1. **Domain Superiority**. If a single intervention within the Immune Modulation Therapy Domain has at least a 99% posterior probability of being in the best regimen for patients in the severe state of the PISOP stratum, this would trigger domain superiority of that intervention.

2. **Intervention Efficacy**. If an intervention is deemed to have at least a 99% posterior probability of being superior to the control, then a declaration of efficacy of that intervention would be declared. This statistical trigger is active for each of the non-control arms in the Immune Modulation Therapy Domain.

3. **Intervention Equivalence**. If two non-control interventions have a 90% probability of equivalence, this would trigger a public disclosure of intervention equivalence.
4. **Intervention Futility**. If an intervention is deemed to have a less than 5% probability of at least a 20% odds ratio improvement compared to the control, then a declaration of futility of that intervention would be declared.

The 99% threshold for efficacy was selected to have good properties for potential outbreak sample sizes. For example, the type I error rate of any conclusion of efficacy for a single intervention 'A' vs. control is less than 2.5% for approximately less than 1000 patients on intervention 'A' with multiple interim analyses (see main and pandemic SAP).

### 5. UNBLINDING

REMAP-CAP has multiple domains to which patients can be randomized and multiple interventions within domains. At the unblinding of the tocilizumab/control interventions, there are other interventions to which patients have been randomized that will not be unblinded at this analysis unless a statistical trigger is hit at the time of the primary analysis. This includes interferon-beta 1a, anakinra and sarilumab interventions within the Immune Modulation Therapy Domain and interventions within other domains. In the analysis plan, there will be analyses conducted by the Statistical Analysis Committee (SAC) using additional randomizations and unblinding of other randomizations. The SAC is unblinded to all interventions and domains as part of their role for REMAP-CAP. There will be other analyses that are conducted with only knowledge of the tocilizumab/control allocation status for patients or the allocation status to other unblinded interventions. These may be conducted by investigators who are blinded to information about other interventions and domains. These analyses are identified below.

### 6. INTERVENTIONS

There are five interventions within the Immune Modulation Therapy Domain. These are

1. No immune modulation for COVID-19 (control)
2. interferon-beta-1a (IFN-β1a)
3. anakinra (interleukin-1 receptor antagonist; IL1Ra)
4. tocilizumab (IL-6 receptor antagonist; IL6Ra)
5. sarilumab (IL-6 receptor antagonist; IL6Ra)

For the primary analysis completed by the SAC, all five arms will be modeled, but only results for tocilizumab relative to control will be reported. For all secondary analyses completed by blinded investigators, tocilizumab will be compared to the control.

The models in this SAP will estimate the interaction effects of the interventions in the Immune Modulation Therapy Domain with the Corticosteroid and Antiviral Domain interventions. The pre-specified interaction effects of tocilizumab and other unblinded interventions will be reported. This includes the interactions between tocilizumab and Corticosteroid Domain interventions (fixed-dose and shock-based hydrocortisone) and tocilizumab and Antiviral Domain interventions (hydroxychloroquine (HCQ), Kaletra, and Kaletra + HCQ).

### 7. DISEASE STATES

There are two disease states in the PAtC, which are **moderate** and **severe**. Tocilizumab has only been open for randomization in patients in the severe state, so only patients in the severe state will be analyzed.

### 8. ANALYSIS POPULATIONS

1. REMAP-CAP COVID-19 severe state intent-to-treat (ITT). This population consists of all PISOP patients in the severe state randomized within at least one domain.
2. Unblinded ITT. All patients randomized to tocilizumab or no immune modulation interventions in the Immune Modulation Therapy Domain, the Corticosteroid Domain, or the Anti-Viral Domain within the PISOP stratum. Note the assignment to the interventions mentioned above will be unblinded, the other intervention assignments will not be unblinded to the analysis team.
3. Unblinded non-negative COVID-19. All patients in the Unblinded ITT population after removing those with ≥1 negative test for COVID-19 **and** no positive tests.
4. Tocilizumab specific ITT. This population consists of only patients eligible to be randomized to tocilizumab that were randomized to tocilizumab or the no immune modulation intervention in the Immune Modulation Therapy Domain within the PISOP stratum
5. Tocilizumab specific per protocol. This consists of the patients in the Tocilizumab ITT population who have been treated as per protocol. In this analysis that is defined as patients randomized to tocilizumab, **and** received at least one dose, or randomized to no immune modulation **and** did not receive any interventions in the Immune Modulation Therapy Domain.

REMAP-CAP is continuing to enroll patients to some of the interventions/domains being analyzed in this SAP. Each of these analysis populations will include only the patients randomized on or before the no immune modulation (control) arm was stopped on 19 November 2020.

### 9. ENDPOINTS

The following end points will be analyzed, displayed graphically, and summarized through descriptive statistics.

1. **Organ Support-Free Days** (OSFD)
    a. An ordinal endpoint with mortality as the worst outcome. The primary endpoint for the REMAP-CAP PISOP stratum. The organ support considered is cardiovascular (vasopressor/inotrope support) and respiratory support. See Appendix A for a detailed description.

2. **In-Hospital Mortality**
    a. A dichotomous endpoint of in-hospital death where the death component corresponds to a –1 on the OSFD endpoint.

3. **Mortality**
    a. This is a time-to-event endpoint through 90-days.
    b. Any patient currently in the hospital or transferred on organ support to an alternative care facility will be censored at their last known status alive.
    c. Any patient successfully discharged from hospital, alive, without organ support, will be censored at the date of discharge, if 90-day mortality data are not yet recorded.

4. **Progression to intubation and mechanical ventilation, extracorporeal membrane oxygenation (ECMO), or death**
    a. A dichotomous endpoint of whether a patient progresses to intubation and mechanical ventilation, ECMO or death in hospital.

5. **Vasopressor/Inotrope Free-Days**
    a. An ordinal outcome of the number of days free of Vasopressor/Inotropes. This is the exact calculation of OSFD, with Vasopressor/Inotropes as the only organ support category. In-hospital death is considered a –1.

6. **Ventilator Free-Days**

a. An ordinal outcome of the number of days free of ventilation. This is the exact calculation of OSFD, with ventilation as the only organ support category. In-hospital death is considered a –1.

7. **Duration of ICU stay**
    a. A time-to-event endpoint of leaving the ICU alive. If a patient is known to leave the ICU and return to the ICU within 14-days that intervening time will be ignored.
    b. This variable will be truncated at 90-days: all deaths in ICU will be considered 90-days with no liberation of ICU.
    c. Patients still in the ICU at data snapshot will be considered censored.

8. **Duration of hospital stay**
    a. A time-to-event endpoint of leaving the hospital alive. If a patient is known to leave and return to the hospital within 14-days that intervening time will be ignored.
    b. This variable will be truncated at 90-days and all deaths in-hospital will be considered 90-days with no events.
    c. Patients still in the hospital at data snapshot will be considered censored.

9. **At least one serious adverse event (SAE)**
    a. A dichotomous endpoint of SAE.
    b. This endpoint will be summarized descriptively. Counts and proportions of SAEs will be provided by intervention.

10. **The World Health Organization (WHO) 8-point ordinal scale, measured at day 14.**
    a. A modified WHO ordinal scale will be used:

    0 + 1 + 2 = No longer hospitalized

    3 = Hospitalized, no oxygen therapy

    4 = Oxygen by mask or nasal prongs

    5 = Non-invasive ventilation or high-flow oxygen

    6 = Intubation and mechanical ventilation

    7 = Ventilation + additional organ support: vasopressors, renal replacement therapy (RRT), ECMO

    8 = Death

### 10. GRAPHICAL DATA SUMMARIES

1. All ordinal endpoints will be plotted using stacked cumulative bar plots and cumulative probability plots.

2. All time-to-event endpoints will be plotted using Kaplan-Meier plots. Positive clinical event outcomes will be plotted as the cumulative rate of event, and negative events will be plotted as the cumulative rate of event-free.

### 11. DESCRIPTIVE STATISTICS

1. Ordinal endpoints will be summarized by the cumulative frequency of each outcome. The 25th, 50th, and 75th percentiles will be summarized.
2. Dichotomous endpoints will be summarized by the proportion in each category.
3. Time-to-event outcomes will be summarized by the 2.5th, 10th, 25th, 50th, 75th, 90th, and 97.5th percentiles from the Kaplan-Meier estimates, as available.

### 12. BASELINE CHARACTERISTICS

The following demographics will be summarized across arms. More may be added as baseline summaries: Age, sex, BMI, ethnicity, APACHE II score (measured from hospital admission to randomization), confirmed SARS CoV-2 infection, preexisting conditions, baseline use of high-frequency nasal oxygenation, non-invasive ventilation, invasive mechanical ventilation, ECMO, vasopressors/inotropes, renal replacement therapy, and miscellaneous physiological values and inflammatory biomarker laboratory values.

### 13. COMPLIANCE

The compliance to immune modulation use will be summarized descriptively as the fraction of use, for each randomized arm.

### 14. ANALYTIC APPROACH

Each inferential analysis will be done using a Bayesian model. Some default frequentist methods are used for exploration and description. A summary of the analyses methods is provided below.

#### 14.1. Primary Analysis of Primary Endpoint

The **primary analysis model** is a Bayesian cumulative logistic model for the ordinal primary endpoint. The model is described below.

The primary endpoint for the severe state has 23 possible, ordered outcomes. Let the outcome for a patient by labeled as $Y_i$, with possible values, –1 (death), 0, 1, …, 21, 22. The outcome of 22 for the severe state (never received organ support) is not possible. Hence there are 23 possible outcomes. A cumulative logistic model is specified. The model is structured so that an odds-ratio >1 implies clinical benefit. The full details of the model are specified in the Current State of The Statistical Model, Version 2.2 dated October 1, 2020. The model has factors for:

- Each level of the ordinal endpoint
- Each global site, nested within country
- Age; ≤39, 40-49, 50-59, 60-69, 70-79, 80+
- Sex
- Time; 2-week buckets of time working backwards from the last enrolled patient, with the most recent bucket being 4 weeks.
- For each domain, an effect for being randomized to the domain
- For each domain, an effect for being eligible for the domain
- An effect for each intervention within each domain
- Specified interactions in the model between interventions across domains

The primary analysis for tocilizumab uses the following rules:

- The high-dose 7-day hydrocortisone arm will be combined with the 7-day hydrocortisone arm (fixed-duration). They were originally nested, which allows their pooling, and there were very few patients randomized to the high-dose 7-day hydrocortisone arm.
- For patients who were randomized as part of REMAP-CAP COVID-19 severe state ITT after the closure of corticosteroid domain (June 17, 2020), the subjects are coded as receiving fixed-dose hydrocortisone.
- All sites within a country that have <5 patients randomized will be combined into a single site within that country.
- If there is an outcome in the ordinal scale that did not occur in the data, then that outcome will be combined with a neighboring outcome (the worse outcome). This is done for model stability. For example, if the outcome 11 never occurred, then a combined outcome of 10 & 11 will be modeled for the analysis.

The primary analysis model will be referenced with certain model assumptions for sensitivity analyses. For example, the “time effects” in the model could be assumed to be 0.

##### 14.1.1. Proportional Odds Assumption

The primary analysis model is based on an assumption of a proportional effect of treatment across the scale of the ordinal outcome. In order to assess the robustness of the results to this assumption, a dichotomous model is fit to every level of the ordinal outcome across the scale and the odds-ratio for each dichotomous break is presented. No statistical test of proportional odds is conducted.

#### 14.2. Analytic Approach for Secondary Dichotomous Endpoints

A Bayesian logistic regression model will be used for each dichotomous outcome. The model will always specify the "event" as the negative outcome and be parameterized so that an odds-ratio >1 implies benefit to patients. The model is the standard logistic link function model:

$$log\left(\frac{\pi}{1-\pi}\right) = \alpha - [factors]$$

References will be made to the factors in the model and their prior distribution. Many of these factors will be the same as the primary analysis model, with the same priors, as the parameters have similar interpretation. For example, all in-hospital mortality models should use the Beta prior distribution implied by the Dirichlet prior in the OSFD model. If not otherwise specified, the prior distribution for the main effect is $\alpha \sim N(0, 1.82^2)$ (similar to a uniform prior on the probability scale).

#### 14.3. Analytic Approach for Secondary Time-To-Event Endpoints

All inferential time-to-event analyses will be done using a Bayesian piecewise exponential model. The Bayesian time-to-event model is intended to mirror a Cox proportional hazards model, with the underlying hazard rate modeled with a piecewise exponential model. The underlying hazard will be modeled with a hazard rate for each 10-day period in the model. The prior distribution for the hazard rate for each day is a gamma distribution with 1 day of exposure and a mean equal to the total exposure divided by the total number of events. This prior will have very little weight but will provide numerical stability to the model. Each factor is incorporated as a proportional hazard rate through an additive linear model of the log-hazard. The default prior for each factor is the same as for the log-odds in the ordinal model. If other non-specified variables are added to the model, then a normal distribution with mean 0 and standard deviation 10 will be utilized.

#### 14.4. Markov Chain Monte Carlo (MCMC) Model Stability

The Bayesian models have many parameters and there may be risk of poor model stability, including convergence and mixing behavior of the MCMC sampler. These instabilities may be based on sparse data on the outcome or covariates. The statisticians running the model may make changes that do not affect the overall interpretation but provide reliable model diagnostics and scientific rigor. Any alterations will be noted.

#### 14.5. Model Outputs

The standard model outputs for each treatment effect will be the mean, standard deviation, median, and 95% credible intervals (all credible intervals will be equal-tailed intervals, so 95% credible intervals will range from the $2.5^{th}$ percentile to the $97.5^{th}$ percentile of the posterior distribution). For the ordinal endpoints, the odds-ratios will be summarized. For the dichotomous endpoints, the odds-ratio will be summarized. For the time-to-event endpoints, the hazard ratios will be summarized. For consistency, all models will be parameterized so that an odds-ratio or hazard-ratio greater than 1 indicates clinical benefit.

For each inferential model, a posterior probability that one arm is superior will be provided for each comparison between arms. This posterior probability has been identified as the primary analysis metric between arms. A posterior probability greater than 99% of superiority has been identified as statistically significant in REMAP-CAP.

#### 14.6. Exploratory Analyses

Exploratory analyses after unblinding will not be considered inferential and no p-values will be presented. Any post-hoc exploratory analyses will use the following methods:

1. Ordinal endpoints will be compared using a cumulative proportional odds model with summaries of the odds-ratio, 95% confidence intervals, and Wilcoxon tests for robustness against a lack of proportional odds.
2. Time-to-Event analyses will utilize a Cox proportional hazards model, summarizing the hazard ratios and 95% confidence intervals.
3. Continuous endpoints will compare means with 95% confidence intervals based on two-sample t-test procedures.
4. Dichotomous proportions will be compared using logistic regression summarizing the odds-ratio and 95% confidence intervals. Differences between proportions will be summarized using observed differences and normal approximations for the 95% credible intervals.

### 15. SPECIFIC PROSPECTIVE ANALYSES

The specific analyses to be undertaken will be as listed in version 1.0 but with the additions / changes described in section 3.1.

### Appendix A. Definition of organ support-free days

This outcome is an ordinal scale of integers from –1 to 22 for each state (Moderate or Severe) derived from a composite of the patient's vital status at the end of acute hospital admission and days spent receiving organ failure support while admitted to an ICU (including a repurposed ICU) during the 21 days (504 hours) after randomisation.

A patient enrolled in the Severe State while still in an Emergency Department is regarded as 'admitted to an ICU' and the time of commencement of organ failure support is the time of randomisation, as it is for all other patients in the Severe State.

Patents who survive to hospital discharge and are enrolled in one or more domains in the Moderate State and are enrolled in one or more domains in the Severe State have a primary end point value for each state, which may be different.

If deceased between first enrolment and ultimate hospital discharge, code OutcomeDay21 as -1

If not deceased, ModerateOutcomeDay21 = 21 – (the sum of the length of time in days and part-days between time of first commencement of organ failure support while admitted to an ICU and the time of last cessation of organ failure support during that ICU admission plus time between first commencement and last cessation of organ failure support during any and all subsequent readmissions to ICU, censored at the 504 hours after enrolment in the Moderate State)

- A patient who is enrolled in the Moderate State who never receives organ failure support while admitted to an ICU has an ModerateOutcomeDay21 = 22.
- A patient who is enrolled in the Moderate State in a ward location who commences organ failure support on the ward and is transferred to an ICU while receiving organ failure support has a commencement time of organ failure support corresponding to the time of ICU admission.

If not deceased, SevereOutcomeDay21 = 21 – (the sum of the length of time in days and part- days between time of enrolment and the time of last cessation of organ failure support during that ICU admission plus the lengths of time between first commencement and last cessation of organ failure support during any and all subsequent readmissions to ICU, censored at 504 hours after the time of enrolment

Decimals are rounded up or down to nearest whole day.

If transferred between hospitals before the last study day 21 and known to be alive at ultimate hospital discharge use all available information to calculate Outcome Day21 with an assumption that no subsequent organ failure support in an ICU was provided.

If transferred between hospitals before the last study day 21 and vital status at ultimate hospital discharge is not known, code as follows:

- If last known to be on a ward use all available information to calculate OutcomeDay21 with an assumption that the patient has not died prior to ultimate hospital discharge and that there were no subsequent ICU admissions.
- If last known to be in an ICU, code OutcomeDay21 as missing (999)

If a patient is discharged alive from the ultimate hospital before 504 hours from each enrolment, assume all subsequent time is alive and without provision of organ failure support in an ICU.

If the patient is alive at the end of one or both censoring time points, the hours will be calculated as above. If the patient dies after the end of one or both of the censoring time points and before hospital discharge, the value will be updated to -1

A patient who remains admitted to an acute hospital and is still alive at the end of study day 90 no further changes to coding will be made.
