## Appendix 2 Primary analysis for "Effectiveness of Tocilizumab, Sarilumab, and Anakinra for critically ill patients with COVID-19 The REMAP-CAP COVID-19 Immune Modulation Therapy Domain Randomized Clinical Trial"

### REMAP-CAP (REMAP-COVID)

#### Analysis of COVID-19 Immune Modulation Domain

May 30, 2021

---

**4 Other Data Summaries** **23**

**5 Analysis Conventions** **30**

**6 Model Stability** **30**

**7 Report Production** **31**

### 1 Introduction

#### 1.1 Overview of the Adaptive Design

This trial is a Randomized, Embedded, Multifactorial Adaptive Platform (REMAP) trial that was originally designed to investigate treatments for Community-Acquired Pneumonia (CAP). The platform trial has the ability to investigate multiple interventions within multiple domains, across different patient strata. The number of interventions, domains, and strata may increase or decrease as the trial progresses. The platform trial includes a pandemic stratum that was activated when COVID-19 emerged. The pandemic stratum-specific protocol details are provided in a Pandemic Appendix to the Core (PAtC) protocol. The PAtC investigates therapies for patients with pandemic infection that are classified as suspected or proven (PISOP). This report focuses on the COVID-19 PISOP stratum.

For the PISOP stratum, patients may be randomized to interventions while they are in a Severe disease state or a Moderate disease state. State definitions are in the PAtC. Patients initially randomized in a Moderate state may progress in their disease severity, and subsequently meet the criteria for Severe state, and have additional randomization and reveal of interventions for Severe state domains.

#### 1.2 Purpose of this Report

This report contains the final analysis of the COVID-19 Immune Modulation domain for both Moderate and Severe disease states.

The international trial steering committee (ITSC) halted randomization to the COVID-19 Immune Modulation domain in the PISOP stratum Severe and Moderate states on April 10, 2021 following the disclosure from the Data and Safety Monitoring Board (DSMB) that the pre-specified threshold for futility had been met for anakinra in Severe state and that the pre-specified threshold for equivalence had been met for tocilizumab and sarilumab. Previously, randomization to the standard of care (control) intervention in the domain had been halted after tocilizumab met the pre-specified threshold for efficacy. Subsequently, the sarilumab intervention also met the pre-specified threshold for efficacy. The results for tocilizumab and sarilumab were publicly disclosed. Randomization within the domain continued for all interventions other than the control until April 10.

Although the ITSC will be unblinded to the interventions within the COVID-19 Immune Modulation domain, they will not be unblinded to the other domains to which patients have been randomized (except the corticosteroid domain, the COVID-19 antiviral domain, the therapeutic anticoagulation domain, and the Immunoglobulin domain, which were previously unblinded). The fully unblinded Statistical Analysis Committee (SAC) performed the set of analyses that use the full statistical model including data from all domains in the PISOP stratum. This report summarizes the data and the results for the Immune Modulation domain interventions resulting from the analyses using the full statistical model. This report is restricted to summaries and results pertaining to the unblinded interventions. Summaries and results for other ongoing domains are contained in a separate unblinded report only viewed by the SAC and DSMB.

#### 1.3 Endpoints

##### 1.3.1 Primary Endpoint: Organ-Support Free-Days (OSFD)

The primary endpoint is organ support-free days (OSFD) (days alive and free of ICU-based respiratory or cardiovascular support) within 21 days, where patients who die before discharge from the index hospitalization, and before day 90, were assigned a $-1$ day, even if the death occurred after day 21. The cumulative hours of organ support are computed and then rounded to the nearest day. Patients who receive no organ support in an ICU will be coded as 22 days. An outcome of 22 days is not possible for patients in Severe state. An outcome of 21 organ-support free-days is only possible in the Severe state if the patient had less than 12 hours of organ support.

##### 1.3.2 Secondary Endpoint: In-Hospital Mortality

The secondary endpoint is a dichotomous endpoint of in-hospital mortality, i.e. those patients with a $-1$ for the OSFD endpoint.

#### 1.4 Vocabulary

- **Domain:** a specific set of competing alternative interventions within a common clinical mode
- **Intervention:** is a treatment option that is subject to variation in clinical practice (comparative effectiveness intervention) or has been proposed for introduction into clinical practice (experimental intervention) and also is being subjected to experimental manipulation within the design of a REMAP.
- **Regimen:** Each patient is assigned a single intervention from each domain. The regimen is the combination of assigned interventions across the domains.
- **Immediate Reveal Domain:** is one for which all participants are eligible, the allocation status is made known, and the intervention is initiated at the time of randomization.
- **Delayed Reveal Domain:** is one for which all participants received a randomization assignment, but the allocation status is only made known and the intervention initiated if and when eligibility occurs. This occurs for example, when a domain is appropriate only for patients in a certain disease state and the patient transitions to that disease state.
- **Deferred Reveal Domain:** is one for which patients receive a randomization assignment and the allocation status is made known based on eligibility criterion known at the time of randomization, but additional information to assess that eligibility becomes known after some time. This occurs for example, when a test results confirming an eligibiltiy criterion are returned after some time.
- **Nest:** A grouping of interventions within a domain that are modeled hierarchically in order to allow for borrowing among the interventions effect estimates.
- **State:** Defined by the disease characteristics of the patient and may change over time as the disease progresses. States are used to define eligibility for certain domains.

#### 1.5 Current Trial Status

Figure 1.1 gives an overview of the interventions, domains, and strata currently being investigated in the COVID-19 pandemic portion of the trial. Each intervention is represented by a colored box, with similar colors used for interventions within the same domain. The figure also indicates features of the statistical model. For example, interactions are represented with an arrow and star (★). Within a domain, interventions that are nested within a hierarchical model are grouped within a curly bracket. Interventions that are closed to enrollment are indicated by an “X”. Table 1.1 is a companion to the current state figure, and provides the mapping of intervention codes to the actual intervention names (e.g. X2 = Lopinavir/ritonavir).

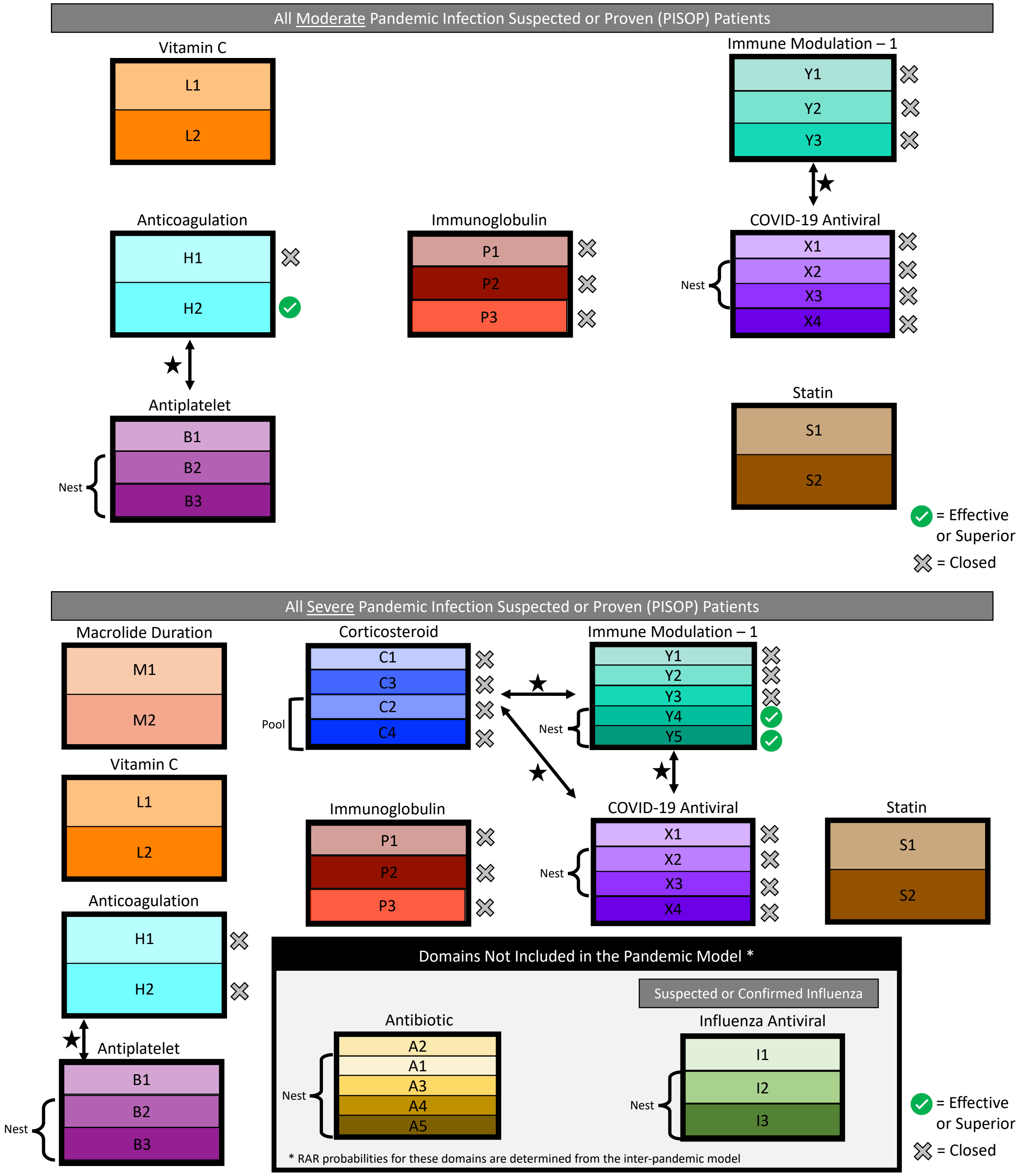

**Figure 1.1:** Current state of the **Moderate State** and **Severe State** pandemic domains and interventions. Each colored box represents an intervention, grouped by domain, with similar colors used for interventions within the same domain. Domains connected with an arrow and indicated with a star (★) will have interaction terms fit between the interventions in those domains. Within a domain, interventions that are grouped with a curly bracket are part of a nest whose main effects are estimated with a hierarchical model. Interventions that are closed to enrollment are indicated by a grey "X". Interventions that have met an Efficacy or Superiority trigger are indicated by a green checkmark. Closure of the Antiviral domain occurred simultaneously with the closure of the control arm (Y1) in the Immune Modulation domain and the superiority trigger of tocilizumab (Y4). The superiority trigger for sarilumab (Y5) occurred subsequently and results were publicly disclosed along with tocilizumab.

**Table 1.1:** List of all interventions to which a patient may be allocated, by disease state.

| Code | Intervention | Moderate | Severe |
|---|---|---|---|
| **Antibiotic** | | | |
| A1 | Ceftriaxone + Macrolide | Not Available | |
| A2 | Moxifloxacin or Levofloxacin | Not Available | |
| A3 | Piperacillin-Tazobactam + Macrolide | Not Available | |
| A4 | Ceftaroline + Macrolide | Not Available | |
| A5 | Amoxicillin-Clavulanate + Macrolide | Not Available | |
| **Macrolide Duration** | | | |
| M1 | Standard course (3 to 5 days) | Not Available | |
| M2 | Extended course (14 days) | Not Available | |
| **Corticosteroid** | | | |
| C1 | No corticosteroids | Not Available | Closed |
| C2 | Hydrocortisone (50mg) | Not Available | Closed |
| C3 | Shock dependent hydrocortisone | Not Available | Closed |
| C4 | High-dose hydrocortisone (100mg) | Not Available | Closed |
| **Influenza Antiviral** | | | |
| I1 | No antiviral | Not Available | |
| I2 | Oseltamivir 5 days | Not Available | |
| I3 | Oseltamivir 10 days | Not Available | |
| **COVID-19 Antiviral** | | | |
| X1 | No antiviral for COVID-19 | Closed | Closed |
| X2 | Lopinavir/ritonavir | Closed | Closed |
| X3 | Hydroxychloroquine | Closed | Closed |
| X4 | Hydroxychloroquine + lopinavir/ritonavir | Closed | Closed |
| **COVID-19 Immune Modulation** | | | |
| Y1 | No immune modulation for COVID-19 | Closed | Closed |
| Y2 | Interferon Beta-1a | Closed | Closed |
| Y3 | Anakinra | Closed | Closed |
| Y4 | Tocilizumab | Not Available | Effective |
| Y5 | Sarilumab | Not Available | Effective |
| **COVID-19 Immunoglobulin** | | | |
| P1 | No Immunoglobulin against COVID-19 | Closed | Closed |
| P2 | Convalescent plasma | Closed | Closed |
| P3 | Delayed convalescent plasma | Closed | Closed |
| **COVID-19 Therapeutic Anticoagulation** | | | |
| H1 | Standard practice thromboprophylaxis | Closed | Closed |
| H2 | Therapeutic anticoagulation | Effective | Closed |
| **Vitamin C** | | | |
| L1 | No vitamin C | | |
| L2 | Vitamin C | | |
| **Statin Therapy** | | | |
| S1 | No simvastatin | | |
| S2 | Simvastatin | | |
| **COVID-19 Antiplatelet** | | | |
| B1 | No antiplatelet therapy | | |
| B2 | Aspirin | | |
| B3 | P2Y12 inhibitor (clopidogrel, prasugrel, or ticagrelor) | | |

#### 1.6 Analysis Population

This report restricts the analysis population to consented patients with pandemic infection suspected or proven (PISOP) that were randomized, including both Moderate and Severe disease states, on or before April 10, 2021. This population is defined as the **REMAP-CAP COVID-19 severe and moderate state ITT population**. The patient population breakdown is as follows:

- 665 PISOP consented patients randomized initially in **Moderate** disease state (inclusive of 20 patients initially randomized while in Moderate disease state that later met the criteria for Severe disease state and had additional randomization for Severe state domains), on or before April 10, 2021;

  - ➔ 663 PISOP consented patients randomized for the first time in Moderate state for whom 21 days have elapsed since randomization and there is a known outcome on the 21-day organ-support free-days endpoint;

- 5292 PISOP consented patients randomized in **Severe** disease state (inclusive of 20 patients initially randomized while in Moderate disease state that later met the criteria for Severe disease state and had additional randomization for Severe state domains), on or before April 10, 2021;

  - ➔ 5189 PISOP consented patients randomized in Severe state for whom 21 days have elapsed since randomization and there is a known outcome on the 21-day organ-support free-days endpoint.

These counts exclude patients that withdrew consent for the use of their data. The patients who withdrew consent do not appear in the SAC data export, so no information is available to the SAC regarding when in the process (e.g. before or after eligibility assessment) consent was withdrawn.

Thus the analysis model is run on the 5852 outcomes for all PISOP patients (663 Moderate and 5189 Severe) for whom 21 days have elapsed since randomization and there is a known outcome on the 21-day OSFD endpoint. A patient randomized in both disease states is expected to have an outcome in each disease state (thus two outcomes in the model).

Although the full statistical model is run on the population defined above, the report only presents data summaries and results for the Immune Modulation domain.

### 2 Data Summaries

#### 2.1 Overview of Descriptive Data Summaries

Data for the PISOP patient population will be summarized, both across all patients, by disease state (without respect to intervention assignments), and at the intervention level for the interventions in the COVID-19 Immune Modulation domain. A description of each of the summary tables and figures is provided here.

**Summary of the availability of data:**

- **Number Eligible:** Eligibility is assessed both at the domain level and the intervention level. We tabulate the number of patients eligible for the domain, and within each category of domain eligibility, the number of patients eligible for each intervention. Eligibility captures both the patient meeting the inclusion criteria, and the domain or intervention being available and active at their site.

- **Number Assigned:** We tabulate the number of patients assigned to each intervention, by eligibility category. No randomized assignment can be given when a patient is ineligible for a domain, or when a patient is eligible for only one intervention within a domain. A patient must be eligible for at least two interventions within a domain to receive a randomized assignment.

- **Number Past 21 Days:** Among the patients eligible and assigned to each intervention, we tabulate the number of patients who have had the opportunity to complete the 21 days of follow-up for the primary endpoint. A patient must have been in the trial at least 21 days to be included in the analysis.

- **Number Missing:** Among the patients eligible and assigned to each intervention, we tabulate the number of patients who have completed 21 days of follow-up but do not have an outcome available on the primary endpoint.

- **Number Known:** Among the patients eligible and assigned to each intervention, we tabulate the number of patients who have completed 21 days of follow-up and have a known outcome on the primary endpoint.

- For the subjects that are eligible for the domain, a stacked bar chart summarizes the number and percent of patients assigned to each intervention.

**Summary of the observed outcomes data:**

For patients that are eligible for the domain and assigned to an intervention, we repeat the tabulation of the number of patients assigned to an intervention and with a known outcome on the 21-day endpoint. Additionally, we provide summaries of the following:

- **Number Deaths:** The number of in-hospital deaths, where the death corresponds to $-1$ on the OSFD endpoint.
- **Mortality Rate:** We calculate the observed in-hospital mortality rate as the number of in-hospital deaths out of the total number of patients with a known 21-day outcome.
- **OSFD median (IQR):** Among the patients with a known 21-day outcome, we compute the 25th, 50th, and 75th percentiles of the Organ-Support Free-Days endpoint. The interquartile range (IQR) is shown in parentheses as the range between the 25th and 75th percentiles.
- **Conditional OSFD:** Among the patients with a known 21-day outcome that were not deceased, we compute the 25th, 50th, and 75th percentiles of the Organ-Support Free-Days endpoint. The interquartile range (IQR) is shown in parentheses as the range between the 25th and 75th percentiles.
- For the subjects that are eligible for the domain, we show a plot of the cumulative distribution function for the OSFD endpoint for each intervention within the domain.
- For the subjects that are eligible for the domain, we show a stacked bar plot for the OSFD outcomes for each intervention within the domain. Red represents worse outcomes and blue represents better outcomes.

#### 2.2 Overall Summaries

Figure 2.1 displays the distribution of outcomes on the primary endpoint for all patients in the analysis population (across all domains), without respect to treatment assignments. Table 2.1 provides descriptive summaries of the OSFD and in-hospital mortality outcomes for all patients in the analysis population.

**Table 2.1:** Overall summary of the OSFD and In-Hospital mortality data (REMAP-CAP COVID-19 severe and moderate state ITT population)

| Participant Group | Number Assigned ($N$) | Number Past Day 21 | Number Known ($n$) | Number Deaths ($y$) | Mortality Rate ($y/n$) | OSFD median (IQR) | Conditional* OSFD median (IQR) |
|---|---|---|---|---|---|---|---|
| Moderate State | 665 | 665 | 663 | 63 | 0.095 | 22.00 (21.00 – 22.00) | 22.00 (22.00 – 22.00) |
| Severe State | 5292 | 5292 | 5189 | 1800 | 0.347 | 3.00 (−1.00 – 16.00) | 14.00 (4.00 – 18.00) |

* Conditional OSFD reports the median and IQR for subjects that did not die.

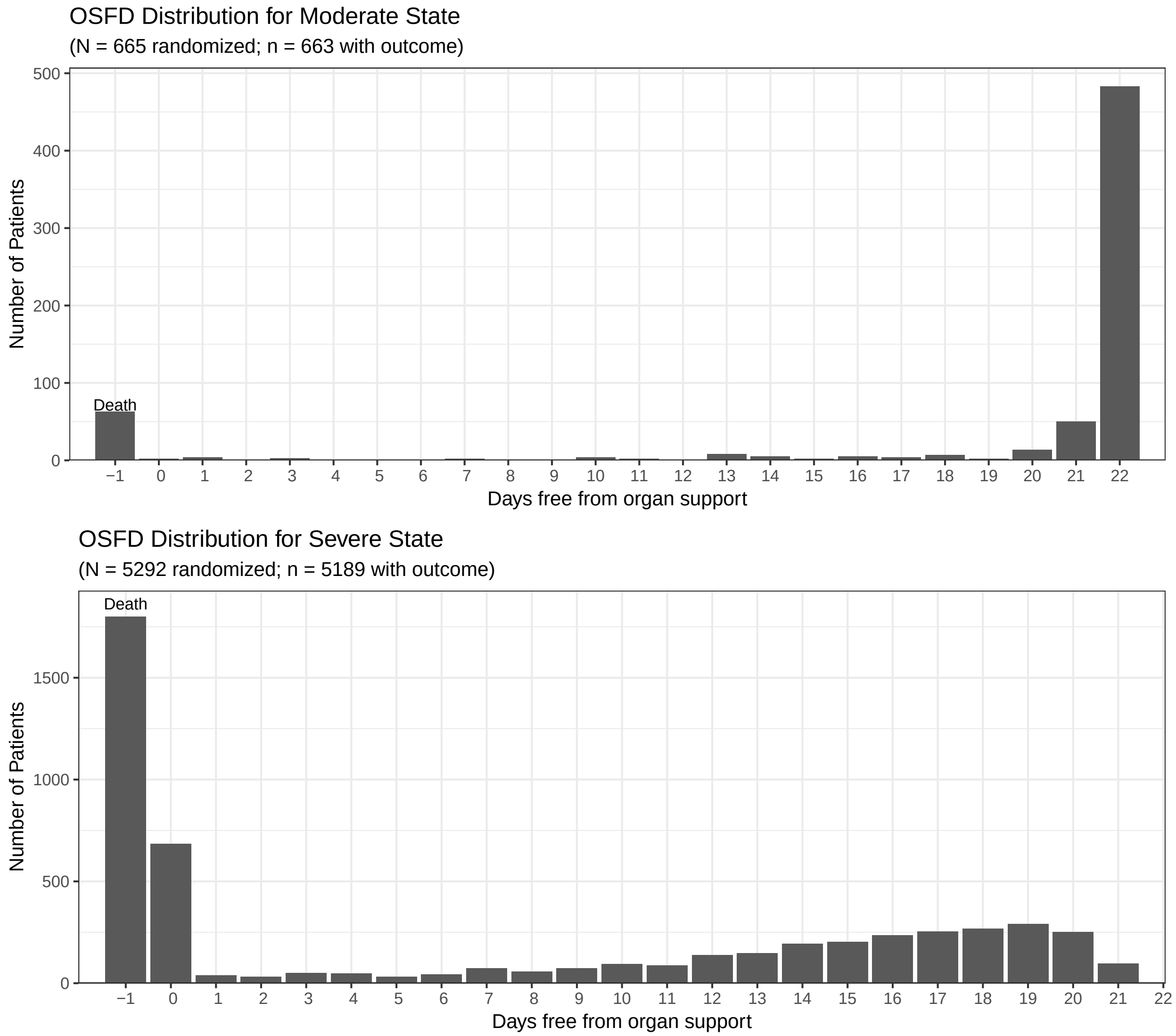

**Figure 2.1:** Overall distribution of the primary organ support free days endpoint for **Moderate** (top) and **Severe** (bottom) disease states.

#### 2.3 COVID-19 Immune Modulation Domain

##### 2.3.1 Description of the COVID-19 Immune Modulation domain

The COVID-19 Immune Modulation domain includes a total of 5 interventions. This domain:

- was available for randomization in both Moderate and Severe disease states;
- closed randomization in both Severe and Moderate states on April 10, 2021;
- was an immediate reveal domain with immediate initiation of the randomized assignment, unless prospective agreement to participate is required, in which case it was deferred reveal domain;
- had no strata identified as being of interest. Analyses and response adaptive randomization were applied to all randomized patients, by disease state;
- has possible interactions modeled with the corticosteroid domain in Severe disease state; A previous study suggested that the interaction of interferon-$\beta$ and corticosteroids may be harmful; therefore an informative prior is used to reflect a harmful interaction. Furthermore, initial (burn-in) randomization probabilities were constructed to limit the number of patients randomized to the combination of corticosteroids and interferon-$\beta$;
- has one nest, comprised of tocilizumab and sarilumab, which are both interleukin-6 inhibitors;
- includes borrowing at the intervention level between the Moderate and Severe disease states;
- has one nest, comprised of tocilizumab and sarilumab, which are both interleukin-6 inhibitors;
- includes borrowing at the intervention level between the Moderate and Severe disease states for interventions that are not nested;
- enforced a minimum marginal RAR probability of 0.05 for each active intervention within the domain, by disease state.

##### 2.3.2 Observed data within the COVID-19 Immune Modulation domain

Table 2.2 shows the number of patients assigned to each intervention within the Immune Modulation domain, by eligibility category within each disease state. The category "Eligibility not assessed in this state" refers to patients who had randomization assignments in one or more domains in both disease states. For example, of the 665 patients in moderate disease state, 9 patients had assignments in other moderate domains, but their assignment the Immune Modulation domain occurred when they were in severe disease state.

**Table 2.2:** Summary of the availability of data (**COVID-19 Immune Modulation** domain)

| Intervention | Number Eligible | Number Assigned | Number Past Day 21 | Number Missing | Number Known |
|---|---|---|---|---|---|
| **Moderate State: N = 665** | | | | | |
| *Eligible for domain: N=5* | | | | | |
| No immune modulation for COVID-19 | 5 | 3 | 3 | 0 | 3 |
| Anakinra | 4 | 2 | 2 | 0 | 2 |
| *Not eligible for domain: N=4* | | | | | |
| No assignment | | 4 | 4 | 0 | 4 |
| *Domain not active/not available: N=647* | | | | | |
| No assignment | | 647 | 647 | 2 | 645 |
| *Eligibility not assessed in this state: N=9* | | | | | |
| No assignment | | 9 | 9 | 0 | 9 |
| **Severe State: N = 5292** | | | | | |
| *Eligible for domain: N=2241* | | | | | |
| No assignment | | 6 | 6 | 0 | 6 |
| No immune modulation for COVID-19 | 872 | 406 | 406 | 0 | 406 |
| Interferon Beta-1a | 62 | 19 | 19 | 0 | 19 |
| Anakinra | 1248 | 373 | 373 | 8 | 365 |
| Tocilizumab | 1989 | 952 | 952 | 9 | 943 |
| Sarilumab | 911 | 485 | 485 | 2 | 483 |
| *Not eligible for domain: N=1151* | | | | | |
| No assignment | | 1151 | 1151 | 7 | 1144 |
| *Domain not active/not available: N=1900* | | | | | |
| No assignment | | 1900 | 1900 | 77 | 1823 |

**Table 2.3:** Summary of the OSFD and In-Hospital mortality data for patients that were eligible for the **COVID-19 Immune Modulation** domain

| Intervention | Number Assigned ($N$) | Number Known ($n$) | Number of Deaths ($y$) | Mortality Rate ($y/n$) | OSFD median (IQR) | Conditional* OSFD median (IQR) |
|---|---|---|---|---|---|---|
| **Moderate State** | | | | | | |
| No immune modulation for COVID-19 | 3 | 3 | 0 | 0.000 | 22.00 (22.00 – 22.00) | 22.00 (22.00 – 22.00) |
| Anakinra | 2 | 2 | 0 | 0.000 | 21.00 (20.50 – 21.50) | 21.00 (20.50 – 21.50) |
| **Severe State** | | | | | | |
| No immune modulation for COVID-19 | 406 | 406 | 150 | 0.369 | 0.00 (−1.00 – 15.00) | 13.00 (4.00 – 17.00) |
| Interferon Beta-1a | 19 | 19 | 7 | 0.368 | 0.00 (−1.00 – 13.50) | 11.00 (0.00 – 17.25) |
| Anakinra | 373 | 365 | 145 | 0.397 | 0.00 (−1.00 – 15.00) | 14.00 (3.75 – 18.00) |
| Tocilizumab | 952 | 943 | 317 | 0.336 | 7.00 (−1.00 – 16.00) | 15.00 (7.25 – 18.00) |
| Sarilumab | 485 | 483 | 158 | 0.327 | 9.00 (−1.00 – 17.00) | 15.00 (9.00 – 18.00) |

* Conditional OSFD reports the median and IQR for subjects that did not die.

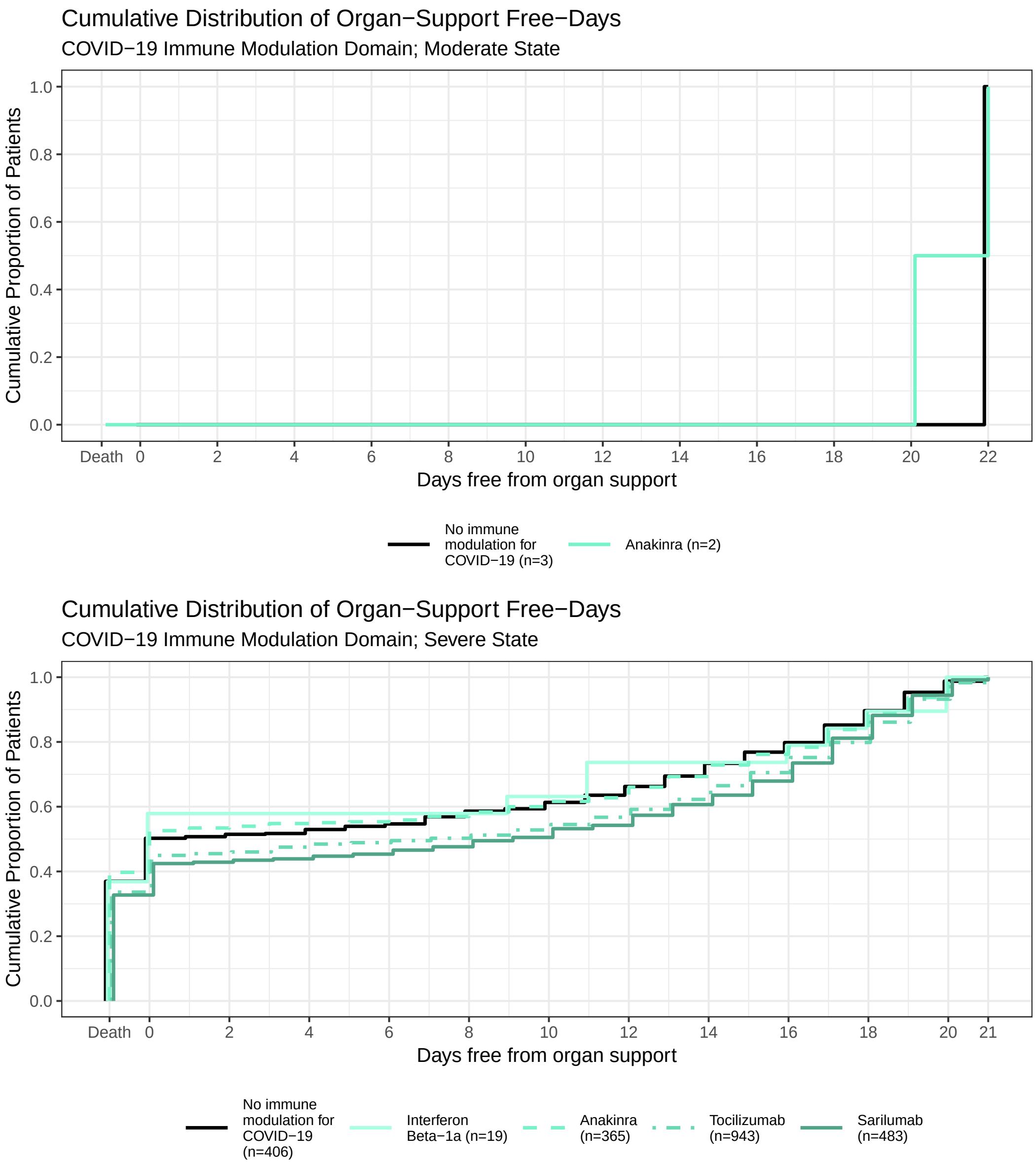

**Figure 2.2:** Empirical cumulative distribution of organ support free days for each intervention in the **COVID-19 Immune Modulation** domain. This plot is restricted to patients who were eligible for the domain and had a revealed assignment.

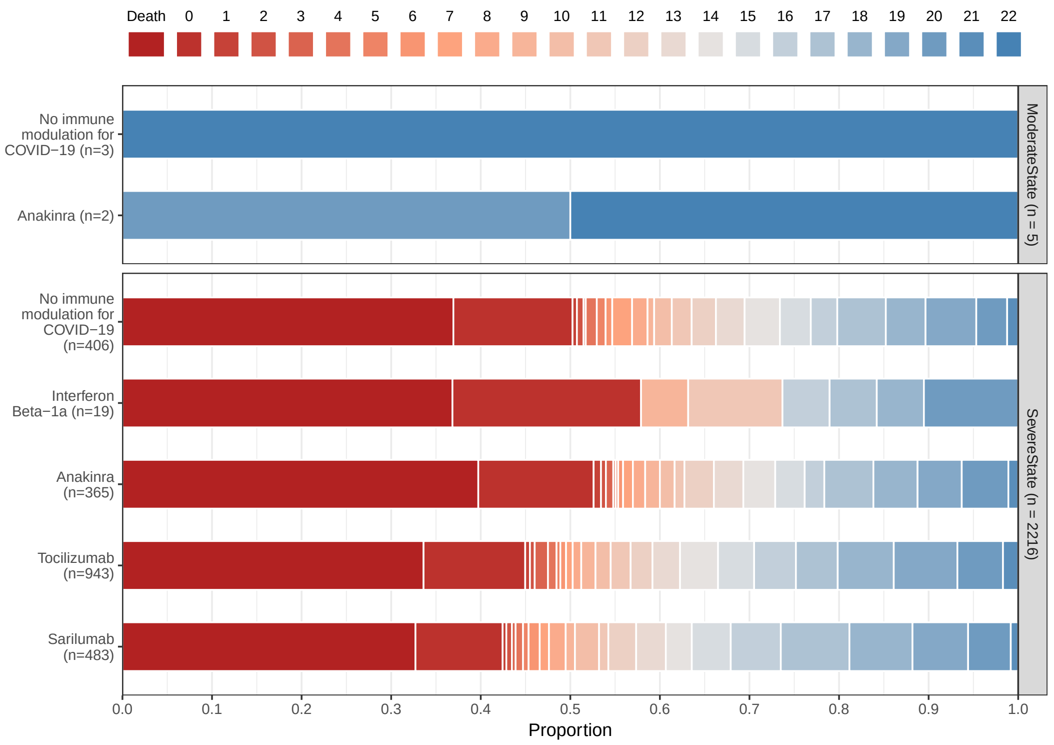

**Figure 2.3:** Stacked proportion of organ support free days for each intervention in the **COVID-19 Immune Modulation** domain. Red represents worse outcomes and blue represents better outcomes. This plot is restricted to patients who were eligible for the domain and have a revealed assignment in the domain.

### 3 Analysis Results and Conclusions

#### 3.1 Overview of the Statistical Model

The primary analysis model is a Bayesian cumulative logistic model for the ordinal primary endpoint. The full statistical model includes data from all interventions within all domains; however this report shows results only for the Immune Modulation domain. The model adjusts for age, sex, site (nested within country), domain ineligibility, randomization within each domain, and time epochs. Borrowing of information between moderate and severe disease states is accomplished through hierarchical modeling. Additionally, the model includes a term for patients that transition from moderate to severe disease state. The model is constructed so that odds-ratios greater than 1 indicate improved outcomes.

#### 3.2 Definition of Statistical Triggers

The adaptive design defines several statistical triggers within the trial, that at any analysis of the trial would result in public disclosure and declaration of a platform conclusion. The following statistical triggers were defined for the COVID-19 Immune Modulation domain:

1. **Domain Superiority.** If a single intervention within a domain has at least a 99% posterior probability of being in the best regimen for patients in the severe state of the PISOP stratum, this would trigger superiority of that intervention.
2. **Intervention Efficacy.** If an intervention is deemed to have at least a 99% posterior probability of being superior to the control, then a declaration of efficacy of that intervention would be declared. This statistical trigger is active for each of the non-control arms in the domain.

3. **Intervention Equivalence.** If two non-control interventions have a 90% probability of equivalence — that is, that the odds ratio comparing the two interventions is between 0.83 (inverse of 1/1.2 and 1.2) — then a declaration of intervention equivalence would be made.

4. **Intervention Futility.** If an intervention is deemed to have less than 5% posterior probability of at least a 20% odds ratio improvement compared to the control, then a declaration of futility of that intervention would be declared. This statistical trigger is active for each of the non-control arms in the domain.

5. **Intervention Inferiority.** If an intervention has low posterior probability of being the optimal intervention within a state, then that intervention will be deemed inferior. Specifically, an intervention is considered inferior if the probability of being the optimal intervention is less than $0.01/(J_d - 1)$, where $J_d$ is the number of interventions within the domain. For the COVID-19 Immune Modulation domain, there were initially $J = 5$ interventions.

#### 3.3 Overview of the model results

The OSFD endpoint is an ordered categorical endpoint that is modeled with a cumulative logistic model. The median and 95% Bayesian credible intervals for the odds-ratios are presented for each intervention, relative to the control intervention in the domain. The model is structured so that an odds-ratio greater than 1 implies patient benefit. We also present the posterior mean and standard deviation, but caution that the mean tends to be higher than the median due to the skewed nature of the posterior distribution.

#### 3.4 Primary analysis for OSFD

Table 3.1 summarizes the model-estimated odds-ratios for the covariates in the model, including age categories, sex at birth, and time effects.

**Table 3.1:** Model-estimated Odds-Ratios for the **OSFD** endpoint; REMAP-CAP COVID-19 severe state ITT population

| Odds-Ratio Parameter | Moderate State | | | Severe State | | |
|---|---|---|---|---|---|---|
| | Median | 95% Credible Interval | Mean (SD) | Median | 95% Credible Interval | Mean (SD) |
| $\leq 39$ | 2.01 | 1.04 – 3.98 | 2.14 (0.78) | 3.96 | 3.27 – 4.83 | 3.98 (0.41) |
| 40-49 | 2.29 | 1.17 – 4.72 | 2.46 (0.91) | 2.38 | 2.03 – 2.81 | 2.39 (0.20) |
| 50-59 | 1.15 | 0.67 – 1.90 | 1.18 (0.31) | 1.82 | 1.59 – 2.08 | 1.82 (0.13) |
| 60-69 (referent) | 1.00 | $NA - NA$ | 1.00 ($NA$) | 1.00 | $NA - NA$ | 1.00 ($NA$) |
| 70-79 | 0.65 | 0.37 – 1.12 | 0.67 (0.20) | 0.54 | 0.46 – 0.63 | 0.54 (0.04) |
| 80+ | 0.45 | 0.23 – 0.91 | 0.48 (0.18) | 0.32 | 0.23 – 0.43 | 0.32 (0.05) |
| Male (referent) | 1.00 | $NA - NA$ | 1.00 ($NA$) | 1.00 | $NA - NA$ | 1.00 ($NA$) |
| Female | 1.38 | 0.92 – 2.05 | 1.41 (0.29) | 1.17 | 1.05 – 1.30 | 1.17 (0.06) |
| Time Epoch-0 (referent) | 1.00 | $NA - NA$ | 1.00 ($NA$) | 1.00 | $NA - NA$ | 1.00 ($NA$) |
| Time Epoch-1 | 0.98 | 0.78 – 1.18 | 0.98 (0.10) | 0.97 | 0.85 – 1.14 | 0.97 (0.07) |
| Time Epoch-2 | 0.95 | 0.62 – 1.33 | 0.95 (0.18) | 0.90 | 0.74 – 1.13 | 0.91 (0.10) |
| Time Epoch-3 | 0.92 | 0.49 – 1.48 | 0.93 (0.25) | 0.80 | 0.63 – 1.02 | 0.81 (0.10) |
| Time Epoch-4 | 0.88 | 0.39 – 1.60 | 0.90 (0.31) | 0.74 | 0.57 – 0.95 | 0.74 (0.10) |
| Time Epoch-5 | 0.83 | 0.32 – 1.66 | 0.86 (0.35) | 0.72 | 0.56 – 0.94 | 0.73 (0.10) |
| Time Epoch-6 | 0.79 | 0.26 – 1.73 | 0.83 (0.39) | 0.75 | 0.58 – 0.99 | 0.76 (0.11) |
| Time Epoch-7 | 0.74 | 0.22 – 1.79 | 0.80 (0.41) | 0.79 | 0.60 – 1.06 | 0.80 (0.12) |
| Time Epoch-8 | 0.70 | 0.18 – 1.78 | 0.76 (0.42) | 0.85 | 0.64 – 1.15 | 0.86 (0.13) |
| Time Epoch-9 | 0.65 | 0.16 – 1.74 | 0.71 (0.42) | 0.89 | 0.66 – 1.21 | 0.90 (0.14) |
| Time Epoch-10 | 0.60 | 0.13 – 1.69 | 0.67 (0.41) | 0.90 | 0.66 – 1.25 | 0.92 (0.15) |
| Time Epoch-11 | 0.54 | 0.12 – 1.62 | 0.62 (0.40) | 0.93 | 0.67 – 1.30 | 0.94 (0.16) |
| Time Epoch-12 | 0.49 | 0.11 – 1.60 | 0.58 (0.39) | 0.95 | 0.68 – 1.37 | 0.97 (0.18) |
| Time Epoch-13 | 0.47 | 0.10 – 1.62 | 0.56 (0.39) | 1.01 | 0.71 – 1.46 | 1.03 (0.19) |
| Time Epoch-14 | 0.46 | 0.10 – 1.65 | 0.55 (0.40) | 1.08 | 0.75 – 1.59 | 1.10 (0.21) |
| Time Epoch-15 | 0.46 | 0.10 – 1.75 | 0.56 (0.42) | 1.14 | 0.78 – 1.69 | 1.17 (0.23) |
| Time Epoch-16 | 0.47 | 0.10 – 1.90 | 0.59 (0.46) | 1.18 | 0.80 – 1.77 | 1.21 (0.24) |
| Time Epoch-17 | 0.50 | 0.10 – 2.07 | 0.63 (0.52) | 1.21 | 0.83 – 1.81 | 1.24 (0.25) |
| Time Epoch-18 | 0.53 | 0.11 – 2.25 | 0.68 (0.60) | 1.23 | 0.85 – 1.84 | 1.26 (0.25) |
| Time Epoch-19 | 0.56 | 0.11 – 2.49 | 0.73 (0.67) | 1.20 | 0.83 – 1.80 | 1.24 (0.25) |
| Time Epoch-20 | 0.60 | 0.12 – 2.88 | 0.80 (0.77) | 1.16 | 0.79 – 1.73 | 1.19 (0.24) |
| Time Epoch-21 | 0.64 | 0.12 – 3.26 | 0.90 (0.91) | 1.07 | 0.72 – 1.62 | 1.09 (0.22) |
| Time Epoch-22 | 0.69 | 0.12 – 3.99 | 1.04 (1.13) | 0.99 | 0.65 – 1.50 | 1.01 (0.21) |
| Time Epoch-23 | 0.75 | 0.12 – 5.23 | 1.23 (1.52) | 0.91 | 0.59 – 1.40 | 0.93 (0.20) |
| Time Epoch-24 | 0.81 | 0.11 – 7.12 | 1.52 (2.17) | 0.87 | 0.56 – 1.35 | 0.89 (0.20) |
| Time Epoch-25 | | | | 0.85 | 0.54 – 1.36 | 0.88 (0.21) |
| Time Epoch-26 | | | | 0.85 | 0.50 – 1.46 | 0.88 (0.25) |
| ModToSevere | | | | 1.12 | 0.53 – 2.32 | 1.20 (0.47) |
| Interferon | 1.71 | 0.18 – 36.87 | 87877.90 (6199194.59) | 1.75 | 0.48 – 6.60 | 2.16 (1.63) |
| Anakinra | 0.99 | 0.26 – 6.47 | 1.67 (4.85) | 0.99 | 0.74 – 1.35 | 1.00 (0.16) |
| Tocilizumab | | | | 1.46 | 1.13 – 1.87 | 1.47 (0.19) |
| Sarilumab | | | | 1.50 | 1.13 – 2.00 | 1.52 (0.22) |

*Note:* For referent categories, the Odds-Ratio is 1.0 by definition. Time bucket X is the Xth 2-week interval prior to the most recent month, and Odds-Ratios are relative to the most recent month.

**Table 3.2:** Posterior Probabilities for the **OSFD** endpoint; REMAP-CAP COVID-19 severe and moderate state ITT population

| Intervention | Decision Quantity | Posterior Probability |
|---|---|---|
| **Moderate** | | |
| *Efficacy* | | |
| Interferon | Prob(OR > 1) | 0.74980 |
| Anakinra | Prob(OR > 1) | 0.49210 |
| *Equivalence* | | |
| Interferon/Anakinra | Prob(equivalent) | 0.12260 |
| *Futility* | | |
| Interferon | Prob(OR < 1.2) | 0.32255 |
| Anakinra | Prob(OR < 1.2) | 0.70985 |
| *Superiority/Inferiority* | | |
| No Immune Modulation | Prob(optimal) | 0.14200 |
| Interferon | Prob(optimal) | 0.66050 |
| Anakinra | Prob(optimal) | 0.19750 |
| **Severe** | | |
| *Efficacy* | | |
| Interferon | Prob(OR > 1) | 0.81630 |
| Anakinra | Prob(OR > 1) | 0.46595 |
| Tocilizumab | Prob(OR > 1) | 0.99785 |
| Sarilumab | Prob(OR > 1) | 0.99750 |
| *Equivalence* | | |
| Interferon/Anakinra | Prob(equivalent) | 0.15025 |
| Interferon/Tocilizumab | Prob(equivalent) | 0.22600 |
| Interferon/Sarilumab | Prob(equivalent) | 0.22480 |
| Anakinra/Tocilizumab | Prob(equivalent) | 0.07250 |
| Anakinra/Sarilumab | Prob(equivalent) | 0.06065 |
| Tocilizumab/Sarilumab | Prob(equivalent) | 0.84910 |
| *Futility* | | |
| Interferon | Prob(OR < 1.2) | 0.26500 |
| Anakinra | Prob(OR < 1.2) | 0.88905 |
| Tocilizumab | Prob(OR < 1.2) | 0.06510 |
| Sarilumab | Prob(OR < 1.2) | 0.05790 |
| *Superiority/Inferiority* | | |
| No Immune Modulation | Prob(optimal) | 0.00005 |
| Interferon | Prob(optimal) | 0.25545 |
| Anakinra | Prob(optimal) | 0.00020 |
| Tocilizumab | Prob(optimal) | 0.27970 |
| Sarilumab | Prob(optimal) | 0.46460 |

**Table 3.3:** Model-estimated Odds-Ratios for the **Mortality** endpoint; REMAP-CAP COVID-19 severe state ITT population

| | Moderate State | | | Severe State | | |
|---|---|---|---|---|---|---|
| Odds-Ratio Parameter | Median | 95% Credible Interval | Mean (SD) | Median | 95% Credible Interval | Mean (SD) |
| $\leq 39$ | 10.26 | 2.55 – 52.30 | 14.45 (14.11) | 8.03 | 5.58 – 11.78 | 8.19 (1.58) |
| 40-49 | 3.41 | 1.19 – 11.25 | 4.13 (2.78) | 3.46 | 2.75 – 4.37 | 3.48 (0.41) |
| 50-59 | 2.47 | 1.15 – 5.87 | 2.74 (1.24) | 2.37 | 2.01 – 2.84 | 2.38 (0.21) |
| 60-69 (referent) | 1.00 | $NA - NA$ | 1.00 ($NA$) | 1.00 | $NA - NA$ | 1.00 ($NA$) |
| 70-79 | 0.47 | 0.23 – 0.96 | 0.50 (0.19) | 0.48 | 0.40 – 0.57 | 0.48 (0.04) |
| 80+ | 0.30 | 0.13 – 0.67 | 0.32 (0.14) | 0.27 | 0.19 – 0.37 | 0.27 (0.05) |
| Male (referent) | 1.00 | $NA - NA$ | 1.00 ($NA$) | 1.00 | $NA - NA$ | 1.00 ($NA$) |
| Female | 2.29 | 1.23 – 4.20 | 2.39 (0.77) | 1.25 | 1.09 – 1.43 | 1.25 (0.09) |
| Time Epoch-0 (referent) | 1.00 | $NA - NA$ | 1.00 ($NA$) | 1.00 | $NA - NA$ | 1.00 ($NA$) |
| Time Epoch-1 | 0.94 | 0.76 – 1.20 | 0.94 (0.11) | 0.92 | 0.80 – 1.08 | 0.92 (0.07) |
| Time Epoch-2 | 0.86 | 0.57 – 1.37 | 0.89 (0.20) | 0.83 | 0.66 – 1.08 | 0.84 (0.11) |
| Time Epoch-3 | 0.80 | 0.45 – 1.52 | 0.85 (0.28) | 0.75 | 0.56 – 1.04 | 0.76 (0.13) |
| Time Epoch-4 | 0.73 | 0.35 – 1.63 | 0.80 (0.33) | 0.69 | 0.49 – 1.01 | 0.71 (0.13) |
| Time Epoch-5 | 0.66 | 0.28 – 1.66 | 0.74 (0.37) | 0.66 | 0.47 – 0.99 | 0.68 (0.13) |
| Time Epoch-6 | 0.60 | 0.24 – 1.69 | 0.70 (0.40) | 0.67 | 0.46 – 1.01 | 0.68 (0.14) |
| Time Epoch-7 | 0.54 | 0.20 – 1.69 | 0.65 (0.41) | 0.70 | 0.48 – 1.08 | 0.72 (0.16) |
| Time Epoch-8 | 0.49 | 0.17 – 1.63 | 0.60 (0.41) | 0.75 | 0.49 – 1.19 | 0.77 (0.18) |
| Time Epoch-9 | 0.44 | 0.15 – 1.57 | 0.55 (0.41) | 0.81 | 0.52 – 1.31 | 0.83 (0.20) |
| Time Epoch-10 | 0.40 | 0.13 – 1.50 | 0.51 (0.40) | 0.86 | 0.54 – 1.44 | 0.89 (0.23) |
| Time Epoch-11 | 0.36 | 0.12 – 1.47 | 0.48 (0.39) | 0.89 | 0.55 – 1.51 | 0.93 (0.25) |
| Time Epoch-12 | 0.34 | 0.11 – 1.46 | 0.45 (0.38) | 0.92 | 0.56 – 1.56 | 0.96 (0.26) |
| Time Epoch-13 | 0.32 | 0.10 – 1.46 | 0.44 (0.38) | 0.97 | 0.58 – 1.68 | 1.01 (0.29) |
| Time Epoch-14 | 0.31 | 0.09 – 1.47 | 0.44 (0.38) | 1.03 | 0.62 – 1.82 | 1.08 (0.31) |
| Time Epoch-15 | 0.31 | 0.09 – 1.50 | 0.44 (0.38) | 1.10 | 0.66 – 1.94 | 1.15 (0.34) |
| Time Epoch-16 | 0.32 | 0.09 – 1.54 | 0.46 (0.40) | 1.16 | 0.70 – 2.03 | 1.21 (0.35) |
| Time Epoch-17 | 0.33 | 0.09 – 1.66 | 0.48 (0.43) | 1.22 | 0.74 – 2.13 | 1.27 (0.37) |
| Time Epoch-18 | 0.35 | 0.09 – 1.85 | 0.51 (0.47) | 1.26 | 0.78 – 2.21 | 1.32 (0.38) |
| Time Epoch-19 | 0.36 | 0.09 – 2.10 | 0.53 (0.52) | 1.29 | 0.81 – 2.25 | 1.35 (0.38) |
| Time Epoch-20 | 0.36 | 0.08 – 2.27 | 0.56 (0.59) | 1.31 | 0.82 – 2.33 | 1.37 (0.39) |
| Time Epoch-21 | 0.37 | 0.07 – 2.49 | 0.61 (0.71) | 1.29 | 0.81 – 2.26 | 1.35 (0.38) |
| Time Epoch-22 | 0.39 | 0.07 – 2.89 | 0.68 (0.91) | 1.24 | 0.77 – 2.15 | 1.29 (0.36) |
| Time Epoch-23 | 0.42 | 0.06 – 3.56 | 0.78 (1.25) | 1.18 | 0.72 – 2.03 | 1.22 (0.34) |
| Time Epoch-24 | 0.45 | 0.05 – 4.66 | 0.93 (1.84) | 1.14 | 0.66 – 1.96 | 1.19 (0.34) |
| Time Epoch-25 | | | | 1.14 | 0.62 – 2.05 | 1.19 (0.37) |
| Time Epoch-26 | | | | 1.16 | 0.57 – 2.34 | 1.23 (0.45) |
| ModToSevere | | | | 0.99 | 0.41 – 2.65 | 1.13 (0.58) |
| Interferon | 1.60 | 0.00 – 14.50 | 9.06 (234.70) | 1.73 | 0.48 – 6.85 | 2.20 (1.73) |
| Anakinra | 1.00 | 0.32 – 7.64 | 28.15 (935.25) | 0.97 | 0.66 – 1.40 | 0.99 (0.19) |
| Tocilizumab | | | | 1.42 | 1.05 – 1.93 | 1.44 (0.23) |
| Sarilumab | | | | 1.51 | 1.06 – 2.20 | 1.54 (0.29) |

*Note:* For referent categories, the Odds-Ratio is 1.0 by definition. Time bucket X is the Xth 2-week interval prior to the most recent month, and Odds-Ratios are relative to the most recent month.

#### 3.5 Primary analysis for in-hospital mortality

**Table 3.4:** Posterior Probabilities for the **Mortality** endpoint; REMAP-CAP COVID-19 severe and moderate state ITT population

| Intervention | Decision Quantity | Posterior Probability |
|---|---|---|
| **Moderate** | | |
| *Efficacy* | | |
| Interferon | Prob(OR > 1) | 0.70860 |
| Anakinra | Prob(OR > 1) | 0.49795 |
| *Equivalence* | | |
| Interferon/Anakinra | Prob(equivalent) | 0.12055 |
| *Futility* | | |
| Interferon | Prob(OR < 1.2) | 0.36735 |
| Anakinra | Prob(OR < 1.2) | 0.70100 |
| *Superiority/Inferiority* | | |
| No Immune Modulation | Prob(optimal) | 0.15410 |
| Interferon | Prob(optimal) | 0.61840 |
| Anakinra | Prob(optimal) | 0.22750 |
| **Severe** | | |
| *Efficacy* | | |
| Interferon | Prob(OR > 1) | 0.80600 |
| Anakinra | Prob(OR > 1) | 0.43615 |
| Tocilizumab | Prob(OR > 1) | 0.98820 |
| Sarilumab | Prob(OR > 1) | 0.98805 |
| *Equivalence* | | |
| Interferon/Anakinra | Prob(equivalent) | 0.15720 |
| Interferon/Tocilizumab | Prob(equivalent) | 0.21070 |
| Interferon/Sarilumab | Prob(equivalent) | 0.20730 |
| Anakinra/Tocilizumab | Prob(equivalent) | 0.11040 |
| Anakinra/Sarilumab | Prob(equivalent) | 0.07930 |
| Tocilizumab/Sarilumab | Prob(equivalent) | 0.71400 |
| *Futility* | | |
| Interferon | Prob(OR < 1.2) | 0.28980 |
| Anakinra | Prob(OR < 1.2) | 0.86480 |
| Tocilizumab | Prob(OR < 1.2) | 0.14570 |
| Sarilumab | Prob(OR < 1.2) | 0.10145 |
| *Superiority/Inferiority* | | |
| No Immune Modulation | Prob(optimal) | 0.00125 |
| Interferon | Prob(optimal) | 0.30940 |
| Anakinra | Prob(optimal) | 0.00050 |
| Tocilizumab | Prob(optimal) | 0.18750 |
| Sarilumab | Prob(optimal) | 0.50135 |

#### 3.6 Sensitivity analysis of OSFD with less informative priors on interaction effects

To assess whether results are strongly influenced by the informative priors on the interaction terms between the immune modulation domain and the hydrocortisone intervention and the antiviral domain interventions, a pre-specified sensitivity analysis was planned in which the model would be evaluated with less informative priors on those interaction terms. However, data support for estimating interactions is weak. As seen in Table 3.6 below, only tocilizumab among the immune modulation interventions has sample support to estimate an interaction with the hydrocortisone intervention but with only 14 patients randomized to a regimen with the combination. The interaction with the antiviral domain has slightly better data support, though only for the lopinavir/ritonavir intervention. As a result of the lack of data support for estimating interactions, model convergence was extremely poor for the model with weakly informative priors. Thus, the results for this planned sensitivity analysis will not be presented.

**Table 3.5:** Number of patients assigned to combinations of interventions in the COVID-19 Immune Modulationdomain and selected other domains (**Moderate State**). This table only includes patients with known 21-day outcomes.

| Intervention | COVID-19 Immune Modulation | | | |
|---|---|---|---|---|
| | No assignment | No immune modulation | Anakinra | Sum |
| **COVID-19 Antiviral** | | | | |
| No assignment | 254 (38%) | 2 (0%) | 0 (0%) | 256 (39%) |
| No antiviral | 14 (2%) | 0 (0%) | 0 (0%) | 14 (2%) |
| Lopinavir/ritonavir | 3 (0%) | 1 (0%) | 2 (0%) | 6 (1%) |
| HCQ | 12 (2%) | 0 (0%) | 0 (0%) | 12 (2%) |
| Antiviral domain closed | 375 (57%) | 0 (0%) | 0 (0%) | 375 (57%) |
| Sum | 658 (99%) | 3 (0%) | 2 (0%) | 663 (100%) |

**Table 3.6:** Number of patients assigned to combinations of interventions in the COVID-19 Immune Modulationdomain and selected other domains (**Severe State**). This table only includes patients with known 21-day outcomes.

| Intervention | COVID-19 Immune Modulation | | | | | | |
|---|---|---|---|---|---|---|---|
| | No assignment | No immune modulation | Interferon Beta-1a | Anakinra | Tocilizumab | Sarilumab | Sum |
| **Corticosteroid** | | | | | | | |
| No assignment | 152 (3%) | 28 (1%) | 2 (0%) | 0 (0%) | 20 (0%) | 0 (0%) | 202 (4%) |
| No corticosteroids | 75 (1%) | 9 (0%) | 1 (0%) | 0 (0%) | 16 (0%) | 0 (0%) | 101 (2%) |
| Hydrocortisone | 94 (2%) | 27 (1%) | 0 (0%) | 0 (0%) | 14 (0%) | 0 (0%) | 135 (3%) |
| Shock dependent hydrocortisone | 108 (2%) | 18 (0%) | 1 (0%) | 1 (0%) | 18 (0%) | 0 (0%) | 146 (3%) |
| High-dose hydrocortisone | 2 (0%) | 0 (0%) | 0 (0%) | 0 (0%) | 0 (0%) | 0 (0%) | 2 (0%) |
| Steroid domain closed | 2542 (49%) | 324 (6%) | 15 (0%) | 364 (7%) | 875 (17%) | 483 (9%) | 4603 (89%) |
| Sum | 2973 (57%) | 406 (8%) | 19 (0%) | 365 (7%) | 943 (18%) | 483 (9%) | 5189 (100%) |
| **COVID-19 Antiviral** | | | | | | | |
| No assignment | 893 (17%) | 185 (4%) | 9 (0%) | 28 (1%) | 187 (4%) | 26 (1%) | 1328 (26%) |
| No antiviral | 132 (3%) | 115 (2%) | 2 (0%) | 6 (0%) | 91 (2%) | 16 (0%) | 362 (7%) |
| Lopinavir/ritonavir | 79 (2%) | 86 (2%) | 5 (0%) | 11 (0%) | 63 (1%) | 10 (0%) | 254 (5%) |
| HCQ | 22 (0%) | 17 (0%) | 0 (0%) | 1 (0%) | 9 (0%) | 0 (0%) | 49 (1%) |
| HCQ + lopinavir/ritonavir | 19 (0%) | 3 (0%) | 0 (0%) | 0 (0%) | 4 (0%) | 0 (0%) | 26 (1%) |
| Antiviral domain closed | 1828 (35%) | 0 (0%) | 3 (0%) | 319 (6%) | 589 (11%) | 431 (8%) | 3170 (61%) |
| Sum | 2973 (57%) | 406 (8%) | 19 (0%) | 365 (7%) | 943 (18%) | 483 (9%) | 5189 (100%) |

#### 3.7 Sensitivity analysis of the proportional odds assumption

An assumption of the ordinal logistic regression model being used to analyze OSFD is that effects have a *proportional* effect on log-odds. That is, a treatment effect that increases the log-odds of OSFD being greater than $X$ is the same for all values of $X$. In order to assess this modeling assumption, a logistic regression model is fit to dichotomized versions of the OSFD values ($\leq X$ versus $> X$) across the possible range of OSFD values, to see if the estimated treatment effect is nearly constant. The prior distribution for the dichotomized outcomes is constructed in like manner, converting the Dirichlet distribution across OSFD values to a Beta distribution by summing across the parameter values for the corresponding OSFD ranges.

The logistic regression model is subject to poor estimation when categories in the model contain only a single outcome type (e.g. all observations are $\leq X$). With the large number of covariate effects being used in the current model, some categories of these covariate crossings may contain single outcomes, particularly as the dichotomization goes to the higher end of the OSFD values with low frequencies. The Bayesian model uses informative priors and thus a model fit can always be constructed. However, because many of the prior distributions in the model are relatively non-informative, the MCMC fitting algorithms can perform poorly in the more extreme dichotomizations. Some poor MCMC behavior was observed for dichotomizations at $\geq 15$ OSFD and higher.

The odds ratio relative to control is reported for each dichtomization in the severe state only.

**Table 3.7:** Sensitivity analysis of proportional odds assumption. Values are Odds-Ratio estimates for the **OSFD** endpoint for different dichotomizations of OSFD; REMAP-CAP COVID-19 severe state ITT population (Interferon)

| | Interferon | | |
|---|---|---|---|
| OSFD Dichotomization | Median | 95% Credible Interval | Mean (SD) |
| ≤-1 vs ≥0 | 1.73 | 0.48 – 6.85 | 2.20 (1.73) |
| ≤0 vs ≥1 | 1.29 | 0.37 – 4.73 | 1.61 (1.23) |
| ≤1 vs ≥2 | 1.25 | 0.36 – 4.61 | 1.54 (1.12) |
| ≤2 vs ≥3 | 1.32 | 0.37 – 5.16 | 1.72 (1.66) |
| ≤3 vs ≥4 | 1.34 | 0.35 – 5.12 | 1.68 (1.28) |
| ≤4 vs ≥5 | 1.46 | 0.34 – 5.24 | 1.80 (1.39) |
| ≤5 vs ≥6 | 1.50 | 0.41 – 6.15 | 1.93 (1.54) |
| ≤6 vs ≥7 | 1.45 | 0.41 – 5.35 | 1.83 (1.37) |
| ≤7 vs ≥8 | 1.66 | 0.48 – 5.92 | 2.05 (1.56) |
| ≤8 vs ≥9 | 1.62 | 0.47 – 5.95 | 2.00 (1.46) |
| ≤9 vs ≥10 | 1.69 | 0.42 – 7.33 | 2.22 (1.94) |
| ≤10 vs ≥11 | 1.61 | 0.44 – 5.69 | 1.97 (1.44) |
| ≤11 vs ≥12 | 1.04 | 0.26 – 3.98 | 1.31 (1.01) |
| ≤12 vs ≥13 | 1.22 | 0.32 – 4.84 | 1.57 (1.30) |
| ≤13 vs ≥14 | 1.36 | 0.28 – 4.80 | 1.66 (1.24) |
| ≤14 vs ≥15 | 1.64 | 0.36 – 6.36 | 2.09 (1.67) |
| ≤15 vs ≥16 | 2.09 | 0.45 – 8.24 | 2.66 (2.26) |
| ≤16 vs ≥17 | 1.74 | 0.43 – 7.64 | 2.32 (2.26) |
| ≤17 vs ≥18 | 1.73 | 0.33 – 7.64 | 2.32 (2.27) |
| ≤18 vs ≥19 | 1.57 | 0.35 – 7.93 | 2.22 (2.16) |
| ≤19 vs ≥20 | 2.43 | 0.43 – 13.84 | 3.61 (4.09) |
| ≤20 vs ≥21 | 0.58 | 0.00 – 6.46 | 1.25 (2.80) |

**Table 3.8:** Sensitivity analysis of proportional odds assumption. Values are Odds-Ratio estimates for the **OSFD** endpoint for different dichotomizations of OSFD; REMAP-CAP COVID-19 severe state ITT population (Anakinra)

| | Anakinra | | |
|---|---|---|---|
| OSFD Dichotomization | Median | 95% Credible Interval | Mean (SD) |
| ≤-1 vs ≥0 | 0.97 | 0.66 – 1.40 | 0.99 (0.19) |
| ≤0 vs ≥1 | 0.98 | 0.69 – 1.39 | 0.99 (0.18) |
| ≤1 vs ≥2 | 0.96 | 0.68 – 1.36 | 0.97 (0.17) |
| ≤2 vs ≥3 | 0.98 | 0.69 – 1.42 | 1.00 (0.18) |
| ≤3 vs ≥4 | 0.92 | 0.66 – 1.33 | 0.94 (0.17) |
| ≤4 vs ≥5 | 0.96 | 0.69 – 1.35 | 0.97 (0.17) |
| ≤5 vs ≥6 | 0.98 | 0.68 – 1.39 | 0.99 (0.18) |
| ≤6 vs ≥7 | 0.98 | 0.69 – 1.40 | 0.99 (0.18) |
| ≤7 vs ≥8 | 1.02 | 0.71 – 1.47 | 1.04 (0.20) |
| ≤8 vs ≥9 | 1.03 | 0.72 – 1.44 | 1.04 (0.19) |
| ≤9 vs ≥10 | 0.97 | 0.70 – 1.38 | 0.99 (0.17) |
| ≤10 vs ≥11 | 1.02 | 0.71 – 1.46 | 1.03 (0.19) |
| ≤11 vs ≥12 | 1.05 | 0.73 – 1.51 | 1.08 (0.20) |
| ≤12 vs ≥13 | 1.06 | 0.73 – 1.51 | 1.07 (0.19) |
| ≤13 vs ≥14 | 1.05 | 0.72 – 1.50 | 1.07 (0.20) |
| ≤14 vs ≥15 | 1.08 | 0.73 – 1.60 | 1.10 (0.22) |
| ≤15 vs ≥16 | 1.04 | 0.69 – 1.60 | 1.06 (0.23) |
| ≤16 vs ≥17 | 1.03 | 0.70 – 1.55 | 1.05 (0.22) |
| ≤17 vs ≥18 | 1.08 | 0.71 – 1.64 | 1.11 (0.24) |
| ≤18 vs ≥19 | 0.99 | 0.60 – 1.60 | 1.02 (0.26) |
| ≤19 vs ≥20 | 0.92 | 0.47 – 1.70 | 0.97 (0.33) |
| ≤20 vs ≥21 | 0.50 | 0.15 – 1.57 | 0.59 (0.37) |

**Table 3.9:** Sensitivity analysis of proportional odds assumption. Values are Odds-Ratio estimates for the **OSFD** endpoint for different dichotomizations of OSFD; REMAP-CAP COVID-19 severe state ITT population (Tocilizumab)

| | Tocilizumab | | |
|---|---|---|---|
| OSFD Dichotomization | Median | 95% Credible Interval | Mean (SD) |
| ≤-1 vs ≥0 | 1.42 | 1.05 – 1.93 | 1.44 (0.23) |
| ≤0 vs ≥1 | 1.47 | 1.10 – 1.97 | 1.49 (0.22) |
| ≤1 vs ≥2 | 1.46 | 1.11 – 1.94 | 1.48 (0.21) |
| ≤2 vs ≥3 | 1.51 | 1.13 – 2.00 | 1.53 (0.22) |
| ≤3 vs ≥4 | 1.41 | 1.04 – 1.87 | 1.42 (0.21) |
| ≤4 vs ≥5 | 1.40 | 1.06 – 1.86 | 1.42 (0.20) |
| ≤5 vs ≥6 | 1.44 | 1.08 – 1.89 | 1.45 (0.21) |
| ≤6 vs ≥7 | 1.43 | 1.07 – 1.91 | 1.45 (0.21) |
| ≤7 vs ≥8 | 1.48 | 1.13 – 2.00 | 1.50 (0.23) |
| ≤8 vs ≥9 | 1.53 | 1.15 – 2.03 | 1.55 (0.23) |
| ≤9 vs ≥10 | 1.48 | 1.11 – 1.97 | 1.49 (0.22) |
| ≤10 vs ≥11 | 1.53 | 1.16 – 2.06 | 1.55 (0.23) |
| ≤11 vs ≥12 | 1.52 | 1.12 – 2.03 | 1.54 (0.23) |
| ≤12 vs ≥13 | 1.53 | 1.15 – 2.02 | 1.55 (0.22) |
| ≤13 vs ≥14 | 1.57 | 1.17 – 2.10 | 1.58 (0.24) |
| ≤14 vs ≥15 | 1.53 | 1.12 – 2.09 | 1.55 (0.25) |
| ≤15 vs ≥16 | 1.42 | 1.03 – 1.98 | 1.45 (0.25) |
| ≤16 vs ≥17 | 1.30 | 0.94 – 1.80 | 1.32 (0.22) |
| ≤17 vs ≥18 | 1.45 | 1.02 – 2.05 | 1.48 (0.26) |
| ≤18 vs ≥19 | 1.36 | 0.91 – 2.11 | 1.39 (0.31) |
| ≤19 vs ≥20 | 1.19 | 0.67 – 2.01 | 1.23 (0.35) |
| ≤20 vs ≥21 | 1.14 | 0.46 – 3.18 | 1.31 (0.71) |

**Table 3.10:** Sensitivity analysis of proportional odds assumption. Values are Odds-Ratio estimates for the **OSFD** endpoint for different dichotomizations of OSFD; REMAP-CAP COVID-19 severe state ITT population (Sarilumab)

| | Sarilumab | | |
|---|---|---|---|
| OSFD Dichotomization | Median | 95% Credible Interval | Mean (SD) |
| ≤-1 vs ≥0 | 1.51 | 1.06 – 2.20 | 1.54 (0.29) |
| ≤0 vs ≥1 | 1.68 | 1.22 – 2.33 | 1.70 (0.29) |
| ≤1 vs ≥2 | 1.67 | 1.21 – 2.32 | 1.70 (0.29) |
| ≤2 vs ≥3 | 1.73 | 1.23 – 2.39 | 1.75 (0.29) |
| ≤3 vs ≥4 | 1.68 | 1.19 – 2.34 | 1.70 (0.30) |
| ≤4 vs ≥5 | 1.70 | 1.22 – 2.40 | 1.72 (0.30) |
| ≤5 vs ≥6 | 1.72 | 1.23 – 2.38 | 1.74 (0.30) |
| ≤6 vs ≥7 | 1.66 | 1.21 – 2.29 | 1.68 (0.28) |
| ≤7 vs ≥8 | 1.75 | 1.23 – 2.42 | 1.76 (0.30) |
| ≤8 vs ≥9 | 1.68 | 1.22 – 2.40 | 1.71 (0.29) |
| ≤9 vs ≥10 | 1.65 | 1.19 – 2.26 | 1.67 (0.28) |
| ≤10 vs ≥11 | 1.62 | 1.16 – 2.27 | 1.64 (0.28) |
| ≤11 vs ≥12 | 1.72 | 1.24 – 2.48 | 1.76 (0.31) |
| ≤12 vs ≥13 | 1.68 | 1.19 – 2.36 | 1.70 (0.30) |
| ≤13 vs ≥14 | 1.65 | 1.17 – 2.37 | 1.68 (0.31) |
| ≤14 vs ≥15 | 1.72 | 1.18 – 2.49 | 1.75 (0.33) |
| ≤15 vs ≥16 | 1.57 | 1.08 – 2.32 | 1.61 (0.32) |
| ≤16 vs ≥17 | 1.34 | 0.93 – 1.94 | 1.37 (0.27) |
| ≤17 vs ≥18 | 1.27 | 0.84 – 1.94 | 1.30 (0.28) |
| ≤18 vs ≥19 | 1.04 | 0.64 – 1.70 | 1.08 (0.27) |
| ≤19 vs ≥20 | 0.84 | 0.44 – 1.61 | 0.89 (0.30) |
| ≤20 vs ≥21 | 0.45 | 0.11 – 1.80 | 0.57 (0.46) |

### 4 Other Data Summaries

This section provides summary tables and graphics for variables that are included as covariates in the model, including age, sex at birth, sites within country, and time effects.

**Table 4.1:** Summary of age groups by sex at birth (REMAP-CAP COVID-19 severe and moderate state ITT population)

| | Age Group (years) | | | | | | | |
|---|---|---|---|---|---|---|---|---|
| | ≤ 39 | 40 − 49 | 50 − 59 | 60 − 69 | 70 − 79 | ≥ 80 | Missing | Total |
| **Moderate State** | | | | | | | | |
| Male | 38 | 51 | 109 | 101 | 61 | 33 | 0 | 393 |
| Female | 25 | 41 | 70 | 57 | 48 | 30 | 1 | 272 |
| Sum | 63 | 92 | 179 | 158 | 109 | 63 | 1 | 665 |
| **Severe State** | | | | | | | | |
| Male | 232 | 457 | 911 | 1124 | 701 | 157 | 1 | 3583 |
| Female | 145 | 231 | 449 | 504 | 308 | 72 | 0 | 1709 |
| Total | 377 | 688 | 1360 | 1628 | 1009 | 229 | 1 | 5292 |

**Table 4.2:** Summary of the number of sites and patients randomized within each country (**Moderate** State)

| | | All Domains | | | COVID-19 Immune Modulation Domain | | |
|---|---|---|---|---|---|---|---|
| Region | Country | Number of Sites | Number of Patients Randomized | Number of Patients w/ Outcomes | Number of Sites | Number of Patients Randomized | Number of Patients w/ Outcomes |
| Americas | Canada | 2 | 15 | 15 | | | |
| | United States of America | 2 | 255 | 255 | | | |
| Asia | India | 3 | 42 | 42 | | | |
| | Nepal | 3 | 11 | 10 | | | |
| | Pakistan | 3 | 8 | 7 | | | |
| Europe | France | 1 | 5 | 5 | | | |
| | Netherlands | 2 | 47 | 47 | | | |
| | United Kingdom | 46 | 279 | 279 | 3 | 4 | 4 |
| Middle East | Saudi Arabia | 1 | 2 | 2 | | | |
| Oceana | Australia | 1 | 1 | 1 | 1 | 1 | 1 |

**Table 4.3:** Summary of the number of sites and patients randomized within each country (**Severe** State)

| | | All Domains | | | COVID-19 Immune Modulation Domain | | |
|---|---|---|---|---|---|---|---|
| Region | Country | Number of Sites | Number of Patients Randomized | Number of Patients w/ Outcomes | Number of Sites | Number of Patients Randomized | Number of Patients w/ Outcomes |
| Americas | Canada | 24 | 292 | 265 | 1 | 4 | 4 |
| | United States of America | 2 | 121 | 121 | | | |
| Asia | India | 4 | 29 | 21 | | | |
| | Nepal | 4 | 52 | 49 | | | |
| | Pakistan | 4 | 60 | 60 | | | |
| Europe | Finland | 1 | 3 | 3 | 1 | 3 | 3 |
| | France | 4 | 45 | 41 | | | |
| | Germany | 5 | 19 | 17 | | | |
| | Ireland | 3 | 66 | 66 | 3 | 38 | 38 |
| | Italy | 1 | 8 | 8 | 1 | 6 | 6 |
| | Netherlands | 11 | 269 | 244 | 11 | 179 | 175 |
| | Portugal | 1 | 3 | 3 | | | |
| | United Kingdom | 132 | 4093 | 4067 | 110 | 1918 | 1903 |
| Middle East | Saudi Arabia | 1 | 147 | 139 | 1 | 83 | 83 |
| Oceana | Australia | 23 | 75 | 75 | 2 | 2 | 2 |
| | New Zealand | 6 | 10 | 10 | 2 | 2 | 2 |

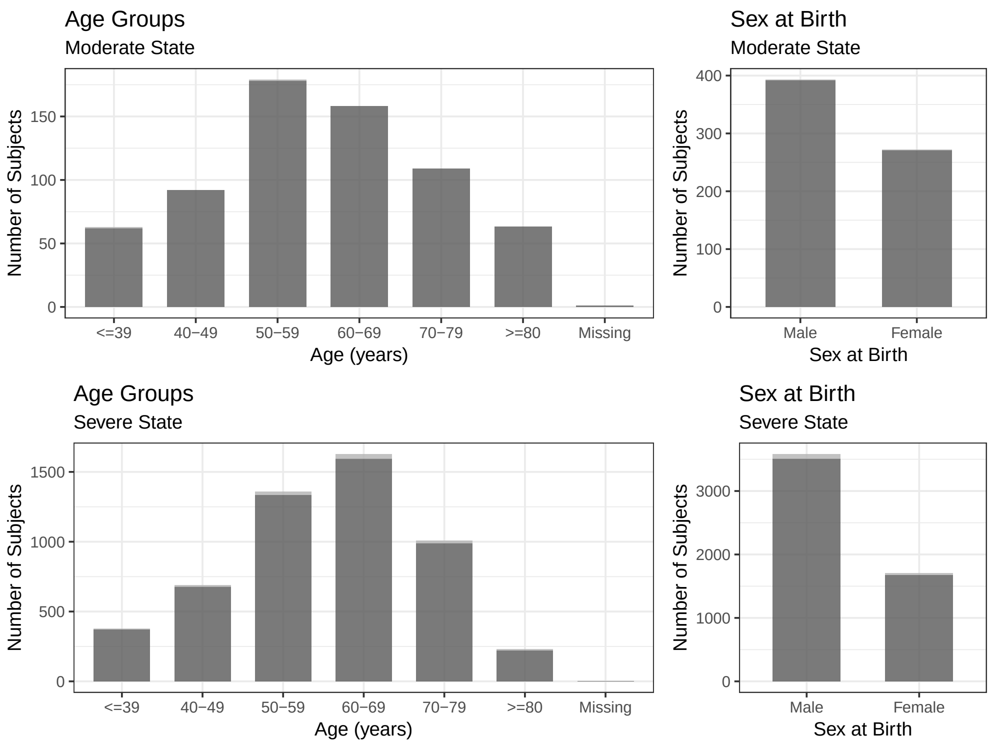

**Figure 4.1:** Distribution of age groups and sex at birth (REMAP-CAP COVID-19 severe and moderate state ITT population). The total height of each bar represents the number of patients in each category. The darker shaded area indicates the number of patients for whom 21 days have elapsed since randomization and have a known OSFD outcome. The lighter shaded area indicates the number of patients who do not have a known OSFD outcome.

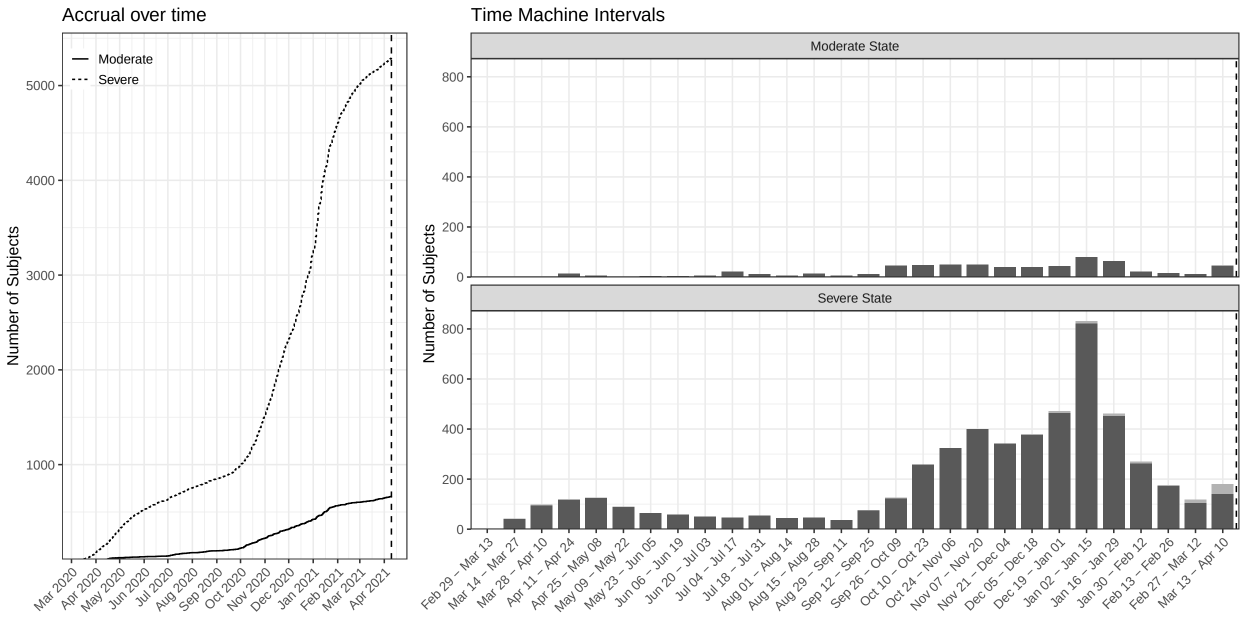

**Figure 4.2:** Accrual over time and distribution of patients within each of the time buckets used to estimate time trends in the analysis model for the REMAP-CAP COVID-19 severe and moderate state ITT population. The time buckets are derived so that the first bucket is the most recent month going backwards in time from the most recently randomized patient in the dataset that has an outcome. Thereafter, each bucket is defined as the next two-week interval backwards in time. The total height of each bar represents the number of patients in each category. The darker shaded area indicates the number of patients for whom 21 days have elapsed since randomization and have a known OSFD outcome. The lighter shaded area indicates the number of patients who do not have a known OSFD outcome. The vertical dashed line indicates the randomization date for the last patient who has passed 21 days and has a known outcome on the primary endpoint at the time of this analysis.

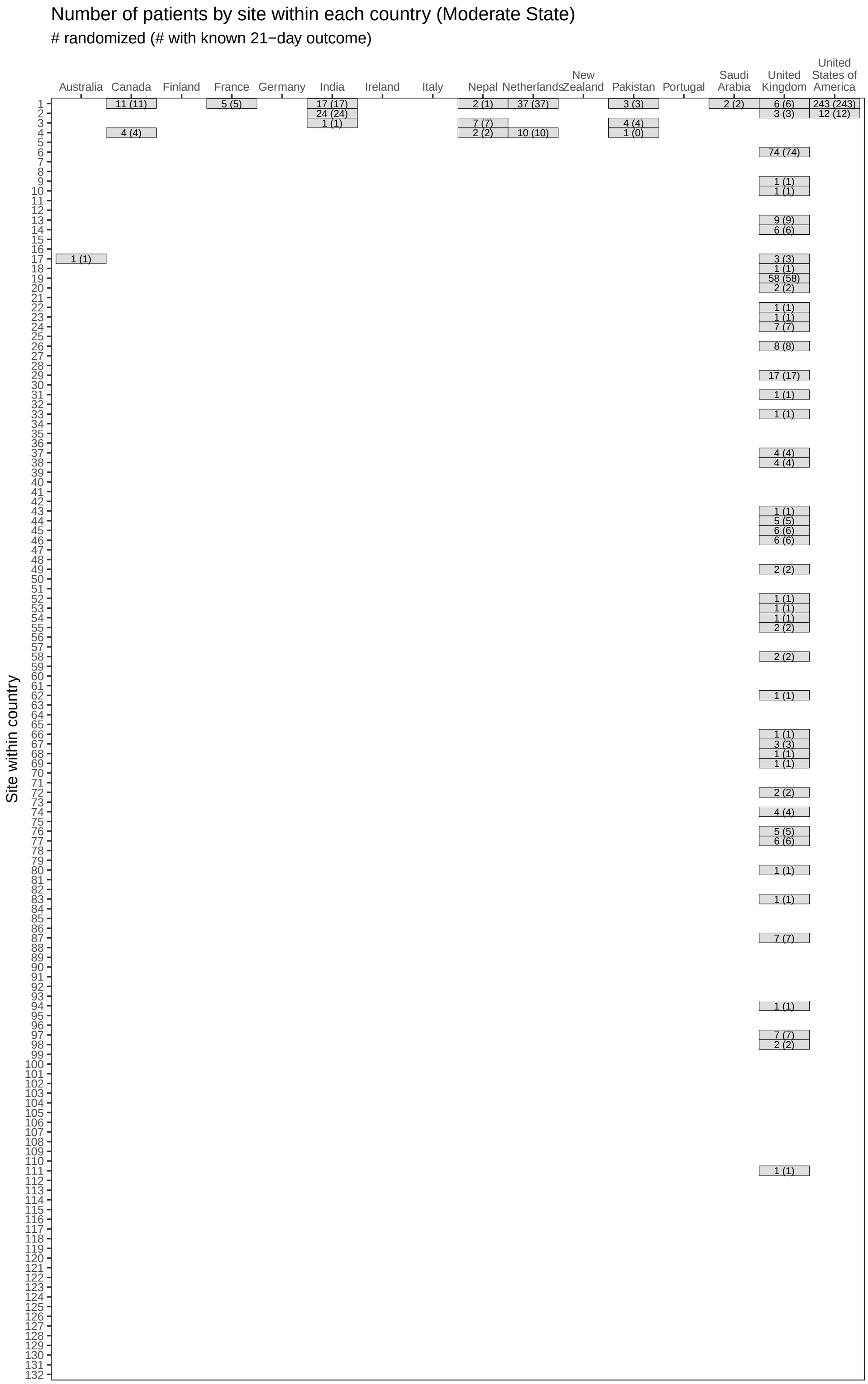

**Figure 4.3:** Sample size at each site within each country (**Moderate** State). The values in each cell represent the number of patients randomized to any domain at that site and, in parentheses, the number of patients for whom the outcome on the 21-day endpoint is known. Within each country, all sites having fewer than 5 randomized patients are combined into a single site for the statistical model.

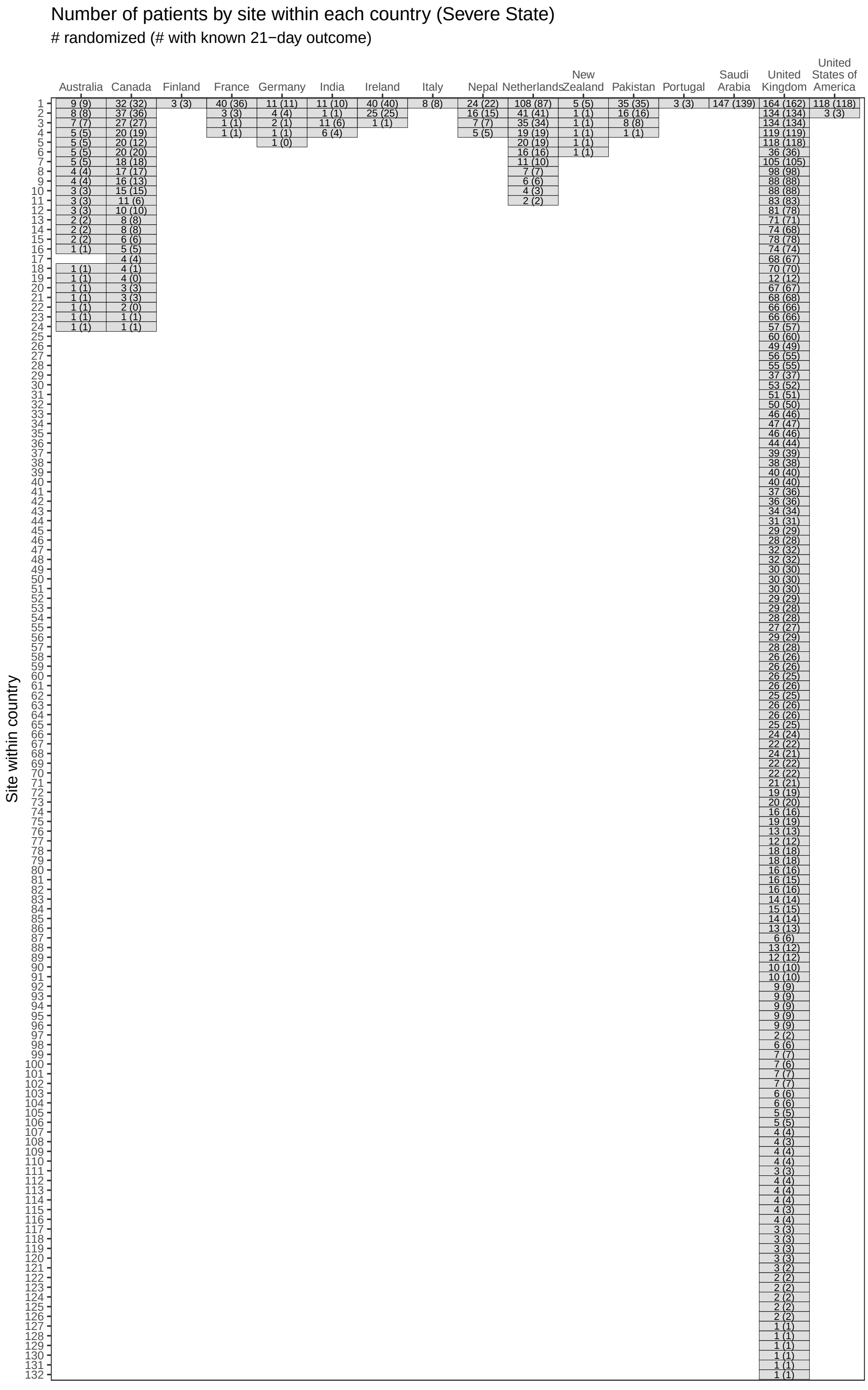

**Figure 4.4:** Sample size at each site within each country (**Severe** State).

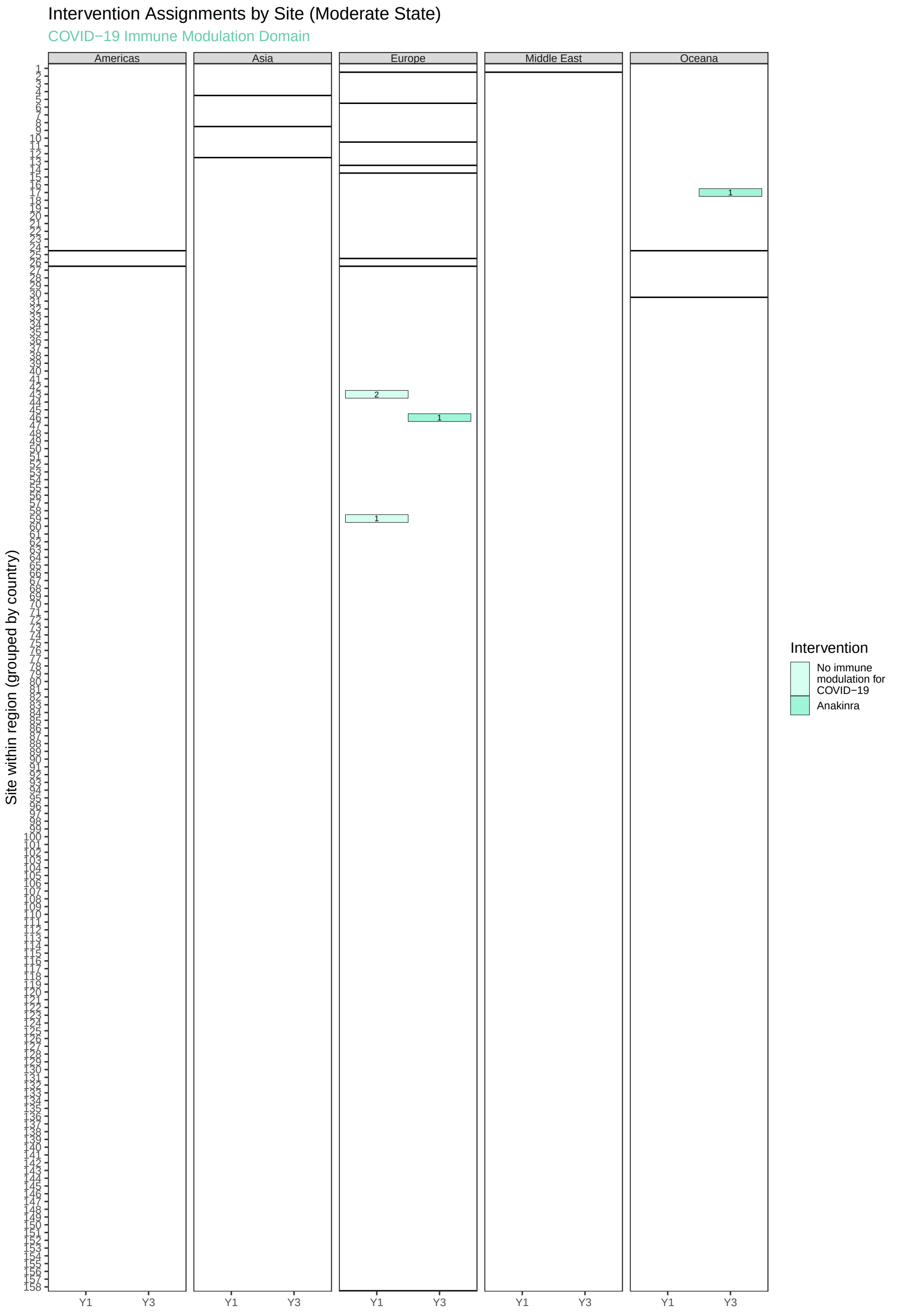

**Figure 4.5:** Allocation of **Moderate State** interventions in the **COVID-19 Immune Modulation** domain by site. The data are summarized in three panels — one for each geographical region. Each panel is a grid with interventions on the x-axis and sites on the y-axis. Each colored cell corresponds to an intervention that has randomized patients at a site. Cells are colored by intervention, with the number in each cell representing how many patients were randomized to the intervention at that site. The solid black horizontal lines distinguish sites located within the same country in the region.

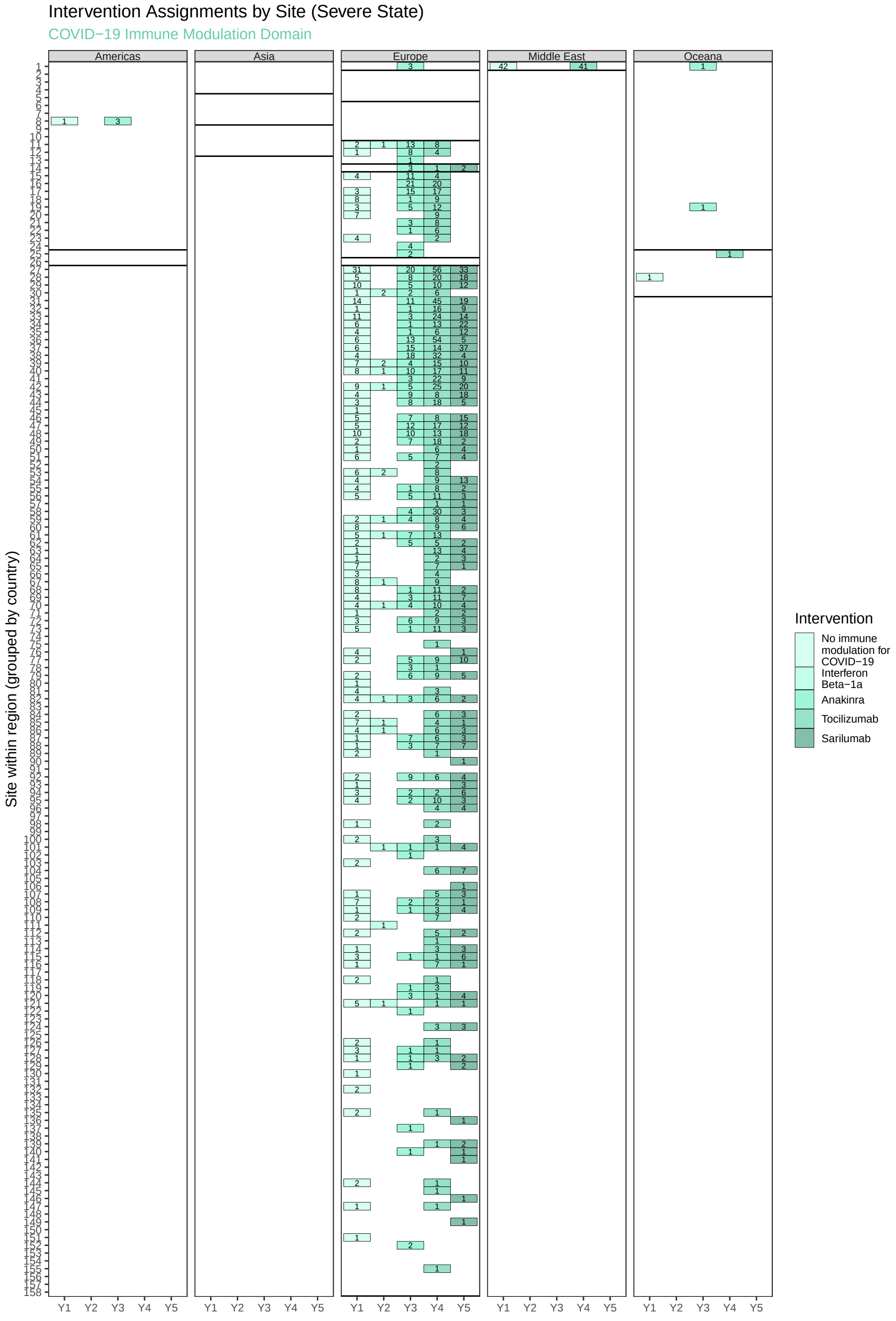

**Figure 4.6:** Allocation of **Severe State** interventions in the **COVID-19 Immune Modulation** domain by site.

### 5 Analysis Conventions

The following conventions were applied to the analyses contained in this report:

- The ITSC closed randomization to the Corticosteroid domain within the PISOP stratum on June 17, 2020. This decision was made following the release of the RECOVERY trial results on June 16, 2020 which reported strong positive effects of dexamethasone. Following this decision, the results from the Corticosteroid domain in REMAP-CAP were publicly disclosed. Patients who are randomized within the PISOP stratum after June 17 receive no randomized assignment within the Corticosteroid domain; however it is asssumed they likely receive fixed duration steroid. Thus, for the statistical model, patients randomized after June 17 are coded identically to patients randomized to fixed duration steroid.
- All sites within a country that have $< 5$ patients randomized in the analysis population within a disease state will have their results combined into a single site within that country.
- For the estimation of time trends in the model, time buckets with $< 5$ patients randomized within the bucket within a disease state were combined with a neighboring bucket.
- All interactions between the shock-based steroid arm and other domains are dropped (assumed to be zero) per the SAP.
- Patients with no randomized assignment in any domain are included in the analysis population when the OSFD outcome is known. Thus these patients contribute to estimates of covariate effects (but no treatment effects) in the model.
- Some patients in the Research Online database have missing randomization assignments for some domains. These patients are treated as having "no assignment" within the respective domains. Only 6 patients are affected.
- Data provided to the SAC only include patients who consented for use of their data.
- Some patients for whom 21 days have elapsed since randomization have missing 21-day OSFD outcomes in the data export. A supplemental file was provided to the SAC in which some additional outcome data was obtained based on a manual review.
- For unique patient identifiers that exist in both the Research Online and Spiral databases, the analysis generally pulls the eligibility and randomization information from the Spiral database and the outcomes from the Research Online database. If outcomes were reported in both places, the reported outcome in Spiral was selected per instructions from the global project manager for the trial (email dated August 6, 2020).
- There are some patients with a missing value for the age covariate. For modeling purposes, these patients are coded as the referent age category, $60 - 69$.

### 6 Model Stability

The Bayesian model was computed in R version 4.0.5 (2021-03-31), using the rstan package version 2.21.0. This package computes the Markov Chain Monte Carlo (MCMC) using the highly efficient Hamiltonian Monte Carlo method. The MCMC used 10 separate chains, with each chain using a burnin of 500 samples, followed by 2000 samples, for a total of 20000 samples. Convergence diagnostics were assessed, and no concerns regarding mixing or convergence were identified. All $\hat{R}$ values were less than 1.05. All model runs used a random number seed of 5062021 for the MCMC initialization.

### 7 Report Production

All analyses in this report are based on the following documents:

- Statistical Analysis Appendix for REMAP-COVID, version 1, dated August 18, 2020;
- Statistical Analysis Plan for the Immune Modulation Domain for Patients with COVID-19 Pandemic Infection Suspected or Proven (PISOP), version 1.1, dated April 15, 2021;
- Current State of the Statistical Model: Pandemic Model Immune Modulation Domain Analysis, version 3.1-IM, dated May 5, 2021;
- Addendum to the Immune Modulation PISOP SAP and Current State Documents, dated May 5, 2021.

Berry Consultants performed the analysis using data received from multiple sources. Table 7.1 shows the file names for the data exports from each database along with the names of supplemental files received by the SAC, and the dates on which each file was received by the SAC.

**Table 7.1:** Summary of data sources.

| File Name | Date Received | Description |
|---|---|---|
| UPMC_SACDataExport_04302021_1109.csv | April 30, 2021 | UPMC data |
| remapcap_spiral_interimexport_2021-05-03_070120_v10.2.3.csv | May 3, 2021 | Spiral data |
| RAR_Unscrambled_RO_20201214_v3.csv | December 14, 2020 | Research Online data |
| missingOSFD_PISOPModerateSevereModeling_forIM1FinalAnalysis2021-05-04_CG.xlsx | May 5, 2021 | Supplemental OSFD outcome data |

All data summaries were completed using the R[1] statistical computing environment R version 3.5.2 (2018-12-20).

[1] R Development Core Team (2005). R: *A language and environment for statistical computing.* R Foundation for Statistical Computing, Vienna, Austria. ISBN 3-900051-07-0. URL http://www.R-project.org.
