## Appendix 3 Secondary analysis for "Effectiveness of Tocilizumab, Sarilumab, and Anakinra for critically ill patients with COVID-19 The REMAP-CAP COVID-19 Immune Modulation Therapy Domain Randomized Clinical Trial"

### REMAP-CAP Unblinded Secondary Analysis Report for the Immune Modulation Domain

Prepared by the ITSC Analysis Committee

June 10, 2021

2.7 Secondary analysis of OSFD for Unblinded ITT population restricted to non-negative COVID-19 20
3.5 Secondary analysis of hospital survival for Unblinded ITT population with pooled IL-6ra effect 31
5.3 Sensitivity analysis of hospital survival for Immune Modulation per protocol population . . 46

**6 Analyses of secondary endpoints 48**

**8 Report production and data sources 95**

### 1 Introduction

This report summarizes the data and the results for the Immune Modulation Therapy domain analyses run by the ITSC analysis committee. The ITSC analysis committee is blinded to all ongoing domains and interventions in REMAP-CAP. The data and analyses in this report are limited to domains/interventions that have been unblinded. All analyses involving blinded randomized assignments, like the primary analysis, are in the primary analysis report created by the unblinded Statistical Analysis Committee (SAC).

#### 1.1 Immune Modulation domain interventions

There are five interventions in the Immune Modulation Therapy Domain. These are:

1. **Control** (No immune modulation for COVID-19)

2. Interferon-beta-1a (IFN-$\beta$1a)
3. Anakinra (interleukin-1 receptor antagonist; IL-1ra)
4. Tocilizumab (IL-6 receptor antagonist; IL-6ra)
5. Sarilumab (IL-6 receptor antagonist; IL-6ra)

These five interventions are mutually exclusive; patients randomized in this domain are assigned to one of the five interventions. As a result of meeting the statistical trigger for equivalence, the two IL-6ra interventions are pooled into a single intervention labeled Pooled IL-6ra in some models/tables/figures.

#### 1.2 Analysis populations

The SAP for the immune modulation domain analysis defines five analysis populations. This report includes results for four of the five analysis populations. The analysis results for the **REMAP-CAP COVID-19 severe and moderate intent-to-treat (ITT)** population are not included in this report since the authors remain blinded to assingments in ongoing domains and interventions. In this report, tables and figures summarize data in the **Immune Modulation specific ITT population**. This report includes data and analyses from the following four analysis populations:

1. The **Unblinded ITT population** is defined as all severe patients randomized in one or more unblinded domains within the pandemic stratum without a prior randomization in the moderate state. The unblinded ITT population consists of 3848 patients randomized in the Immune Modulation Therapy, Corticosteroid, Antiviral, Anticoagulation, or Immunoglobulin domains. There are 31 patients within this population that are missing values of OSFD and in-hospital mortality.

- 2226 patients randomized to the Immune Modulation domain
  - 405 patients randomized to control of which 405 have known OSFD outcome
  - 19 patients randomized to interferon-beta-1a of which 19 have known OSFD outcomes
  - 373 patients randomized to anakinra of which 365 have known OSFD outcomes
  - 947 patients randomized to tocilizumab of which 938 have known OSFD outcomes
  - 482 patients randomized to sarilumab of which 480 have known OSFD outcomes
- 386 patients randomized to the Corticosteroid domain of which 379 have known OSFD outcomes
  - 3339 patients randomized after the closure of the Corticosteroid domain
- 693 patients randomized to the Antiviral domain of which 691 have known OSFD outcomes.
- 1010 patients randomized to the Anticoagulation domain of which 1007 have known OSFD outcomes.
- 1980 patients randomized to the Immunoglobulin domain of which 1973 have known OSFD out-

comes.

2. The **Unblinded non-negative COVID-19 population** is defined as all patients in the Unblinded ITT population after removing those with >1 negative test for COVID-19 and no positive tests. The unblinded ITT population restricted to non-negative COVID-19 consists of 3613 patients. There are 29 patients within this population that are missing values of OSFD and in-hospital mortality.

- 2096 patients randomized to the Immune Modulation domain
  - 364 patients randomized to **control** of which 364 have known OSFD outcome
  - 15 patients randomized to **interferon-beta-1a** of which 15 have known OSFD outcomes
  - 363 patients randomized to **anakinra** of which 355 have known OSFD outcomes
  - 887 patients randomized to **tocilizumab** of which 879 have known OSFD outcomes
  - 467 patients randomized to **sarilumab** of which 465 have known OSFD outcomes
- 316 patients randomized to the Corticosteroid domain of which 310 have known OSFD outcomes
  - 3203 patients randomized after the closure of the Corticosteroid domain
- 614 patients randomized to the Antiviral domain of which 612 have known OSFD outcomes.
- 955 patients randomized to the Anticoagulation domain of which 952 have known OSFD outcomes.
- 1955 patients randomized to the Immunoglobulin domain of which 1948 have known OSFD outcomes.

3. The **Immune Modulation ITT population** consists of patients in the Unblinded ITT population that were randomized to the Immune Modulation domain within the pandemic stratum. The Immune Modulation ITT population consists of 2226 patients. There are 19 patients within this population that are missing values of OSFD and in-hospital mortality.

- 405 patients randomized to **control** of which 405 have known OSFD outcomes
- 19 patients randomized to **interferon-beta-1a** of which 19 have known OSFD outcomes
- 373 patients randomized to **anakinra** of which 365 have known OSFD outcomes
- 947 patients randomized to **tocilizumab** of which 938 have known OSFD outcomes
- 482 patients randomized to **sarilumab** of which 480 have known OSFD outcomes

4. The **Immune Modulation Per Protocol (PP) population** consists of the patients in the Immune Modulation ITT population who have been treated as per protocol. The Immune Modulation per protocol population consists of 2107 patients. There are 13 patients within this population that are missing values of OSFD and in-hospital mortality.

- 394 patients randomized to **control** of which 394 have known OSFD outcomes
- 14 patients randomized to **interferon-beta-1a** of which 14 have known OSFD outcomes
- 349 patients randomized to **anakinra** of which 343 have known OSFD outcomes
- 901 patients randomized to **tocilizumab** of which 896 have known OSFD outcomes
- 449 patients randomized to **sarilumab** of which 447 have known OSFD outcomes

#### 1.3 Modeling conventions

- All reported credible intervals (CrIs) are 95% equal-tailed intervals.
- Results from models of ordinal and dichotomous endpoints are reported as odds ratios (ORs). Results from models of time to event endpoints are reported as hazard ratios (HRs). **For consistency of interpretation, all models are parameterized so that an OR/HR greater than 1 indicates patient benefit relative to the reference group and an OR/HR less than 1 indicates patient harm relative to the reference group.**
- The reference group for the age category OR/HRs is the age category from 60-69 years old.
- The reference group for the time epochs OR/HRs is the most recent time epoch consisting of the four-week period preceding Apr 9, 2021. Time epoch 0 is the most recent epoch and the epochs move backwards in time in two-week periods from epoch 1 to 27.
- The reference group for the sex at birth OR/HRs is the male category.
- **Interferon-beta-1a**, **Anakinra**, **Tocilizumab**, and **Sarilumab** are compared to the **Control** intervention. A posterior probability of at least 99% that the odds ratio is greater than 1 is used as a statistical trigger for efficacy (or superiority to control). Similarly, a probability of harm is reported as the probability that the odds ratio is less than 1 relative to **Control**, or 1 minus the probability of efficacy.
- **Interferon-beta-1a**, **Anakinra**, **Tocilizumab**, and **Sarilumab** are compared to **Control** for futility. A 95% probability of a smaller than 1.2 odds ratio for **Interferon-beta-1a**, **Anakinra**, **Tocilizumab**, and **Sarilumab** relative to **Control** is used as a statistical trigger for futility.
- Pairwise comparisons of **Interferon-beta-1a**, **Anakinra**, **Tocilizumab**, and **Sarilumab** are completed to assess equivalence. A 90% probability of equivalence (defined as an odds ratio of two active interventions between 1/1.2 and 1.2) is used as a statistical trigger for equivalence.
- For each intervention in the Immune Modulation domain, the probability of being in the optimal regimen is calculated and used for statistical triggers of domain superiority and inferiority. A greater than 99% probability of being in the optimal regimen is used as a statistical trigger for superiority for each active Immune Modulation intervention. A less than 0.33% probability of being in the optimal

regimen is used as a statistical trigger of inferiority for all Immune Modulation interventions.

- OR/HR effects for pre-specified **combinations** of interventions from the Immune Modulation domain with the Corticosteroid and Antiviral domain are reported relative to receiving control in each domain. These OR/HRs incorporate the effect of each individual intervention and the interaction term for the combination of interventions.
- OR/HR effects for pre-specified **interaction effects** are reported relative to an additive effect (on the log odds ratio scale). For example, if an interaction effect OR/HR is equal to 1, the combination of interventions is additive (equal to the sum of the log OR effects of the two interventions taken separately). If an interaction OR/HR is greater than 1, the effect of the combination of interventions is synergistic (greater than the sum of the log OR effects of the two interventions taken separately). If the interaction OR/HR is less than 1, the effect of the combination is sub-additive (less than the sum of the log OR effects of the two interventions taken separately).

#### 1.4 Overall summaries of OSFD in the Unblinded ITT population

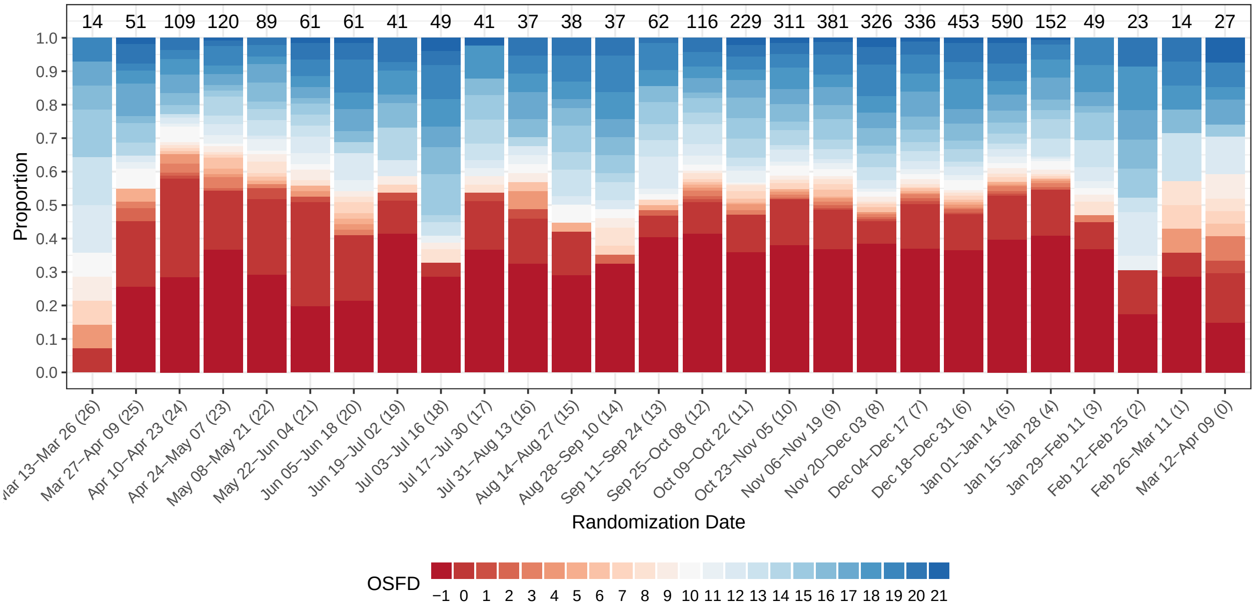

Figure 1: Empirical distribution of organ support free days (OSFD) by time epoch. The Mar 12-Apr 9 category (epoch 0) is the reference time epoch in all models. This plot shows the time epochs defined for the Unblinded ITT population, with the labeled epoch number shown in parentheses.

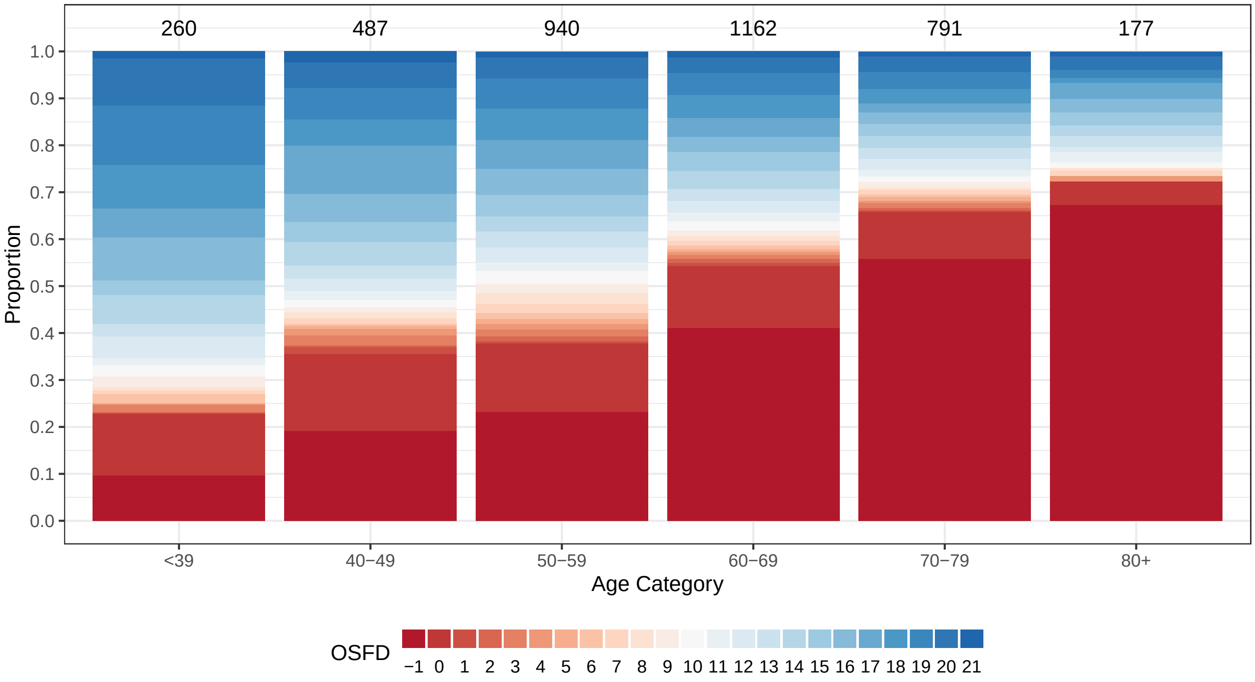

Figure 2: Empirical distribution of organ support free days (OSFD) by age category for the Unblinded ITT population. The 60—69 year age category is the reference group in all models.

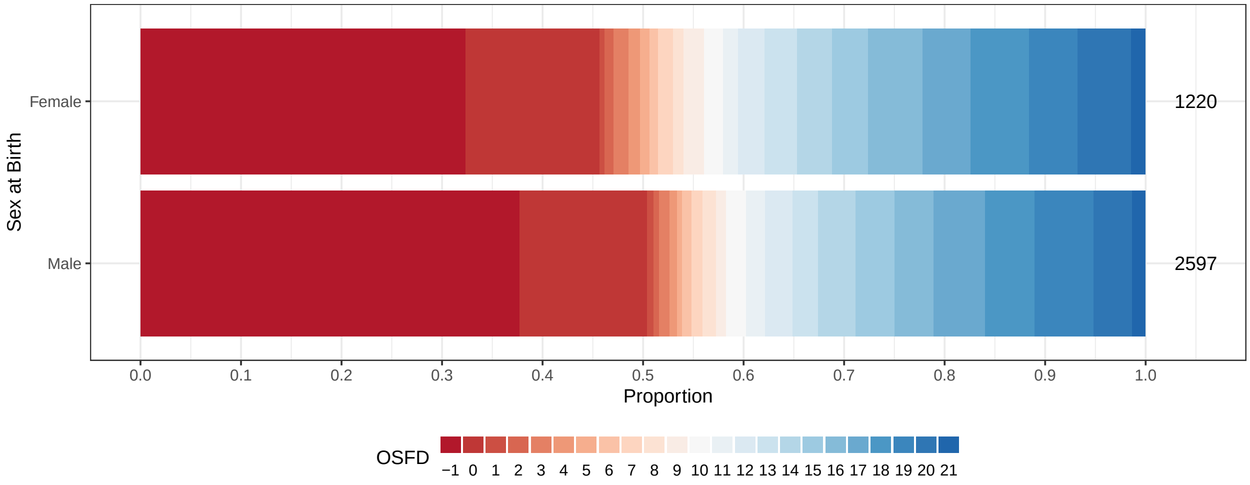

Figure 3: Empirical distribution of organ support free days (OSFD) by sex at birth for the Unblinded ITT population. The male category is the reference group in all models.

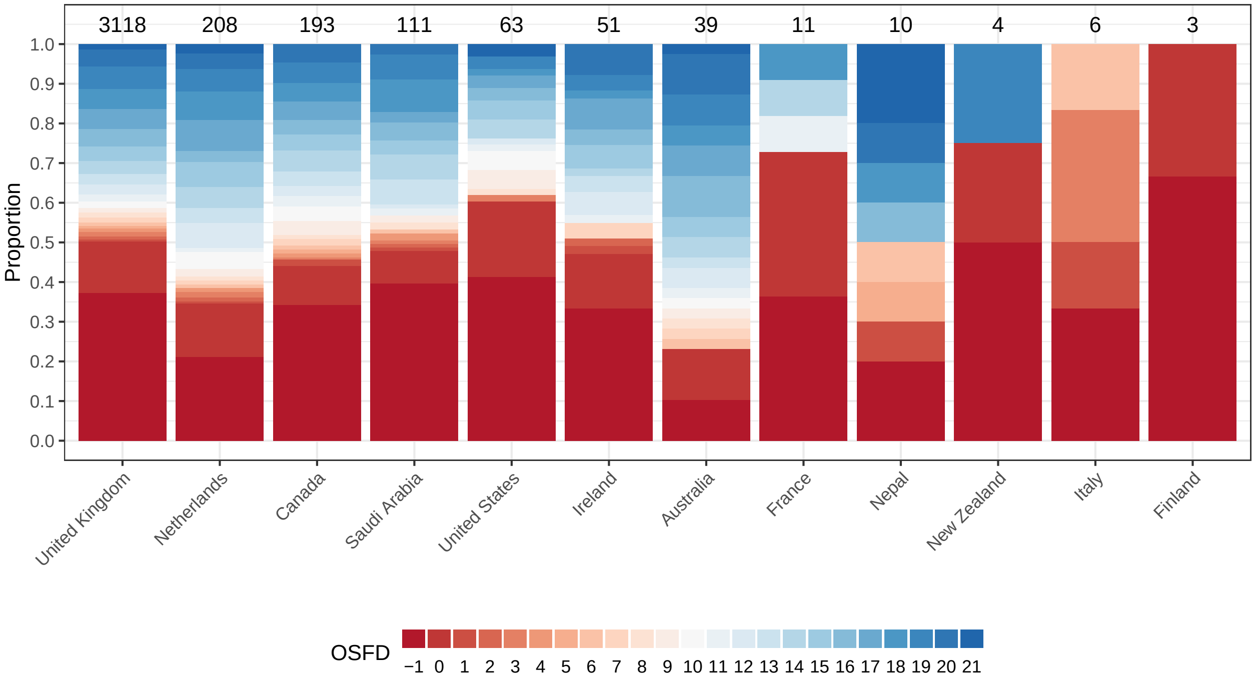

Figure 4: Empirical distribution of organ support free days (OSFD) by country for the Unblinded ITT population.

### 2 Secondary analyses of OSFD

#### 2.1 Empirical distribution of OSFD in Immune Modulation ITT population

Table 1: Summary of OSFD for the Immune Modulation ITT population

| Intervention | # Patients | # Known | In-hospital deaths | OSFD median (IQR) | OSFD in survivors* median (IQR) |
|---|---|---|---|---|---|
| Interferon-beta-1a | 19 | 19 | 7 (36.8%) | 0 (-1, 13.5) | 11 (0, 17.25) |
| Anakinra | 373 | 365 | 145 (39.7%) | 0 (-1, 15) | 14 (3.75, 18) |
| Tocilizumab | 947 | 938 | 317 (33.8%) | 7 (-1, 16) | 15 (7, 18) |
| Sarilumab | 482 | 480 | 157 (32.7%) | 9 (-1, 17) | 15 (9, 18) |
| Control | 405 | 405 | 150 (37%) | 0 (-1, 15) | 13 (4, 17) |
| Pooled IL-6ra | 1429 | 1418 | 474 (33.4%) | 8 (-1, 17) | 15 (8, 18) |

* Days free of organ support in survivors within 21 days

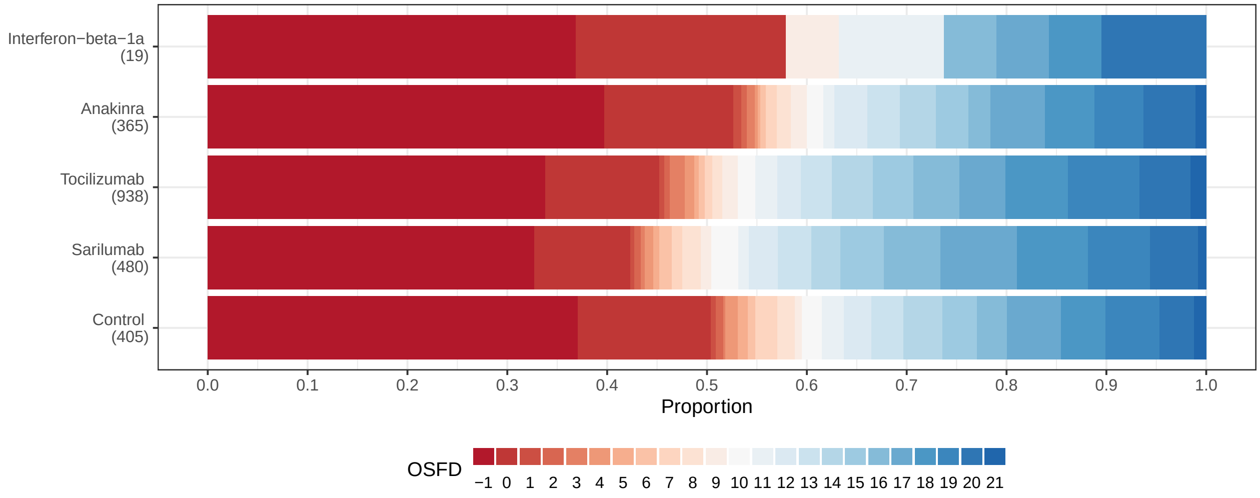

Figure 5: Empirical distribution of organ support free days (OSFD) for interferon-beta-1a, anakinra, tocilizumab, sarilumab, and control. This plot is restricted to the Immune Modulation ITT population.

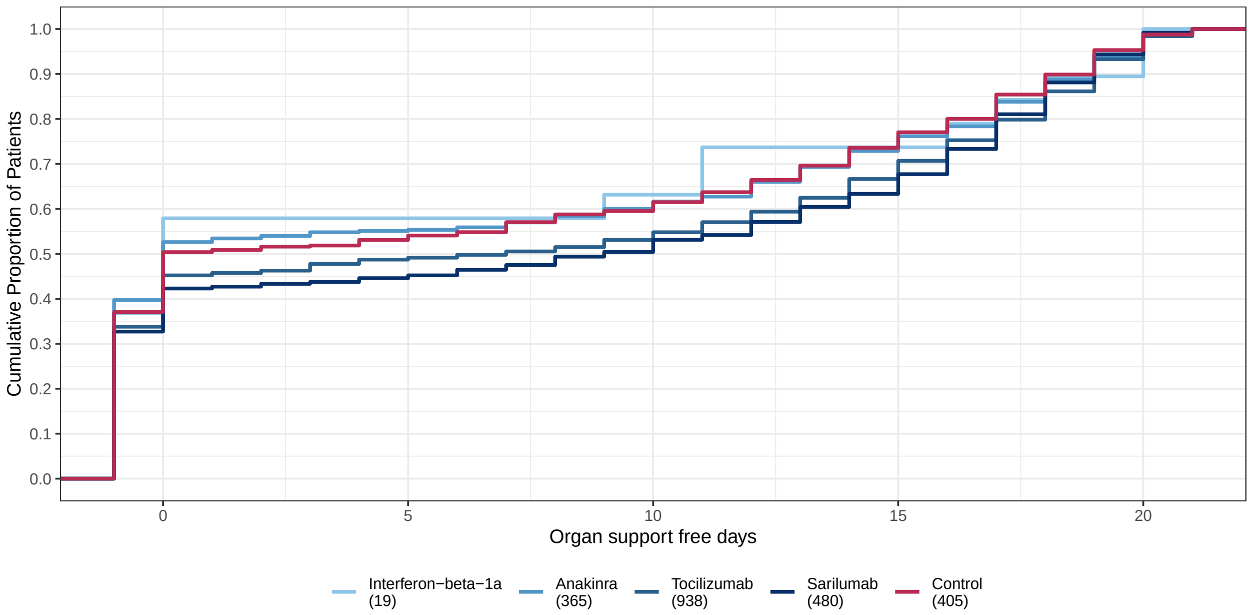

Figure 6: Empirical cumulative distribution of organ support free days (OSFD) for interferon-beta-1a, anakinra, tocilizumab, sarilumab, and control. This plot is restricted to the Immune Modulation ITT population.

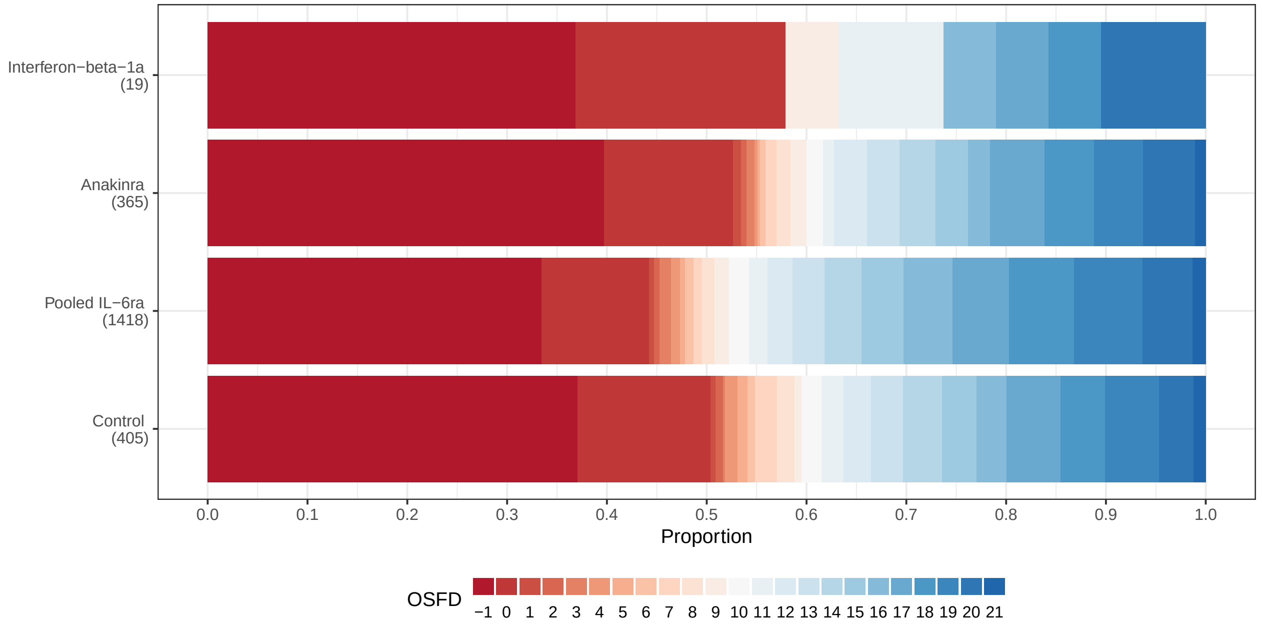

Figure 7: Empirical cumulative distribution of organ support free days (OSFD) for Immune Modulation interventions and control. This plot is restricted to the Immune Modulation ITT population.

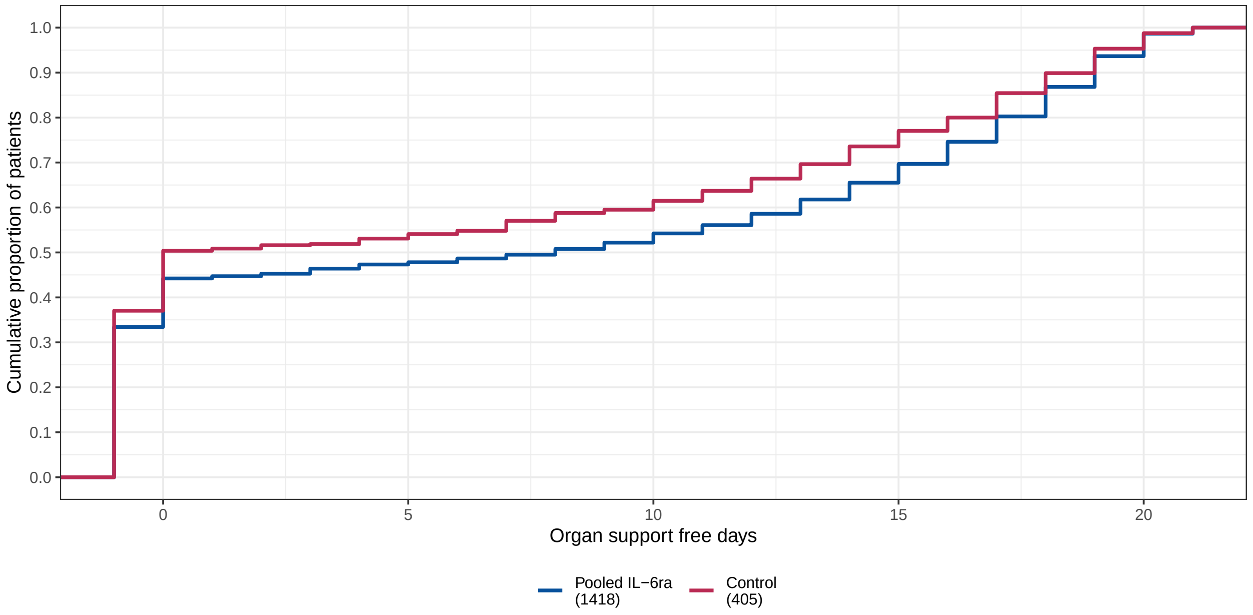

Figure 8: Empirical cumulative distribution of organ support free days (OSFD) for Immune Modulation and control interventions. This plot is restricted to the Immune Modulation ITT population.

#### 2.2 Summary of OSFD odds ratios for IL-6ra interventions

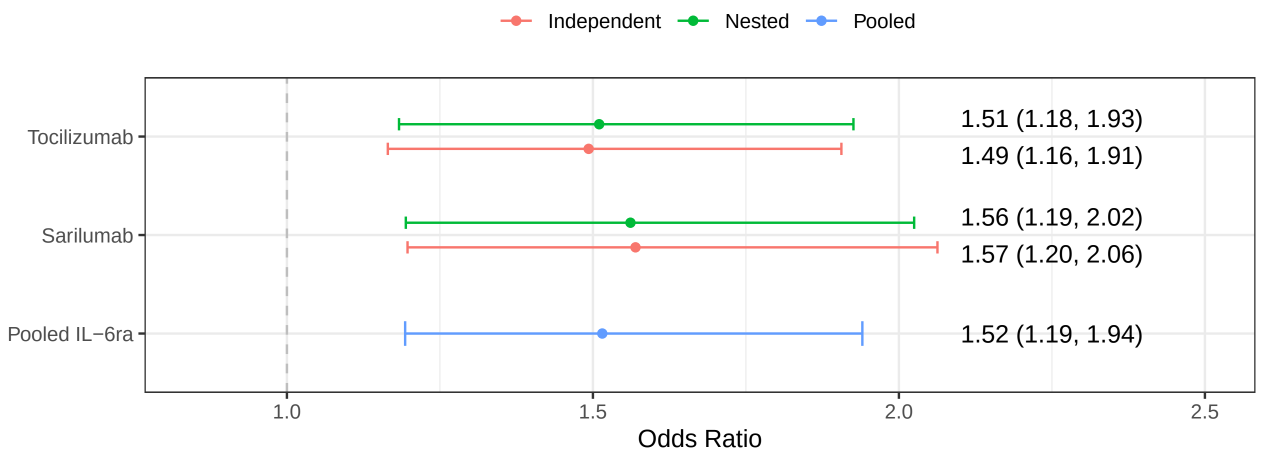

Figure 9: OSFD odds ratios for tocilizumab, sarilumab, and pooled IL-6ra in models with nested, independent, and pooled treatment effects.

#### 2.3 Secondary analysis of OSFD for Unblinded ITT population with nesting between IL-6ra intervention effects

- Model: Primary analysis ordinal model
- Factors: Age, sex, site, time, Immune Modulation Therapy Domain interventions (control, interferon-beta-1a, anakinra, tocilizumab, and sarilumab), COVID-19 Antiviral domain interventions (no antiviral, lopinavir−ritonavir, hydroxychloroquine, combination therapy), Corticosteroid domain interventions (no corticosteroid, fixed-duration corticosteroids, shock-dependent corticosteroids), Immunoglobulin domain interventions (no convalescent plasma, convalescent plasma, delayed convalescent plasma), and Anticoagulation interventions (usual care thromboprophylaxis, therapeutic anticoagulation)
- Treatment effects for tocilizumab and sarilumab are nested with a hierarchical prior

Table 2: Odds ratio parameters for secondary analysis of OSFD for Unblinded ITT population with nested tocilizumab/sarilumab effects

| | Mean | SD | Median | CrI |
|---|---|---|---|---|
| Interferon-beta-1a | 1.37 | 0.56 | 1.27 | (0.58, 2.75) |
| Anakinra | 1.07 | 0.17 | 1.06 | (0.77, 1.44) |
| Tocilizumab | 1.52 | 0.19 | 1.51 | (1.18, 1.93) |
| Sarilumab | 1.57 | 0.21 | 1.56 | (1.19, 2.02) |
| Lopinavir−ritonavir | 0.80 | 0.12 | 0.79 | (0.60, 1.05) |
| Hydroxychloroquine | 0.61 | 0.13 | 0.61 | (0.37, 0.88) |
| Fixed-dose corticosteroids | 1.46 | 0.32 | 1.42 | (0.93, 2.19) |
| Shock-dependent corticosteroids | 1.16 | 0.26 | 1.13 | (0.74, 1.76) |
| Therapeutic anticogulation | 0.86 | 0.10 | 0.86 | (0.68, 1.08) |
| Convalescent plasma | 0.95 | 0.08 | 0.95 | (0.80, 1.11) |
| Delayed convalescent plasma | 0.54 | 0.30 | 0.47 | (0.16, 1.30) |
| Lopinavir−ritonavir*Tocilizumab combination | 1.23 | 0.24 | 1.21 | (0.83, 1.76) |
| Hydroxychloroquine*Tocilizumab combination | 0.93 | 0.24 | 0.91 | (0.54, 1.45) |
| Tocilizumab*Fixed-dose corticosteroids combination | 2.21 | 0.56 | 2.14 | (1.31, 3.49) |
| Lopinavir−ritonavir*Tocilizumab interaction | 1.01 | 0.05 | 1.01 | (0.92, 1.11) |
| Tocilizumab*Fixed-dose corticosteroids interaction | 1.00 | 0.05 | 1.00 | (0.91, 1.10) |
| Hydroxychloroquine*Tocilizumab interaction | 1.00 | 0.05 | 1.00 | (0.91, 1.09) |
| Age<39 | 4.30 | 0.53 | 4.27 | (3.36, 5.46) |
| Age 40-49 | 2.38 | 0.23 | 2.37 | (1.96, 2.85) |
| Age 50-59 | 1.85 | 0.15 | 1.84 | (1.58, 2.15) |
| Age 70-79 | 0.52 | 0.05 | 0.52 | (0.43, 0.62) |
| Age 80+ | 0.31 | 0.05 | 0.30 | (0.21, 0.42) |
| Female | 1.15 | 0.07 | 1.15 | (1.01, 1.30) |

Table 3: Posterior probabilities for secondary analysis of OSFD for Unblinded ITT population with nested tocilizumab/sarilumab effects

| | Posterior Probability |
|---|---|
| Interferon-beta-1a is optimal | 0.283 |
| Interferon-beta-1a is superior to control | 0.728 |
| Interferon-beta-1a is futile (OR < 1.2) | 0.443 |
| Interferon-beta-1a is harmful (OR < 1) | 0.273 |
| Anakinra is optimal | 0.000 |
| Anakinra is superior to control | 0.647 |
| Anakinra is futile (OR < 1.2) | 0.786 |
| Anakinra is harmful (OR < 1) | 0.353 |
| Tocilizumab is optimal | 0.248 |
| Tocilizumab is superior to control | 1.000 |
| Tocilizumab is futile (OR < 1.2) | 0.032 |
| Tocilizumab is harmful (OR < 1) | 0.001 |
| Sarilumab is optimal | 0.469 |
| Sarilumab is superior to control | 0.999 |
| Sarilumab is futile (OR < 1.2) | 0.028 |
| Sarilumab is harmful (OR < 1) | 0.001 |
| Lopinavir−ritonavir*tocilizumab combination OR > 1 | 0.837 |
| Hydroxychloroquine*tocilizumab combination OR > 1 | 0.358 |
| Fixed-dose steroids*tocilizumab combination OR > 1 | 0.998 |
| Interferon-beta-1a equivalent to anakinra | 0.314 |
| Interferon-beta-1a equivalent to tocilizumab | 0.324 |
| Interferon-beta-1a equivalent to sarilumab | 0.307 |
| Anakinra equivalent to tocilizumab | 0.094 |
| Anakinra equivalent to sarilumab | 0.062 |
| Tocilizumab equivalent to sarilumab | 0.929 |
| Anakinra superior to tocilizumab | 0.004 |
| Anakinra superior to sarilumab | 0.002 |
| Tocilizumab superior to sarilumab | 0.361 |

#### 2.4 Secondary analysis of OSFD for Unblinded ITT population with independent IL-6ra intervention effects

- Model: Primary analysis ordinal model
- Factors: Age, sex, site, time, Immune Modulation Therapy Domain interventions (control, interferon-beta-1a, anakinra, tocilizumab, and sarilumab), COVID-19 Antiviral domain interventions (no antiviral, lopinavir−ritonavir, hydroxychloroquine, combination therapy), Corticosteroid domain interventions (no corticosteroid, fixed-duration corticosteroids, shock-dependent corticosteroids), Immunoglobulin

domain interventions (no convalescent plasma, convalescent plasma, delayed convalescent plasma), and Anticoagulation interventions (usual care thromboprophylaxis, therapeutic anticoagulation)

- Treatment effects for tocilizumab and sarilumab are modeled with independent priors.

Table 4: Odds ratio parameters for secondary analysis of OSFD for Unblinded ITT population with independent tocilizumab/sarilumab effects

| | Mean | SD | Median | CrI |
|---|---|---|---|---|
| Interferon-beta-1a | 1.36 | 0.55 | 1.26 | (0.58, 2.65) |
| Anakinra | 1.07 | 0.17 | 1.06 | (0.78, 1.44) |
| Tocilizumab | 1.50 | 0.19 | 1.49 | (1.16, 1.91) |
| Sarilumab | 1.59 | 0.22 | 1.57 | (1.20, 2.06) |
| Lopinavir−ritonavir | 0.80 | 0.12 | 0.79 | (0.59, 1.06) |
| Hydroxychloroquine | 0.61 | 0.14 | 0.61 | (0.37, 0.89) |
| Fixed-dose corticosteroids | 1.46 | 0.32 | 1.42 | (0.93, 2.18) |
| Shock-dependent corticosteroids | 1.16 | 0.26 | 1.13 | (0.73, 1.76) |
| Therapeutic anticogulation | 0.86 | 0.10 | 0.85 | (0.68, 1.07) |
| Convalescent plasma | 0.95 | 0.08 | 0.95 | (0.80, 1.11) |
| Delayed convalescent plasma | 0.54 | 0.32 | 0.47 | (0.16, 1.35) |
| Lopinavir−ritonavir*Tocilizumab combination | 1.21 | 0.24 | 1.19 | (0.80, 1.74) |
| Hydroxychloroquine*Tocilizumab combination | 0.92 | 0.24 | 0.90 | (0.52, 1.45) |
| Tocilizumab*Fixed-dose corticosteroids combination | 2.19 | 0.56 | 2.12 | (1.29, 3.44) |
| Lopinavir−ritonavir*Tocilizumab interaction | 1.01 | 0.05 | 1.01 | (0.92, 1.11) |
| Tocilizumab*Fixed-dose corticosteroids interaction | 1.00 | 0.05 | 1.00 | (0.91, 1.10) |
| Hydroxychloroquine*Tocilizumab interaction | 1.00 | 0.05 | 1.00 | (0.91, 1.10) |
| Age<39 | 4.30 | 0.53 | 4.27 | (3.34, 5.43) |
| Age 40-49 | 2.38 | 0.23 | 2.37 | (1.96, 2.86) |
| Age 50-59 | 1.85 | 0.15 | 1.84 | (1.57, 2.15) |
| Age 70-79 | 0.52 | 0.05 | 0.52 | (0.43, 0.62) |
| Age 80+ | 0.31 | 0.05 | 0.30 | (0.21, 0.42) |
| Female | 1.15 | 0.07 | 1.15 | (1.01, 1.30) |

Table 5: Posterior probabilities for secondary analysis of OSFD for Unblinded ITT population with independent tocilizumab/sarilumab effects

| | Posterior Probability |
|---|---|
| Interferon-beta-1a is optimal | 0.272 |
| Interferon-beta-1a is superior to control | 0.719 |
| Interferon-beta-1a is futile (OR < 1.2) | 0.453 |
| Interferon-beta-1a is harmful (OR < 1) | 0.281 |
| Anakinra is optimal | 0.001 |
| Anakinra is superior to control | 0.646 |
| Anakinra is futile (OR < 1.2) | 0.787 |
| Anakinra is harmful (OR < 1) | 0.354 |
| Tocilizumab is optimal | 0.222 |
| Tocilizumab is superior to control | 0.999 |
| Tocilizumab is futile (OR < 1.2) | 0.043 |
| Tocilizumab is harmful (OR < 1) | 0.001 |
| Sarilumab is optimal | 0.506 |
| Sarilumab is superior to control | 0.999 |
| Sarilumab is futile (OR < 1.2) | 0.026 |
| Sarilumab is harmful (OR < 1) | 0.001 |
| Lopinavir−ritonavir*tocilizumab combination OR > 1 | 0.810 |
| Hydroxychloroquine*tocilizumab combination OR > 1 | 0.342 |
| Fixed-dose steroids*tocilizumab combination OR > 1 | 0.999 |
| Interferon-beta-1a equivalent to anakinra | 0.311 |
| Interferon-beta-1a equivalent to tocilizumab | 0.325 |
| Interferon-beta-1a equivalent to sarilumab | 0.301 |
| Anakinra equivalent to tocilizumab | 0.114 |
| Anakinra equivalent to sarilumab | 0.063 |
| Tocilizumab equivalent to sarilumab | 0.837 |
| Anakinra superior to tocilizumab | 0.005 |
| Anakinra superior to sarilumab | 0.002 |
| Tocilizumab superior to sarilumab | 0.333 |

#### 2.5 Secondary analysis of OSFD for Unblinded ITT population with pooled IL-6ra effect

- Model: Primary analysis ordinal model
- Factors: Age, sex, site, time, Immune Modulation Therapy Domain interventions (control, interferon-beta-1a, anakinra, tocilizumab, and sarilumab), COVID-19 Antiviral domain interventions (no antiviral, lopinavir−ritonavir, hydroxychloroquine, combination therapy), Corticosteroid domain interventions (no corticosteroid, fixed-duration corticosteroids, shock-dependent corticosteroids), Immunoglobulin domain interventions (no convalescent plasma, convalescent plasma, delayed convalescent plasma),

and Anticoagulation interventions (usual care thromboprophylaxis, therapeutic anticoagulation)

- Treatment effects for tocilizumab and sarilumab are pooled into a single IL-6ra effect.

Table 6: Odds ratio parameters for secondary analysis of OSFD for Unblinded ITT population with pooled tocilizumab/sarilumab effects

| | Mean | SD | Median | CrI |
|---|---|---|---|---|
| Interferon-beta-1a | 1.36 | 0.56 | 1.26 | (0.58, 2.69) |
| Anakinra | 1.07 | 0.17 | 1.05 | (0.78, 1.45) |
| Pooled IL-6ra | 1.53 | 0.19 | 1.52 | (1.19, 1.94) |
| Lopinavir−ritonavir | 0.80 | 0.12 | 0.79 | (0.60, 1.05) |
| Hydroxychloroquine | 0.61 | 0.13 | 0.61 | (0.37, 0.89) |
| Fixed-dose corticosteroids | 1.46 | 0.32 | 1.42 | (0.94, 2.16) |
| Shock-dependent corticosteroids | 1.16 | 0.26 | 1.13 | (0.74, 1.74) |
| Therapeutic anticogulation | 0.86 | 0.10 | 0.86 | (0.68, 1.07) |
| Convalescent plasma | 0.95 | 0.08 | 0.94 | (0.80, 1.11) |
| Delayed convalescent plasma | 0.54 | 0.31 | 0.47 | (0.16, 1.34) |
| Lopinavir−ritonavir*Pooled IL-6ra combination | 1.23 | 0.24 | 1.21 | (0.83, 1.77) |
| Hydroxychloroquine*Pooled IL-6ra combination | 0.94 | 0.24 | 0.92 | (0.53, 1.47) |
| Pooled IL-6ra*Fixed-dose corticosteroids combination | 2.22 | 0.56 | 2.16 | (1.32, 3.50) |
| Lopinavir−ritonavir*Pooled IL-6ra interaction | 1.01 | 0.05 | 1.01 | (0.92, 1.11) |
| Pooled IL-6ra*Fixed-dose corticosteroids interaction | 1.00 | 0.05 | 1.00 | (0.91, 1.11) |
| Hydroxychloroquine*Pooled IL-6ra interaction | 1.00 | 0.05 | 1.00 | (0.91, 1.11) |
| Age<39 | 4.31 | 0.52 | 4.29 | (3.38, 5.40) |
| Age 40-49 | 2.38 | 0.23 | 2.37 | (1.97, 2.86) |
| Age 50-59 | 1.85 | 0.15 | 1.84 | (1.58, 2.15) |
| Age 70-79 | 0.52 | 0.05 | 0.52 | (0.43, 0.62) |
| Age 80+ | 0.31 | 0.05 | 0.30 | (0.21, 0.42) |
| Female | 1.15 | 0.07 | 1.15 | (1.02, 1.30) |

Table 7: Posterior probabilities for secondary analysis of OSFD for Unblinded ITT population with pooled tocilizumab/sarilumab effects

| | Posterior Probability |
|---|---|
| Interferon-beta-1a is optimal | 0.317 |
| Interferon-beta-1a is superior to control | 0.724 |
| Interferon-beta-1a is futile (OR < 1.2) | 0.452 |
| Interferon-beta-1a is harmful (OR < 1) | 0.277 |
| Anakinra is optimal | 0.001 |
| Anakinra is superior to control | 0.639 |
| Anakinra is futile (OR < 1.2) | 0.794 |
| Anakinra is harmful (OR < 1) | 0.361 |
| Pooled IL-6ra is optimal | 0.682 |
| Pooled IL-6ra is superior to control | 1.000 |
| Pooled IL-6ra is futile (OR < 1.2) | 0.028 |
| Pooled IL-6ra is harmful (OR < 1) | 0.000 |
| Lopinavir−ritonavir*Pooled IL-6ra combination OR > 1 | 0.837 |
| Hydroxychloroquine*Pooled IL-6ra combination OR > 1 | 0.366 |
| Fixed-dose steroids*Pooled IL-6ra combination OR > 1 | 0.999 |
| Interferon-beta-1a equivalent to anakinra | 0.320 |
| Interferon-beta-1a equivalent to pooled IL-6ra | 0.327 |
| Anakinra equivalent to pooled IL-6ra | 0.083 |
| Anakinra superior to pooled IL-6ra | 0.003 |

#### 2.6 Secondary analysis of OSFD for Unblinded ITT population with additional unblinded interaction effects

- Model: Primary analysis ordinal model
- Factors: Age, sex, site, time, Immune Modulation Therapy Domain interventions (control, interferon-beta-1a, anakinra, tocilizumab, and sarilumab), COVID-19 Antiviral domain interventions (no antiviral, lopinavir−ritonavir, hydroxychloroquine, combination therapy), Corticosteroid domain interventions (no corticosteroid, fixed-duration corticosteroids, shock-dependent corticosteroids), Immunoglobulin domain interventions (no convalescent plasma, convalescent plasma, delayed convalescent plasma), and Anticoagulation interventions (usual care thromboprophylaxis, therapeutic anticoagulation)
- Treatment effects for tocilizumab and sarilumab are pooled into a single IL-6ra effect.

Table 8: Number of patients randomized in the Immune Modulation and Immunoglobulin domains

| | No convalescent plasma | Convalescent plasma |
|---|---|---|
| Interferon-beta-1a | 7 | 4 |
| Anakinra | 39 | 52 |
| Tocilizumab | 208 | 252 |
| Sarilumab | 119 | 143 |
| Control | 95 | 108 |

Table 9: Number of patients randomized in the Immune Modulation and Antiviral domains

| | No antiviral | Lopinavir-ritonavir | HCQ | Combination therapy |
|---|---|---|---|---|
| Interferon-beta-1a | 2 | 5 | 0 | 0 |
| Anakinra | 6 | 11 | 1 | 0 |
| Tocilizumab | 91 | 63 | 10 | 5 |
| Sarilumab | 16 | 10 | 0 | 0 |
| Control | 115 | 86 | 17 | 3 |

Table 10: Number of patients randomized in the Immune Modulation and Anticoagulation domains

| | Venous thromboprophylaxis | Therapeutic dose anticoagulation |
|---|---|---|
| Interferon-beta-1a | 3 | 7 |
| Anakinra | 18 | 22 |
| Tocilizumab | 80 | 90 |
| Sarilumab | 54 | 40 |
| Control | 71 | 77 |

Table 11: Number of patients randomized in the Immune Modulation and Corticosteroid domains. Note that this includes patients randomized after the closure of the steroid domain in the fixed-dose corticosteroids intervention

| | No corticosteroid | Fixed-dose corticosteroids | Shock-dependent corticosteroids |
|---|---|---|---|
| Interferon-beta-1a | 1 | 15 | 1 |
| Anakinra | 0 | 372 | 1 |
| Tocilizumab | 16 | 891 | 19 |
| Sarilumab | 0 | 482 | 0 |
| Control | 9 | 350 | 18 |

Table 12: Odds ratio parameters for secondary analysis of OSFD with additional unblinded interaction effects

| | Mean | SD | Median | CrI |
|---|---|---|---|---|
| Interferon-beta-1a | 1.35 | 0.55 | 1.25 | (0.56, 2.68) |
| Anakinra | 1.08 | 0.19 | 1.07 | (0.76, 1.49) |
| Pooled IL-6ra | 1.45 | 0.40 | 1.39 | (0.82, 2.38) |
| Lopinavir−ritonavir | 0.75 | 0.12 | 0.74 | (0.55, 1.01) |
| Hydroxychloroquine | 0.59 | 0.14 | 0.58 | (0.35, 0.87) |
| Fixed-dose corticosteroids | 1.45 | 0.32 | 1.41 | (0.91, 2.18) |
| Shock-dependent corticosteroids | 1.15 | 0.26 | 1.13 | (0.73, 1.75) |
| Therapeutic anticogulation | 0.88 | 0.11 | 0.87 | (0.68, 1.12) |
| Convalescent plasma | 0.92 | 0.10 | 0.92 | (0.75, 1.12) |
| Delayed convalescent plasma | 0.54 | 0.32 | 0.47 | (0.16, 1.36) |
| Lopinavir−ritonavir*Pooled IL-6ra combination | 1.46 | 0.50 | 1.37 | (0.71, 2.63) |
| Hydroxychloroquine*Pooled IL-6ra combination | 1.03 | 0.50 | 0.93 | (0.38, 2.26) |
| Pooled IL-6ra*Fixed-dose corticosteroids combination | 2.08 | 0.55 | 2.02 | (1.21, 3.31) |
| Pooled IL-6ra*Therapeutic anticoagulation combination | 1.20 | 0.39 | 1.14 | (0.61, 2.16) |
| Pooled IL-6ra*Convalescent plasma combination | 1.50 | 0.45 | 1.44 | (0.81, 2.57) |
| Anakinra*Convalescent plasma combination | 0.78 | 0.24 | 0.75 | (0.42, 1.33) |
| Lopinavir−ritonavir*Pooled IL-6ra interaction | 1.38 | 0.36 | 1.33 | (0.80, 2.20) |
| Pooled IL-6ra*Fixed-dose corticosteroids interaction | 1.31 | 0.66 | 1.17 | (0.46, 2.96) |
| Hydroxychloroquine*Pooled IL-6ra interaction | 1.06 | 0.28 | 1.02 | (0.61, 1.70) |
| Pooled IL-6ra*Therapeutic anticoagulation interaction | 0.96 | 0.19 | 0.94 | (0.64, 1.40) |
| Pooled IL-6ra*Convalescent plasma interaction | 1.14 | 0.17 | 1.12 | (0.84, 1.50) |
| Anakinra*Convalescent plasma interaction | 0.80 | 0.25 | 0.76 | (0.42, 1.38) |
| Age<39 | 4.29 | 0.52 | 4.26 | (3.37, 5.40) |
| Age 40-49 | 2.39 | 0.23 | 2.38 | (1.97, 2.88) |
| Age 50-59 | 1.85 | 0.15 | 1.84 | (1.57, 2.16) |
| Age 70-79 | 0.52 | 0.05 | 0.52 | (0.43, 0.62) |
| Age 80+ | 0.30 | 0.05 | 0.30 | (0.21, 0.42) |
| Female | 1.15 | 0.07 | 1.15 | (1.02, 1.31) |

Table 13: Posterior probabilities for secondary analysis of OSFD for Unblinded ITT population with additional unblinded interaction effects

| | Posterior Probability |
|---|---|
| Interferon-beta-1a is optimal | 0.212 |
| Interferon-beta-1a is superior to control | 0.715 |
| Interferon-beta-1a is futile (OR < 1.2) | 0.458 |
| Interferon-beta-1a is harmful (OR < 1) | 0.285 |
| Anakinra is optimal | 0.006 |
| Anakinra is superior to control | 0.649 |
| Anakinra is futile (OR < 1.2) | 0.749 |
| Anakinra is harmful (OR < 1) | 0.351 |
| Pooled IL-6ra is optimal | 0.782 |
| Pooled IL-6ra is superior to control | 0.889 |
| Pooled IL-6ra is futile (OR < 1.2) | 0.290 |
| Pooled IL-6ra is harmful (OR < 1) | 0.111 |
| Lopinavir−ritonavir*Pooled IL-6ra combination OR > 1 | 0.829 |
| Hydroxychloroquine*Pooled IL-6ra combination OR > 1 | 0.438 |
| Fixed-dose steroids*Pooled IL-6ra combination OR > 1 | 0.996 |
| Therapeutic anticoagulation*Pooled IL-6ra combination OR > 1 | 0.664 |
| Convalescent plasma*Pooled IL-6ra combination OR > 1 | 0.894 |
| Convalescent plasma*Anakinra OR > 1 | 0.161 |
| Interferon-beta-1a equivalent to anakinra | 0.315 |
| Interferon-beta-1a equivalent to pooled IL-6ra | 0.301 |
| Anakinra equivalent to pooled IL-6ra | 0.326 |
| Anakinra superior to pooled IL-6ra | 0.176 |

#### 2.7 Secondary analysis of OSFD for Unblinded ITT population restricted to non-negative COVID-19

- Model: Primary analysis ordinal model
- Factors: Age, sex, site, time, Immune Modulation Therapy Domain interventions (control, interferon-beta-1a, anakinra, tocilizumab, and sarilumab), COVID-19 Antiviral domain interventions (no antiviral, lopinavir−ritonavir, hydroxychloroquine, combination therapy), Corticosteroid domain interventions (no corticosteroid, fixed-duration corticosteroids, shock-dependent corticosteroids), Immunoglobulin domain interventions (no convalescent plasma, convalescent plasma, delayed convalescent plasma), and Anticoagulation interventions (usual care thromboprophylaxis, therapeutic anticoagulation)
- Treatment effects for tocilizumab and sarilumab are modeled with independent priors.

Table 14: Odds ratio parameters for secondary analysis of OSFD for the non-negative population

| | Mean | SD | Median | CrI |
|---|---|---|---|---|
| Interferon-beta-1a | 1.08 | 0.50 | 0.99 | (0.41, 2.31) |
| Anakinra | 1.11 | 0.18 | 1.10 | (0.80, 1.51) |
| Tocilizumab | 1.55 | 0.20 | 1.53 | (1.18, 1.99) |
| Sarilumab | 1.69 | 0.25 | 1.68 | (1.26, 2.23) |
| Lopinavir−ritonavir | 0.87 | 0.14 | 0.86 | (0.63, 1.18) |
| Hydroxychloroquine | 0.58 | 0.15 | 0.57 | (0.33, 0.89) |
| Fixed-dose corticosteroids | 1.35 | 0.32 | 1.32 | (0.83, 2.10) |
| Shock-dependent corticosteroids | 1.02 | 0.25 | 0.99 | (0.62, 1.61) |
| Therapeutic anticogulation | 0.83 | 0.10 | 0.82 | (0.65, 1.05) |
| Convalescent plasma | 0.95 | 0.08 | 0.95 | (0.81, 1.12) |
| Delayed convalescent plasma | 0.59 | 0.34 | 0.51 | (0.17, 1.45) |
| Lopinavir−ritonavir*Tocilizumab combination | 1.36 | 0.29 | 1.33 | (0.88, 2.02) |
| Hydroxychloroquine*Tocilizumab combination | 0.89 | 0.26 | 0.87 | (0.48, 1.46) |
| Tocilizumab*Fixed-dose corticosteroids combination | 2.09 | 0.57 | 2.01 | (1.21, 3.42) |
| Lopinavir−ritonavir*Tocilizumab interaction | 1.01 | 0.05 | 1.01 | (0.92, 1.11) |
| Tocilizumab*Fixed-dose corticosteroids interaction | 1.00 | 0.05 | 1.00 | (0.91, 1.10) |
| Hydroxychloroquine*Tocilizumab interaction | 1.00 | 0.05 | 1.00 | (0.91, 1.10) |
| Age<39 | 4.61 | 0.59 | 4.57 | (3.56, 5.87) |
| Age 40-49 | 2.47 | 0.24 | 2.46 | (2.02, 2.98) |
| Age 50-59 | 1.88 | 0.15 | 1.87 | (1.60, 2.19) |
| Age 70-79 | 0.52 | 0.05 | 0.52 | (0.43, 0.63) |
| Age 80+ | 0.28 | 0.05 | 0.27 | (0.19, 0.39) |
| Female | 1.16 | 0.08 | 1.16 | (1.02, 1.32) |

Table 15: Posterior probabilities for secondary analysis of OSFD for the non-negative population

| | Posterior Probability |
|---|---|
| Interferon-beta-1a is optimal | 0.104 |
| Interferon-beta-1a is superior to control | 0.493 |
| Interferon-beta-1a is futile (OR < 1.2) | 0.670 |
| Interferon-beta-1a is harmful (OR < 1) | 0.507 |
| Anakinra is optimal | 0.000 |
| Anakinra is superior to control | 0.721 |
| Anakinra is futile (OR < 1.2) | 0.712 |
| Anakinra is harmful (OR < 1) | 0.279 |
| Tocilizumab is optimal | 0.202 |
| Tocilizumab is superior to control | 0.999 |
| Tocilizumab is futile (OR < 1.2) | 0.031 |
| Tocilizumab is harmful (OR < 1) | 0.001 |
| Sarilumab is optimal | 0.693 |
| Sarilumab is superior to control | 1.000 |
| Sarilumab is futile (OR < 1.2) | 0.010 |
| Sarilumab is harmful (OR < 1) | 0.000 |
| Lopinavir−ritonavir*tocilizumab combination OR > 1 | 0.913 |
| Hydroxychloroquine*tocilizumab combination OR > 1 | 0.312 |
| Fixed-dose steroids*tocilizumab combination OR > 1 | 0.996 |
| Interferon-beta-1a equivalent to anakinra | 0.310 |
| Interferon-beta-1a equivalent to tocilizumab | 0.203 |
| Interferon-beta-1a equivalent to sarilumab | 0.160 |
| Anakinra equivalent to tocilizumab | 0.128 |
| Anakinra equivalent to sarilumab | 0.042 |
| Tocilizumab equivalent to sarilumab | 0.767 |
| Anakinra superior to tocilizumab | 0.006 |
| Anakinra superior to sarilumab | 0.001 |
| Tocilizumab superior to sarilumab | 0.234 |

#### 2.8 Secondary analysis of OSFD for Immune Modulation ITT population

- Model: Primary analysis ordinal model
- Factors: Age, sex, site, time, Immune Modulation Therapy Domain interventions (control, interferon-beta-1a, anakinra, tocilizumab, and sarilumab)
- Treatment effects for tocilizumab and sarilumab are modeled with independent priors.

Table 16: Odds ratio parameters for secondary analysis of OSFD for Immune Modulation ITT population with independent tocilizumab/sarilumab effects

| | Mean | SD | Median | CrI |
|---|---|---|---|---|
| Interferon-beta-1a | 1.29 | 0.53 | 1.20 | (0.55, 2.59) |
| Anakinra | 1.16 | 0.19 | 1.14 | (0.83, 1.57) |
| Tocilizumab | 1.55 | 0.19 | 1.54 | (1.22, 1.95) |
| Sarilumab | 1.67 | 0.25 | 1.65 | (1.23, 2.22) |
| Age<39 | 4.59 | 0.75 | 4.53 | (3.30, 6.20) |
| Age 40-49 | 2.53 | 0.33 | 2.50 | (1.95, 3.24) |
| Age 50-59 | 2.19 | 0.23 | 2.17 | (1.78, 2.66) |
| Age 70-79 | 0.56 | 0.07 | 0.56 | (0.44, 0.71) |
| Age 80+ | 0.31 | 0.07 | 0.30 | (0.19, 0.47) |
| Female | 1.27 | 0.11 | 1.26 | (1.07, 1.49) |

Table 17: Posterior probabilities for secondary analysis of OSFD for Immune Modulation ITT population with independent tocilizumab/sarilumab effects

| | Posterior Probability |
|---|---|
| Interferon-beta-1a is optimal | 0.195 |
| Interferon-beta-1a is superior to control | 0.683 |
| Interferon-beta-1a is futile (OR < 1.2) | 0.497 |
| Interferon-beta-1a is harmful (OR < 1) | 0.317 |
| Anakinra is optimal | 0.001 |
| Anakinra is superior to control | 0.796 |
| Anakinra is futile (OR < 1.2) | 0.622 |
| Anakinra is harmful (OR < 1) | 0.204 |
| Tocilizumab is optimal | 0.204 |
| Tocilizumab is superior to control | 1.000 |
| Tocilizumab is futile (OR < 1.2) | 0.018 |
| Tocilizumab is harmful (OR < 1) | 0.000 |
| Sarilumab is optimal | 0.600 |
| Sarilumab is superior to control | 1.000 |
| Sarilumab is futile (OR < 1.2) | 0.017 |
| Sarilumab is harmful (OR < 1) | 0.000 |
| Interferon-beta-1a equivalent to anakinra | 0.357 |
| Interferon-beta-1a equivalent to tocilizumab | 0.300 |
| Interferon-beta-1a equivalent to sarilumab | 0.263 |
| Anakinra equivalent to tocilizumab | 0.177 |
| Anakinra equivalent to sarilumab | 0.091 |
| Tocilizumab equivalent to sarilumab | 0.839 |
| Anakinra superior to tocilizumab | 0.008 |
| Anakinra superior to sarilumab | 0.004 |
| Tocilizumab superior to sarilumab | 0.268 |

#### 2.9 Sensitivity analysis of OSFD for Unblinded ITT population with site and time factors removed

- Model: Primary analysis ordinal model
- Factors: Age, sex, Immune Modulation Therapy Domain interventions (control, interferon-beta-1a, anakinra, tocilizumab, and sarilumab), COVID-19 Antiviral domain interventions (no antiviral, lopinavir–ritonavir, hydroxychloroquine, combination therapy), Corticosteroid domain interventions (no corticosteroid, fixed-duration corticosteroids, shock-dependent corticosteroids), Immunoglobulin domain interventions (no convalescent plasma, convalescent plasma, delayed convalescent plasma), and Anticoagulation interventions (usual care thromboprophylaxis, therapeutic anticoag-

ulation) corticosteroids, and shock-dependent steroids and reported interventions of the Immune Modulation Therapy Domain: tocilizumab, sarilumab (combined to a single IL-6 arm) and no immune modulation

- Treatment effects for tocilizumab and sarilumab are modeled with independent priors.

Table 18: Odds ratio parameters for the sensitivity analysis of OSFD with site and time removed

| | Mean | SD | Median | CrI |
|---|---|---|---|---|
| Interferon-beta-1a | 1.12 | 0.46 | 1.03 | (0.48, 2.23) |
| Anakinra | 0.99 | 0.14 | 0.98 | (0.74, 1.30) |
| Tocilizumab | 1.42 | 0.17 | 1.41 | (1.12, 1.79) |
| Sarilumab | 1.39 | 0.18 | 1.37 | (1.07, 1.77) |
| Lopinavir−ritonavir | 0.82 | 0.12 | 0.81 | (0.61, 1.08) |
| Hydroxychloroquine | 0.53 | 0.12 | 0.52 | (0.32, 0.79) |
| Fixed-dose corticosteroids | 1.32 | 0.23 | 1.30 | (0.92, 1.83) |
| Shock-dependent corticosteroids | 1.20 | 0.26 | 1.17 | (0.78, 1.79) |
| Therapeutic anticogulation | 0.89 | 0.10 | 0.88 | (0.70, 1.10) |
| Convalescent plasma | 0.96 | 0.08 | 0.96 | (0.82, 1.12) |
| Delayed convalescent plasma | 0.55 | 0.29 | 0.49 | (0.18, 1.29) |
| Lopinavir−ritonavir*Tocilizumab combination | 1.17 | 0.23 | 1.14 | (0.79, 1.67) |
| Hydroxychloroquine*Tocilizumab combination | 0.75 | 0.20 | 0.73 | (0.43, 1.20) |
| Tocilizumab*Fixed-dose corticosteroids combination | 1.87 | 0.39 | 1.84 | (1.24, 2.77) |
| Lopinavir−ritonavir*Tocilizumab interaction | 1.01 | 0.05 | 1.01 | (0.92, 1.11) |
| Tocilizumab*Fixed-dose corticosteroids interaction | 1.00 | 0.05 | 1.00 | (0.91, 1.11) |
| Hydroxychloroquine*Tocilizumab interaction | 1.00 | 0.05 | 1.00 | (0.91, 1.10) |
| Age<39 | 3.79 | 0.46 | 3.76 | (2.97, 4.76) |
| Age 40-49 | 2.24 | 0.21 | 2.22 | (1.85, 2.68) |
| Age 50-59 | 1.80 | 0.14 | 1.80 | (1.54, 2.09) |
| Age 70-79 | 0.57 | 0.05 | 0.57 | (0.48, 0.68) |
| Age 80+ | 0.37 | 0.06 | 0.36 | (0.26, 0.50) |
| Female | 1.16 | 0.07 | 1.16 | (1.03, 1.31) |

Table 19: Posterior probabilities for the sensitivity analysis of OSFD with site and time removed

| | Posterior Probability |
|---|---|
| Interferon-beta-1a is optimal | 0.198 |
| Interferon-beta-1a is superior to control | 0.536 |
| Interferon-beta-1a is futile (OR < 1.2) | 0.642 |
| Interferon-beta-1a is harmful (OR < 1) | 0.464 |
| Anakinra is optimal | 0.000 |
| Anakinra is superior to control | 0.436 |
| Anakinra is futile (OR < 1.2) | 0.926 |
| Anakinra is harmful (OR < 1) | 0.564 |
| Tocilizumab is optimal | 0.485 |
| Tocilizumab is superior to control | 0.998 |
| Tocilizumab is futile (OR < 1.2) | 0.087 |
| Tocilizumab is harmful (OR < 1) | 0.002 |
| Sarilumab is optimal | 0.317 |
| Sarilumab is superior to control | 0.992 |
| Sarilumab is futile (OR < 1.2) | 0.143 |
| Sarilumab is harmful (OR < 1) | 0.008 |
| Lopinavir–ritonavir*tocilizumab combination OR > 1 | 0.758 |
| Hydroxychloroquine*tocilizumab combination OR > 1 | 0.116 |
| Fixed-dose steroids*tocilizumab combination OR > 1 | 1.000 |
| Interferon-beta-1a equivalent to anakinra | 0.338 |
| Interferon-beta-1a equivalent to tocilizumab | 0.266 |
| Interferon-beta-1a equivalent to sarilumab | 0.283 |
| Anakinra equivalent to tocilizumab | 0.074 |
| Anakinra equivalent to sarilumab | 0.120 |
| Tocilizumab equivalent to sarilumab | 0.892 |
| Anakinra superior to tocilizumab | 0.002 |
| Anakinra superior to sarilumab | 0.004 |
| Tocilizumab superior to sarilumab | 0.593 |

### 3 Secondary analyses of in-hospital mortality

All models in this report are parameterized so that an OR/HR greater than 1 indicates patient benefit relative to the reference group and an OR/HR less than 1 indicates patient harm relative to the reference group. In this section, the ORs are summarized for the outcome of “hospital survival” which is defined as the complement of in-hospital survival.

##### 3.1 Empirical distribution of in-hospital mortality in Immune Modulation ITT population

Table 20: Summary of in-hospital mortality for the Immune Modulation ITT population

| Intervention | # Patients (N) | # Known (n) | Deaths (y) | Mortality Rate (y/n) |
|---|---|---|---|---|
| Interferon-beta-1a | 19 | 19 | 7 | 0.368 |
| Anakinra | 373 | 365 | 145 | 0.397 |
| Tocilizumab | 947 | 938 | 317 | 0.338 |
| Sarilumab | 482 | 480 | 157 | 0.327 |
| Control | 405 | 405 | 150 | 0.370 |
| Pooled IL-6ra | 1429 | 1418 | 474 | 0.334 |

##### 3.2 Summary of hospital survival odds ratios for IL-6ra interventions

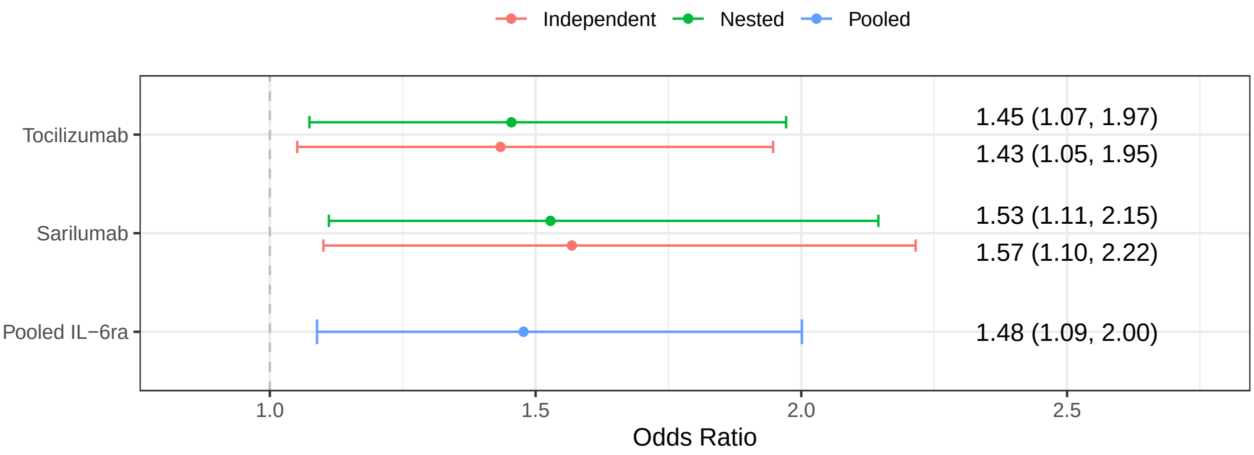

Figure 10: Hospital survival odds ratios for tocilizumab, sarilumab, and pooled IL-6ra in models with nested, independent, and pooled treatment effects.

##### 3.3 Secondary analysis of hospital survival for Unblinded ITT population with nesting between IL-6ra intervention effects

- Model: Primary dichotomous model
- Factors: Age, sex, site, time, Immune Modulation Therapy Domain interventions (control, interferon-beta-1a, anakinra, tocilizumab, and sarilumab), COVID-19 Antiviral domain interventions (no antiviral, lopinavir–ritonavir, hydroxychloroquine, combination therapy), Corticosteroid domain interventions (no corticosteroid, fixed-duration corticosteroids, shock-dependent corticosteroids), Immunoglobulin

domain interventions (no convalescent plasma, convalescent plasma, delayed convalescent plasma), and Anticoagulation interventions (usual care thromboprophylaxis, therapeutic anticoagulation)

- Treatment effects for tocilizumab and sarilumab are nested with a hierarchical prior

Table 21: Odds ratio parameters for secondary analysis of hospital survival for Unblinded ITT population with nested tocilizumab/sarilumab effects

| | Mean | SD | Median | CrI |
|---|---|---|---|---|
| Interferon-beta-1a | 1.42 | 0.72 | 1.26 | (0.52, 3.22) |
| Anakinra | 1.03 | 0.20 | 1.01 | (0.70, 1.49) |
| Tocilizumab | 1.47 | 0.23 | 1.45 | (1.07, 1.97) |
| Sarilumab | 1.55 | 0.27 | 1.53 | (1.11, 2.15) |
| Lopinavir−ritonavir | 0.72 | 0.13 | 0.70 | (0.50, 1.02) |
| Hydroxychloroquine | 0.57 | 0.15 | 0.56 | (0.30, 0.88) |
| Fixed-dose corticosteroids | 1.02 | 0.29 | 0.98 | (0.58, 1.68) |
| Shock-dependent corticosteroids | 1.19 | 0.36 | 1.14 | (0.65, 2.04) |
| Therapeutic anticogulation | 0.83 | 0.12 | 0.82 | (0.62, 1.09) |
| Convalescent plasma | 1.02 | 0.11 | 1.01 | (0.82, 1.25) |
| Delayed convalescent plasma | 0.84 | 0.58 | 0.69 | (0.20, 2.35) |
| Lopinavir−ritonavir*Tocilizumab combination | 1.06 | 0.26 | 1.03 | (0.65, 1.65) |
| Hydroxychloroquine*Tocilizumab combination | 0.84 | 0.27 | 0.81 | (0.40, 1.43) |
| Tocilizumab*Fixed-dose corticosteroids combination | 1.48 | 0.48 | 1.40 | (0.77, 2.62) |
| Lopinavir−ritonavir*Tocilizumab interaction | 1.01 | 0.05 | 1.00 | (0.91, 1.10) |
| Tocilizumab*Fixed-dose corticosteroids interaction | 1.01 | 0.05 | 1.00 | (0.91, 1.11) |
| Hydroxychloroquine*Tocilizumab interaction | 0.99 | 0.05 | 0.99 | (0.90, 1.09) |
| Age<39 | 8.54 | 1.95 | 8.31 | (5.44, 13.07) |
| Age 40-49 | 3.48 | 0.47 | 3.45 | (2.65, 4.52) |
| Age 50-59 | 2.45 | 0.26 | 2.44 | (1.99, 2.99) |
| Age 70-79 | 0.47 | 0.05 | 0.47 | (0.38, 0.57) |
| Age 80+ | 0.26 | 0.05 | 0.26 | (0.18, 0.37) |
| Female | 1.25 | 0.10 | 1.25 | (1.06, 1.47) |

Table 22: Posterior probabilities for secondary analysis of hospital survival for Unblinded ITT population with nested tocilizumab/sarilumab effects

| | Posterior Probability |
|---|---|
| Interferon-beta-1a is optimal | 0.317 |
| Interferon-beta-1a is superior to control | 0.689 |
| Interferon-beta-1a is futile (OR < 1.2) | 0.460 |
| Interferon-beta-1a is harmful (OR < 1) | 0.311 |
| Anakinra is optimal | 0.001 |
| Anakinra is superior to control | 0.524 |
| Anakinra is futile (OR < 1.2) | 0.813 |
| Anakinra is harmful (OR < 1) | 0.476 |
| Tocilizumab is optimal | 0.212 |
| Tocilizumab is superior to control | 0.993 |
| Tocilizumab is futile (OR < 1.2) | 0.103 |
| Tocilizumab is harmful (OR < 1) | 0.007 |
| Sarilumab is optimal | 0.469 |
| Sarilumab is superior to control | 0.996 |
| Sarilumab is futile (OR < 1.2) | 0.072 |
| Sarilumab is harmful (OR < 1) | 0.004 |
| Lopinavir–ritonavir*tocilizumab combination OR > 1 | 0.549 |
| Hydroxychloroquine*tocilizumab combination OR > 1 | 0.250 |
| Fixed-dose steroids*tocilizumab combination OR > 1 | 0.862 |
| Interferon-beta-1a equivalent to anakinra | 0.272 |
| Interferon-beta-1a equivalent to tocilizumab | 0.290 |
| Interferon-beta-1a equivalent to sarilumab | 0.279 |
| Anakinra equivalent to tocilizumab | 0.126 |
| Anakinra equivalent to sarilumab | 0.076 |
| Tocilizumab equivalent to sarilumab | 0.858 |
| Anakinra superior to tocilizumab | 0.011 |
| Anakinra superior to sarilumab | 0.006 |
| Tocilizumab superior to sarilumab | 0.326 |

#### 3.4 Secondary analysis of hospital survival for Unblinded ITT population with independent IL-6ra intervention effects

- Model: Primary dichotomous model
- Factors: Age, sex, site, time, Immune Modulation Therapy Domain interventions (control, interferon-beta-1a, anakinra, tocilizumab, and sarilumab), COVID-19 Antiviral domain interventions (no antiviral, lopinavir–ritonavir, hydroxychloroquine, combination therapy), Corticosteroid domain interventions (no corticosteroid, fixed-duration corticosteroids, shock-dependent corticosteroids), Immunoglobulin

domain interventions (no convalescent plasma, convalescent plasma, delayed convalescent plasma), and Anticoagulation interventions (usual care thromboprophylaxis, therapeutic anticoagulation)

- Treatment effects for tocilizumab and sarilumab are modeled with independent priors.

Table 23: Odds ratio parameters for secondary analysis of hospital survival for Unblinded ITT population with independent tocilizumab/sarilumab effects

| | Mean | SD | Median | CrI |
|---|---|---|---|---|
| Interferon-beta-1a | 1.42 | 0.72 | 1.26 | (0.51, 3.27) |
| Anakinra | 1.03 | 0.20 | 1.01 | (0.70, 1.49) |
| Tocilizumab | 1.45 | 0.23 | 1.43 | (1.05, 1.95) |
| Sarilumab | 1.59 | 0.29 | 1.57 | (1.10, 2.22) |
| Lopinavir−ritonavir | 0.72 | 0.13 | 0.70 | (0.50, 1.02) |
| Hydroxychloroquine | 0.57 | 0.15 | 0.56 | (0.30, 0.89) |
| Fixed-dose corticosteroids | 1.02 | 0.30 | 0.98 | (0.55, 1.69) |
| Shock-dependent corticosteroids | 1.20 | 0.38 | 1.14 | (0.62, 2.08) |
| Therapeutic anticogulation | 0.83 | 0.12 | 0.82 | (0.62, 1.08) |
| Convalescent plasma | 1.02 | 0.11 | 1.01 | (0.83, 1.24) |
| Delayed convalescent plasma | 0.84 | 0.58 | 0.69 | (0.21, 2.41) |
| Lopinavir−ritonavir*Tocilizumab combination | 1.05 | 0.26 | 1.02 | (0.63, 1.64) |
| Hydroxychloroquine*Tocilizumab combination | 0.83 | 0.26 | 0.80 | (0.40, 1.41) |
| Tocilizumab*Fixed-dose corticosteroids combination | 1.47 | 0.49 | 1.40 | (0.73, 2.62) |
| Lopinavir−ritonavir*Tocilizumab interaction | 1.01 | 0.05 | 1.00 | (0.91, 1.10) |
| Tocilizumab*Fixed-dose corticosteroids interaction | 1.01 | 0.05 | 1.00 | (0.91, 1.11) |
| Hydroxychloroquine*Tocilizumab interaction | 0.99 | 0.05 | 0.99 | (0.90, 1.09) |
| Age<39 | 8.56 | 1.94 | 8.33 | (5.49, 13.02) |
| Age 40-49 | 3.49 | 0.48 | 3.46 | (2.67, 4.53) |
| Age 50-59 | 2.46 | 0.25 | 2.45 | (1.99, 2.98) |
| Age 70-79 | 0.47 | 0.05 | 0.47 | (0.38, 0.57) |
| Age 80+ | 0.26 | 0.05 | 0.26 | (0.18, 0.37) |
| Female | 1.25 | 0.11 | 1.25 | (1.06, 1.47) |

Table 24: Posterior probabilities for secondary analysis of hospital survival for Unblinded ITT population with independent tocilizumab/sarilumab effects

| | Posterior Probability |
|---|---|
| Interferon-beta-1a is optimal | 0.306 |
| Interferon-beta-1a is superior to control | 0.691 |
| Interferon-beta-1a is futile (OR < 1.2) | 0.456 |
| Interferon-beta-1a is harmful (OR < 1) | 0.309 |
| Anakinra is optimal | 0.001 |
| Anakinra is superior to control | 0.529 |
| Anakinra is futile (OR < 1.2) | 0.809 |
| Anakinra is harmful (OR < 1) | 0.470 |
| Tocilizumab is optimal | 0.176 |
| Tocilizumab is superior to control | 0.990 |
| Tocilizumab is futile (OR < 1.2) | 0.131 |
| Tocilizumab is harmful (OR < 1) | 0.011 |
| Sarilumab is optimal | 0.516 |
| Sarilumab is superior to control | 0.993 |
| Sarilumab is futile (OR < 1.2) | 0.069 |
| Sarilumab is harmful (OR < 1) | 0.007 |
| Lopinavir–ritonavir*tocilizumab combination OR > 1 | 0.523 |
| Hydroxychloroquine*tocilizumab combination OR > 1 | 0.232 |
| Fixed-dose steroids*tocilizumab combination OR > 1 | 0.850 |
| Interferon-beta-1a equivalent to anakinra | 0.263 |
| Interferon-beta-1a equivalent to tocilizumab | 0.286 |
| Interferon-beta-1a equivalent to sarilumab | 0.271 |
| Anakinra equivalent to tocilizumab | 0.148 |
| Anakinra equivalent to sarilumab | 0.069 |
| Tocilizumab equivalent to sarilumab | 0.701 |
| Anakinra superior to tocilizumab | 0.015 |
| Anakinra superior to sarilumab | 0.005 |
| Tocilizumab superior to sarilumab | 0.272 |

#### 3.5 Secondary analysis of hospital survival for Unblinded ITT population with pooled IL-6ra effect

- Model: Primary dichotomous model
- Factors: Age, sex, site, time, Immune Modulation Therapy Domain interventions (control, interferon-beta-1a, anakinra, tocilizumab, and sarilumab), COVID-19 Antiviral domain interventions (no antiviral, lopinavir–ritonavir, hydroxychloroquine, combination therapy), Corticosteroid domain interventions (no corticosteroid, fixed-duration corticosteroids, shock-dependent corticosteroids), Immunoglobulin

domain interventions (no convalescent plasma, convalescent plasma, delayed convalescent plasma), and Anticoagulation interventions (usual care thromboprophylaxis, therapeutic anticoagulation)

- Treatment effects for tocilizumab and sarilumab are pooled into a single IL-6ra effect.

Table 25: Odds ratio parameters for secondary analysis of hospital survival for Unblinded ITT population with pooled tocilizumab/sarilumab effects

| | Mean | SD | Median | CrI |
|---|---|---|---|---|
| Interferon-beta-1a | 1.42 | 0.72 | 1.27 | (0.51, 3.25) |
| Anakinra | 1.03 | 0.20 | 1.01 | (0.69, 1.47) |
| Pooled IL-6ra | 1.49 | 0.23 | 1.48 | (1.09, 2.00) |
| Lopinavir−ritonavir | 0.72 | 0.13 | 0.70 | (0.50, 1.01) |
| Hydroxychloroquine | 0.56 | 0.15 | 0.55 | (0.30, 0.89) |
| Fixed-dose corticosteroids | 1.03 | 0.30 | 0.98 | (0.57, 1.72) |
| Shock-dependent corticosteroids | 1.20 | 0.38 | 1.15 | (0.64, 2.10) |
| Therapeutic anticogulation | 0.83 | 0.12 | 0.82 | (0.61, 1.09) |
| Convalescent plasma | 1.02 | 0.11 | 1.01 | (0.83, 1.24) |
| Delayed convalescent plasma | 0.84 | 0.58 | 0.69 | (0.20, 2.32) |
| Lopinavir−ritonavir*Pooled IL-6ra combination | 1.08 | 0.27 | 1.04 | (0.66, 1.68) |
| Hydroxychloroquine*Pooled IL-6ra combination | 0.85 | 0.27 | 0.82 | (0.42, 1.44) |
| Pooled IL-6ra*Fixed-dose corticosteroids combination | 1.52 | 0.51 | 1.44 | (0.76, 2.74) |
| Lopinavir−ritonavir*Pooled IL-6ra interaction | 1.01 | 0.05 | 1.00 | (0.91, 1.11) |
| Pooled IL-6ra*Fixed-dose corticosteroids interaction | 1.01 | 0.05 | 1.00 | (0.91, 1.11) |
| Hydroxychloroquine*Pooled IL-6ra interaction | 0.99 | 0.05 | 0.99 | (0.90, 1.09) |
| Age<39 | 8.53 | 1.95 | 8.29 | (5.47, 12.98) |
| Age 40-49 | 3.49 | 0.48 | 3.46 | (2.64, 4.53) |
| Age 50-59 | 2.45 | 0.25 | 2.44 | (2.00, 2.97) |
| Age 70-79 | 0.47 | 0.05 | 0.47 | (0.38, 0.57) |
| Age 80+ | 0.27 | 0.05 | 0.26 | (0.18, 0.37) |
| Female | 1.25 | 0.10 | 1.25 | (1.07, 1.47) |

Table 26: Posterior probabilities for secondary analysis of hospital survival for Unblinded ITT population with pooled tocilizumab/sarilumab effects

| | Posterior Probability |
|---|---|
| Interferon-beta-1a is optimal | 0.377 |
| Interferon-beta-1a is superior to control | 0.692 |
| Interferon-beta-1a is futile (OR < 1.2) | 0.452 |
| Interferon-beta-1a is harmful (OR < 1) | 0.308 |
| Anakinra is optimal | 0.003 |
| Anakinra is superior to control | 0.516 |
| Anakinra is futile (OR < 1.2) | 0.821 |
| Anakinra is harmful (OR < 1) | 0.484 |
| Pooled IL-6ra is optimal | 0.618 |
| Pooled IL-6ra is superior to control | 0.994 |
| Pooled IL-6ra is futile (OR < 1.2) | 0.089 |
| Pooled IL-6ra is harmful (OR < 1) | 0.006 |
| Lopinavir−ritonavir*Pooled IL-6ra combination OR > 1 | 0.571 |
| Hydroxychloroquine*Pooled IL-6ra combination OR > 1 | 0.257 |
| Fixed-dose steroids*Pooled IL-6ra combination OR > 1 | 0.874 |
| Interferon-beta-1a equivalent to anakinra | 0.267 |
| Interferon-beta-1a equivalent to pooled IL-6ra | 0.277 |
| Anakinra equivalent to Pooled IL-6ra | 0.092 |
| Anakinra superior to Pooled IL-6ra | 0.006 |

#### 3.6 Secondary analysis of hospital survival for Unblinded ITT population with additional unblinded interaction effects

- Model: Primary dichotomous model
- Factors: Age, sex, site, time, Immune Modulation Therapy Domain interventions (control, interferon-beta-1a, anakinra, tocilizumab, and sarilumab), COVID-19 Antiviral domain interventions (no antiviral, lopinavir−ritonavir, hydroxychloroquine, combination therapy), Corticosteroid domain interventions (no corticosteroid, fixed-duration corticosteroids, shock-dependent corticosteroids), Immunoglobulin domain interventions (no convalescent plasma, convalescent plasma, delayed convalescent plasma), and Anticoagulation interventions (usual care thromboprophylaxis, therapeutic anticoagulation)
- Treatment effects for tocilizumab and sarilumab are pooled into a single IL-6ra effect.

Table 27: Odds ratio parameters for secondary analysis of hospital survival with additional unblinded interaction effects

| | Mean | SD | Median | CrI |
|---|---|---|---|---|
| Interferon-beta-1a | 1.41 | 0.72 | 1.26 | (0.50, 3.26) |
| Anakinra | 1.05 | 0.23 | 1.02 | (0.68, 1.56) |
| Pooled IL-6ra | 2.11 | 0.90 | 1.93 | (0.90, 4.35) |
| Lopinavir−ritonavir | 0.70 | 0.14 | 0.68 | (0.47, 1.03) |
| Hydroxychloroquine | 0.53 | 0.15 | 0.52 | (0.26, 0.85) |
| Fixed-dose corticosteroids | 1.09 | 0.32 | 1.04 | (0.60, 1.83) |
| Shock-dependent corticosteroids | 1.21 | 0.37 | 1.15 | (0.64, 2.09) |
| Therapeutic anticogulation | 0.84 | 0.13 | 0.83 | (0.61, 1.13) |
| Convalescent plasma | 0.97 | 0.12 | 0.96 | (0.75, 1.23) |
| Delayed convalescent plasma | 0.83 | 0.55 | 0.70 | (0.21, 2.26) |
| Lopinavir−ritonavir*Pooled IL-6ra combination | 1.71 | 0.87 | 1.53 | (0.61, 3.83) |
| Hydroxychloroquine*Pooled IL-6ra combination | 2.19 | 1.82 | 1.68 | (0.45, 6.99) |
| Pooled IL-6ra*Fixed-dose corticosteroids combination | 1.45 | 0.49 | 1.37 | (0.73, 2.64) |
| Pooled IL-6ra*Therapeutic anticoagulation combination | 1.72 | 0.84 | 1.55 | (0.64, 3.81) |
| Pooled IL-6ra*Convalescent plasma combination | 2.49 | 1.12 | 2.28 | (0.99, 5.31) |
| Anakinra*Convalescent plasma combination | 0.73 | 0.24 | 0.69 | (0.36, 1.30) |
| Lopinavir−ritonavir*Pooled IL-6ra interaction | 1.21 | 0.39 | 1.16 | (0.62, 2.12) |
| Pooled IL-6ra*Fixed-dose corticosteroids interaction | 2.19 | 1.68 | 1.73 | (0.49, 6.61) |
| Hydroxychloroquine*Pooled IL-6ra interaction | 0.73 | 0.30 | 0.68 | (0.31, 1.45) |
| Pooled IL-6ra*Therapeutic anticoagulation interaction | 1.00 | 0.26 | 0.96 | (0.59, 1.60) |
| Pooled IL-6ra*Convalescent plasma interaction | 1.23 | 0.23 | 1.21 | (0.85, 1.74) |
| Anakinra*Convalescent plasma interaction | 0.74 | 0.26 | 0.70 | (0.36, 1.39) |
| Age<39 | 8.57 | 1.96 | 8.32 | (5.47, 13.09) |
| Age 40-49 | 3.52 | 0.48 | 3.49 | (2.68, 4.55) |
| Age 50-59 | 2.47 | 0.26 | 2.45 | (2.01, 3.01) |
| Age 70-79 | 0.47 | 0.05 | 0.47 | (0.38, 0.57) |
| Age 80+ | 0.27 | 0.05 | 0.26 | (0.18, 0.37) |
| Female | 1.25 | 0.10 | 1.25 | (1.06, 1.47) |

Table 28: Posterior probabilities for secondary analysis of hospital survival for Unblinded ITT population with additional unblinded interaction effects

| | Posterior Probability |
|---|---|
| Interferon-beta-1a is optimal | 0.109 |
| Interferon-beta-1a is superior to control | 0.686 |
| Interferon-beta-1a is futile (OR < 1.2) | 0.460 |
| Interferon-beta-1a is harmful (OR < 1) | 0.314 |
| Anakinra is optimal | 0.003 |
| Anakinra is superior to control | 0.539 |
| Anakinra is futile (OR < 1.2) | 0.772 |
| Anakinra is harmful (OR < 1) | 0.461 |
| Pooled IL-6ra is optimal | 0.886 |
| Pooled IL-6ra is superior to control | 0.954 |
| Pooled IL-6ra is futile (OR < 1.2) | 0.113 |
| Pooled IL-6ra is harmful (OR < 1) | 0.046 |
| Lopinavir−ritonavir*Pooled IL-6ra combination OR > 1 | 0.820 |
| Hydroxychloroquine*Pooled IL-6ra combination OR > 1 | 0.780 |
| Fixed-dose steroids*Pooled IL-6ra combination OR > 1 | 0.842 |
| Therapeutic anticoagulation*Pooled IL-6ra combination OR > 1 | 0.830 |
| Convalescent plasma*Pooled IL-6ra combination OR > 1 | 0.974 |
| Convalescent plasma*Anakinra OR > 1 | 0.127 |
| Interferon-beta-1a equivalent to anakinra | 0.266 |
| Interferon-beta-1a equivalent to pooled IL-6ra | 0.185 |
| Anakinra equivalent to pooled IL-6ra | 0.114 |
| Anakinra superior to pooled IL-6ra | 0.061 |

#### 3.7 Secondary analysis of hospital survival for Unblinded ITT population restricted to non-negative COVID-19

- Model: Primary dichotomous model
- Factors: Age, sex, site, time, Immune Modulation Therapy Domain interventions (control, interferon-beta-1a, anakinra, tocilizumab, and sarilumab), COVID-19 Antiviral domain interventions (no antiviral, lopinavir−ritonavir, hydroxychloroquine, combination therapy), Corticosteroid domain interventions (no corticosteroid, fixed-duration corticosteroids, shock-dependent corticosteroids), Immunoglobulin domain interventions (no convalescent plasma, convalescent plasma, delayed convalescent plasma), and Anticoagulation interventions (usual care thromboprophylaxis, therapeutic anticoagulation)
- Treatment effects for tocilizumab and sarilumab are modeled with independent priors.

Table 29: Odds ratio parameters for secondary analysis of hospital survival for the non-negative population

| | Mean | SD | Median | CrI |
|---|---|---|---|---|
| Interferon-beta-1a | 1.04 | 0.58 | 0.90 | (0.33, 2.51) |
| Anakinra | 1.06 | 0.21 | 1.04 | (0.70, 1.52) |
| Tocilizumab | 1.48 | 0.24 | 1.46 | (1.06, 2.00) |
| Sarilumab | 1.68 | 0.31 | 1.65 | (1.15, 2.37) |
| Lopinavir−ritonavir | 0.75 | 0.15 | 0.73 | (0.50, 1.10) |
| Hydroxychloroquine | 0.49 | 0.16 | 0.48 | (0.23, 0.82) |
| Fixed-dose corticosteroids | 0.94 | 0.29 | 0.90 | (0.48, 1.62) |
| Shock-dependent corticosteroids | 1.07 | 0.36 | 1.01 | (0.53, 1.92) |
| Therapeutic anticogulation | 0.81 | 0.12 | 0.80 | (0.60, 1.07) |
| Convalescent plasma | 1.02 | 0.11 | 1.01 | (0.83, 1.24) |
| Delayed convalescent plasma | 0.92 | 0.65 | 0.76 | (0.22, 2.57) |
| Lopinavir−ritonavir*Tocilizumab combination | 1.12 | 0.30 | 1.08 | (0.65, 1.82) |
| Hydroxychloroquine*Tocilizumab combination | 0.73 | 0.27 | 0.69 | (0.32, 1.34) |
| Tocilizumab*Fixed-dose corticosteroids combination | 1.37 | 0.49 | 1.29 | (0.65, 2.54) |
| Lopinavir−ritonavir*Tocilizumab interaction | 1.01 | 0.05 | 1.00 | (0.91, 1.11) |
| Tocilizumab*Fixed-dose corticosteroids interaction | 1.01 | 0.05 | 1.00 | (0.91, 1.11) |
| Hydroxychloroquine*Tocilizumab interaction | 0.99 | 0.05 | 0.99 | (0.90, 1.09) |
| Age<39 | 8.80 | 2.05 | 8.51 | (5.60, 13.47) |
| Age 40-49 | 3.54 | 0.50 | 3.50 | (2.67, 4.61) |
| Age 50-59 | 2.44 | 0.26 | 2.42 | (1.97, 2.98) |
| Age 70-79 | 0.48 | 0.05 | 0.48 | (0.39, 0.59) |
| Age 80+ | 0.26 | 0.05 | 0.26 | (0.17, 0.37) |
| Female | 1.26 | 0.11 | 1.25 | (1.06, 1.48) |

Table 30: Posterior probabilities for secondary analysis of hospital survival for the non-negative population

| | Posterior Probability |
|---|---|
| Interferon-beta-1a is optimal | 0.116 |
| Interferon-beta-1a is superior to control | 0.421 |
| Interferon-beta-1a is futile (OR < 1.2) | 0.712 |
| Interferon-beta-1a is harmful (OR < 1) | 0.579 |
| Anakinra is optimal | 0.001 |
| Anakinra is superior to control | 0.576 |
| Anakinra is futile (OR < 1.2) | 0.780 |
| Anakinra is harmful (OR < 1) | 0.424 |
| Tocilizumab is optimal | 0.169 |
| Tocilizumab is superior to control | 0.990 |
| Tocilizumab is futile (OR < 1.2) | 0.110 |
| Tocilizumab is harmful (OR < 1) | 0.010 |
| Sarilumab is optimal | 0.713 |
| Sarilumab is superior to control | 0.998 |
| Sarilumab is futile (OR < 1.2) | 0.040 |
| Sarilumab is harmful (OR < 1) | 0.002 |
| Lopinavir−ritonavir*tocilizumab combination OR > 1 | 0.616 |
| Hydroxychloroquine*tocilizumab combination OR > 1 | 0.150 |
| Fixed-dose steroids*tocilizumab combination OR > 1 | 0.767 |
| Interferon-beta-1a equivalent to anakinra | 0.268 |
| Interferon-beta-1a equivalent to tocilizumab | 0.183 |
| Interferon-beta-1a equivalent to sarilumab | 0.141 |
| Anakinra equivalent to tocilizumab | 0.156 |
| Anakinra equivalent to sarilumab | 0.045 |
| Tocilizumab equivalent to sarilumab | 0.630 |
| Anakinra superior to tocilizumab | 0.018 |
| Anakinra superior to sarilumab | 0.003 |
| Tocilizumab superior to sarilumab | 0.201 |

#### 3.8 Secondary analysis of hospital survival for Immune Modulation ITT population

- Model: Primary dichotomous model
- Factors: Age, sex, site, time, Immune Modulation Therapy Domain interventions (control, interferon-beta-1a, anakinra, tocilizumab, and sarilumab)
- Treatment effects for tocilizumab and sarilumab are modeled with independent priors.

Table 31: Odds ratio parameters for secondary analysis of hospital survival for Immune Modulation ITT population with independent tocilizumab/sarilumab effects

| | Mean | SD | Median | CrI |
|---|---|---|---|---|
| Interferon-beta-1a | 1.42 | 0.71 | 1.27 | (0.51, 3.19) |
| Anakinra | 1.14 | 0.23 | 1.12 | (0.76, 1.66) |
| Tocilizumab | 1.55 | 0.25 | 1.53 | (1.13, 2.09) |
| Sarilumab | 1.78 | 0.34 | 1.74 | (1.21, 2.54) |
| Age<39 | 9.07 | 2.83 | 8.61 | (4.96, 15.83) |
| Age 40-49 | 3.50 | 0.64 | 3.43 | (2.43, 4.94) |
| Age 50-59 | 2.86 | 0.38 | 2.83 | (2.17, 3.68) |
| Age 70-79 | 0.54 | 0.07 | 0.53 | (0.41, 0.69) |
| Age 80+ | 0.26 | 0.06 | 0.25 | (0.16, 0.41) |
| Female | 1.44 | 0.16 | 1.43 | (1.16, 1.78) |

Table 32: Posterior probabilities for secondary analysis of hospital survival for Immune Modulation ITT population with independent tocilizumab/sarilumab effects

| | Posterior Probability |
|---|---|
| Interferon-beta-1a is optimal | 0.244 |
| Interferon-beta-1a is superior to control | 0.694 |
| Interferon-beta-1a is futile (OR < 1.2) | 0.452 |
| Interferon-beta-1a is harmful (OR < 1) | 0.306 |
| Anakinra is optimal | 0.001 |
| Anakinra is superior to control | 0.709 |
| Anakinra is futile (OR < 1.2) | 0.635 |
| Anakinra is harmful (OR < 1) | 0.291 |
| Tocilizumab is optimal | 0.127 |
| Tocilizumab is superior to control | 0.998 |
| Tocilizumab is futile (OR < 1.2) | 0.061 |
| Tocilizumab is harmful (OR < 1) | 0.003 |
| Sarilumab is optimal | 0.627 |
| Sarilumab is superior to control | 0.999 |
| Sarilumab is futile (OR < 1.2) | 0.023 |
| Sarilumab is harmful (OR < 1) | 0.001 |
| Interferon-beta-1a equivalent to anakinra | 0.280 |
| Interferon-beta-1a equivalent to tocilizumab | 0.279 |
| Interferon-beta-1a equivalent to sarilumab | 0.236 |
| Anakinra equivalent to tocilizumab | 0.193 |
| Anakinra equivalent to sarilumab | 0.059 |
| Tocilizumab equivalent to sarilumab | 0.639 |
| Anakinra superior to tocilizumab | 0.020 |
| Anakinra superior to sarilumab | 0.005 |
| Tocilizumab superior to sarilumab | 0.183 |

#### 3.9 Sensitivity analysis of hospital survival for Unblinded ITT population with site and time factors removed

- Model: Primary dichotomous model
- Factors: Age, sex, Immune Modulation Therapy Domain interventions (control, interferon-beta-1a, anakinra, tocilizumab, and sarilumab), COVID-19 Antiviral domain interventions (no antiviral, lopinavir–ritonavir, hydroxychloroquine, combination therapy), Corticosteroid domain interventions (no corticosteroid, fixed-duration corticosteroids, shock-dependent corticosteroids), Immunoglobulin domain interventions (no convalescent plasma, convalescent plasma, delayed convalescent plasma), and Anticoagulation interventions (usual care thromboprophylaxis, therapeutic anticoagu-

lation)

- Treatment effects for tocilizumab and sarilumab are modeled with independent priors.

Table 33: Odds ratio parameters for the sensitivity analysis of hospital survival with site and time removed

| | Mean | SD | Median | CrI |
|---|---|---|---|---|
| Interferon-beta-1a | 1.26 | 0.62 | 1.13 | (0.47, 2.87) |
| Anakinra | 0.97 | 0.17 | 0.95 | (0.68, 1.33) |
| Tocilizumab | 1.37 | 0.20 | 1.35 | (1.02, 1.80) |
| Sarilumab | 1.37 | 0.22 | 1.35 | (1.00, 1.88) |
| Lopinavir−ritonavir | 0.73 | 0.13 | 0.72 | (0.51, 1.03) |
| Hydroxychloroquine | 0.52 | 0.14 | 0.51 | (0.27, 0.82) |
| Fixed-dose corticosteroids | 0.83 | 0.20 | 0.81 | (0.51, 1.28) |
| Shock-dependent corticosteroids | 1.35 | 0.40 | 1.30 | (0.74, 2.30) |
| Therapeutic anticogulation | 0.85 | 0.12 | 0.84 | (0.64, 1.12) |
| Convalescent plasma | 1.04 | 0.10 | 1.03 | (0.85, 1.26) |
| Delayed convalescent plasma | 0.97 | 0.60 | 0.82 | (0.26, 2.54) |
| Lopinavir−ritonavir*Tocilizumab combination | 1.00 | 0.24 | 0.97 | (0.62, 1.55) |
| Hydroxychloroquine*Tocilizumab combination | 0.72 | 0.22 | 0.69 | (0.35, 1.20) |
| Tocilizumab*Fixed-dose corticosteroids combination | 1.13 | 0.30 | 1.09 | (0.65, 1.82) |
| Lopinavir−ritonavir*Tocilizumab interaction | 1.00 | 0.05 | 1.00 | (0.91, 1.10) |
| Tocilizumab*Fixed-dose corticosteroids interaction | 1.01 | 0.05 | 1.00 | (0.91, 1.11) |
| Hydroxychloroquine*Tocilizumab interaction | 0.99 | 0.05 | 0.99 | (0.90, 1.09) |
| Age<39 | 6.88 | 1.53 | 6.69 | (4.45, 10.43) |
| Age 40-49 | 3.10 | 0.41 | 3.07 | (2.38, 4.00) |
| Age 50-59 | 2.32 | 0.23 | 2.31 | (1.91, 2.80) |
| Age 70-79 | 0.54 | 0.05 | 0.54 | (0.45, 0.65) |
| Age 80+ | 0.33 | 0.06 | 0.32 | (0.23, 0.45) |
| Female | 1.27 | 0.10 | 1.26 | (1.08, 1.47) |

Table 34: Posterior probabilities for the sensitivity analysis of hospital survival with site and time removed

| | Posterior Probability |
|---|---|
| Interferon-beta-1a is optimal | 0.299 |
| Interferon-beta-1a is superior to control | 0.603 |
| Interferon-beta-1a is futile (OR < 1.2) | 0.552 |
| Interferon-beta-1a is harmful (OR < 1) | 0.397 |
| Anakinra is optimal | 0.001 |
| Anakinra is superior to control | 0.387 |
| Anakinra is futile (OR < 1.2) | 0.912 |
| Anakinra is harmful (OR < 1) | 0.613 |
| Tocilizumab is optimal | 0.342 |
| Tocilizumab is superior to control | 0.981 |
| Tocilizumab is futile (OR < 1.2) | 0.205 |
| Tocilizumab is harmful (OR < 1) | 0.019 |
| Sarilumab is optimal | 0.355 |
| Sarilumab is superior to control | 0.975 |
| Sarilumab is futile (OR < 1.2) | 0.224 |
| Sarilumab is harmful (OR < 1) | 0.025 |
| Lopinavir−ritonavir*tocilizumab combination OR > 1 | 0.455 |
| Hydroxychloroquine*tocilizumab combination OR > 1 | 0.111 |
| Fixed-dose steroids*tocilizumab combination OR > 1 | 0.629 |
| Interferon-beta-1a equivalent to anakinra | 0.289 |
| Interferon-beta-1a equivalent to tocilizumab | 0.292 |
| Interferon-beta-1a equivalent to sarilumab | 0.288 |
| Anakinra equivalent to tocilizumab | 0.123 |
| Anakinra equivalent to sarilumab | 0.136 |
| Tocilizumab equivalent to sarilumab | 0.818 |
| Anakinra superior to tocilizumab | 0.008 |
| Anakinra superior to sarilumab | 0.011 |
| Tocilizumab superior to sarilumab | 0.495 |

### 4 Safety analyses

#### 4.1 Empirical distribution of serious adverse events in Immune Modulation ITT population

Table 35: Summary of any serious adverse event (SAE) for the Immune Modulation ITT population. Note that this table shows the number of patients with any serious adverse event rather than the total number of serious adverse events observed.

| Intervention | # Patients (N) | # Known (n) | Any serious adverse event (y) | Rate of any serious adverse event (y/n) |
|---|---|---|---|---|
| Interferon-beta-1a | 19 | 19 | 1 | 0.053 |
| Anakinra | 373 | 373 | 10 | 0.027 |
| Tocilizumab | 947 | 947 | 24 | 0.025 |
| Sarilumab | 482 | 482 | 8 | 0.017 |
| Control | 405 | 405 | 11 | 0.027 |

#### 4.2 Primary safety analysis of serious adverse events in Immune Modulation ITT population

- Model: Primary dichotomous model
- Factors: Age, sex, site, Immune Modulation Therapy Domain interventions (control, interferon-beta-1a, anakinra, tocilizumab, and sarilumab)
- Treatment effects for tocilizumab and sarilumab are modeled with independent priors.

Table 36: Odds ratio parameters for the safety analysis of freedom from serious adverse events. ORs above 1 indicate fewer SAEs, and ORs below 1 indicate more SAEs.

| | Mean | SD | Median | CrI |
|---|---|---|---|---|
| Interferon-beta-1a | 1.26 | 1.16 | 0.92 | (0.22, 4.33) |
| Anakinra | 1.32 | 0.60 | 1.20 | (0.51, 2.77) |
| Tocilizumab | 1.15 | 0.41 | 1.08 | (0.53, 2.15) |
| Sarilumab | 1.68 | 0.79 | 1.52 | (0.65, 3.66) |
| Age<39 | 2.67 | 2.01 | 2.13 | (0.72, 7.70) |
| Age 40-49 | 2.22 | 1.24 | 1.91 | (0.80, 5.40) |
| Age 50-59 | 2.27 | 0.94 | 2.07 | (1.02, 4.64) |
| Age 70-79 | 0.99 | 0.34 | 0.93 | (0.49, 1.84) |
| Age 80+ | 1.89 | 1.50 | 1.47 | (0.45, 5.70) |
| Female | 1.00 | 0.31 | 0.96 | (0.54, 1.74) |

Table 37: Posterior probabilities for the primary safety analysis of freedom from serious adverse events

| | Posterior Probability |
|---|---|
| Interferon-beta-1a is superior to control | 0.456 |
| Anakinra is superior to control | 0.669 |
| Tocilizumab is superior to control | 0.592 |
| Sarilumab is superior to control | 0.840 |

### 5 Per protocol analyses

#### 5.1 Data summaries for per protocol analyses

Table 38: Summary of patients treated per protocol by treatment arm

| Intervention | Patients | Known | Treated per protocol |
|---|---|---|---|
| Interferon-beta-1a | 19 | 19 | 14 (73.7%) |
| Anakinra | 373 | 369 | 349 (94.6%) |
| Tocilizumab | 947 | 932 | 901 (96.7%) |
| Sarilumab | 482 | 468 | 449 (95.9%) |
| Control | 405 | 401 | 394 (98.3%) |
| Pooled IL-6ra | 1429 | 1400 | 1350 (96.4%) |

Table 39: Summary of OSFD for the Immune Modulation Per Protocol population

| Intervention | # Patients | # Known | In-hospital deaths | OSFD median (IQR) | OSFD in survivors* median (IQR) |
|---|---|---|---|---|---|
| Interferon-beta-1a | 14 | 14 | 5 (35.7%) | 0 (-1, 10.5) | 9 (0, 17) |
| Anakinra | 349 | 343 | 140 (40.8%) | 0 (-1, 15) | 14 (6.5, 18) |
| Tocilizumab | 901 | 896 | 294 (32.8%) | 8 (-1, 16.25) | 14.5 (7.25, 18) |
| Sarilumab | 449 | 447 | 145 (32.4%) | 9 (-1, 17) | 15 (8.25, 18) |
| Control | 394 | 394 | 143 (36.3%) | 0.5 (-1, 15) | 13 (4, 17) |
| Pooled IL-6ra | 1350 | 1343 | 439 (32.7%) | 8 (-1, 17) | 15 (8, 18) |

* Days free of organ support in survivors within 21 days

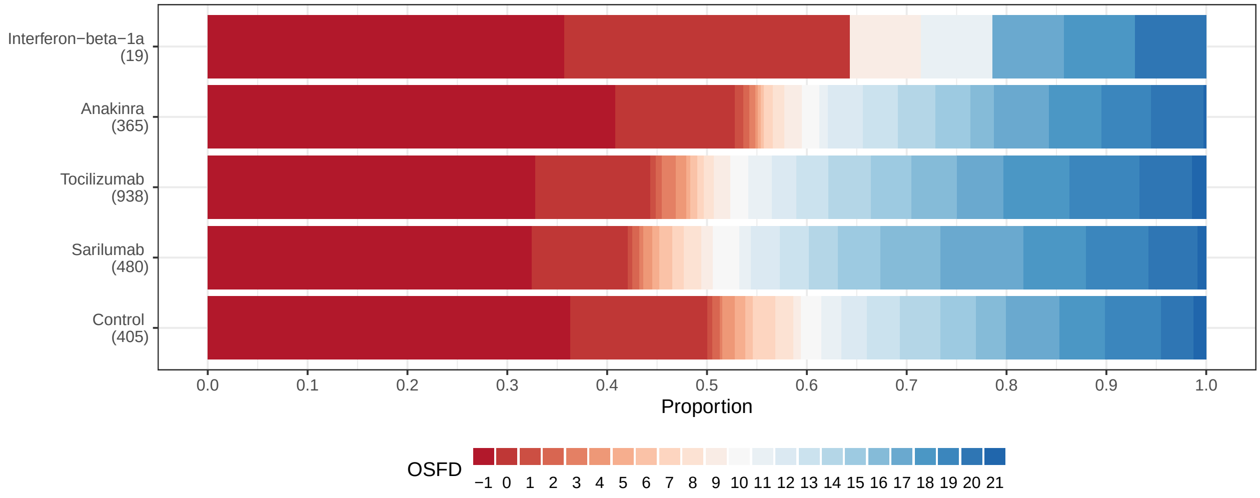

Figure 11: Empirical distribution of organ support free days (OSFD) for interferon-beta-1a, anakinra, tocilizumab, sarilumab, and control. This plot is restricted to the Immune Modulation Per Protocol population.

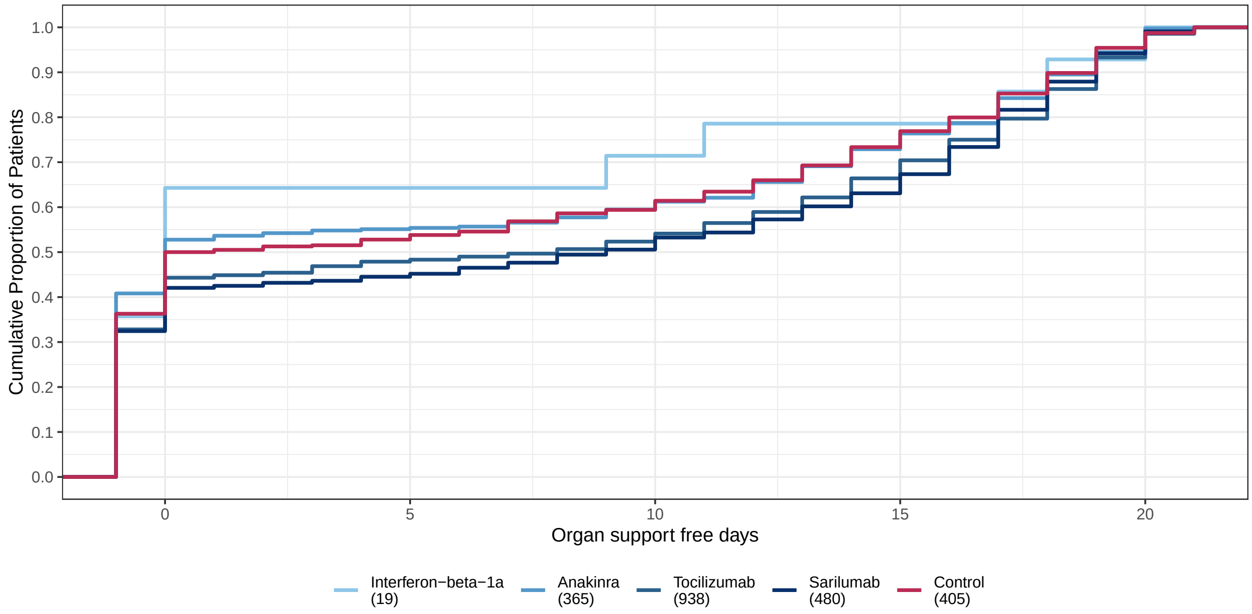

Figure 12: Empirical cumulative distribution of organ support free days (OSFD) for interferon-beta-1a, anakinra, tocilizumab, sarilumab, and control. This plot is restricted to the Immune Modulation Per Protocol population.

#### 5.2 Sensitivity analysis of OSFD for Immune Modulation per protocol population

- Model: Primary analysis ordinal model
- Factors: Age, sex, site, time, Immune Modulation Therapy Domain interventions (control, interferon-beta-1a, anakinra, tocilizumab, and sarilumab)
- Population: Immune modulation per protocol
- Treatment effects for tocilizumab and sarilumab are modeled with independent priors.

Table 40: Odds ratio parameters for secondary analysis of OSFD in the per protocol population

| | Mean | SD | Median | CrI |
|---|---|---|---|---|
| Interferon-beta-1a | 1.20 | 0.55 | 1.09 | (0.46, 2.55) |
| Anakinra | 1.14 | 0.19 | 1.13 | (0.81, 1.57) |
| Tocilizumab | 1.58 | 0.20 | 1.56 | (1.23, 1.99) |
| Sarilumab | 1.67 | 0.26 | 1.65 | (1.23, 2.24) |
| Age<39 | 4.57 | 0.77 | 4.52 | (3.25, 6.24) |
| Age 40-49 | 2.51 | 0.33 | 2.49 | (1.92, 3.22) |
| Age 50-59 | 2.19 | 0.23 | 2.18 | (1.77, 2.68) |
| Age 70-79 | 0.54 | 0.07 | 0.54 | (0.43, 0.68) |
| Age 80+ | 0.30 | 0.07 | 0.30 | (0.18, 0.47) |
| Female | 1.28 | 0.11 | 1.28 | (1.08, 1.51) |

Table 41: Posterior probabilities for secondary analysis of OSFD in the per protocol population

| | Posterior Probability |
|---|---|
| Interferon-beta-1a is optimal | 0.162 |
| Interferon-beta-1a is superior to control | 0.579 |
| Interferon-beta-1a is futile (OR < 1.2) | 0.587 |
| Interferon-beta-1a is harmful (OR < 1) | 0.421 |
| Anakinra is optimal | 0.000 |
| Anakinra is superior to control | 0.758 |
| Anakinra is futile (OR < 1.2) | 0.650 |
| Anakinra is harmful (OR < 1) | 0.242 |
| Tocilizumab is optimal | 0.254 |
| Tocilizumab is superior to control | 1.000 |
| Tocilizumab is futile (OR < 1.2) | 0.017 |
| Tocilizumab is harmful (OR < 1) | 0.000 |
| Sarilumab is optimal | 0.584 |
| Sarilumab is superior to control | 1.000 |
| Sarilumab is futile (OR < 1.2) | 0.018 |
| Sarilumab is harmful (OR < 1) | 0.001 |
| Interferon-beta-1a equivalent to anakinra | 0.309 |
| Interferon-beta-1a equivalent to tocilizumab | 0.227 |
| Interferon-beta-1a equivalent to sarilumab | 0.208 |
| Anakinra equivalent to tocilizumab | 0.124 |
| Anakinra equivalent to sarilumab | 0.075 |
| Tocilizumab equivalent to sarilumab | 0.841 |
| Anakinra superior to tocilizumab | 0.006 |
| Anakinra superior to sarilumab | 0.002 |
| Tocilizumab superior to sarilumab | 0.318 |

#### 5.3 Sensitivity analysis of hospital survival for Immune Modulation per protocol population

- Model: Primary dichotomous model
- Factors: Age, sex, site, time, Immune Modulation Therapy Domain interventions (control, interferon-beta-1a, anakinra, tocilizumab, and sarilumab)
- Population: Immune modulation per protocol
- Treatment effects for tocilizumab and sarilumab are modeled with independent priors.

Table 42: Odds ratio parameters for secondary analysis of hospital survival in the per protocol population

| | Mean | SD | Median | CrI |
|---|---|---|---|---|
| Interferon-beta-1a | 1.50 | 0.89 | 1.28 | (0.46, 3.78) |
| Anakinra | 1.10 | 0.23 | 1.07 | (0.71, 1.60) |
| Tocilizumab | 1.60 | 0.26 | 1.58 | (1.14, 2.16) |
| Sarilumab | 1.80 | 0.36 | 1.76 | (1.19, 2.61) |
| Age<39 | 9.21 | 2.95 | 8.71 | (4.93, 16.24) |
| Age 40-49 | 3.50 | 0.67 | 3.43 | (2.39, 5.03) |
| Age 50-59 | 2.84 | 0.40 | 2.81 | (2.13, 3.71) |
| Age 70-79 | 0.52 | 0.07 | 0.52 | (0.40, 0.68) |
| Age 80+ | 0.26 | 0.06 | 0.25 | (0.15, 0.40) |
| Female | 1.49 | 0.17 | 1.48 | (1.19, 1.83) |

Table 43: Posterior probabilities for secondary analysis of hospital survival in the per protocol population

| | Posterior Probability |
|---|---|
| Interferon-beta-1a is optimal | 0.279 |
| Interferon-beta-1a is superior to control | 0.685 |
| Interferon-beta-1a is futile (OR < 1.2) | 0.450 |
| Interferon-beta-1a is harmful (OR < 1) | 0.315 |
| Anakinra is optimal | 0.000 |
| Anakinra is superior to control | 0.639 |
| Anakinra is futile (OR < 1.2) | 0.701 |
| Anakinra is harmful (OR < 1) | 0.361 |
| Tocilizumab is optimal | 0.152 |
| Tocilizumab is superior to control | 0.998 |
| Tocilizumab is futile (OR < 1.2) | 0.046 |
| Tocilizumab is harmful (OR < 1) | 0.003 |
| Sarilumab is optimal | 0.568 |
| Sarilumab is superior to control | 0.998 |
| Sarilumab is futile (OR < 1.2) | 0.026 |
| Sarilumab is harmful (OR < 1) | 0.002 |
| Interferon-beta-1a equivalent to anakinra | 0.247 |
| Interferon-beta-1a equivalent to tocilizumab | 0.242 |
| Interferon-beta-1a equivalent to sarilumab | 0.220 |
| Anakinra equivalent to tocilizumab | 0.097 |
| Anakinra equivalent to sarilumab | 0.037 |
| Tocilizumab equivalent to sarilumab | 0.662 |
| Anakinra superior to tocilizumab | 0.007 |
| Anakinra superior to sarilumab | 0.003 |
| Tocilizumab superior to sarilumab | 0.229 |

### 6 Analyses of secondary endpoints

#### 6.1 Secondary analysis of 90-day mortality

- Model: Primary TTE model
- Factors: Age, sex, time, Immune Modulation Therapy Domain interventions (control, interferon-beta-1a, anakinra, tocilizumab, and sarilumab)
- Treatment effects for tocilizumab and sarilumab are modeled with independent priors.
- Note: The pre-specified time-to-event model did not converge. As a result, the statisticians running the model made the following modifications:
  - The analysis population was changed from the Unblinded ITT population to the Immune Modulation ITT population, and intervention/interactions for domains other than Immune Modulation were removed from the model
  - Site factors were removed from the model (set to 0)

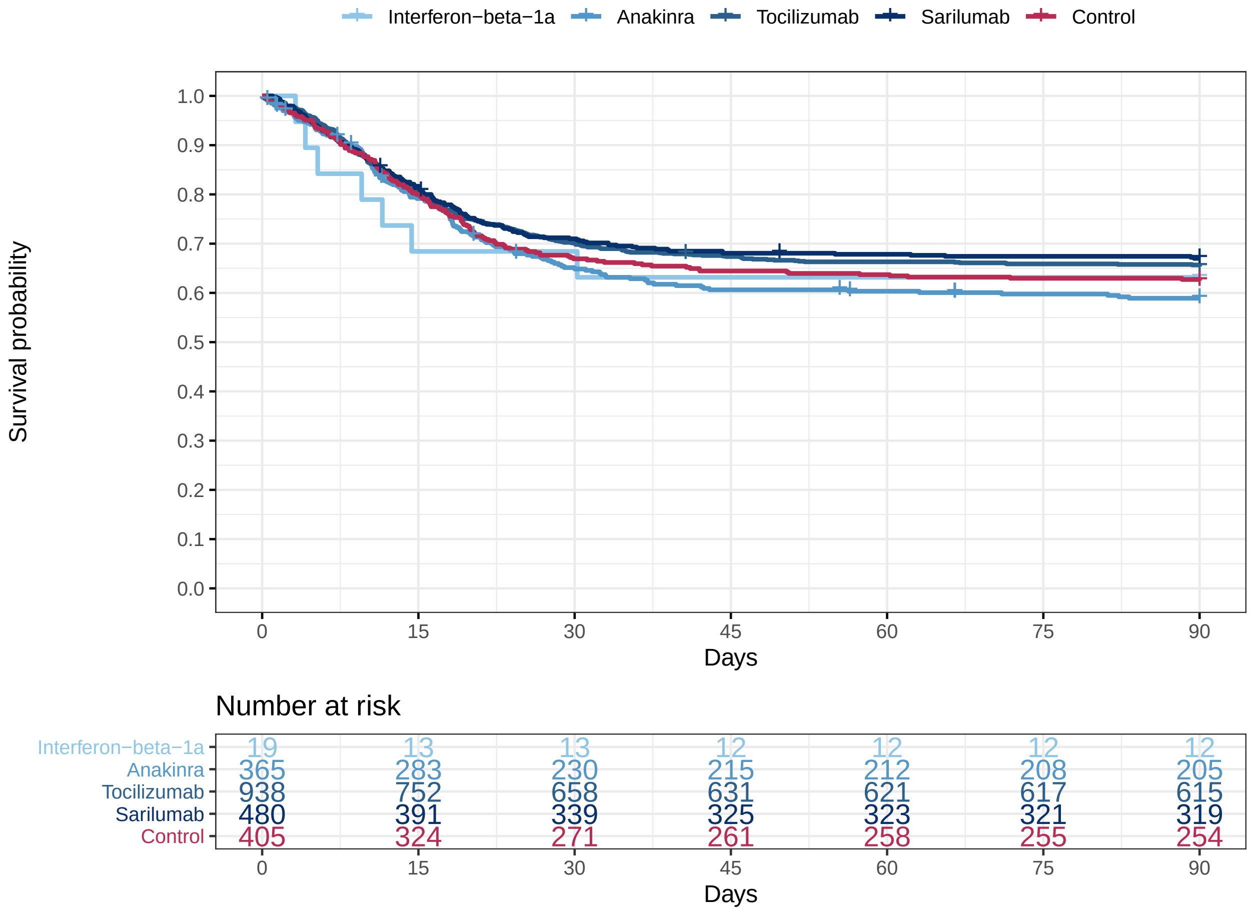

Figure 13: Empirical distribution of 90-day mortality for interferon-beta-1a, anakinra, tocilizumab, sarilumab, and control. This plot is restricted to the Immune Modulation ITT population.

Table 44: Summary of 2.5th, 10th, 25th, 50th, 75th, 90th, and 97.5th percentiles from the Kaplan-Meier estimates for mortality (in days). Displaying the observed percentiles for this outcome.

| | 2.5 | 10.0 | 25.0 | 50.0 | 75.0 | 90.0 | 97.5 |
|---|---|---|---|---|---|---|---|
| Interferon-beta-1a | 3.2 | 4.15 | 11.54 | - | - | - | - |
| Anakinra | 1.99 | 8.62 | 18.04 | - | - | - | - |
| Tocilizumab | 3.27 | 8.15 | 20.03 | - | - | - | - |
| Sarilumab | 3.08 | 8.26 | 20.38 | - | - | - | - |
| Control | 2.21 | 7.89 | 19.09 | - | - | - | - |

Table 45: Hazard ratio parameters for secondary analysis of 90-day survival. Hazard ratios above 1 indicate benefit, and hazard ratios below 1 indicate harm.

| | Mean | SD | Median | CrI |
|---|---|---|---|---|
| Interferon-beta-1a | 1.26 | 0.51 | 1.16 | (0.60, 2.53) |
| Anakinra | 1.15 | 0.16 | 1.13 | (0.87, 1.49) |
| Tocilizumab | 1.40 | 0.16 | 1.39 | (1.11, 1.74) |
| Sarilumab | 1.46 | 0.20 | 1.44 | (1.11, 1.89) |
| Age<39 | 4.45 | 1.15 | 4.27 | (2.71, 7.17) |
| Age 40-49 | 2.25 | 0.33 | 2.22 | (1.69, 2.97) |
| Age 50-59 | 2.10 | 0.23 | 2.09 | (1.70, 2.59) |
| Age 70-79 | 0.64 | 0.06 | 0.64 | (0.54, 0.76) |
| Age 80+ | 0.36 | 0.05 | 0.36 | (0.28, 0.48) |
| Female | 1.31 | 0.11 | 1.30 | (1.11, 1.53) |

Table 46: Posterior probabilities for secondary analysis of 90-day survival

| | Posterior Probability |
|---|---|
| Interferon-beta-1a is optimal | 0.262 |
| Interferon-beta-1a is superior to control | 0.658 |
| Interferon-beta-1a is futile (OR < 1.2) | 0.539 |
| Interferon-beta-1a is harmful (OR < 1) | 0.342 |
| Anakinra is optimal | 0.005 |
| Anakinra is superior to control | 0.823 |
| Anakinra is futile (OR < 1.2) | 0.661 |
| Anakinra is harmful (OR < 1) | 0.177 |
| Tocilizumab is optimal | 0.254 |
| Tocilizumab is superior to control | 0.999 |
| Tocilizumab is futile (OR < 1.2) | 0.092 |
| Tocilizumab is harmful (OR < 1) | 0.002 |
| Sarilumab is optimal | 0.479 |
| Sarilumab is superior to control | 0.996 |
| Sarilumab is futile (OR < 1.2) | 0.085 |
| Sarilumab is harmful (OR < 1) | 0.004 |
| Interferon-beta-1a equivalent to anakinra | 0.374 |
| Interferon-beta-1a equivalent to tocilizumab | 0.327 |
| Interferon-beta-1a equivalent to sarilumab | 0.305 |
| Anakinra equivalent to tocilizumab | 0.411 |
| Anakinra equivalent to sarilumab | 0.306 |
| Tocilizumab equivalent to sarilumab | 0.917 |
| Anakinra superior to tocilizumab | 0.027 |
| Anakinra superior to sarilumab | 0.020 |
| Tocilizumab superior to sarilumab | 0.364 |

#### 6.2 Secondary analysis of progression to intubation, ECMO, or death

- Model: Primary dichotomous model
- Factors: Age, sex, site, time, Immune Modulation Therapy Domain interventions (control, interferon-beta-1a, anakinra, tocilizumab, and sarilumab), COVID-19 Antiviral domain interventions (no antiviral, lopinavir−ritonavir, hydroxychloroquine, combination therapy), Corticosteroid domain interventions (no corticosteroid, fixed-duration corticosteroids, shock-dependent corticosteroids), Immunoglobulin domain interventions (no convalescent plasma, convalescent plasma, delayed convalescent plasma), and Anticoagulation interventions (usual care thromboprophylaxis, therapeutic anticoagulation)
- Treatment effects for tocilizumab and sarilumab are modeled with independent priors.

Table 47: Summary of progression to intubation, ECMO, or death for the Unblinded ITT population restricted to patients not on mechanical ventilation or ECMO at baseline

| Intervention | # Patients (N) | # Progressors (y) | Progression Rate (y/N) |
|---|---|---|---|
| Interferon-beta-1a | 12 | 6 | 0.500 |
| Anakinra | 228 | 122 | 0.535 |
| Tocilizumab | 613 | 266 | 0.434 |
| Sarilumab | 331 | 149 | 0.450 |
| Control | 276 | 147 | 0.533 |

Table 48: Summary of progression to intubation, ECMO, or death by component for the Unblinded ITT population restricted to patients not on mechanical ventilation or ECMO at baseline. Note that a patient may progress on more than one component of the composite endpoint, so the sum of events in this table may not match the total number of progressors.

| Intervention | # Patients (N) | Death, n (%) | Intubation, n (%) | ECMO, n (%) |
|---|---|---|---|---|
| Interferon-beta-1a | 12 | 3 (25) | 6 (50) | 0 (0) |
| Anakinra | 228 | 75 (32.9) | 103 (45.2) | 2 (0.9) |
| Tocilizumab | 613 | 173 (28.2) | 214 (34.9) | 6 (1) |
| Sarilumab | 331 | 102 (30.8) | 110 (33.2) | 3 (0.9) |
| Control | 276 | 88 (31.9) | 121 (43.8) | 3 (1.1) |

Table 49: Odds ratio parameters for secondary analysis of freedom from progression to intubation, ECMO, or death

| | Mean | SD | Median | CrI |
|---|---|---|---|---|
| Interferon-beta-1a | 1.33 | 0.79 | 1.15 | (0.39, 3.35) |
| Anakinra | 1.19 | 0.26 | 1.16 | (0.76, 1.78) |
| Tocilizumab | 1.70 | 0.29 | 1.67 | (1.20, 2.36) |
| Sarilumab | 1.46 | 0.29 | 1.43 | (0.98, 2.10) |
| Lopinavir−ritonavir | 0.75 | 0.16 | 0.73 | (0.49, 1.10) |
| Hydroxychloroquine | 0.48 | 0.19 | 0.47 | (0.16, 0.88) |
| Fixed-dose corticosteroids | 2.98 | 1.17 | 2.78 | (1.32, 5.87) |
| Shock-dependent corticosteroids | 1.13 | 0.46 | 1.04 | (0.50, 2.26) |
| Therapeutic anticogulation | 0.89 | 0.15 | 0.88 | (0.64, 1.21) |
| Convalescent plasma | 0.84 | 0.10 | 0.83 | (0.66, 1.05) |
| Delayed convalescent plasma | 0.38 | 0.35 | 0.29 | (0.06, 1.23) |
| Lopinavir−ritonavir*Tocilizumab combination | 1.28 | 0.36 | 1.23 | (0.73, 2.12) |
| Hydroxychloroquine*Tocilizumab combination | 0.81 | 0.36 | 0.77 | (0.25, 1.62) |
| Tocilizumab*Fixed-dose corticosteroids combination | 5.10 | 2.24 | 4.68 | (2.07, 10.75) |
| Lopinavir−ritonavir*Tocilizumab interaction | 1.01 | 0.05 | 1.01 | (0.91, 1.11) |
| Tocilizumab*Fixed-dose corticosteroids interaction | 1.00 | 0.05 | 1.00 | (0.91, 1.10) |
| Hydroxychloroquine*Tocilizumab interaction | 1.01 | 0.05 | 1.01 | (0.91, 1.11) |
| Age<39 | 4.75 | 1.05 | 4.62 | (3.09, 7.17) |
| Age 40-49 | 2.29 | 0.33 | 2.27 | (1.70, 3.00) |
| Age 50-59 | 1.74 | 0.21 | 1.73 | (1.37, 2.18) |
| Age 70-79 | 0.64 | 0.08 | 0.63 | (0.49, 0.82) |
| Age 80+ | 0.44 | 0.09 | 0.43 | (0.28, 0.64) |
| Female | 1.13 | 0.11 | 1.13 | (0.94, 1.36) |

Table 50: Posterior probabilities for secondary analysis of freedom from progression to intubation, ECMO, or death

| | Posterior Probability |
|---|---|
| Interferon-beta-1a is optimal | 0.239 |
| Interferon-beta-1a is superior to control | 0.596 |
| Interferon-beta-1a is futile (OR < 1.2) | 0.533 |
| Interferon-beta-1a is harmful (OR < 1) | 0.405 |
| Anakinra is optimal | 0.011 |
| Anakinra is superior to control | 0.750 |
| Anakinra is futile (OR < 1.2) | 0.564 |
| Anakinra is harmful (OR < 1) | 0.251 |
| Tocilizumab is optimal | 0.639 |
| Tocilizumab is superior to control | 0.999 |
| Tocilizumab is futile (OR < 1.2) | 0.025 |
| Tocilizumab is harmful (OR < 1) | 0.001 |
| Sarilumab is optimal | 0.111 |
| Sarilumab is superior to control | 0.969 |
| Sarilumab is futile (OR < 1.2) | 0.180 |
| Sarilumab is harmful (OR < 1) | 0.031 |
| Lopinavir−ritonavir*tocilizumab combination OR > 1 | 0.774 |
| Hydroxychloroquine*tocilizumab combination OR > 1 | 0.269 |
| Fixed-dose steroids*tocilizumab combination OR > 1 | 1.000 |
| Interferon-beta-1a equivalent to anakinra | 0.244 |
| Interferon-beta-1a equivalent to tocilizumab | 0.212 |
| Interferon-beta-1a equivalent to sarilumab | 0.231 |
| Anakinra equivalent to tocilizumab | 0.158 |
| Anakinra equivalent to sarilumab | 0.421 |
| Tocilizumab equivalent to sarilumab | 0.548 |
| Anakinra superior to tocilizumab | 0.025 |
| Anakinra superior to sarilumab | 0.140 |
| Tocilizumab superior to sarilumab | 0.831 |

#### 6.3 Secondary analysis of cardiovascular support-free days

- Model: Primary analysis ordinal model
- Factors: Age, sex, site, time, Immune Modulation Therapy Domain interventions (control, interferon-beta-1a, anakinra, tocilizumab, and sarilumab), COVID-19 Antiviral domain interventions (no antiviral, lopinavir−ritonavir, hydroxychloroquine, combination therapy), Corticosteroid domain interventions (no corticosteroid, fixed-duration corticosteroids, shock-dependent corticosteroids), Immunoglobulin domain interventions (no convalescent plasma, convalescent plasma, delayed convalescent plasma),

and Anticoagulation interventions (usual care thromboprophylaxis, therapeutic anticoagulation)

- Treatment effects for tocilizumab and sarilumab are modeled with independent priors.
- There are 26 missing values of days free of vasopressors/inotropes in the Unblinded ITT population due to missing daily data. These patients are removed from this analysis.

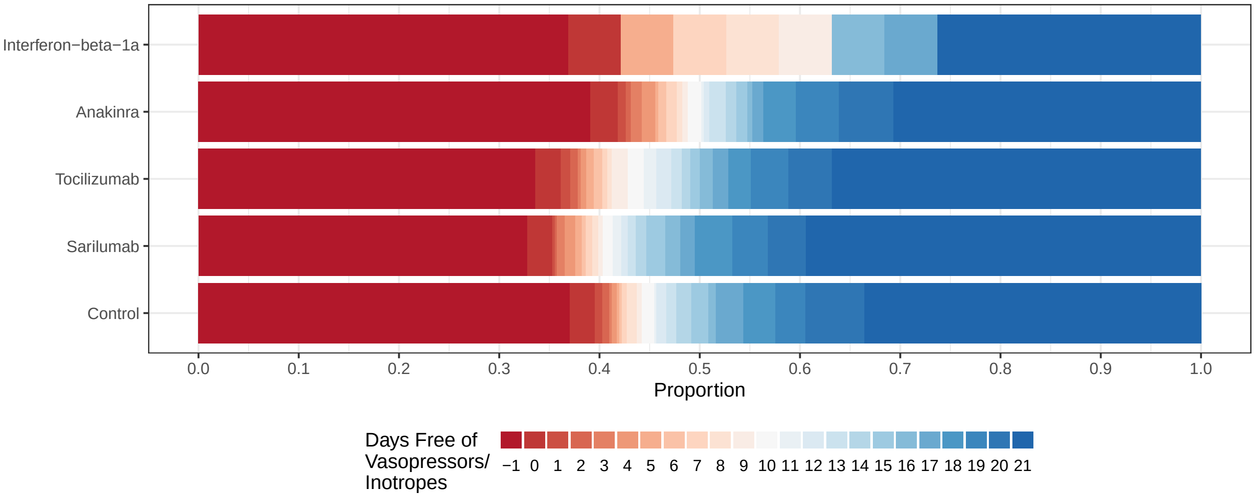

Figure 14: Empirical distribution of cardiovascular support-free days for interferon-beta-1a, anakinra, tocilizumab, sarilumab, and control. This plot is restricted to the Immune Modulation ITT population.

Table 51: Summary of cardiovascular support-free days in the Immune Modulation ITT population

| Intervention | # Patients | # Known | Cardiovascular support-free days median (IQR) | Days free of cardiovascular support in survivors* median (IQR) |
|---|---|---|---|---|
| Interferon-beta-1a | 19 | 19 | 7 (-1, 19) | 16.5 (7.75, 21) |
| Anakinra | 373 | 371 | 10 (-1, 21) | 21 (15, 21) |
| Tocilizumab | 947 | 944 | 15.5 (-1, 21) | 21 (16, 21) |
| Sarilumab | 482 | 479 | 18 (-1, 21) | 21 (18, 21) |
| Control | 405 | 405 | 15 (-1, 21) | 21 (17, 21) |

* Days free of cardiovascular support in survivors within 21 days

Table 52: Odds ratio parameters for secondary analysis of cardiovascular support-free days

| | Mean | SD | Median | CrI |
|---|---|---|---|---|
| Interferon-beta-1a | 1.08 | 0.47 | 0.99 | (0.44, 2.20) |
| Anakinra | 1.09 | 0.18 | 1.07 | (0.78, 1.47) |
| Tocilizumab | 1.46 | 0.19 | 1.44 | (1.12, 1.87) |
| Sarilumab | 1.60 | 0.24 | 1.58 | (1.19, 2.12) |
| Lopinavir−ritonavir | 0.73 | 0.11 | 0.72 | (0.54, 0.96) |
| Hydroxychloroquine | 0.64 | 0.13 | 0.63 | (0.41, 0.90) |
| Fixed-dose corticosteroids | 1.46 | 0.32 | 1.43 | (0.93, 2.18) |
| Shock-dependent corticosteroids | 1.19 | 0.27 | 1.16 | (0.75, 1.78) |
| Therapeutic anticogulation | 0.91 | 0.11 | 0.90 | (0.71, 1.15) |
| Convalescent plasma | 0.95 | 0.08 | 0.95 | (0.80, 1.13) |
| Delayed convalescent plasma | 0.89 | 0.54 | 0.76 | (0.25, 2.33) |
| Lopinavir−ritonavir*Tocilizumab combination | 1.07 | 0.21 | 1.05 | (0.71, 1.56) |
| Hydroxychloroquine*Tocilizumab combination | 0.93 | 0.23 | 0.91 | (0.55, 1.44) |
| Tocilizumab*Fixed-dose corticosteroids combination | 2.13 | 0.55 | 2.06 | (1.24, 3.40) |
| Lopinavir−ritonavir*Tocilizumab interaction | 1.01 | 0.05 | 1.01 | (0.92, 1.11) |
| Tocilizumab*Fixed-dose corticosteroids interaction | 1.01 | 0.05 | 1.01 | (0.91, 1.11) |
| Hydroxychloroquine*Tocilizumab interaction | 1.00 | 0.05 | 1.00 | (0.91, 1.10) |
| Age<39 | 4.40 | 0.59 | 4.37 | (3.38, 5.66) |
| Age 40-49 | 2.72 | 0.28 | 2.70 | (2.22, 3.31) |
| Age 50-59 | 2.01 | 0.17 | 2.00 | (1.70, 2.35) |
| Age 70-79 | 0.51 | 0.05 | 0.51 | (0.43, 0.62) |
| Age 80+ | 0.28 | 0.05 | 0.28 | (0.19, 0.39) |
| Female | 1.12 | 0.07 | 1.12 | (0.98, 1.27) |

Table 53: Posterior probabilities for secondary analysis of cardiovascular support-free days

| | Posterior Probability |
|---|---|
| Interferon-beta-1a is optimal | 0.119 |
| Interferon-beta-1a is superior to control | 0.493 |
| Interferon-beta-1a is futile (OR < 1.2) | 0.674 |
| Interferon-beta-1a is harmful (OR < 1) | 0.507 |
| Anakinra is optimal | 0.001 |
| Anakinra is superior to control | 0.666 |
| Anakinra is futile (OR < 1.2) | 0.755 |
| Anakinra is harmful (OR < 1) | 0.334 |
| Tocilizumab is optimal | 0.183 |
| Tocilizumab is superior to control | 0.998 |
| Tocilizumab is futile (OR < 1.2) | 0.083 |
| Tocilizumab is harmful (OR < 1) | 0.003 |
| Sarilumab is optimal | 0.697 |
| Sarilumab is superior to control | 0.999 |
| Sarilumab is futile (OR < 1.2) | 0.028 |
| Sarilumab is harmful (OR < 1) | 0.001 |
| Lopinavir–ritonavir*tocilizumab combination OR > 1 | 0.589 |
| Hydroxychloroquine*tocilizumab combination OR > 1 | 0.346 |
| Fixed-dose steroids*tocilizumab combination OR > 1 | 0.998 |
| Interferon-beta-1a equivalent to anakinra | 0.324 |
| Interferon-beta-1a equivalent to tocilizumab | 0.235 |
| Interferon-beta-1a equivalent to sarilumab | 0.191 |
| Anakinra equivalent to tocilizumab | 0.203 |
| Anakinra equivalent to sarilumab | 0.075 |
| Tocilizumab equivalent to sarilumab | 0.745 |
| Anakinra superior to tocilizumab | 0.014 |
| Anakinra superior to sarilumab | 0.003 |
| Tocilizumab superior to sarilumab | 0.224 |

#### 6.4 Secondary analysis of respiratory support-free days

- Model: Primary analysis ordinal model
- Factors: Age, sex, site, time, Immune Modulation Therapy Domain interventions (control, interferon-beta-1a, anakinra, tocilizumab, and sarilumab), COVID-19 Antiviral domain interventions (no antiviral, lopinavir–ritonavir, hydroxychloroquine, combination therapy), Corticosteroid domain interventions (no corticosteroid, fixed-duration corticosteroids, shock-dependent corticosteroids), Immunoglobulin domain interventions (no convalescent plasma, convalescent plasma, delayed convalescent plasma), and Anticoagulation interventions (usual care thromboprophylaxis, therapeutic anticoagulation)

- Treatment effects for tocilizumab and sarilumab are modeled with independent priors.
- There are 26 missing values of days free of ventilation in the Unblinded ITT population due to missing daily data. These patients are removed from this analysis.

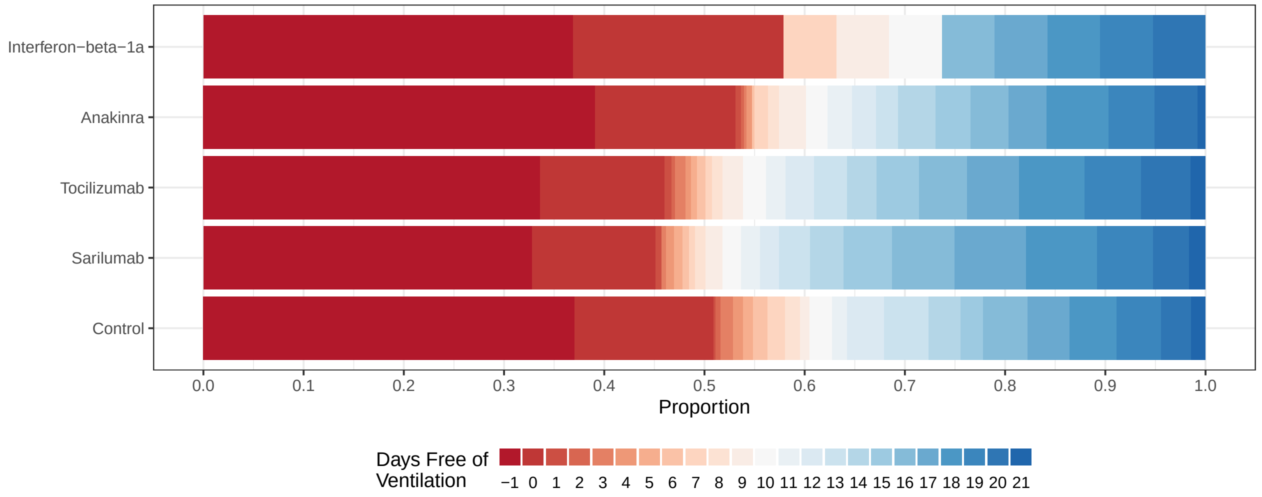

Figure 15: Empirical distribution of respiratory support-free days for interferon-beta-1a, anakinra, tocilizumab, sarilumab, and control. This plot is restricted to the Immune Modulation ITT population.

Table 54: Summary of respiratory support-free days for the Immune Modulation ITT population

| Intervention | # Patients | # Known | Respiratory support-free days median (IQR) | Respiratory support-free days in survivors* median (IQR) |
|---|---|---|---|---|
| Interferon-beta-1a | 19 | 19 | 0 (-1, 13) | 9.5 (0, 17.25) |
| Anakinra | 373 | 371 | 0 (-1, 15) | 13.5 (3.75, 18) |
| Tocilizumab | 947 | 944 | 6 (-1, 16) | 14 (6, 18) |
| Sarilumab | 482 | 479 | 8 (-1, 16.5) | 15 (8, 18) |
| Control | 405 | 405 | 0 (-1, 14) | 13 (3.5, 17) |

* Days free of respiratory support in survivors within 21 days

Table 55: Odds ratio parameters for secondary analysis of respiratory support-free days

| | Mean | SD | Median | CrI |
|---|---|---|---|---|
| Interferon-beta-1a | 1.36 | 0.55 | 1.27 | (0.58, 2.72) |
| Anakinra | 1.07 | 0.17 | 1.06 | (0.78, 1.43) |
| Tocilizumab | 1.52 | 0.19 | 1.51 | (1.18, 1.92) |
| Sarilumab | 1.61 | 0.23 | 1.60 | (1.21, 2.09) |
| Lopinavir−ritonavir | 0.81 | 0.12 | 0.80 | (0.61, 1.07) |
| Hydroxychloroquine | 0.65 | 0.14 | 0.65 | (0.39, 0.93) |
| Fixed-dose corticosteroids | 1.47 | 0.32 | 1.44 | (0.94, 2.18) |
| Shock-dependent corticosteroids | 1.25 | 0.28 | 1.22 | (0.79, 1.87) |
| Therapeutic anticogulation | 0.82 | 0.10 | 0.82 | (0.65, 1.02) |
| Convalescent plasma | 0.94 | 0.08 | 0.94 | (0.80, 1.11) |
| Delayed convalescent plasma | 0.61 | 0.35 | 0.53 | (0.18, 1.49) |
| Lopinavir−ritonavir*Tocilizumab combination | 1.24 | 0.24 | 1.22 | (0.84, 1.79) |
| Hydroxychloroquine*Tocilizumab combination | 0.99 | 0.25 | 0.97 | (0.56, 1.55) |
| Tocilizumab*Fixed-dose corticosteroids combination | 2.23 | 0.56 | 2.16 | (1.32, 3.50) |
| Lopinavir−ritonavir*Tocilizumab interaction | 1.01 | 0.05 | 1.01 | (0.92, 1.11) |
| Tocilizumab*Fixed-dose corticosteroids interaction | 1.01 | 0.05 | 1.00 | (0.91, 1.11) |
| Hydroxychloroquine*Tocilizumab interaction | 1.00 | 0.05 | 1.00 | (0.91, 1.10) |
| Age<39 | 4.22 | 0.51 | 4.19 | (3.32, 5.32) |
| Age 40-49 | 2.48 | 0.24 | 2.47 | (2.03, 2.99) |
| Age 50-59 | 1.84 | 0.15 | 1.83 | (1.56, 2.15) |
| Age 70-79 | 0.52 | 0.05 | 0.52 | (0.43, 0.62) |
| Age 80+ | 0.30 | 0.05 | 0.30 | (0.21, 0.42) |
| Female | 1.14 | 0.07 | 1.14 | (1.00, 1.28) |

Table 56: Posterior probabilities for secondary analysis of respiratory support-free days

| | Posterior Probability |
|---|---|
| Interferon-beta-1a is optimal | 0.259 |
| Interferon-beta-1a is superior to control | 0.723 |
| Interferon-beta-1a is futile (OR < 1.2) | 0.445 |
| Interferon-beta-1a is harmful (OR < 1) | 0.277 |
| Anakinra is optimal | 0.000 |
| Anakinra is superior to control | 0.633 |
| Anakinra is futile (OR < 1.2) | 0.802 |
| Anakinra is harmful (OR < 1) | 0.367 |
| Tocilizumab is optimal | 0.220 |
| Tocilizumab is superior to control | 1.000 |
| Tocilizumab is futile (OR < 1.2) | 0.032 |
| Tocilizumab is harmful (OR < 1) | 0.000 |
| Sarilumab is optimal | 0.522 |
| Sarilumab is superior to control | 1.000 |
| Sarilumab is futile (OR < 1.2) | 0.021 |
| Sarilumab is harmful (OR < 1) | 0.000 |
| Lopinavir−ritonavir*tocilizumab combination OR > 1 | 0.845 |
| Hydroxychloroquine*tocilizumab combination OR > 1 | 0.455 |
| Fixed-dose steroids*tocilizumab combination OR > 1 | 1.000 |
| Interferon-beta-1a equivalent to anakinra | 0.319 |
| Interferon-beta-1a equivalent to tocilizumab | 0.325 |
| Interferon-beta-1a equivalent to sarilumab | 0.297 |
| Anakinra equivalent to tocilizumab | 0.092 |
| Anakinra equivalent to sarilumab | 0.046 |
| Tocilizumab equivalent to sarilumab | 0.823 |
| Anakinra superior to tocilizumab | 0.004 |
| Anakinra superior to sarilumab | 0.002 |
| Tocilizumab superior to sarilumab | 0.310 |

#### 6.5 Secondary analysis of length of ICU stay

- Model: Primary TTE model
- Factors: Age, sex, time, Immune Modulation Therapy Domain interventions (control, interferon-beta-1a, anakinra, tocilizumab, and sarilumab)
- Treatment effects for tocilizumab and sarilumab are modeled with independent priors.
- Note: The pre-specified time-to-event model did not converge. As a result, the statisticians running the model made the following modifications:
  - The analysis population was changed from the Unblinded ITT population to the Immune Modula-

tion ITT population, and intervention/interactions for domains other than Immune Modulation were removed from the model

– Site factors were removed from the model (set to 0)

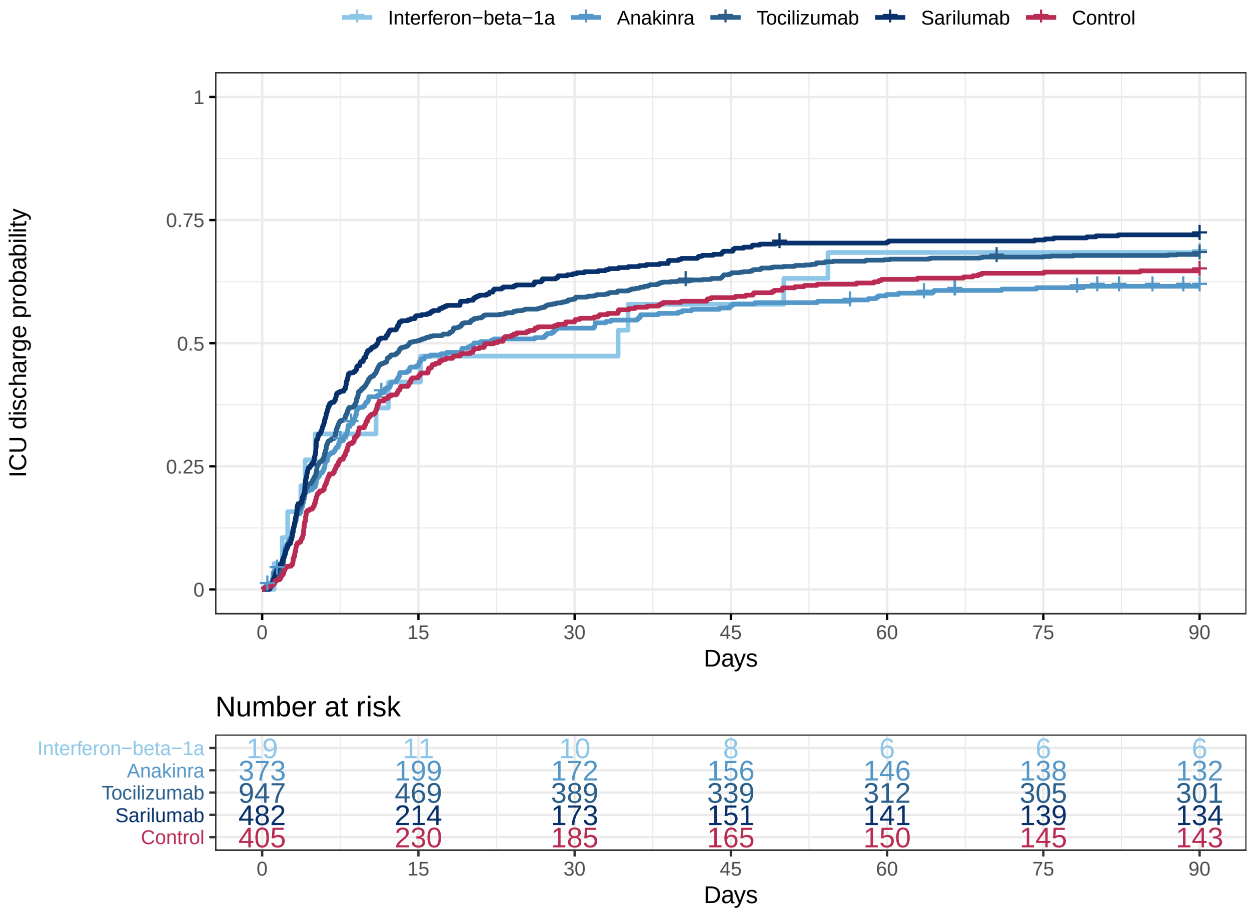

Figure 16: Empirical distribution of length of ICU stay for interferon-beta-1a, anakinra, tocilizumab, sarilumab, and control. This plot is restricted to the Immune Modulation ITT population.

Table 57: Summary of 2.5th, 10th, 25th, 50th, 75th, 90th, and 97.5th percentiles from the Kaplan-Meier estimates for length of ICU stay (in days). Displaying the observed percentiles for this outcome.

| | 2.5 | 10.0 | 25.0 | 50.0 | 75.0 | 90.0 | 97.5 |
|---|---|---|---|---|---|---|---|
| Interferon-beta-1a | 1.15 | 1.92 | 4.12 | 34.17 | - | - | - |
| Anakinra | 1.03 | 2.68 | 6.01 | 20.33 | - | - | - |
| Tocilizumab | 1.17 | 2.75 | 5.29 | 14.17 | - | - | - |
| Sarilumab | 1.21 | 2.78 | 4.6 | 11.09 | - | - | - |
| Control | 1.82 | 3.73 | 7.13 | 22.23 | - | - | - |

Table 58: Hazard ratio parameters for secondary analysis of length of ICU stay

| | Mean | SD | Median | CrI |
|---|---|---|---|---|
| Interferon-beta-1a | 1.21 | 0.33 | 1.17 | (0.65, 1.95) |
| Anakinra | 1.10 | 0.12 | 1.10 | (0.89, 1.36) |
| Tocilizumab | 1.30 | 0.11 | 1.29 | (1.10, 1.52) |
| Sarilumab | 1.42 | 0.14 | 1.41 | (1.16, 1.72) |
| Age<39 | 2.61 | 0.26 | 2.60 | (2.12, 3.16) |
| Age 40-49 | 1.70 | 0.14 | 1.69 | (1.44, 1.99) |
| Age 50-59 | 1.62 | 0.11 | 1.62 | (1.42, 1.85) |
| Age 70-79 | 0.73 | 0.06 | 0.73 | (0.62, 0.86) |
| Age 80+ | 0.63 | 0.10 | 0.62 | (0.45, 0.84) |
| Female | 1.18 | 0.06 | 1.18 | (1.06, 1.32) |

Table 59: Posterior probabilities for secondary analysis of length of ICU stay

| | Posterior Probability |
|---|---|
| Interferon-beta-1a is optimal | 0.246 |
| Interferon-beta-1a is superior to control | 0.710 |
| Interferon-beta-1a is futile (OR < 1.2) | 0.534 |
| Interferon-beta-1a is harmful (OR < 1) | 0.290 |
| Anakinra is optimal | 0.001 |
| Anakinra is superior to control | 0.802 |
| Anakinra is futile (OR < 1.2) | 0.802 |
| Anakinra is harmful (OR < 1) | 0.198 |
| Tocilizumab is optimal | 0.066 |
| Tocilizumab is superior to control | 0.999 |
| Tocilizumab is futile (OR < 1.2) | 0.184 |
| Tocilizumab is harmful (OR < 1) | 0.001 |
| Sarilumab is optimal | 0.687 |
| Sarilumab is superior to control | 1.000 |
| Sarilumab is futile (OR < 1.2) | 0.050 |
| Sarilumab is harmful (OR < 1) | 0.000 |
| Interferon-beta-1a equivalent to anakinra | 0.455 |
| Interferon-beta-1a equivalent to tocilizumab | 0.467 |
| Interferon-beta-1a equivalent to sarilumab | 0.408 |
| Anakinra equivalent to tocilizumab | 0.591 |
| Anakinra equivalent to sarilumab | 0.209 |
| Tocilizumab equivalent to sarilumab | 0.906 |
| Anakinra superior to tocilizumab | 0.026 |
| Anakinra superior to sarilumab | 0.002 |
| Tocilizumab superior to sarilumab | 0.099 |

#### 6.6 Secondary analysis of length of hospital stay

- Model: Primary TTE model
- Factors: Age, sex, time, Immune Modulation Therapy Domain interventions (control, interferon-beta-1a, anakinra, tocilizumab, and sarilumab)
- Treatment effects for tocilizumab and sarilumab are modeled with independent priors.
- Note: The pre-specified time-to-event model did not converge. As a result, the statisticians running the model made the following modifications:
  - The analysis population was changed from the Unblinded ITT population to the Immune Modulation ITT population, and intervention/interactions for domains other than Immune Modulation were removed from the model

– Site factors were removed from the model (set to 0)

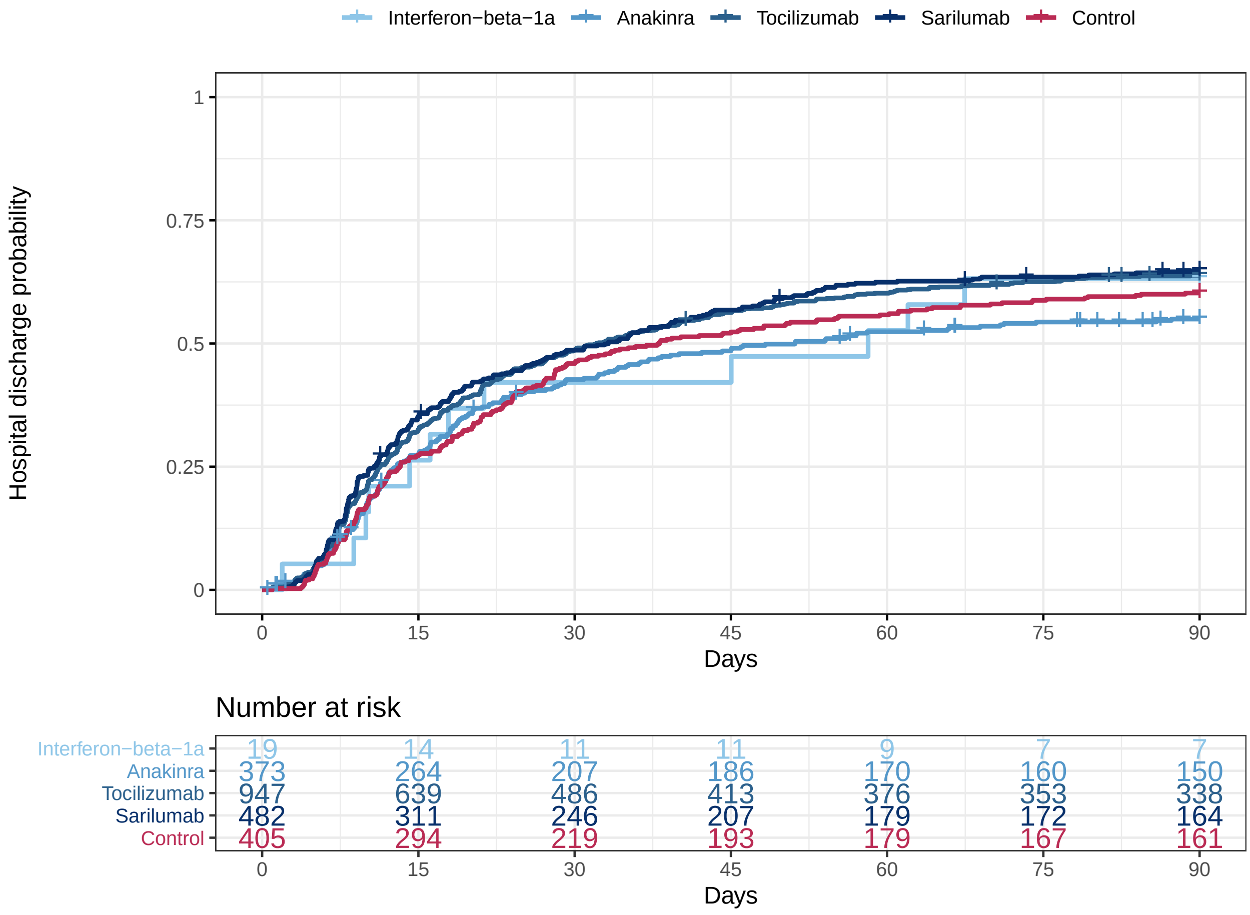

Figure 17: Empirical distribution of length of hospital stay for interferon-beta-1a, anakinra, tocilizumab, sarilumab, and control. This plot is restricted to the Immune Modulation ITT population.

Table 60: Summary of 2.5th, 10th, 25th, 50th, 75th, 90th, and 97.5th percentiles from the Kaplan-Meier estimates for length of hospital stay (in days). Displaying the observed percentiles for this outcome.

| | 2.5 | 10.0 | 25.0 | 50.0 | 75.0 | 90.0 | 97.5 |
|---|---|---|---|---|---|---|---|
| Interferon-beta-1a | 1.92 | 8.8 | 14.16 | 58.18 | - | - | - |
| Anakinra | 3.94 | 7.07 | 12.92 | 51.14 | - | - | - |
| Tocilizumab | 3.75 | 7.03 | 11.29 | 32.11 | - | - | - |
| Sarilumab | 4.18 | 6.45 | 10.75 | 33.22 | - | - | - |
| Control | 4.89 | 7.36 | 13.26 | 38.11 | - | - | - |

Table 61: Hazard ratio parameters for secondary analysis of length of hospital stay

| | Mean | SD | Median | CrI |
|---|---|---|---|---|
| Interferon-beta-1a | 1.18 | 0.34 | 1.15 | (0.62, 1.94) |
| Anakinra | 1.05 | 0.12 | 1.04 | (0.84, 1.30) |
| Tocilizumab | 1.31 | 0.11 | 1.31 | (1.11, 1.55) |
| Sarilumab | 1.31 | 0.14 | 1.30 | (1.06, 1.59) |
| Age<39 | 3.16 | 0.33 | 3.15 | (2.57, 3.84) |
| Age 40-49 | 1.90 | 0.16 | 1.89 | (1.60, 2.23) |
| Age 50-59 | 1.81 | 0.13 | 1.81 | (1.57, 2.07) |
| Age 70-79 | 0.70 | 0.06 | 0.70 | (0.59, 0.83) |
| Age 80+ | 0.44 | 0.08 | 0.43 | (0.29, 0.62) |
| Female | 1.22 | 0.07 | 1.22 | (1.09, 1.37) |

Table 62: Posterior probabilities for secondary analysis of length of hospital stay

| | Posterior Probability |
|---|---|
| Interferon-beta-1a is optimal | 0.292 |
| Interferon-beta-1a is superior to control | 0.686 |
| Interferon-beta-1a is futile (OR < 1.2) | 0.558 |
| Interferon-beta-1a is harmful (OR < 1) | 0.314 |
| Anakinra is optimal | 0.001 |
| Anakinra is superior to control | 0.647 |
| Anakinra is futile (OR < 1.2) | 0.897 |
| Anakinra is harmful (OR < 1) | 0.352 |
| Tocilizumab is optimal | 0.373 |
| Tocilizumab is superior to control | 1.000 |
| Tocilizumab is futile (OR < 1.2) | 0.156 |
| Tocilizumab is harmful (OR < 1) | 0.001 |
| Sarilumab is optimal | 0.334 |
| Sarilumab is superior to control | 0.994 |
| Sarilumab is futile (OR < 1.2) | 0.224 |
| Sarilumab is harmful (OR < 1) | 0.006 |
| Interferon-beta-1a equivalent to anakinra | 0.430 |
| Interferon-beta-1a equivalent to tocilizumab | 0.446 |
| Interferon-beta-1a equivalent to sarilumab | 0.447 |
| Anakinra equivalent to tocilizumab | 0.301 |
| Anakinra equivalent to sarilumab | 0.348 |
| Tocilizumab equivalent to sarilumab | 0.985 |
| Anakinra superior to tocilizumab | 0.005 |
| Anakinra superior to sarilumab | 0.008 |
| Tocilizumab superior to sarilumab | 0.539 |

#### 6.7 Secondary analysis of WHO scale at 14 days

- Model: Primary analysis ordinal model
- Factors: Age, sex, site, time, Immune Modulation Therapy Domain interventions (control, interferon-beta-1a, anakinra, tocilizumab, and sarilumab), COVID-19 Antiviral domain interventions (no antiviral, lopinavir−ritonavir, hydroxychloroquine, combination therapy), Corticosteroid domain interventions (no corticosteroid, fixed-duration corticosteroids, shock-dependent corticosteroids), Immunoglobulin domain interventions (no convalescent plasma, convalescent plasma, delayed convalescent plasma), and Anticoagulation interventions (usual care thromboprophylaxis, therapeutic anticoagulation)
- Treatment effects for tocilizumab and sarilumab are modeled with independent priors.
- There are 2 missing values of WHO scale at day 14 due to missing daily data. These patients are removed from this analysis.

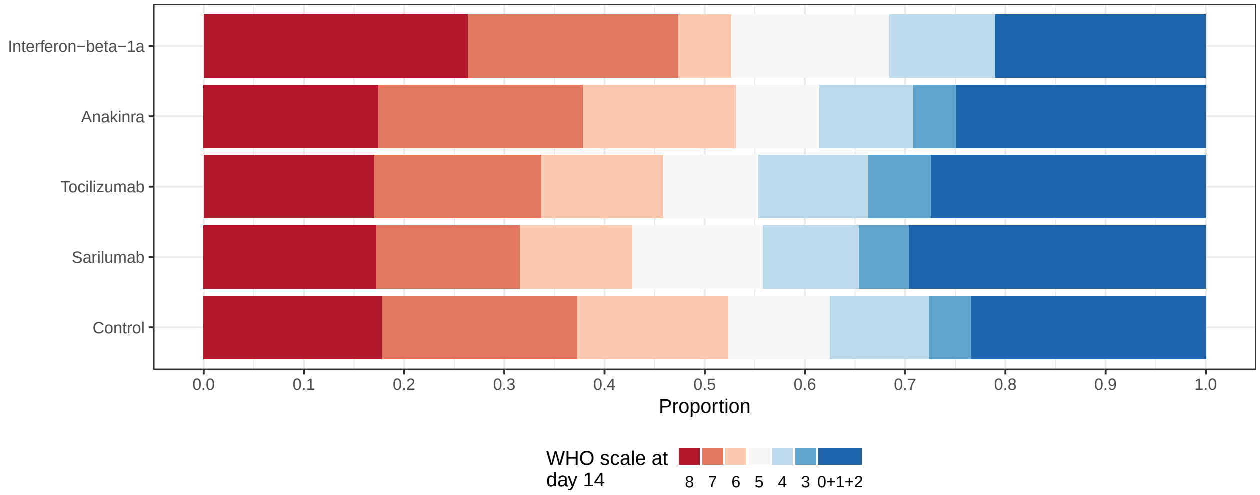

Figure 18: Empirical distribution of modified WHO scale at day 14 for interferon-beta-1a, anakinra, tocilizumab, sarilumab, and control. This plot is restricted to the Immune Modulation ITT population.

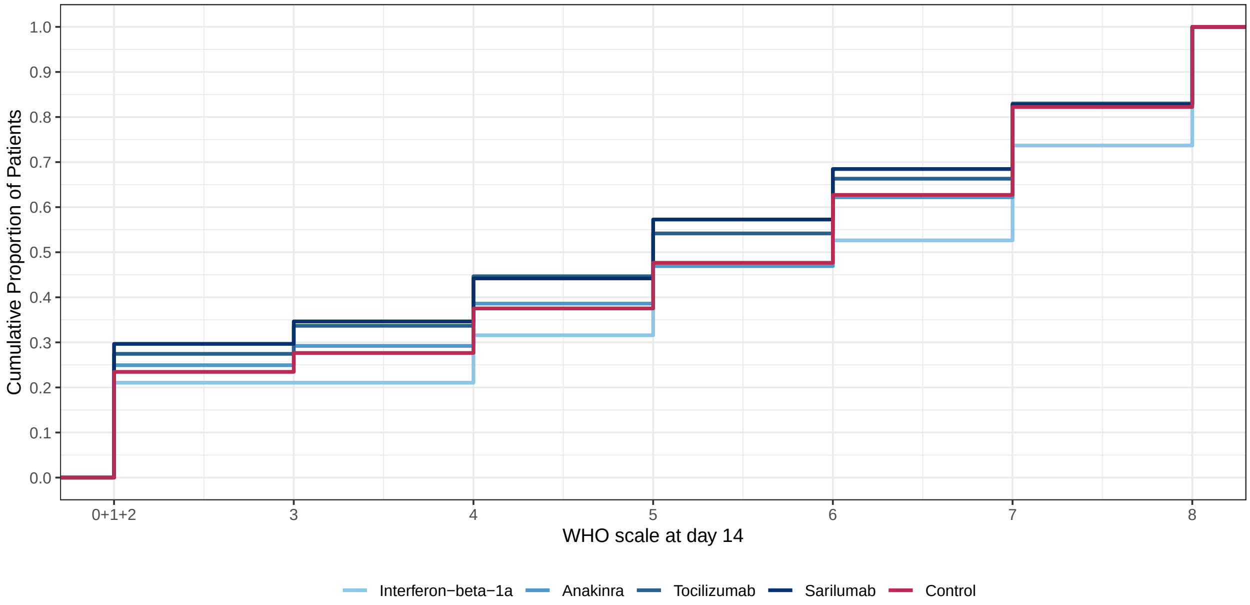

Figure 19: Empirical cumulative distribution of WHO scale for interferon-beta-1a, anakinra, tocilizumab, sarilumab, and control. This plot is restricted to the Immune Modulation ITT population.

Table 63: Odds ratio parameters for secondary analysis of WHO scale at 14 days

| | Mean | SD | Median | CrI |
|---|---|---|---|---|
| Interferon-beta-1a | 1.22 | 0.50 | 1.13 | (0.52, 2.46) |
| Anakinra | 1.19 | 0.18 | 1.18 | (0.87, 1.60) |
| Tocilizumab | 1.54 | 0.19 | 1.53 | (1.20, 1.95) |
| Sarilumab | 1.67 | 0.24 | 1.65 | (1.25, 2.18) |
| Lopinavir−ritonavir | 0.88 | 0.12 | 0.88 | (0.67, 1.15) |
| Hydroxychloroquine | 0.79 | 0.15 | 0.78 | (0.52, 1.10) |
| Fixed-dose corticosteroids | 1.34 | 0.29 | 1.31 | (0.86, 2.00) |
| Shock-dependent corticosteroids | 0.99 | 0.22 | 0.96 | (0.63, 1.49) |
| Therapeutic anticogulation | 0.79 | 0.09 | 0.78 | (0.62, 0.98) |
| Convalescent plasma | 0.92 | 0.08 | 0.92 | (0.78, 1.08) |
| Delayed convalescent plasma | 0.71 | 0.38 | 0.63 | (0.23, 1.66) |
| Lopinavir−ritonavir*Tocilizumab combination | 1.38 | 0.26 | 1.36 | (0.95, 1.95) |
| Hydroxychloroquine*Tocilizumab combination | 1.21 | 0.28 | 1.19 | (0.74, 1.81) |
| Tocilizumab*Fixed-dose corticosteroids combination | 2.06 | 0.51 | 2.00 | (1.23, 3.26) |
| Lopinavir−ritonavir*Tocilizumab interaction | 1.01 | 0.05 | 1.01 | (0.92, 1.12) |
| Tocilizumab*Fixed-dose corticosteroids interaction | 1.00 | 0.05 | 1.00 | (0.91, 1.11) |
| Hydroxychloroquine*Tocilizumab interaction | 1.00 | 0.05 | 1.00 | (0.90, 1.10) |
| Age<39 | 4.35 | 0.58 | 4.31 | (3.35, 5.58) |
| Age 40-49 | 2.31 | 0.22 | 2.29 | (1.92, 2.78) |
| Age 50-59 | 1.74 | 0.14 | 1.74 | (1.49, 2.03) |
| Age 70-79 | 0.55 | 0.05 | 0.54 | (0.46, 0.64) |
| Age 80+ | 0.27 | 0.04 | 0.27 | (0.19, 0.37) |
| Female | 1.05 | 0.07 | 1.05 | (0.92, 1.19) |

Table 64: Posterior probabilities for secondary analysis of WHO scale at 14 days

| | Posterior Probability |
|---|---|
| Interferon-beta-1a is optimal | 0.160 |
| Interferon-beta-1a is superior to control | 0.620 |
| Interferon-beta-1a is futile (OR < 1.2) | 0.556 |
| Interferon-beta-1a is harmful (OR < 1) | 0.380 |
| Anakinra is optimal | 0.002 |
| Anakinra is superior to control | 0.856 |
| Anakinra is futile (OR < 1.2) | 0.545 |
| Anakinra is harmful (OR < 1) | 0.144 |
| Tocilizumab is optimal | 0.206 |
| Tocilizumab is superior to control | 1.000 |
| Tocilizumab is futile (OR < 1.2) | 0.025 |
| Tocilizumab is harmful (OR < 1) | 0.000 |
| Sarilumab is optimal | 0.632 |
| Sarilumab is superior to control | 1.000 |
| Sarilumab is futile (OR < 1.2) | 0.013 |
| Sarilumab is harmful (OR < 1) | 0.000 |
| Lopinavir−ritonavir*tocilizumab combination OR > 1 | 0.949 |
| Hydroxychloroquine*tocilizumab combination OR > 1 | 0.776 |
| Fixed-dose steroids*tocilizumab combination OR > 1 | 0.998 |
| Interferon-beta-1a equivalent to anakinra | 0.347 |
| Interferon-beta-1a equivalent to tocilizumab | 0.276 |
| Interferon-beta-1a equivalent to sarilumab | 0.233 |
| Anakinra equivalent to tocilizumab | 0.273 |
| Anakinra equivalent to sarilumab | 0.125 |
| Tocilizumab equivalent to sarilumab | 0.802 |
| Anakinra superior to tocilizumab | 0.021 |
| Anakinra superior to sarilumab | 0.006 |
| Tocilizumab superior to sarilumab | 0.260 |

#### 6.8 Secondary analysis of major thrombotic events or death

Table 65: Summary of major thrombotic events or death by intervention in the Immune Modulation ITT population. Note that this table displays the number of patients with one or more major thrombotic events rather than the total number of major thrombotic events.

| Intervention | Patients (N) | Known (n) | Death | Major thrombotic event | Death or major thrombotic event (y) | Death or major thrombotic event rate (y/n) |
|---|---|---|---|---|---|---|
| Overall | 2226 | 1137 | 409 | 97 | 456 | 0.401 |
| Interferon-beta-1a | 19 | 14 | 5 | 0 | 5 | 0.357 |
| Anakinra | 373 | 97 | 45 | 6 | 48 | 0.495 |
| Tocilizumab | 947 | 505 | 174 | 40 | 192 | 0.380 |
| Sarilumab | 482 | 276 | 91 | 26 | 104 | 0.377 |
| Control | 405 | 245 | 94 | 25 | 107 | 0.437 |
| Pooled IL-6ra | 1429 | 781 | 265 | 66 | 296 | 0.379 |

Table 66: Summary of the number of major thrombotic events by intervention in the Immune Modulation ITT population

| Intervention | Patients | Known | Patients with major thrombotic events | Total number of major thrombotic events |
|---|---|---|---|---|
| Overall | 2226 | 1138 | 97 | 106 |
| Interferon-beta-1a | 19 | 14 | 0 | 0 |
| Anakinra | 373 | 97 | 6 | 8 |
| Tocilizumab | 947 | 506 | 40 | 42 |
| Sarilumab | 482 | 276 | 26 | 31 |
| Control | 405 | 245 | 25 | 25 |

Table 67: Summary of the components of major thrombotic events by intervention in the Immune Modulation ITT population

| Intervention | # Major thrombotic events | Pulmonary embolism | Myocardial infarction | Ischemic cerebrovascular event | Systemic arterial thromboembolism |
|---|---|---|---|---|---|
| Overall | 106 | 61 | 16 | 21 | 8 |
| Interferon-beta-1a | 0 | 0 | 0 | 0 | 0 |
| Anakinra | 8 | 4 | 2 | 2 | 0 |
| Tocilizumab | 42 | 20 | 8 | 12 | 2 |
| Sarilumab | 31 | 19 | 6 | 3 | 3 |
| Control | 25 | 18 | 0 | 4 | 3 |

- Model: Primary dichotomous model
- Factors: Age, sex, site, time, Immune Modulation Therapy Domain interventions (control, interferon-beta-1a, anakinra, tocilizumab, and sarilumab)
- Treatment effects for tocilizumab and sarilumab are pooled into a single IL-6ra effect.

Table 68: Odds ratio parameters for the secondary analysis of freedom from major thrombotic events and death

| | Mean | SD | Median | CrI |
|---|---|---|---|---|
| Interferon-beta-1a | 1.63 | 1.00 | 1.38 | (0.51, 4.17) |
| Anakinra | 1.01 | 0.29 | 0.97 | (0.57, 1.69) |
| Pooled IL-6ra | 1.78 | 0.34 | 1.75 | (1.22, 2.55) |
| Age<39 | 8.51 | 3.63 | 7.75 | (3.76, 17.65) |
| Age 40-49 | 3.18 | 0.76 | 3.08 | (1.97, 4.91) |
| Age 50-59 | 2.88 | 0.52 | 2.82 | (2.01, 3.99) |
| Age 70-79 | 0.54 | 0.10 | 0.53 | (0.37, 0.75) |
| Age 80+ | 0.28 | 0.09 | 0.26 | (0.14, 0.48) |
| Female | 1.36 | 0.21 | 1.35 | (1.00, 1.80) |

Table 69: Posterior probabilities for the secondary analysis of freedom from major thrombotic events and death

| | Posterior Probability |
|---|---|
| Interferon-beta-1a is optimal | 0.335 |
| Interferon-beta-1a is superior to control | 0.730 |
| Interferon-beta-1a is futile (OR < 1.2) | 0.392 |
| Interferon-beta-1a is harmful (OR < 1) | 0.270 |
| Anakinra is optimal | 0.005 |
| Anakinra is superior to control | 0.460 |
| Anakinra is futile (OR < 1.2) | 0.772 |
| Anakinra is harmful (OR < 1) | 0.540 |
| Pooled IL-6ra is optimal | 0.659 |
| Pooled IL-6ra is superior to control | 0.999 |
| Pooled IL-6ra is futile (OR < 1.2) | 0.021 |
| Pooled IL-6ra is harmful (OR < 1) | 0.001 |
| Interferon-beta-1a equivalent to anakinra | 0.203 |
| Interferon-beta-1a equivalent to pooled IL-6ra | 0.236 |
| Anakinra equivalent to pooled IL-6ra | 0.046 |
| Anakinra superior to pooled IL-6ra | 0.007 |

#### 6.9 Secondary analysis of major thrombotic events

Table 70: Summary of major thrombotic events by intervention in the Immune Modulation ITT population. Note that this table displays the number of patients with one or more major thrombotic events rather than the total number of major thrombotic events.

| Intervention | Patients (N) | Known (n) | Major thrombotic event | Major thrombotic event rate (y/n) |
|---|---|---|---|---|
| Overall | 2226 | 1138 | 97 | 0.085 |
| Interferon-beta-1a | 19 | 14 | 0 | 0.000 |
| Anakinra | 373 | 97 | 6 | 0.062 |
| Tocilizumab | 947 | 506 | 40 | 0.079 |
| Sarilumab | 482 | 276 | 26 | 0.094 |
| Control | 405 | 245 | 25 | 0.102 |
| Pooled IL-6ra | 1429 | 782 | 66 | 0.084 |

- Model: Primary dichotomous model

- Factors: Age, sex, site, time, Immune Modulation Therapy Domain interventions (control, interferon-beta-1a, anakinra, tocilizumab, and sarilumab)
- Treatment effects for tocilizumab and sarilumab are pooled into a single IL-6ra effect.

Table 71: Odds ratio parameters for the secondary analysis of freedom from major thrombotic events

| | Mean | SD | Median | CrI |
|---|---|---|---|---|
| Interferon-beta-1a | 3.07 | 3.07 | 2.16 | (0.52, 11.15) |
| Anakinra | 1.79 | 0.91 | 1.58 | (0.66, 4.10) |
| Pooled IL-6ra | 1.24 | 0.37 | 1.19 | (0.68, 2.09) |
| Age<39 | 2.83 | 1.80 | 2.40 | (0.92, 7.18) |
| Age 40-49 | 1.40 | 0.51 | 1.31 | (0.67, 2.66) |
| Age 50-59 | 1.83 | 0.55 | 1.75 | (1.00, 3.14) |
| Age 70-79 | 1.01 | 0.29 | 0.97 | (0.57, 1.70) |
| Age 80+ | 2.93 | 2.22 | 2.35 | (0.79, 8.49) |
| Female | 1.10 | 0.27 | 1.07 | (0.67, 1.71) |

Table 72: Posterior probabilities for the secondary analysis of freedom from major thrombotic events

| | Posterior Probability |
|---|---|
| Interferon-beta-1a is optimal | 0.612 |
| Interferon-beta-1a is superior to control | 0.843 |
| Interferon-beta-1a is futile (OR < 1.2) | 0.220 |
| Interferon-beta-1a is harmful (OR < 1) | 0.157 |
| Anakinra is optimal | 0.312 |
| Anakinra is superior to control | 0.844 |
| Anakinra is futile (OR < 1.2) | 0.272 |
| Anakinra is harmful (OR < 1) | 0.156 |
| Pooled IL-6ra is optimal | 0.057 |
| Pooled IL-6ra is superior to control | 0.729 |
| Pooled IL-6ra is futile (OR < 1.2) | 0.513 |
| Pooled IL-6ra is harmful (OR < 1) | 0.271 |
| Interferon-beta-1a equivalent to anakinra | 0.157 |
| Interferon-beta-1a equivalent to pooled IL-6ra | 0.145 |
| Anakinra equivalent to pooled IL-6ra | 0.277 |
| Anakinra superior to pooled IL-6ra | 0.745 |

#### 6.10 Secondary analysis of major bleeding events

Table 73: Summary of major bleeds by intervention for the Immune Modulation ITT population

| Intervention | Patients (N) | Known (n) | Patients with Major Bleeds (y) | Major Bleed Rate (y/n) |
|---|---|---|---|---|
| Overall | 2226 | 458 | 8 | 0.017 |
| Interferon-beta-1a | 19 | 10 | 1 | 0.100 |
| Anakinra | 373 | 37 | 1 | 0.027 |
| Tocilizumab | 947 | 170 | 4 | 0.024 |
| Sarilumab | 482 | 93 | 0 | 0.000 |
| Control | 405 | 148 | 2 | 0.014 |
| Pooled IL-6ra | 1429 | 263 | 4 | 0.015 |

- Model: Primary dichotomous model
- Factors: Age, sex, Immune Modulation Therapy Domain interventions (control, interferon-beta-1a, anakinra, tocilizumab, and sarilumab)
- Treatment effects for tocilizumab and sarilumab are pooled into a single IL-6ra effect.
- Due to the small number of major bleeding events and concerns about model stability, site and time factors were removed from this model.

Table 74: Odds ratio parameters for the secondary analysis of freedom from major bleeding events

| | Mean | SD | Median | CrI |
|---|---|---|---|---|
| Interferon-beta-1a | 0.88 | 0.96 | 0.59 | (0.12, 3.47) |
| Anakinra | 1.31 | 1.26 | 0.96 | (0.23, 4.44) |
| Pooled IL-6ra | 1.46 | 0.93 | 1.24 | (0.39, 3.89) |
| Age<39 | 8.35 | 10.10 | 5.25 | (0.86, 34.70) |
| Age 40-49 | 1.88 | 1.70 | 1.39 | (0.34, 6.41) |
| Age 50-59 | 5.26 | 5.37 | 3.71 | (0.85, 19.63) |
| Age 70-79 | 0.79 | 0.57 | 0.64 | (0.19, 2.30) |
| Age 80+ | 0.78 | 0.85 | 0.54 | (0.13, 2.87) |
| Female | 0.49 | 0.33 | 0.41 | (0.13, 1.32) |

Table 75: Posterior probabilities for the secondary analysis of freedom from major bleeding events

| | Posterior Probability |
|---|---|
| Interferon-beta-1a is optimal | 0.134 |
| Interferon-beta-1a is superior to control | 0.270 |
| Interferon-beta-1a is harmful (OR < 1) | 0.730 |
| Anakinra is optimal | 0.280 |
| Anakinra is superior to control | 0.474 |
| Anakinra is harmful (OR < 1) | 0.526 |
| Pooled IL-6ra is optimal | 0.414 |
| Pooled IL-6ra is superior to control | 0.637 |
| Pooled IL-6ra is harmful (OR < 1) | 0.363 |
| Interferon-beta-1a equivalent to anakinra | 0.123 |
| Interferon-beta-1a equivalent to pooled IL-6ra | 0.112 |
| Anakinra equivalent to pooled IL-6ra | 0.159 |
| Anakinra superior to pooled IL-6ra | 0.386 |

### 7 Subgroup analyses

#### 7.1 Subgroup analyses by baseline C-reactive protein subgroup

In this section, subgroup analyses are performed by baseline C-reactive protein (CRP) subgroup. The CRP subgroups are defined based on terciles of CRP. Patients with CRP lower than 85 $\mu$g/ml are included in the lowest CRP subgroup. Patients with CRP greater/equal to 85 and less than 169 $\mu$g/ml are included in the middle CRP subgroup. Patients with CRP greater or equal to 169 $\mu$g/ml are included in the highest CRP subgroup. Patients with unknown CRP are included in a fourth "unknown" subgroup.

##### 7.1.1 Data summaries of baseline CRP in the Immune Modulation ITT population

Table 76: Summary of baseline CRP subgroup by intervention

| Intervention | Patients | Lower CRP tercile | Middle CRP tercile | Upper CRP tercile | Unknown CRP |
|---|---|---|---|---|---|
| Interferon-beta-1a | 19 | 6 (31.6%) | 5 (26.3%) | 3 (15.8%) | 5 (26.3%) |
| Anakinra | 373 | 110 (29.5%) | 118 (31.6%) | 96 (25.7%) | 49 (13.1%) |
| Tocilizumab | 947 | 237 (25%) | 261 (27.6%) | 280 (29.6%) | 169 (17.8%) |
| Sarilumab | 482 | 135 (28%) | 139 (28.8%) | 142 (29.5%) | 66 (13.7%) |
| Control | 405 | 78 (19.3%) | 85 (21%) | 91 (22.5%) | 151 (37.3%) |
| Pooled IL-6ra | 1429 | 372 (26%) | 400 (28%) | 422 (29.5%) | 235 (16.4%) |
| Overall | 2226 | 566 (25.4%) | 608 (27.3%) | 612 (27.5%) | 440 (19.8%) |

Table 77: Summary of subjects by baseline CRP subgroup in the Immune Modulation ITT population

| Intervention | Patients | Known | Deaths (%) | OSFD median (IQR) | OSFD in survivors* median (IQR) |
|---|---|---|---|---|---|
| **Lower tercile of CRP** | | | | | |
| Interferon-beta-1a | 6 | 6 | 2 (33.3%) | 4.5 (-0.75, 10.5) | 10 (6.75, 12.75) |
| Anakinra | 110 | 109 | 42 (38.5%) | 1 (-1, 17) | 15 (3, 19) |
| Tocilizumab | 237 | 235 | 79 (33.6%) | 10 (-1, 18) | 17 (10, 19) |
| Sarilumab | 135 | 135 | 45 (33.3%) | 11 (-1, 17) | 16 (11.25, 18) |
| Control | 78 | 78 | 30 (38.5%) | 10 (-1, 17) | 16.5 (11.75, 18.25) |
| Pooled IL-6ra | 372 | 370 | 124 (33.5%) | 10 (-1, 18) | 17 (10, 19) |
| **Middle tercile of CRP** | | | | | |
| Interferon-beta-1a | 5 | 5 | 3 (60%) | -1 (-1, 11) | 15.5 (13.25, 17.75) |
| Anakinra | 118 | 116 | 45 (38.8%) | 0 (-1, 14) | 13 (5.5, 17) |
| Tocilizumab | 261 | 260 | 83 (31.9%) | 8.5 (-1, 16) | 14 (8, 18) |
| Sarilumab | 139 | 138 | 43 (31.2%) | 9.5 (-1, 17) | 15 (8.5, 18) |
| Control | 85 | 85 | 29 (34.1%) | 9 (-1, 15) | 14 (9, 17) |
| Pooled IL-6ra | 400 | 398 | 126 (31.7%) | 9 (-1, 16) | 14.5 (8, 18) |
| **Upper tercile of CRP** | | | | | |
| Interferon-beta-1a | 3 | 3 | 0 (0%) | 16 (8, 16.5) | 16 (8, 16.5) |
| Anakinra | 96 | 94 | 40 (42.6%) | 0 (-1, 14) | 13 (0.5, 16.75) |
| Tocilizumab | 280 | 278 | 103 (37.1%) | 1.5 (-1, 16) | 14 (4.5, 18) |
| Sarilumab | 142 | 141 | 48 (34%) | 8 (-1, 16) | 15 (9, 17) |
| Control | 91 | 91 | 32 (35.2%) | 0 (-1, 14) | 12 (0, 16.5) |
| Pooled IL-6ra | 422 | 419 | 151 (36%) | 3 (-1, 16) | 14 (7, 17) |
| **Unknown CRP** | | | | | |
| Interferon-beta-1a | 5 | 5 | 2 (40%) | 0 (-1, 0) | 0 (0, 10) |
| Anakinra | 49 | 46 | 18 (39.1%) | 3 (-1, 14.75) | 13.5 (7.75, 17.25) |
| Tocilizumab | 169 | 165 | 52 (31.5%) | 7 (-1, 15) | 14 (6, 17) |
| Sarilumab | 66 | 66 | 21 (31.8%) | 7 (-1, 16.75) | 14 (7, 18) |
| Control | 151 | 151 | 59 (39.1%) | 0 (-1, 13.5) | 11 (0, 17) |
| Pooled IL-6ra | 235 | 231 | 73 (31.6%) | 7 (-1, 15.5) | 14 (6, 18) |

* Days free of organ support in survivors within 21 days

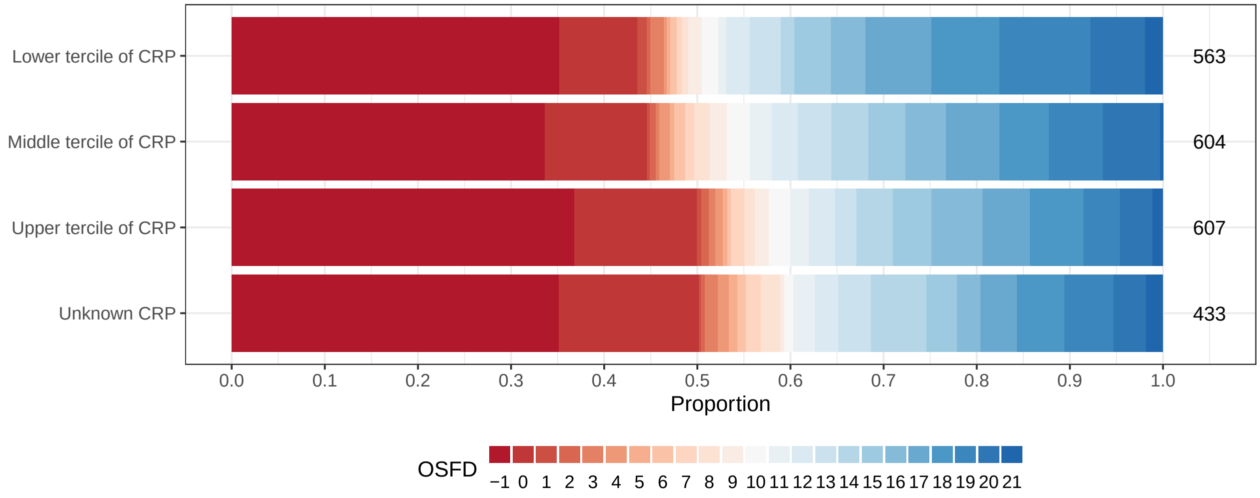

Figure 20: Empirical distribution of organ support free days (OSFD) by baseline CRP subgroup. This plot is restricted to the Immune Modulation ITT population.

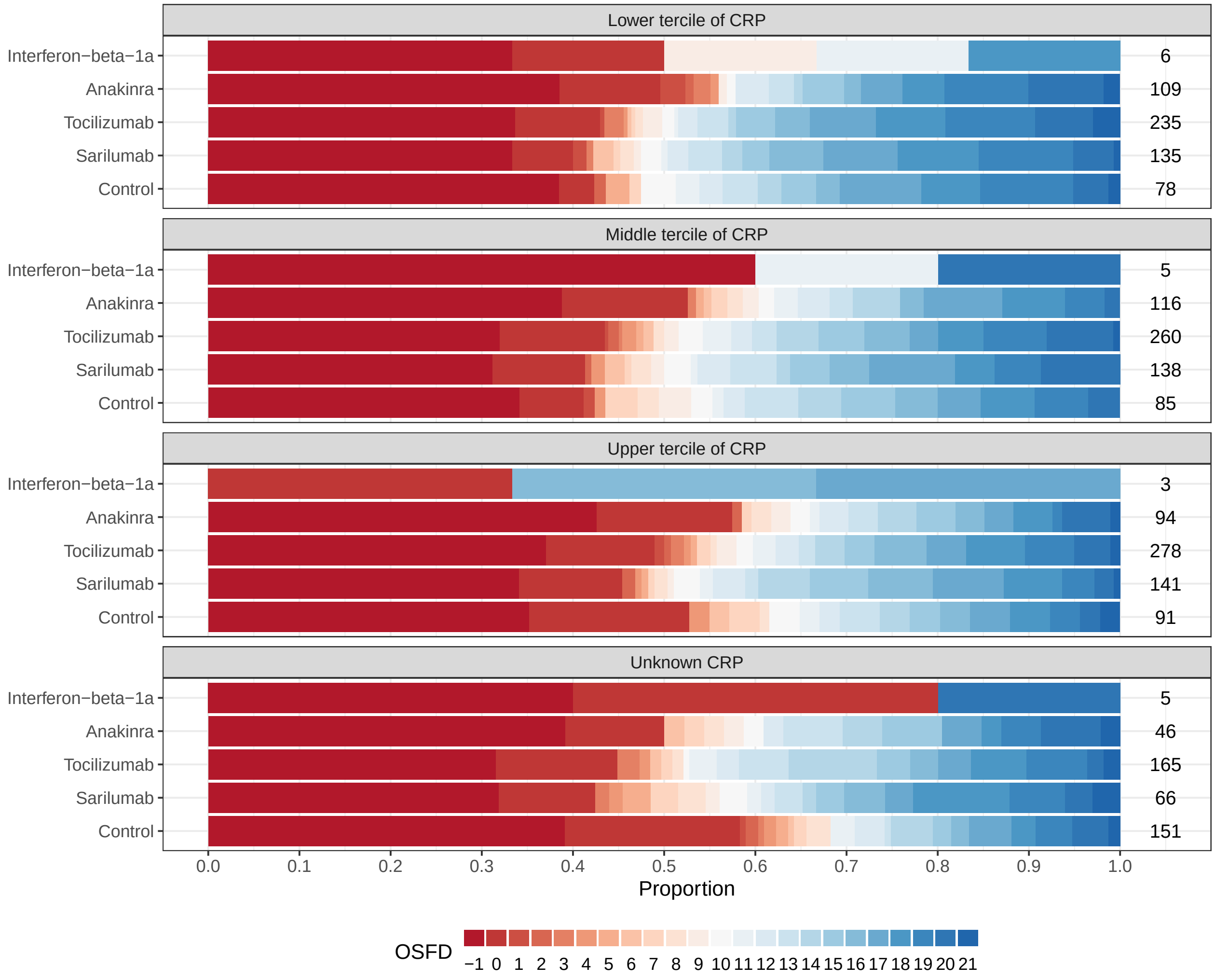

Figure 21: Empirical distribution of organ support free days (OSFD) for interferon-beta-1a, anakinra, tocilizumab, sarilumab, and control by baseline CRP subgroup. This plot is restricted to the Immune Modulation ITT population.

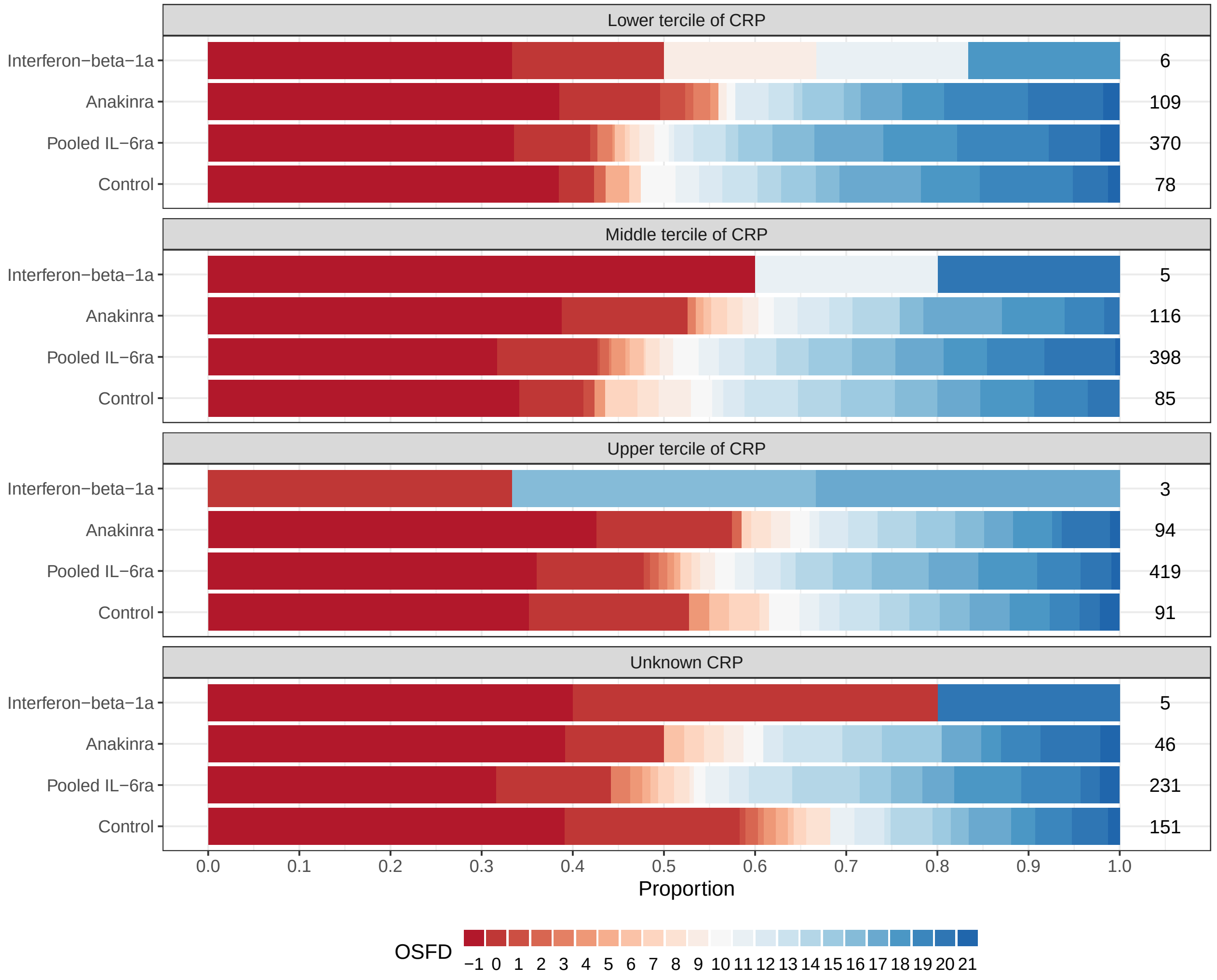

Figure 22: Empirical distribution of organ support free days (OSFD) for pooled IL-6ra and control interventions by baseline CRP subgroup. This plot is restricted to the Immune Modulation ITT population.

##### 7.1.2 Subgroup analysis with differential effect on OSFD by baseline CRP subgroup

- Model: Primary analysis ordinal model
- Factors: Age, sex, site, time, Immune Modulation Therapy Domain interventions (control, interferon-beta-1a, anakinra, tocilizumab, and sarilumab), COVID-19 Antiviral domain interventions (no antiviral, lopinavir−ritonavir, hydroxychloroquine, combination therapy), Corticosteroid domain interventions (no corticosteroid, fixed-duration corticosteroids, shock-dependent corticosteroids), Immunoglobulin domain interventions (no convalescent plasma, convalescent plasma, delayed convalescent plasma), and Anticoagulation interventions (usual care thromboprophylaxis, therapeutic anticoagulation)

- Treatment effects for tocilizumab and sarilumab are modeled with independent priors.
- Differential treatment effects are modeled by baseline CRP tercile (low/middle/high/unknown). Due to small sample sizes, a differential effect by CRP subgroup is not estimated for Interferon-beta-1a.

Table 78: Odds ratio parameters for subgroup analysis with differential effect on OSFD by baseline CRP subgroup

| | Mean | SD | Median | CrI |
|---|---|---|---|---|
| Interferon-beta-1a | 1.24 | 0.52 | 1.14 | (0.52, 2.47) |
| Anakinra, lower CRP tercile | 1.04 | 0.25 | 1.01 | (0.64, 1.60) |
| Anakinra, middle CRP tercile | 0.92 | 0.20 | 0.90 | (0.58, 1.38) |
| Anakinra, upper CRP tercile | 1.34 | 0.32 | 1.30 | (0.83, 2.05) |
| Anakinra, unknown CRP | 0.94 | 0.29 | 0.90 | (0.50, 1.64) |
| Tocilizumab, lower CRP tercile | 1.30 | 0.23 | 1.28 | (0.91, 1.82) |
| Tocilizumab, middle CRP tercile | 1.33 | 0.23 | 1.31 | (0.95, 1.83) |
| Tocilizumab, upper CRP tercile | 1.90 | 0.32 | 1.87 | (1.35, 2.59) |
| Tocilizumab, unknown CRP | 1.43 | 0.25 | 1.41 | (1.00, 1.99) |
| Sarilumab, lower CRP tercile | 1.42 | 0.30 | 1.39 | (0.93, 2.09) |
| Sarilumab, middle CRP tercile | 1.37 | 0.28 | 1.34 | (0.91, 2.01) |
| Sarilumab, upper CRP tercile | 1.88 | 0.38 | 1.85 | (1.24, 2.69) |
| Sarilumab, unknown CRP | 1.69 | 0.45 | 1.63 | (0.97, 2.71) |
| Age<39 | 4.24 | 0.52 | 4.21 | (3.31, 5.34) |
| Age 40-49 | 2.42 | 0.24 | 2.41 | (1.99, 2.93) |
| Age 50-59 | 1.84 | 0.15 | 1.83 | (1.56, 2.14) |
| Age 70-79 | 0.52 | 0.05 | 0.51 | (0.43, 0.62) |
| Age 80+ | 0.31 | 0.05 | 0.30 | (0.22, 0.43) |
| Female | 1.15 | 0.07 | 1.15 | (1.01, 1.30) |
| Middle tercile CRP subgroup (relative to low) | 0.96 | 0.12 | 0.95 | (0.74, 1.22) |
| Upper tercile CRP subgroup (relative to low) | 0.55 | 0.07 | 0.55 | (0.42, 0.70) |
| Unknown CRP subgroup (relative to low) | 0.68 | 0.10 | 0.68 | (0.51, 0.90) |

Table 79: Posterior probabilities for subgroup analysis with differential effect on OSFD by baseline CRP subgroup

| | Posterior Probability |
|---|---|
| Interferon-beta-1a is superior to control | 0.627 |
| Interferon-beta-1a is futile | 0.548 |
| Anakinra is superior to control in lower CRP tercile | 0.525 |
| Anakinra is futile in lower CRP tercile | 0.767 |
| Anakinra is superior to control in middle CRP tercile | 0.310 |
| Anakinra is futile in middle CRP tercile | 0.910 |
| Anakinra is superior to control in upper CRP tercile | 0.871 |
| Anakinra is futile in upper CRP tercile | 0.359 |
| Anakinra is superior to control with unknown CRP | 0.364 |

Table 79: Posterior probabilities for subgroup analysis with differential effect on OSFD by baseline CRP subgroup *(continued)*

| | Posterior Probability |
|---|---|
| Anakinra is futile with unknown CRP | 0.833 |
| Tocilizumab is superior to control in lower CRP tercile | 0.924 |
| Tocilizumab is futile in lower CRP tercile | 0.349 |
| Tocilizumab is superior to control in middle CRP tercile | 0.948 |
| Tocilizumab is futile in middle CRP tercile | 0.304 |
| Tocilizumab is superior to control in upper CRP tercile | 1.000 |
| Tocilizumab is futile in upper CRP tercile | 0.004 |
| Tocilizumab is superior to control with unknown CRP | 0.975 |
| Tocilizumab is futile with unknown CRP | 0.185 |
| Sarilumab is superior to control in lower CRP tercile | 0.944 |
| Sarilumab is futile in lower CRP tercile | 0.232 |
| Sarilumab is superior to control in middle CRP tercile | 0.933 |
| Sarilumab is futile in middle CRP tercile | 0.288 |
| Sarilumab is superior to control in upper CRP tercile | 0.998 |
| Sarilumab is futile in upper CRP tercile | 0.017 |
| Sarilumab is superior to control with unknown CRP | 0.966 |
| Sarilumab is futile with unknown CRP | 0.122 |

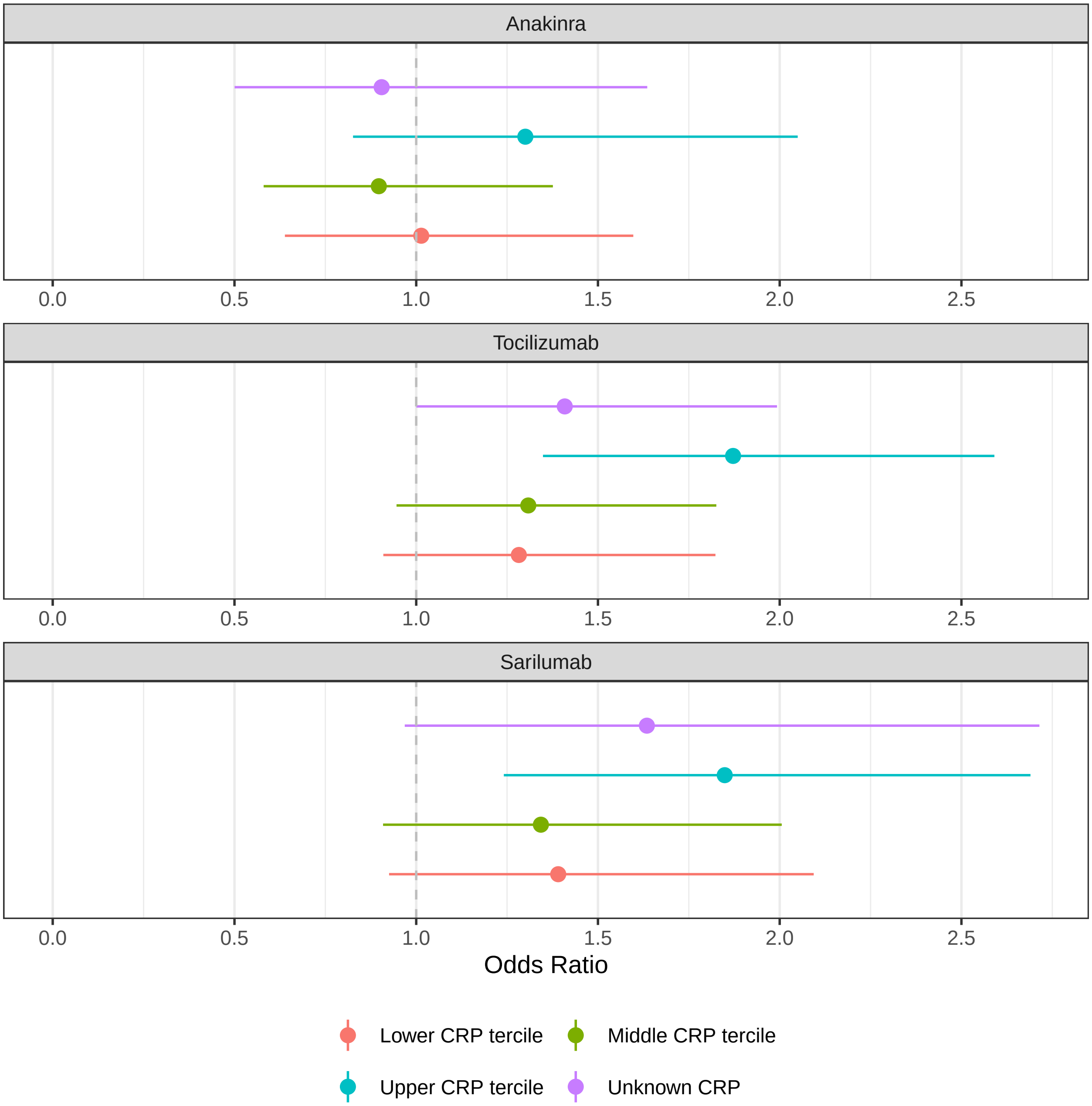

Figure 23: Estimated OSFD odds ratios for anakinra, tocilizumab, and sarilumab by CRP subgroup.

##### 7.1.3 Subgroup analysis with differential effect on hospital survival by baseline CRP subgroup

- Model: Primary dichotomous model
- Factors: Age, sex, site, time, Immune Modulation Therapy Domain interventions (control, interferon-beta-1a, anakinra, tocilizumab, and sarilumab), COVID-19 Antiviral domain interventions (no antiviral,

lopinavir–ritonavir, hydroxychloroquine, combination therapy), Corticosteroid domain interventions (no corticosteroid, fixed-duration corticosteroids, shock-dependent corticosteroids), Immunoglobulin domain interventions (no convalescent plasma, convalescent plasma, delayed convalescent plasma), and Anticoagulation interventions (usual care thromboprophylaxis, therapeutic anticoagulation)

- Treatment effects for tocilizumab and sarilumab are modeled with independent priors.
- Differential treatment effects are modeled by baseline CRP tercile (low/middle/high/unknown). Due to small sample sizes, a differential effect by CRP subgroup is not estimated for Interferon-beta-1a.

Table 80: Odds ratio parameters for subgroup analysis with differential effect on hospital survival by baseline CRP subgroup

| | Mean | SD | Median | CrI |
|---|---|---|---|---|
| Interferon-beta-1a | 1.32 | 0.70 | 1.17 | (0.46, 3.10) |
| Anakinra, lower CRP tercile | 0.85 | 0.24 | 0.82 | (0.48, 1.41) |
| Anakinra, middle CRP tercile | 0.99 | 0.27 | 0.95 | (0.57, 1.62) |
| Anakinra, upper CRP tercile | 1.23 | 0.35 | 1.18 | (0.68, 2.06) |
| Anakinra, unknown CRP | 0.94 | 0.36 | 0.89 | (0.43, 1.80) |
| Tocilizumab, lower CRP tercile | 1.09 | 0.24 | 1.07 | (0.70, 1.63) |
| Tocilizumab, middle CRP tercile | 1.32 | 0.29 | 1.29 | (0.85, 1.95) |
| Tocilizumab, upper CRP tercile | 1.80 | 0.38 | 1.76 | (1.18, 2.65) |
| Tocilizumab, unknown CRP | 1.45 | 0.34 | 1.41 | (0.90, 2.21) |
| Sarilumab, lower CRP tercile | 1.33 | 0.35 | 1.29 | (0.79, 2.15) |
| Sarilumab, middle CRP tercile | 1.38 | 0.36 | 1.34 | (0.81, 2.20) |
| Sarilumab, upper CRP tercile | 1.86 | 0.47 | 1.80 | (1.11, 2.95) |
| Sarilumab, unknown CRP | 1.76 | 0.60 | 1.66 | (0.89, 3.23) |
| Age<39 | 8.51 | 1.92 | 8.25 | (5.48, 12.90) |
| Age 40-49 | 3.56 | 0.50 | 3.52 | (2.69, 4.65) |
| Age 50-59 | 2.47 | 0.26 | 2.45 | (2.00, 3.02) |
| Age 70-79 | 0.47 | 0.05 | 0.47 | (0.38, 0.57) |
| Age 80+ | 0.26 | 0.05 | 0.26 | (0.18, 0.37) |
| Female | 1.26 | 0.11 | 1.26 | (1.07, 1.48) |
| Middle tercile CRP subgroup (relative to low) | 1.09 | 0.17 | 1.08 | (0.79, 1.48) |
| Upper tercile CRP subgroup (relative to low) | 0.66 | 0.10 | 0.65 | (0.48, 0.87) |
| Unknown CRP subgroup (relative to low) | 0.79 | 0.14 | 0.78 | (0.56, 1.10) |

Table 81: Posterior probabilities for subgroup analysis with differential effect on hospital survival by baseline CRP subgroup

| | Posterior Probability |
|---|---|
| Interferon-beta-1a is superior to control | 0.631 |
| Interferon-beta-1a is futile | 0.519 |
| Anakinra is superior to control in lower CRP tercile | 0.236 |

Table 81: Posterior probabilities for subgroup analysis with differential effect on hospital survival by baseline CRP subgroup *(continued)*

| | Posterior Probability |
|---|---|
| Anakinra is futile in lower CRP tercile | 0.915 |
| Anakinra is superior to control in middle CRP tercile | 0.433 |
| Anakinra is futile in middle CRP tercile | 0.793 |
| Anakinra is superior to control in upper CRP tercile | 0.728 |
| Anakinra is futile in upper CRP tercile | 0.520 |
| Anakinra is superior to control with unknown CRP | 0.365 |
| Anakinra is futile with unknown CRP | 0.802 |
| Tocilizumab is superior to control in lower CRP tercile | 0.624 |
| Tocilizumab is futile in lower CRP tercile | 0.706 |
| Tocilizumab is superior to control in middle CRP tercile | 0.876 |
| Tocilizumab is futile in middle CRP tercile | 0.373 |
| Tocilizumab is superior to control in upper CRP tercile | 0.997 |
| Tocilizumab is futile in upper CRP tercile | 0.030 |
| Tocilizumab is superior to control with unknown CRP | 0.935 |
| Tocilizumab is futile with unknown CRP | 0.238 |
| Sarilumab is superior to control in lower CRP tercile | 0.840 |
| Sarilumab is futile in lower CRP tercile | 0.386 |
| Sarilumab is superior to control in middle CRP tercile | 0.871 |
| Sarilumab is futile in middle CRP tercile | 0.329 |
| Sarilumab is superior to control in upper CRP tercile | 0.991 |
| Sarilumab is futile in upper CRP tercile | 0.049 |
| Sarilumab is superior to control with unknown CRP | 0.942 |
| Sarilumab is futile with unknown CRP | 0.158 |

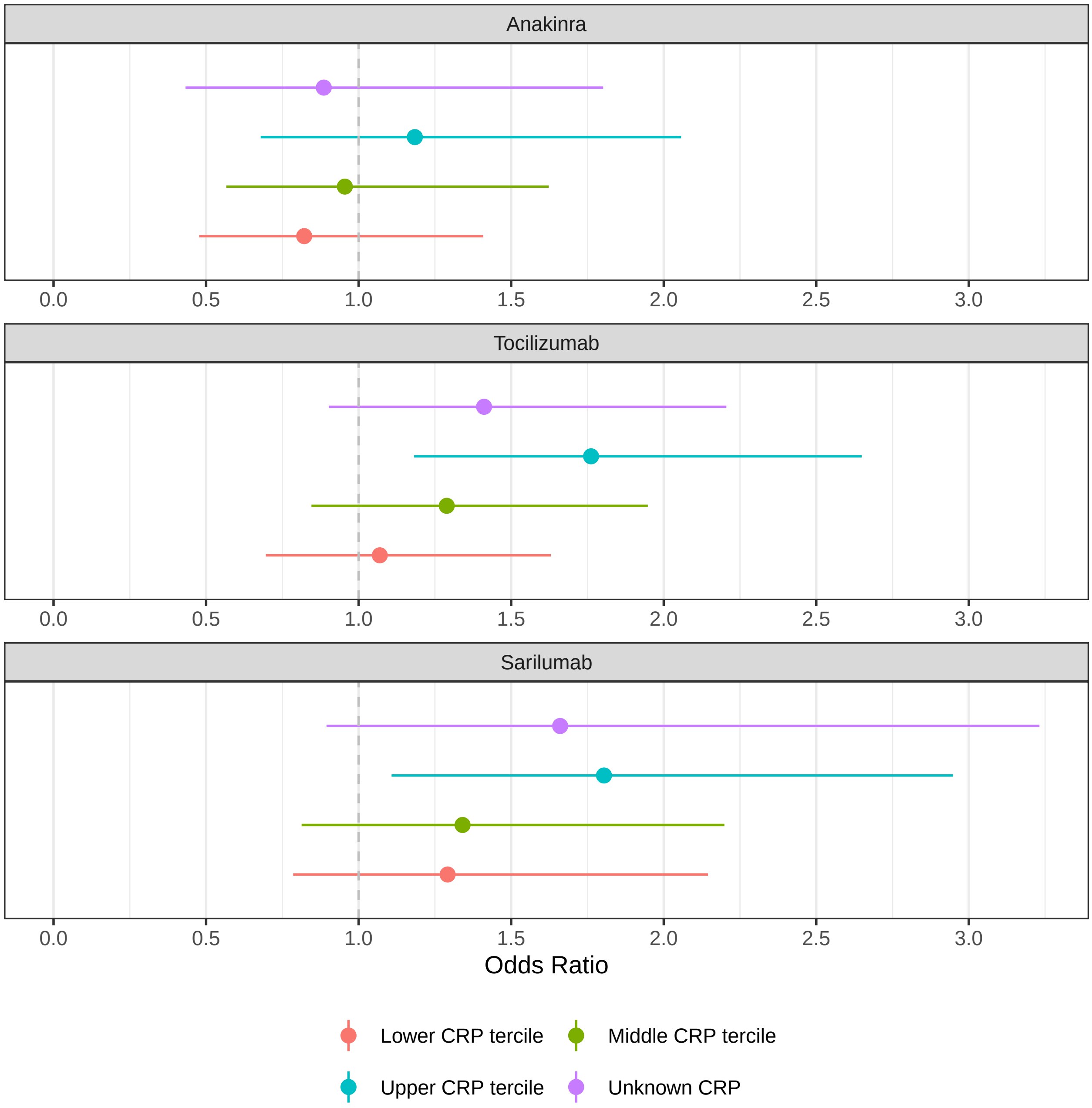

Figure 24: Estimated hospital survival odds ratios for anakinra, tocilizumab, and sarilumab by CRP subgroup.

#### 7.2 Subgroup analyses by baseline mechanical ventilation status

##### 7.2.1 Data summaries of baseline mechanical ventilation status in the Immune Modulation ITT population

Table 82: Summary of baseline invasive mechanical ventilation by intervention

| Intervention | Patients | No IMV at baseline | IMV at baseline |
|---|---|---|---|
| Interferon-beta-1a | 19 | 12 (63.2%) | 7 (36.8%) |
| Anakinra | 373 | 235 (63%) | 138 (37%) |
| Tocilizumab | 947 | 628 (66.3%) | 319 (33.7%) |
| Sarilumab | 482 | 336 (69.7%) | 146 (30.3%) |
| Control | 405 | 282 (69.6%) | 123 (30.4%) |
| Pooled IL-6ra | 1429 | 964 (67.5%) | 465 (32.5%) |
| Overall | 2226 | 1493 (67.1%) | 733 (32.9%) |

Table 83: Summary of subjects by baseline mechanical ventilation status in the Immune Modulation ITT population

| Intervention | Patients | Known | Deaths (%) | OSFD median (IQR) | OSFD in survivors* median (IQR) |
|---|---|---|---|---|---|
| **No invasive mechanical ventilation at baseline** | | | | | |
| Interferon-beta-1a | 12 | 12 | 3 (25%) | 10 (-0.25, 17.25) | 16 (9, 18) |
| Anakinra | 235 | 230 | 76 (33%) | 10 (-1, 17) | 16 (10, 18) |
| Tocilizumab | 628 | 621 | 175 (28.2%) | 13 (-1, 18) | 16 (11, 19) |
| Sarilumab | 336 | 335 | 103 (30.7%) | 13 (-1, 17) | 16 (12, 18) |
| Control | 282 | 282 | 90 (31.9%) | 7 (-1, 16.75) | 14 (6.75, 18) |
| Pooled IL-6ra | 964 | 956 | 278 (29.1%) | 13 (-1, 18) | 16 (12, 18) |
| **Invasive mechanical ventilation at baseline** | | | | | |
| Interferon-beta-1a | 7 | 7 | 4 (57.1%) | -1 (-1, 0) | 0 (0, 5.5) |
| Anakinra | 138 | 135 | 69 (51.1%) | -1 (-1, 7.5) | 8 (0, 13) |
| Tocilizumab | 319 | 317 | 142 (44.8%) | 0 (-1, 11) | 10 (0, 14) |
| Sarilumab | 146 | 145 | 54 (37.2%) | 0 (-1, 12) | 9 (0, 15) |
| Control | 123 | 123 | 60 (48.8%) | 0 (-1, 9.5) | 9 (0, 14) |
| Pooled IL-6ra | 465 | 462 | 196 (42.4%) | 0 (-1, 11) | 9.5 (0, 15) |

* Days free of organ support in survivors within 21 days

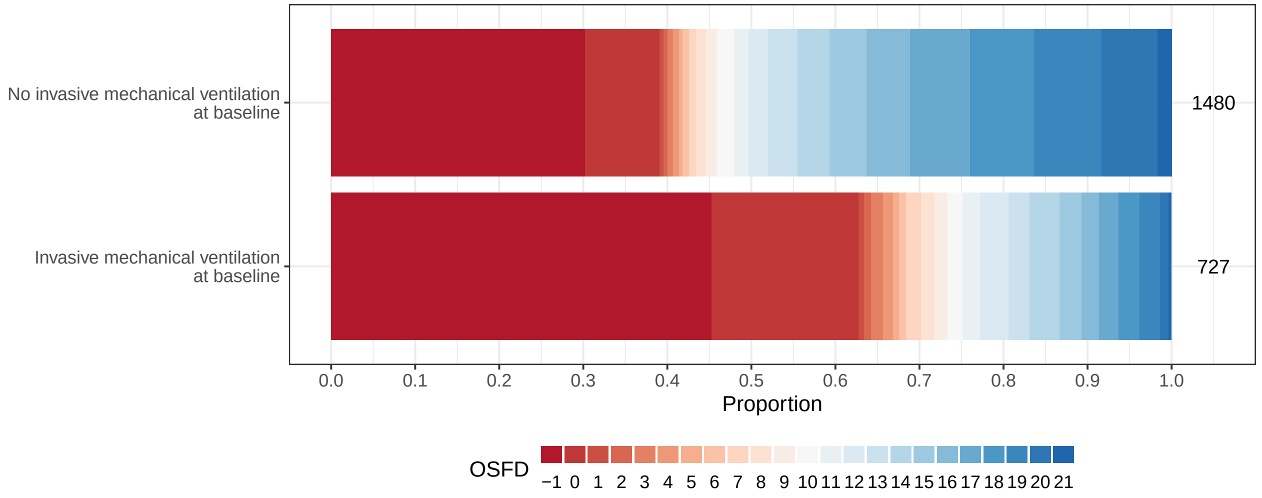

Figure 25: Empirical distribution of organ support free days (OSFD) by baseline invasive mechanical ventilation status. This plot is restricted to the Immune Modulation ITT population.

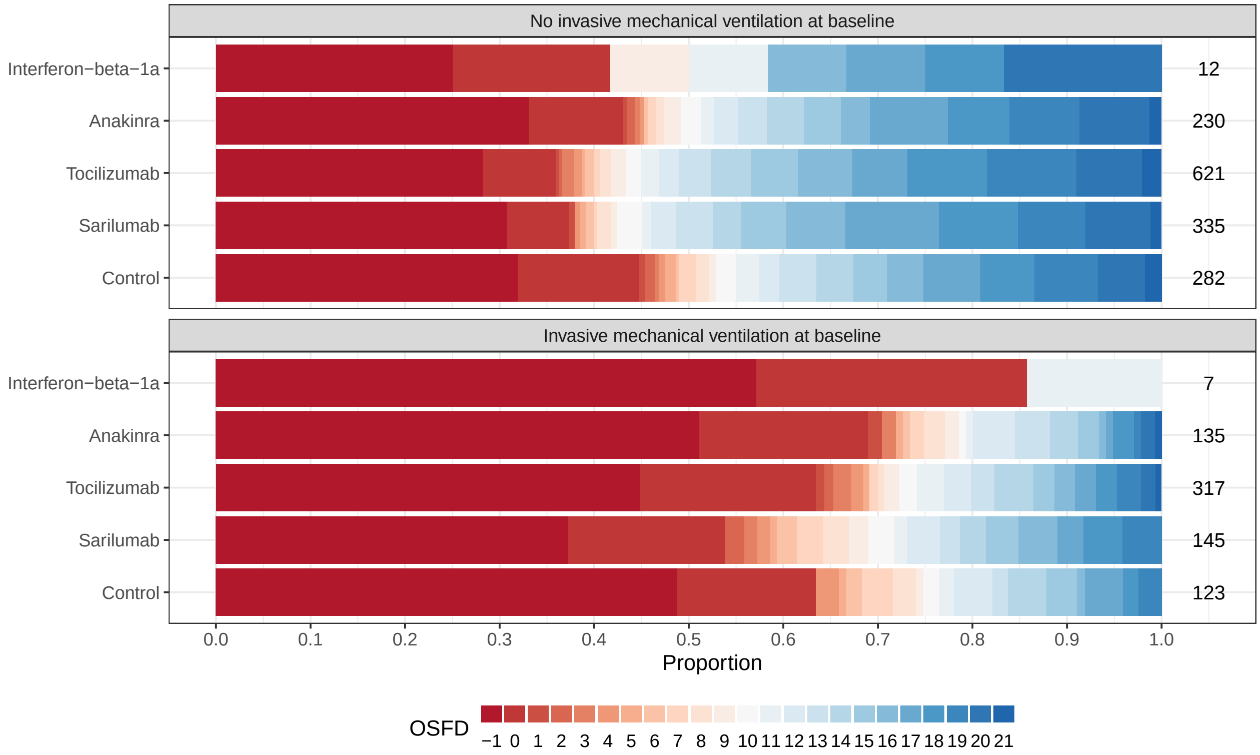

Figure 26: Empirical distribution of organ support free days (OSFD) for interferon-beta-1a, anakinra, tocilizumab, sarilumab, and control by baseline invasive mechanical ventilation status. This plot is restricted to the Immune Modulation ITT population.

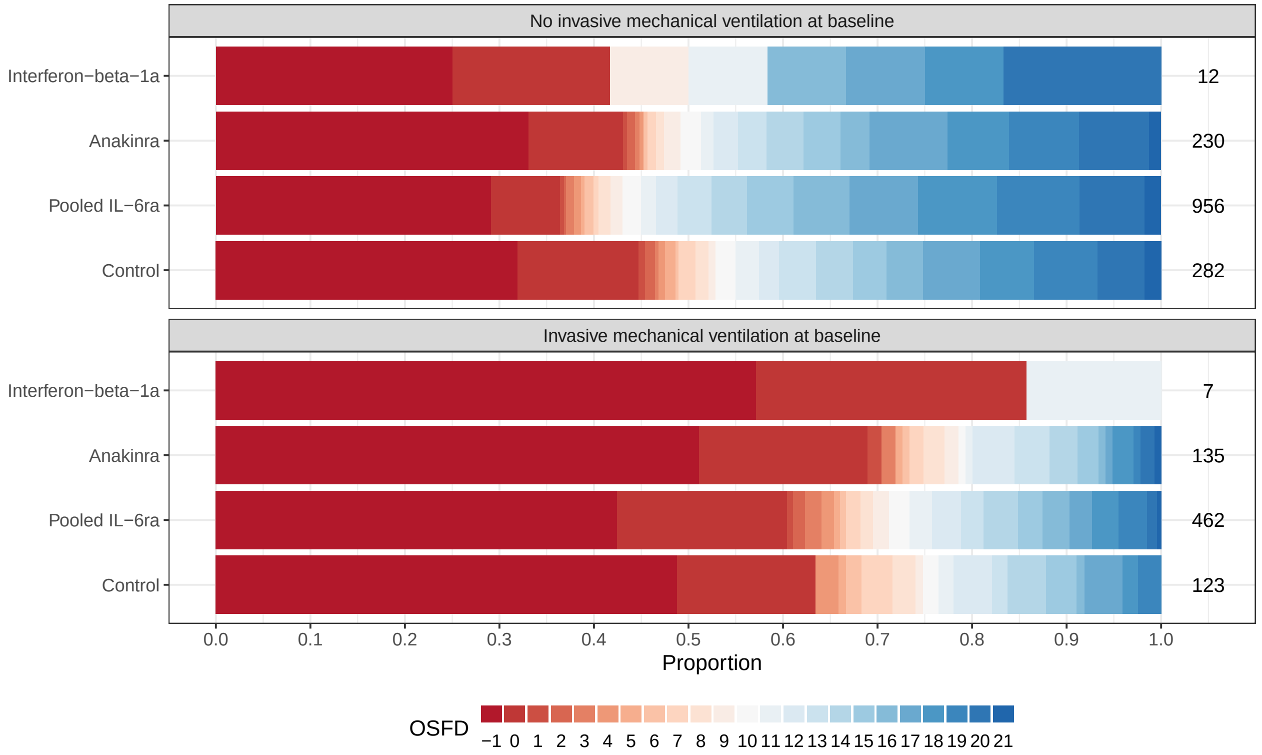

Figure 27: Empirical distribution of organ support free days (OSFD) for pooled IL-6ra and control interventions by baseline invasive mechanical ventilation status. This plot is restricted to the Immune Modulation ITT population.

##### 7.2.2 Subgroup analysis with differential effect on OSFD by baseline mechanical ventilation status

- Model: Primary analysis ordinal model
- Factors: Age, sex, site, time, Immune Modulation Therapy Domain interventions (control, interferon-beta-1a, anakinra, tocilizumab, and sarilumab), COVID-19 Antiviral domain interventions (no antiviral, lopinavir−ritonavir, hydroxychloroquine, combination therapy), Corticosteroid domain interventions (no corticosteroid, fixed-duration corticosteroids, shock-dependent corticosteroids), Immunoglobulin domain interventions (no convalescent plasma, convalescent plasma, delayed convalescent plasma), and Anticoagulation interventions (usual care thromboprophylaxis, therapeutic anticoagulation)
- Differential treatment effects for immune modulation interventions by baseline invasive mechanical ventilation status will be compared to the control arm. A posterior probability of superiority of 99% will be used as a statistical trigger for efficacy for each subgroup.
- Population: Unblinded ITT

Table 84: Odds ratio parameters for subgroup analysis with differential effect on OSFD by mechanical ventilation

| | Mean | SD | Median | CrI |
|---|---|---|---|---|
| Interferon-beta-1a, no IMV | 1.87 | 0.95 | 1.67 | (0.66, 4.24) |
| Interferon-beta-1a, IMV | 0.83 | 0.53 | 0.70 | (0.21, 2.24) |
| Anakinra, no IMV | 1.11 | 0.19 | 1.09 | (0.78, 1.53) |
| Anakinra, IMV | 0.89 | 0.19 | 0.87 | (0.57, 1.31) |
| Tocilizumab, no IMV | 1.62 | 0.20 | 1.61 | (1.26, 2.07) |
| Tocilizumab, IMV | 1.22 | 0.19 | 1.20 | (0.88, 1.64) |
| Sarilumab, no IMV | 1.56 | 0.23 | 1.54 | (1.16, 2.08) |
| Sarilumab, IMV | 1.40 | 0.28 | 1.38 | (0.94, 2.02) |
| Age<39 | 4.39 | 0.54 | 4.36 | (3.43, 5.53) |
| Age 40-49 | 2.29 | 0.22 | 2.28 | (1.88, 2.76) |
| Age 50-59 | 1.83 | 0.15 | 1.82 | (1.56, 2.13) |
| Age 70-79 | 0.51 | 0.05 | 0.50 | (0.42, 0.60) |
| Age 80+ | 0.28 | 0.05 | 0.27 | (0.19, 0.38) |
| Female | 1.20 | 0.08 | 1.20 | (1.06, 1.36) |
| Mechanical ventilation (relative no IMV) | 0.41 | 0.04 | 0.41 | (0.34, 0.48) |

Table 85: Posterior probabilities for subgroup analysis with differential effect on OSFD by mechanical ventilation with antiviral interventions

| | Posterior Probability |
|---|---|
| Interferon-beta-1a is superior to control with no IMV | 0.861 |
| Interferon-beta-1a is futile with no IMV | 0.247 |
| Interferon-beta-1a is superior to control with IMV | 0.274 |
| Interferon-beta-1a is futile with IMV | 0.821 |
| Anakinra is superior to control with no IMV | 0.695 |
| Anakinra is futile with no IMV | 0.710 |
| Anakinra is superior to control with IMV | 0.254 |
| Anakinra is futile with IMV | 0.941 |
| Tocilizumab is superior to control with no IMV | 1.000 |
| Tocilizumab is futile with no IMV | 0.009 |
| Tocilizumab is superior to control with IMV | 0.876 |
| Tocilizumab is futile with IMV | 0.495 |
| Sarilumab is superior to control with no IMV | 0.999 |
| Sarilumab is futile with no IMV | 0.045 |
| Sarilumab is superior to control with IMV | 0.948 |
| Sarilumab is futile with IMV | 0.240 |

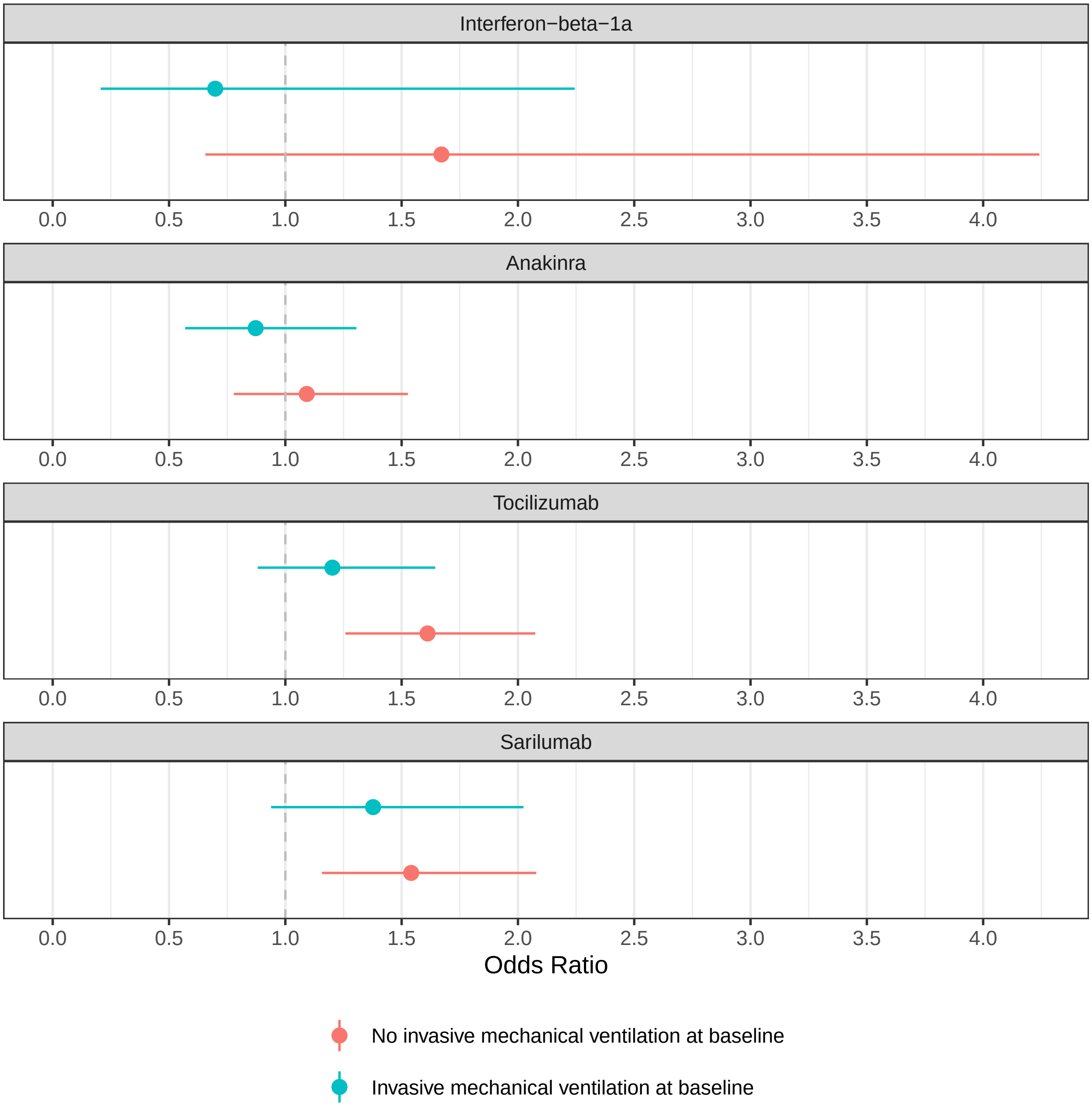

Figure 28: Estimated OSFD odds ratios for interferon, anakinra, tocilizumab, and sarilumab by baseline mechanical ventilation subgroup.

##### 7.2.3 Subgroup analysis with differential effect on hospital survival by baseline mechanical ventilation status

- Model: Primary dichotomous model

- Factors: Age, sex, site, time, Immune Modulation Therapy Domain interventions (control, interferon-beta-1a, anakinra, tocilizumab, and sarilumab), COVID-19 Antiviral domain interventions (no antiviral, lopinavir–ritonavir, hydroxychloroquine, combination therapy), Corticosteroid domain interventions (no corticosteroid, fixed-duration corticosteroids, shock-dependent corticosteroids), Immunoglobulin domain interventions (no convalescent plasma, convalescent plasma, delayed convalescent plasma), and Anticoagulation interventions (usual care thromboprophylaxis, therapeutic anticoagulation)
- Differential treatment effects for immune modulation interventions by baseline invasive mechanical ventilation status will be compared to the control arm. A posterior probability of superiority of 99% will be used as a statistical trigger for efficacy for each subgroup.
- Population: Unblinded ITT

Table 86: Odds ratio parameters for subgroup analysis with differential effect on hospital survival by mechanical ventilation

| | Mean | SD | Median | CrI |
|---|---|---|---|---|
| Interferon-beta-1a, no IMV | 2.13 | 1.38 | 1.78 | (0.59, 5.74) |
| Interferon-beta-1a, IMV | 0.91 | 0.66 | 0.74 | (0.21, 2.59) |
| Anakinra, no IMV | 1.13 | 0.25 | 1.10 | (0.72, 1.70) |
| Anakinra, IMV | 0.84 | 0.22 | 0.81 | (0.50, 1.34) |
| Tocilizumab, no IMV | 1.56 | 0.25 | 1.54 | (1.12, 2.11) |
| Tocilizumab, IMV | 1.18 | 0.23 | 1.16 | (0.79, 1.70) |
| Sarilumab, no IMV | 1.51 | 0.29 | 1.48 | (1.02, 2.16) |
| Sarilumab, IMV | 1.57 | 0.39 | 1.53 | (0.95, 2.45) |
| Age<39 | 8.65 | 1.95 | 8.41 | (5.53, 13.16) |
| Age 40-49 | 3.42 | 0.47 | 3.38 | (2.60, 4.45) |
| Age 50-59 | 2.44 | 0.25 | 2.43 | (1.98, 2.96) |
| Age 70-79 | 0.46 | 0.05 | 0.46 | (0.38, 0.56) |
| Age 80+ | 0.25 | 0.05 | 0.25 | (0.17, 0.36) |
| Female | 1.29 | 0.11 | 1.28 | (1.09, 1.51) |
| Mechanical ventilation (relative no IMV) | 0.59 | 0.07 | 0.59 | (0.47, 0.74) |

Table 87: Posterior probabilities for subgroup analysis with differential effect on hospital survival by mechanical ventilation with antiviral interventions

| | Posterior Probability |
|---|---|
| Interferon-beta-1a is superior to control with no IMV | 0.840 |
| Interferon-beta-1a is futile with no IMV | 0.250 |
| Interferon-beta-1a is superior to control with IMV | 0.325 |
| Interferon-beta-1a is futile with IMV | 0.768 |

Table 87: Posterior probabilities for subgroup analysis with differential effect on hospital survival by mechanical ventilation with antiviral interventions *(continued)*

| | Posterior Probability |
|---|---|
| Anakinra is superior to control with no IMV | 0.678 |
| Anakinra is futile with no IMV | 0.647 |
| Anakinra is superior to control with IMV | 0.208 |
| Anakinra is futile with IMV | 0.938 |
| Tocilizumab is superior to control with no IMV | 0.996 |
| Tocilizumab is futile with no IMV | 0.063 |
| Tocilizumab is superior to control with IMV | 0.768 |
| Tocilizumab is futile with IMV | 0.576 |
| Sarilumab is superior to control with no IMV | 0.980 |
| Sarilumab is futile with no IMV | 0.132 |
| Sarilumab is superior to control with IMV | 0.959 |
| Sarilumab is futile with IMV | 0.163 |

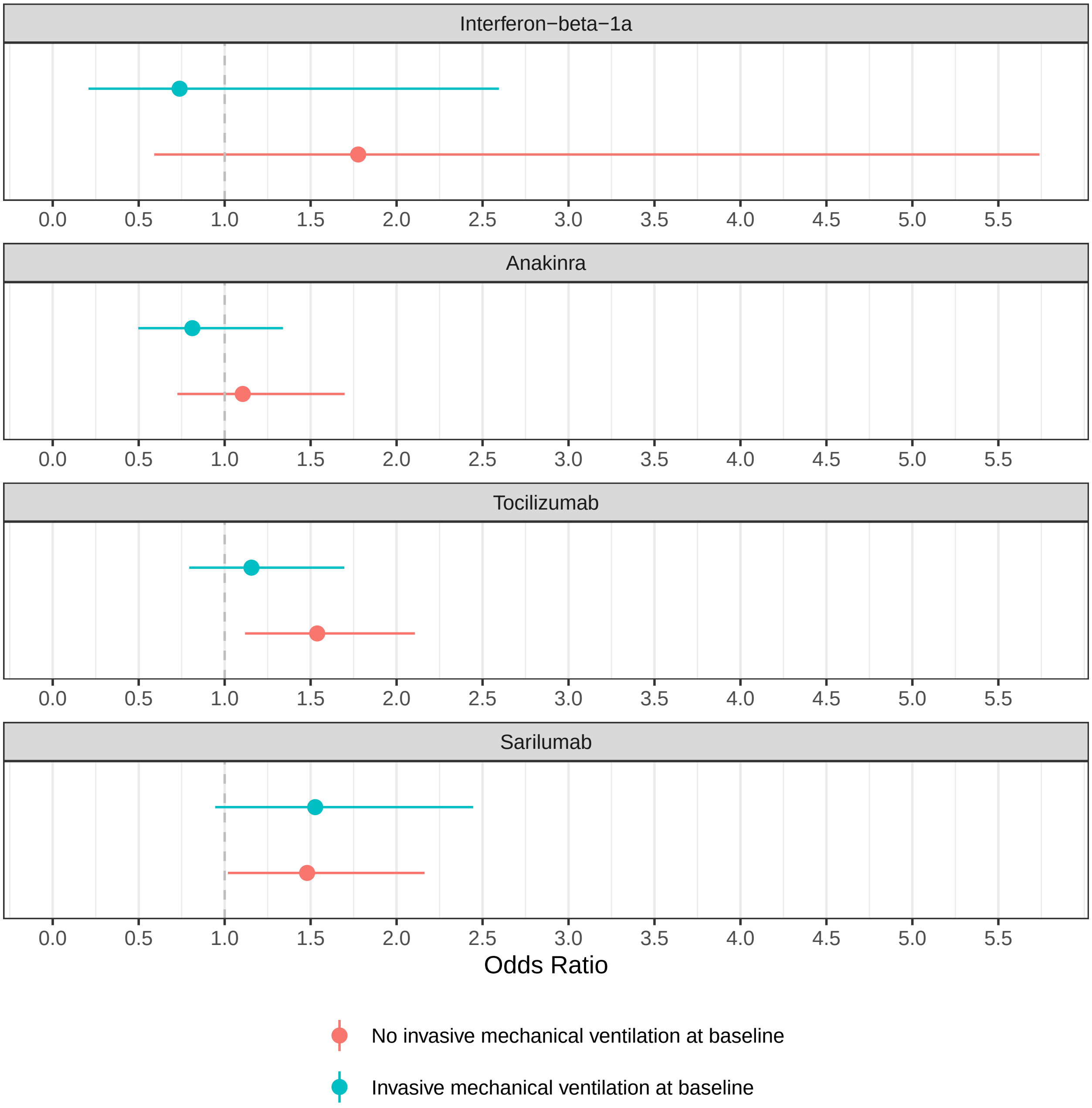

Figure 29: Estimated hospital survival odds ratios for interferon, anakinra, tocilizumab, and sarilumab by baseline mechanical ventilation subgroup.

### 8 Report production and data sources

All analyses in this report are based on the following documents:

- Statistical Analysis Appendix for REMAP-COVID, version 1, dated August 18, 2020;
- Statistical Analysis Plan for the Immune Modulation Therapy Domain for Patients with COVID-19 Pandemic Infection Suspected Or Proven (PISOP), version 1.1, dated April 15, 2021;
- Statistical Analysis Plan for the Tocilizumab Analysis of the Immune Modulation Therapy Domain for Patients with COVID-19 Pandemic Infection Suspected Or Proven (PISOP); version 1.0, dated November 25, 2020;
- Current State of the Statistical Model: Pandemic Model, version 3.1-IM, dated May 5, 2021;
- Addendum to the Immune Modulation PISOP SAP and Current State Documents, dated May 5, 2021

The blinded ITSC analysis team at Berry Consultants performed the analyses in this report using data received from multiple sources. The OSFD outcomes and treatment assignments for patients in unblinded domains were sent from the unblinded Statistical Analysis Committee (SAC) with all blinded information removed. The baseline, discharge and daily data for patients randomized in unblinded domains were sent from an unblinded data coordination team at Monash University. The table below shows the file names for the data exports from each data source and the date each file was received by the blinded ITSC analysis committee. All merging and summarization of data was done using the R statistical computing environment. This report was generated from an Rmarkdown document in the Rstudio software.

Table 88: Summary of data sources

| Filename | Date received | Description |
|---|---|---|
| merged_REMAP_PISOPSevere_IM1_2021-05-11.csv | May 11, 2021 | OSFD and treatment assignments from unblinded SAC |
| Immune modulation baseline discharge compliance data_May3_Designteam.csv | May 10, 2021 | Baseline and discharge data for unblinded ITT |
| Immune Modulation daily data_May3_Designteam.csv | May 10, 2021 | Daily ICU data for unblinded ITT |
